# The Eating Disorders Genetics Initiative (EDGI) United Kingdom

**DOI:** 10.1101/2022.11.11.22282083

**Authors:** Dina Monssen, Helena L Davies, Shannon Bristow, Saakshi Kakar, Susannah C B Curzons, Molly R Davies, Zain Ahmad, John R Bradley, Steven Bright, Jonathan R I Coleman, Kiran Glen, Matthew Hotopf, Emily J Kelly, Abigail R Ter Kuile, Chelsea Mika Malouf, Gursharan Kalsi, Nathalie Kingston, Monika McAtarsney-Kovacs, Jessica Mundy, Alicia J Peel, Alish B Palmos, Henry C Rogers, Megan Skelton, Brett N Adey, Sang Hyuck Lee, Hope Virgo, Tom Quinn, Tom Price, Johan Zvrskovec, Thalia C Eley, Janet Treasure, Christopher Hübel, Gerome Breen

## Abstract

**Objective:** The Eating Disorders Genetics Initiative United Kingdom (EDGI UK), part of the National Institute for Health and Care Research (NIHR) Mental Health BioResource, aims to deepen our understanding of the environmental and genetic aetiology of eating disorders. EDGI UK launched in February 2020 and is partnered with the UK eating disorders charity, Beat. There are multiple EDGI branches worldwide.

**Method:** EDGI UK recruits via media and clinical services. Anyone living in England, at least 16 years old, with a lifetime probable or clinical eating disorder is eligible to sign up online: edgiuk.org. Participants complete online questionnaires, donate a saliva sample for genetic analysis, and consent to medical record linkage and recontact for future studies.

**Results:** As of September 2022, EDGI UK has recruited 8,397 survey participants: 98% female, 93% white, 97.7% cisgender, 67% heterosexual, and 52% have a university degree. Half (51.7%) of participants have returned their saliva kit. The most common diagnoses are anorexia nervosa (42.7%), atypical anorexia nervosa (31.4%), bulimia nervosa (33.2%), binge-eating disorder (14.6%), and purging disorder (33.5%).

**Conclusion:** EDGI UK is the largest UK eating disorders study but needs to increase its diversity, and efforts are underway to do so. It also offers a unique opportunity to accelerate eating disorder research, and collaboration between researchers and participants with lived experience, with unparalleled sample size.

## 1. Introduction

### 1.1 Eating disorders

Anorexia nervosa, bulimia nervosa, binge-eating disorder, and other specified feeding or eating disorders (OSFED) affect ∼8% of the global population, predominantly females, with OSFED being the most prevalent (Galmiche et al., 2019). Preliminary prevalence estimates of the newly recognised eating disorder, avoidant restrictive food intake disorder (ARFID), lie at 3% in children (Kurz et al., 2015). Eating disorders are often chronic, with high relapse rates, but about half of those with anorexia or bulimia nervosa reach remission with available treatments (Berkman et al., 2007; Eddy et al., 2017). Barriers such as stigma around eating disorders, secrecy, and lack of research funding result in small samples. Research to fully understand eating disorders is therefore challenging (Ali et al., 2017; Regan et al., 2017).

Eating disorders cause substantial costs to healthcare systems and reduce workforce participation, yet many patients’ medical needs are unmet (Ágh et al., 2016; Schaumberg et al., 2017). Individuals with eating disorder symptoms often do not seek help, therefore numbers of affected individuals are likely much higher than reported (Hart et al., 2011). Only ∼1% of the UK Government’s annual mental health funding was allocated to eating disorder research between 2015 to 2019, despite eating disorders comprising 9% of those with a psychiatric disorder in the UK (All-Party Parliamentary Group on Eating Disorders, 2021). Given such funding constraints, large-scale research studies are difficult to conduct.

Eating disorders are highly comorbid with psychiatric and somatic illnesses. Approximately 70% of individuals with eating disorders have at least one other lifetime psychiatric disorder, including mood and anxiety disorders, obsessive-compulsive disorders, and substance use disorders (Eskander et al., 2020; Momen et al., 2022). Common comorbid somatic diseases in binge-eating disorder include obesity and weight-related problems, such as hypertension, diabetes, and osteoporosis. Individuals with anorexia nervosa often experience reproductive complications and osteoporosis, whereas people with bulimia nervosa experience gastrointestinal diseases. These include gastric acid reflux, dysphagia, dyspepsia, and cathartic colon syndrome, often due to compensatory behaviours such as self-induced vomiting (Giel et al., 2022; Gosseaume et al., 2019; Momen et al., 2021; Westmoreland et al., 2016). As psychiatric and somatic comorbidities are common, it is important to explore shared biological pathways with eating disorders and how they affect therapy outcomes.

### 1.2 Genetic factors

Independently replicated heritability estimates from family and twin studies show a substantial genetic component of anorexia nervosa (58-74%), bulimia nervosa (59-83%), and binge-eating disorder (41-57%) (Thornton et al., 2011). Moreover, a twin study estimated the heritability of ARFID at 79%, using a combination of characteristics to assign twins a probable diagnosis (Dinkler et al., 2022). Genome-wide association studies (GWASs) can help us to identify eating disorder-associated common genetic variants that each contribute a small effect to overall risk; however, these in concert, can explain a sizable percentages of trait variance, so called single-nucleotide polymorphism-(SNP)-based heritability. For example, attention-deficit/hyperactivity disorder (ADHD) has a SNP-based heritability of 14% (Demontis et al., 2022), major depressive disorder (MDD) has a SNP-based heritability of 8.9% (Howard et al., 2019; Wray et al., 2018), whilst anorexia nervosa has a SNP-based heritability of 17% (Hübel, Gaspar, Coleman, Finucane, et al., 2019; Hübel, Gaspar, Coleman, Hanscombe, et al., 2019; Watson et al., 2019). Results from GWASs of eating disorders can be used to understand the biological aetiology of the disorder and pinpoint potential drug targets. Additionally, these common genetic variants can be included in risk prediction models to stratify patients into groups bearing different risks for therapy success or medication side effects (Sullivan et al., 2018).

Large sample sizes are essential for the success of GWASs of complex polygenic traits like psychiatric disorders, because they provide the high precision necessary to estimate the very small effects of the individual genetic variants. These can be achieved by combining international datasets (Schizophrenia Working Group of the Psychiatric Genomics Consortium, 2014; Sullivan et al., 2018). Several GWASs have successfully identified genomic regions (i.e., loci on chromosomes) associated with psychiatric disorders that commonly co-occur with eating disorders, like ADHD and MDD (Hübel et al., 2018). For example, in ADHD, a sample of 38,691 individuals with ADHD and 186,843 controls was required to identify 27 independent genome-wide significant loci (Demontis et al., 2022). For MDD, 246,363 cases and 561,190 controls were required to identify 101 independent loci (Howard et al., 2019; Wray et al., 2018). These genetic findings from twin studies and GWASs of other psychiatric disorders motivated molecular genetic research in eating disorders and clearly indicated the likely required sample sizes.

Genetic research in eating disorders lags behind other psychiatric disorders due to funding constraints that complicate recruiting large samples with sufficient case numbers, resulting in low statistical power. Despite this, in 2019, the Eating Disorders Working Group of the Psychiatric Genomics Consortium (PGC-ED) successfully combined 33 datasets in the largest GWAS of anorexia nervosa. Eight significant loci were identified, along with several statistically significant genetic correlations with other psychiatric disorders (e.g., obsessive-compulsive disorder [OCD], major depressive disorder [MDD], schizophrenia), metabolic traits (e.g., high-density lipoprotein cholesterol, fasting insulin concentrations), and accelerometer-measured physical activity (Hübel, Gaspar, Coleman, Finucane, et al., 2019; Hübel, Gaspar, Coleman, Hanscombe, et al., 2019; Watson et al., 2019). This reconceptualised anorexia nervosa as a metabo-psychiatric disorder. No GWASs of clinically diagnosed bulimia nervosa, binge-eating disorder, or ARFID have yet been conducted, but GWASs using proxy phenotyping approaches have been performed. In a GWAS of machine learning predicted binge-eating disorder based on details in medical records, three loci were genome-wide significant, after adjusting for body mass index (BMI; (Burstein et al., 2022). The first GWAS of ARFID used proxy phenotyping within a subsample of the autism study SPARK, including 3,142 autistic children and 2,205 of their parents (Koomar et al., 2021). A probable ARFID diagnosis was assigned using a score on the Nine-Item ARFID Screen (NIAS; (Zickgraf & Ellis, 2018) alongside other measures, and one genetic variant reached genome-wide significance (Koomar et al., 2021). Generalisability of the ARFID GWAS is limited as it was statistically underpowered and based on a highly selected sample, however, it enables future power calculations. To understand the genetic and biological underpinnings of eating disorders, statistically well-powered GWASs of all eating disorders in large diverse samples are urgently needed.

### 1.3 Environmental and psychological factors

Environmental and psychological factors play a role in the aetiology of eating disorders, with some occurring in early life (McClelland et al., 2020; Solmi et al., 2021). Environmentally, premature birth and older parental age have been associated with increased risk of an eating disorder in a nationwide Danish study (Larsen et al., 2021) as well as traumatic experiences during early life in systematic reviews (Molendijk et al., 2017; Palmisano et al., 2016). Another potential environmental risk factor is social media use, which has been longitudinally associated with having a negative body image (de Valle et al., 2021). In contrast, positive relationships with parents and family members, including regular family meal times, not commenting on the young person’s weight, and body appreciation can be protective against eating disorder development (Langdon-Daly & Serpell, 2017). Psychologically, low body satisfaction in early adolescent girls is a strong risk factor for an eating disorder in later adolescence (Stice et al., 2011). In some females, dietary restriction to control weight, self-induced vomiting, or laxative intake precedes binge-eating disorder and bulimia nervosa (Allen et al., 2013). If dieting or drive for thinness co-occurs with depressive or anxious symptoms, this may increase risk for eating disorders (McClelland et al., 2020). As longitudinal studies are rare, it is difficult to distinguish prodromal symptoms from risk factors. However, how environmental factors interact with genetic liability to trigger eating disorder development is not yet understood.

### 1.4 Aims of EDGI UK

The aim of the Eating Disorders Genetics Initiative United Kingdom (EDGI UK; edgiuk.org) is to improve our understanding of all eating disorders by collecting genetic and phenotypic data from at least 10,000 people, and linking this with their medical records. Furthermore, EDGI UK acts as a resource for recontact of volunteers, as part of the National Institute for Health and Care Research (NIHR) BioResource, enabling secondary analysis. The NIHR BioResource is a UK-based database of >250,000 volunteers who are willing to be recontacted for future research. All EDGI UK survey participants provide consent to take part both in EDGI UK and in the NIHR Mental Health BioResource. Internationally, EDGI UK is part of a research collaboration, EDGI World (Bulik et al., 2021). Across research teams based in the USA, Australia, New Zealand, Sweden, Puerto Rico, and Mexico, EDGI World aims to collect 100,000 genetic samples of each eating disorder. As of 2022, more countries (e.g., Italy) are opening EDGI branches. EDGI UK represents a novel and vital resource for research both in the eating disorder field and for associated comorbidities.

## 2 Methods

### 2.1 Pre-launch

#### Collaboration with EDGI World and Beat

EDGI World is the follow-up study of the Anorexia Nervosa Genetics Initiative (ANGI; **Figure 1**). The UK branch pioneered the use of detailed phenotyping, and other branches around the world adopted the UK approach by adapting its assessment batteries to their local needs. Additionally, EDGI UK formed a partnership with Beat, a UK eating disorder charity (beateatingdisorders.org.uk). Beat’s experience and insight as part of the UK’s All-Party Parliamentary Group for Eating Disorders (a group of UK Government members and experts working to improve policy for eating disorders) was invaluable to the success of EDGI UK. Beat also offers helplines, raises awareness, disseminates information on eating disorders, and promotes research projects including EDGI UK. For instance, EDGI UK participates in Beat’s annual conference during Eating Disorders Awareness Week, and EDGI UK has featured on Beat’s social media platforms, website, and mailing lists. In addition to this, Beat’s ambassadors – volunteers with lived experience or caring for someone with an eating disorder – offered their time to be featured in promotional materials and provided insight into the user experience. Beat ambassadors are also a part of EDGI UK’s Steering Committee.

**Figure 1.**
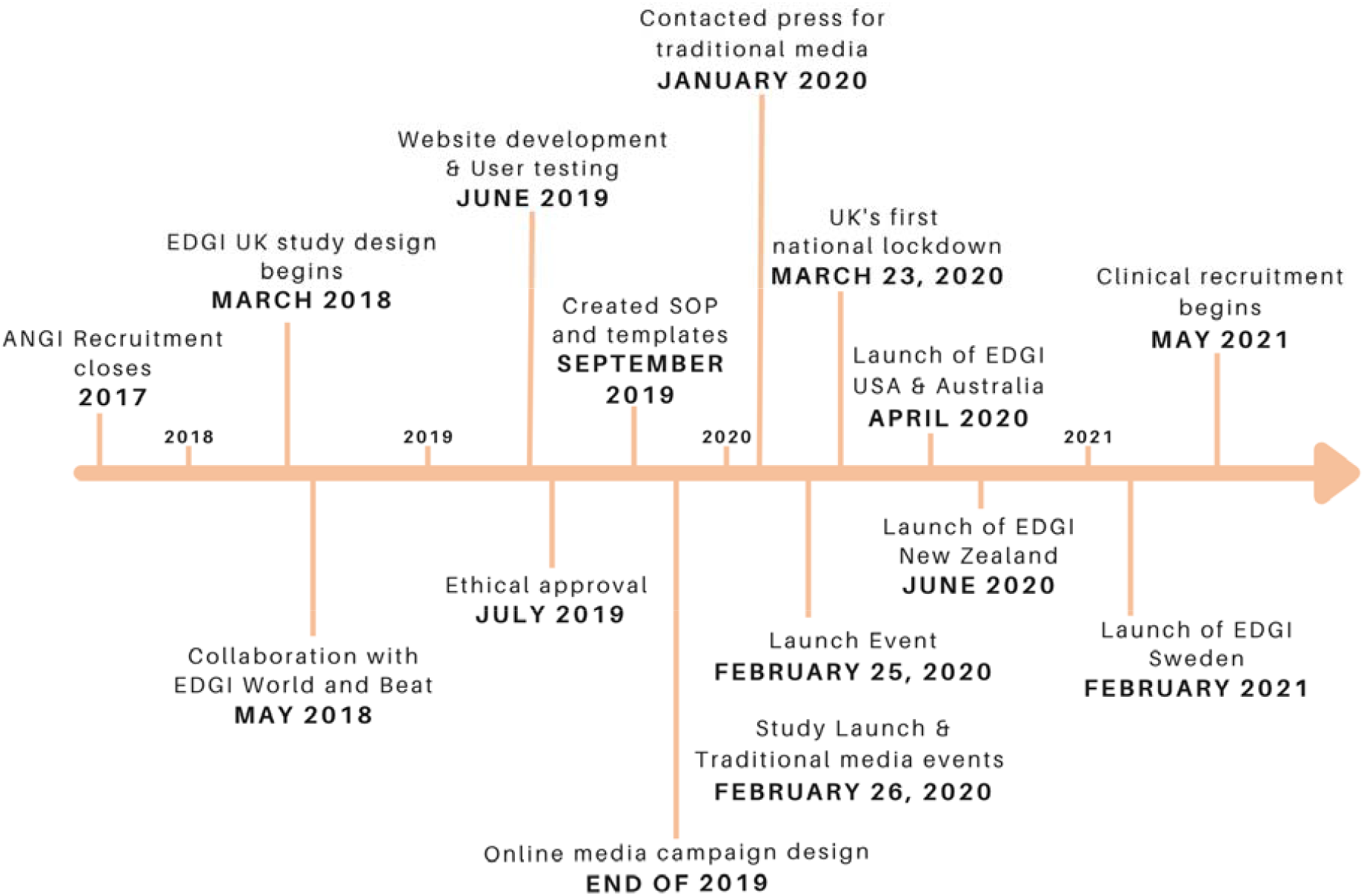
Timeline of Eating Disorders Genetics Initiative United Kingdom (EDGI UK). The timeline begins at the end of the Anorexia Nervosa Genetics Initiative (ANGI) in 2017 and ends with the writing of this manuscript in September 2022.

**Figure 2.**
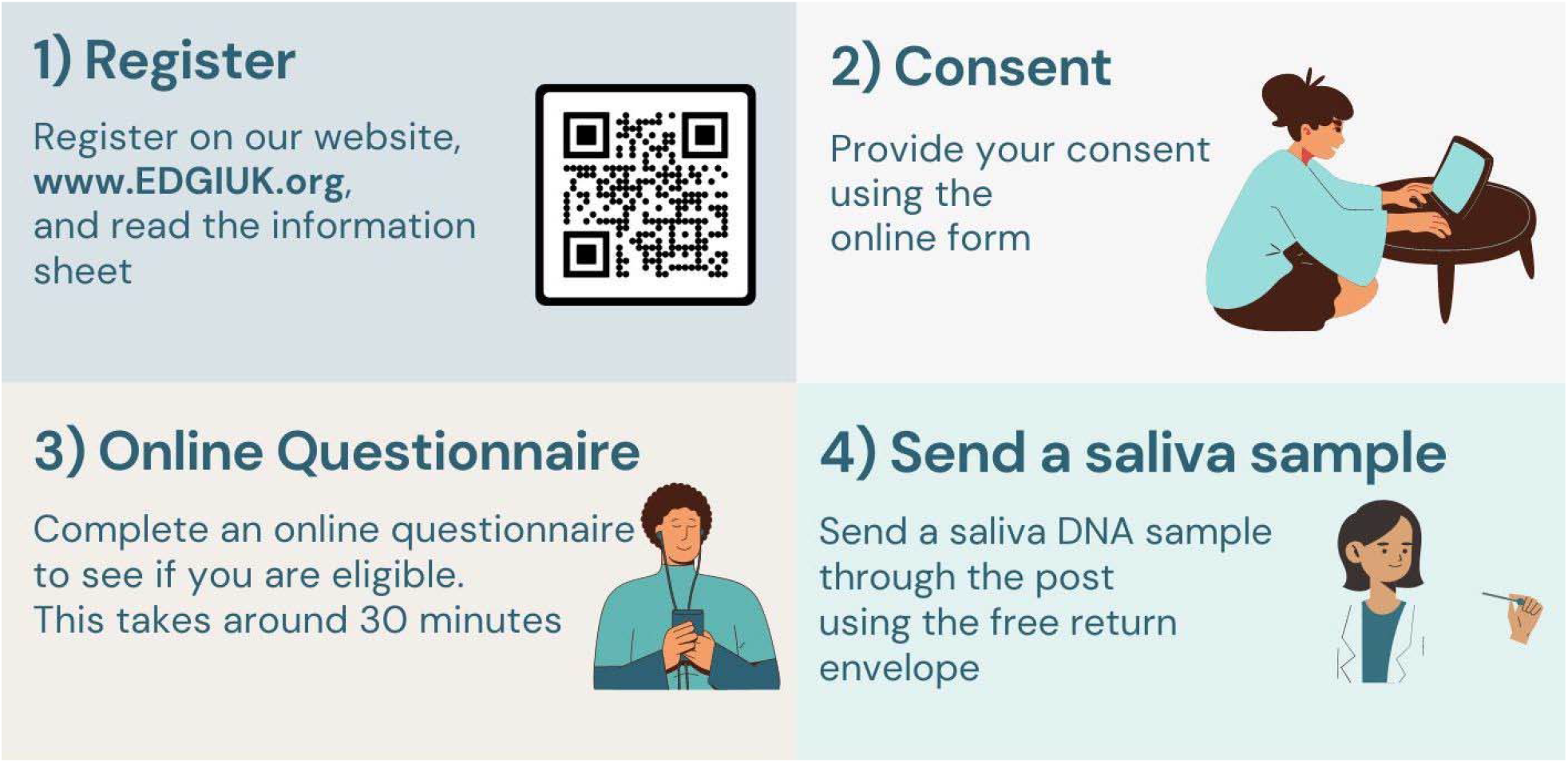
Sign-up process of Eating Disorders Genetics Initiative United Kingdom (EDGI UK). The four-step sign-up process for participants of EDGI UK, highlighting registration, consent, completing the online questionnaire, and the return of their saliva sample.

**Figure 3.**
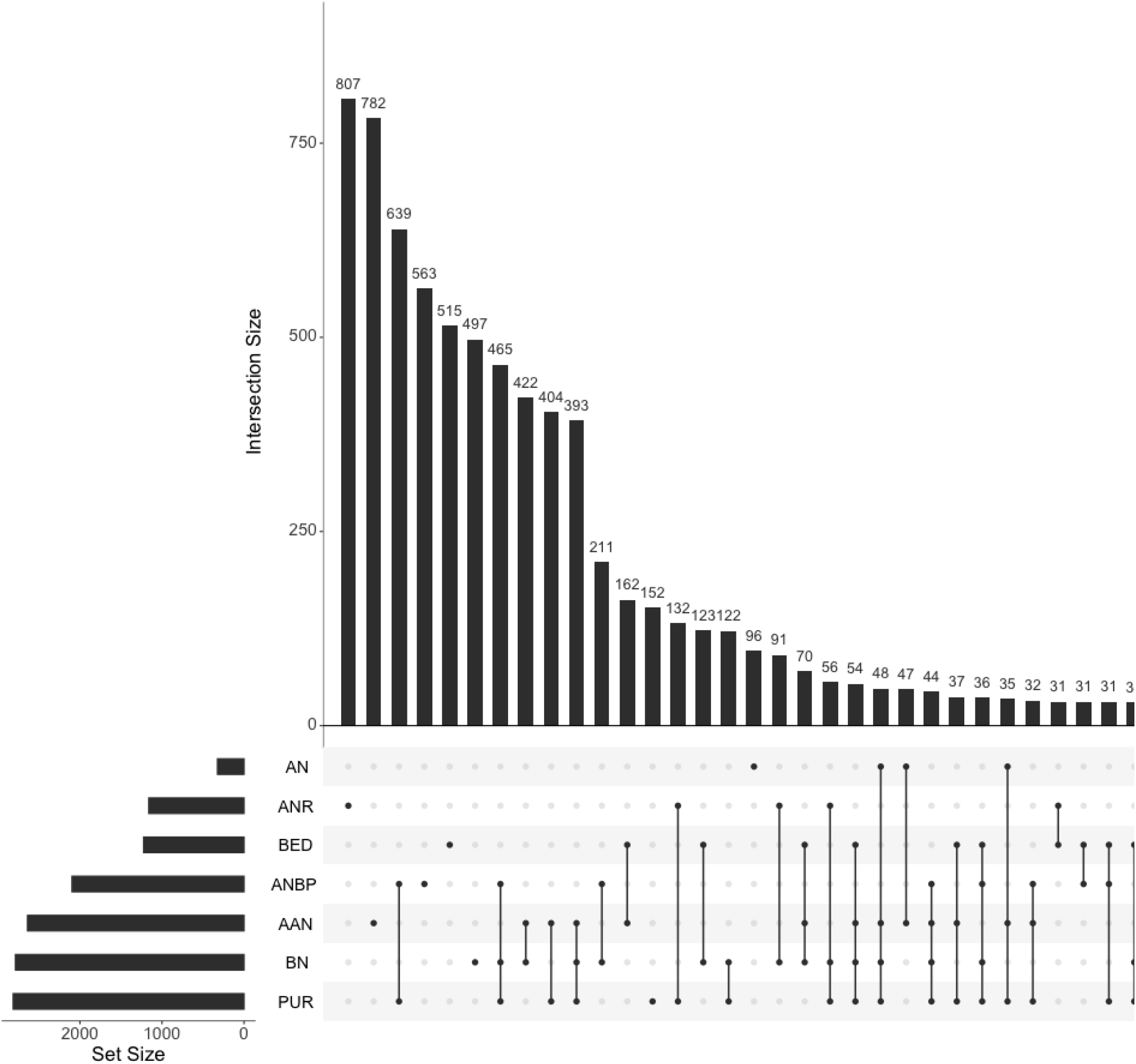
Overlap of lifetime eating disorder diagnoses in Eating Disorders Genetics Initiative United Kingdom (EDGI UK) survey participants. Diagnoses are algorithm-derived via responses to the ED100K and/or self-report in the Mental Health Diagnosis (MHD) questionnaire. Some eating disorders are algorithm-derived only (anorexia nervosa binge-eating/purging and anorexia nervosa restricting). Set size represents overall size of each group, whilst intersection size indicates size of overlapping groups. Groups smaller than 20 are not presented. See Supplementary Figure 1 for more information on smaller groups. Abbreviations: AN = anorexia nervosa (no subtype), ANR = anorexia nervosa restricting, BED = binge-eating disorder, ANBP = anorexia nervosa binge-eating/purging, AAN = atypical anorexia nervosa, BN = bulimia nervosa, PUR = purging disorder.

#### Website development

Participation for EDGI UK takes place on our website edgiuk.org. The website development was outsourced to Mindwave Ventures who based the design on our sister study, the Genetic Links to Anxiety and Depression (GLAD) Study (gladstudy.org.uk) (M. R. Davies et al., 2019). The colour scheme showcases our partnership with Beat whose Public Relations and Communications team advised on the website design. Beat ambassadors and other volunteers participated in user testing for ∼1 hour and provided feedback through in-person group sessions or online. We trialled participant-facing materials, website design, and functionality. Volunteers were compensated with a £10 Amazon voucher. The research team reviewed and incorporated the feedback to optimise user experience.

#### Standard operating procedures (SOPs) and templates

The EDGI UK team created SOPs including response templates for participant contact, database management, use of postal services for saliva kit posting, and record-keeping of saliva kits. To provide timely, standardised responses to participant queries, we created email, social media, and voicemail template responses, which explained the purpose of the study, website usage, saliva sample procedures, and an overview of genetics.

#### Endorsement through organisations and individuals

We approached organisations, charities, politicians, and mental health activists prior to the launch to ask for their endorsement by advertising EDGI UK on their public platforms (e.g., charity websites, social media profiles). EDGI UK sent an information package, detailing recruitment support strategies, for example, by being a spokesperson or providing a quote for social media. The package included infographics with sign-up information, hashtags, and handles for social media posts. We were endorsed by mental health organisations, including F.E.A.S.T, Body Dysmorphic Disorder Foundation, MQ Mental Health, The Motherhood Group, and Mental Health Foundation. Mental health activists Hope Virgo and Dave Chawner are spokespeople for EDGI UK. On EDGI UK’s one year anniversary, further influencers, athletes, and mental health activists supported and endorsed EDGI UK.

### 2.2 Launch event

We hosted an EDGI UK launch event with members of the eating disorder community, National Health Service (NHS) staff including eating disorder clinicians, representatives from different universities, the NIHR BioResource, and Beat. Beat, the NIHR BioResource, EDGI UK’s chief investigators, and people with lived experience presented on the importance of our research. The event was concluded by a panel discussion with an audience-guided question-and-answer session.

### 2.3 Traditional media campaign

The media agency Bright Star Digital, the NIHR BioResource and the King’s College London Communications and Engagement teams contacted media outlets for traditional media coverage (e.g., newspapers, radio stations). Sky News, a UK television news channel, interviewed Professor Gerome Breen and Hope Virgo. Professor Janet Treasure and Andrew Radford, Beat’s Chief Executive, spoke to BBC Radio 4 Woman’s Hour. Ten regional radio stations played pre-recorded content by Professor Gerome Breen. Recordings are not available due to copyright and must be bought from the publisher. Tom Quinn, the Director of External Affairs for Beat, spoke to ITV News, the UK’s largest commercial news organisation. ITV News, Pharma Times, the NIHR BioResource, and Beat published online articles (**Supplementary Material 1**). Media coverage was not as large as hoped, as COVID-19 reporting started to dominate the news agenda (EDGI UK launched three weeks prior to the UK Government’s first national COVID-19 lockdown in March 2020).

### 2.4 Social media campaign

EDGI UK used several online strategies for advertisement. The agency, Born Social (bornsocial.com) developed and disseminated paid social media advertisements on Instagram® and Facebook®, targeting young people of all genders. We encouraged participation by describing the simple process involved, using key statistics, and emphasising that EDGI UK does not discriminate by eating disorder type, gender, or the receipt of a clinical diagnosis. To capture a wide audience, the first advertisement addressed all eating disorders, whereas the second specifically focused on binge-eating disorder, which is underdiagnosed and often overlooked in research (Kornstein et al., 2016). To align with Beat’s media values, we used animated body-image neutral figurines or included a range of different body types. The advertisement captions initially included trigger warnings, which were later removed due to inadequate evidence demonstrating their efficacy (Sanson et al., 2019), and the consideration that trigger warnings may perpetuate stigma, as outlined in edgiuk.org/trigger-warning-statement. Participants were sent an “opt-out” form if they did not want to see our paid advertisements on their Instagram® feeds.

In addition to the agency-created content, we created a six-week social media content plan for Instagram®, Facebook®, and Twitter®. Posts included quotes from EDGI UK spokespeople, infographics, and information on EDGI UK and eating disorders. They also carried Beat’s, EDGI UK’s, and the NIHR BioResource’s logos to showcase our collaborations. We pinned an in-house produced “Call to Action” video to the top of our Twitter® page: lived experience volunteers spoke about their eating disorders and our chief investigators highlighted the urgent research need and explained the participation process.

### 2.5 Clinical recruitment

NHS England Trusts are hospital and community services that provide patient care in circumscribed geographical regions. By September 2022, EDGI UK recruited 200 participants through 13 of England’s NHS trusts. The 13 trusts included ten specialist First Episode Rapid Early Intervention for Eating Disorders (FREED) services (Richards et al., 2022). Clinical recruitment was delayed until early 2021 due to COVID-19 restrictions and availability of medical staff. Each trust displays posters, distributes leaflets, sends texts or letters to patients with eating disorders, or approaches patients in-clinic to assist with registration and completing questionnaires.

### 2.6 Eligibility criteria

To take part in EDGI UK, participants must be aged 16 years or over, reside in England, and have a lifetime experience of an eating disorder as defined in the Diagnostic and Statistical Manual of Mental Health, Fifth Edition, (DSM-5(American Psychiatric Association, 2013) or the International Classification of Diseases, Eleventh Revision (ICD-11; (World Health Organisation (WHO), 2019). Individuals who have not been clinically diagnosed or treated for an eating disorder can still participate as eligibility is determined through the sign-up questionnaire. As of September 2022, due to funding limitations and ethical approval, EDGI UK is limited to England. However, participants living abroad with a home address in England are eligible (e.g., university students). Ineligible individuals who self-report lifetime anxiety and/or depressive disorders are invited to the GLAD Study (gladstudy.org.uk). Approximately half of EDGI UK survey participants have returned saliva samples for genotyping thus far, so some participants are not full members of the study. This paper includes all participants who have completed the sign-up questionnaire regardless of whether they have returned a saliva sample, therefore throughout this manuscript we refer to them as “EDGI UK survey participants”.

### 2.7 Sign-up process

Interested individuals can watch an animated video explaining EDGI UK and the NIHR BioResource before deciding to take part. On edgiuk.org, participants sign up by creating an account, filling in their name, contact details, sex, gender, and date of birth. Subsequently, they are required to read the participant information sheet and must fully consent electronically. Mandatory consent for EDGI UK includes joining the NIHR BioResource, permitting long-term de-personalised storage of data, donating a biological sample, being recontacted to take part in future research projects based on their genetic and/or phenotypic data, being contacted for additional questionnaires, and linkage of their medical records. Once consented, participants must complete the EDGI UK sign-up questionnaire. Questionnaire responses are used to assess eligibility before participants are mailed a saliva kit.

### 2.8 Measures

The online assessment process is split into two elements: a sign-up questionnaire and numerous optional questionnaires. First, the sign-up questionnaire assesses basic demographics, psychiatric and somatic comorbidity, and family history via the NIHR BioResource Health and Lifestyle questionnaire. Subsequently, the ED100K questionnaire assesses lifetime eating disorder symptoms, their severity, and duration (Thornton et al., 2018). The ED100K is augmented by questions covering atypical anorexia nervosa, the validated 7-Item Binge Eating Disorder Screener (BEDS-7; (Herman et al., 2016), the Night Eating Diagnostic Questionnaire (NEDQ; (Gluck et al., 2001), the Muscle Dysmorphic Disorder Inventory (MDDI; (Hildebrandt et al., 2004), and the Nine-Item Avoidant/Restrictive Food Intake Disorder Screen (NIAS; (Zickgraf & Ellis, 2018) (**Supplementary Material 2**). Moreover, at sign-up, participants can answer or skip the Childhood Trauma Screener (CTS-5; (Glaesmer et al., 2013), questionnaires regarding adverse life experiences including domestic violence and catastrophic trauma (Davis et al., 2020), and the Post-Traumatic Stress Disorder Checklist (PCL-6; (Lang et al., 2012). The sign-up questionnaire takes ∼30-45 minutes depending on routing. Participants can then access 32 additional optional questionnaires to provide further information on symptomatology, comorbidity, treatment experience, and social impairment. The optional questionnaires are grouped into six categories: eating behaviours and physical activity, thoughts and behaviours, lifestyle, quality of life, personality, and treatment (for a full list of measures, see **Supplementary Material 3**). Our assessment battery mirrors large-scale data collections: the UK Biobank (Davis et al., 2020), two Swedish eating disorder quality registers (Stepwise and Riksät; (Birgegård et al., 2010), and EDGI World.

### 2.9 Biological samples, genotyping, genetic analysis

We send eligible participants a parcel with a GFX Isohelix Saliva Collector through the post containing an instructions leaflet (**Supplementary Material 4**) for self-sampling at home. Participants can return their samples free of charge via post. Participants receive up to six emails and two text message reminders to complete and post their kit, along with tips on how to provide a saliva sample. It is the responsibility of the participants to post their sample to the National Biosample Centre in Milton Keynes, where DNA is extracted following the manufacturer’s instructions, then handled and stored in the UK Biobank storage and sample handling facility. Genome-wide genotyping and standard quality control will be undertaken using the Thermo Fisher microarray. Under the umbrella of the PGC-ED, we will perform international GWAS meta-analyses by combining the genotypes of our participants with data from the UK Biobank, EDGI World, and the Lundbeck Foundation Initiative for Integrative Psychiatric Research, Denmark.

### 2.10 Participant recontact & access to EDGI UK

As a recontactable database, researchers can submit proposals for participant recontact and access de-identified data for their own research projects. During the COVID-19 pandemic, participants were invited to take part in the COVID-19 Psychiatry and Neurological Genetics (COPING) study, which began at the first national lockdown to monitor the effect of the pandemic on mental health and in which >900 EDGI UK survey participants took part (H. L. Davies et al., 2022; Young et al., 2021). As of 2022, EDGI UK is part of the UK Longitudinal Linkage Collaboration (UK LLC), a national collaborative effort for collating and harmonising health data to support longitudinal research. This will allow for the exploration of research topics including physical and mental health to inform health research and policy.

### 2.11 Ethical approval

EDGI UK was approved by the London - Fulham Research Ethics Committee on 29th July, 2019 (REC reference: 19/LO/1254) following a full review. The NIHR BioResource has been approved as a Research Tissue Bank by the East of England - Cambridge Central Committee (REC reference: 17/EE/0025).

### 2.12 Statistical analysis

Participants were included for analysis if they had complete data on age, sex, and reported lifetime experience of an eating disorder (*n* = 8,397). For continuous variables, we calculated means, standard deviations, medians, and interquartile ranges. We also show minimum and maximum values. For categorical variables, we present absolute numbers and percentages of the total sample, unless otherwise stated. Diagnoses were derived from DSM-5-based algorithms using responses to the ED100K and/or self-report via the Mental Health Diagnosis (MHD) questionnaire (Davis et al., 2020; Thornton et al., 2018). Some eating disorders are algorithm-derived only (i.e., anorexia nervosa binge-eating/purging and anorexia nervosa restricting) whilst others are self-report only (i.e., pica, rumination disorder, and ARFID).

## 3 Results

### 3.1 Sample descriptors

Recruitment of EDGI UK is ongoing and results are from participants recruited before 16^th^ September 2022. In the first week of recruitment, 2,236 participants signed up to EDGI UK and 780 of these participants (35%) completed the questionnaire. Within the first month, 4,389 had signed up and 1,537 (36%) had completed their questionnaire. As of 16^th^ September 2022, 15,339 people had registered, 10,319 (67%) people had consented to participate, 8,397 (54%) people had completed the sign-up questionnaire (constituting the EDGI UK survey participants), and 4,294 (28%) had returned their saliva kits for genotyping. We recruited 5,758 participants (∼73% of eligible individuals) via social media who self-reported hearing about the study on Facebook, Twitter®, or Instagram. By September 2022, @edgi_uk had over 4,000 Instagram followers. Most of these followers are women (96%), aged between 25-34 years, and based in the UK, predominantly London. On Twitter, @edgi_uk has 1,014 followers, and on Facebook, @edgi.uk has 814. On the latter platforms, no detailed information is available.

EDGI UK survey participants are on average 30 years old (**Figure 4**), and mostly female (98%), cisgender (97.7%), white (93%), heterosexual (67%), have a university degree or equivalent (52%), and are in paid employment (55%) (**Table 1**). The five most common lifetime eating disorders reported are anorexia nervosa (42.8% of females, 33.6% of males), atypical anorexia nervosa (31% of females, 29% of males), bulimia nervosa (33% of females, 30% of males), binge eating disorder (15% of females, 15% of males), and purging disorder (34% of females, 21% of males) (**Table 2**). Demographic characteristics are largely consistent across the whole sample and in each of the five most common eating disorder groups (**Table 1**). Over half of participants report that they have received treatment at some point in their life (61%; **Table 1**). Treatment receipt was most often reported by those with lifetime anorexia nervosa (85%), and least often by those with binge-eating disorder (44%).

**Table 1.**
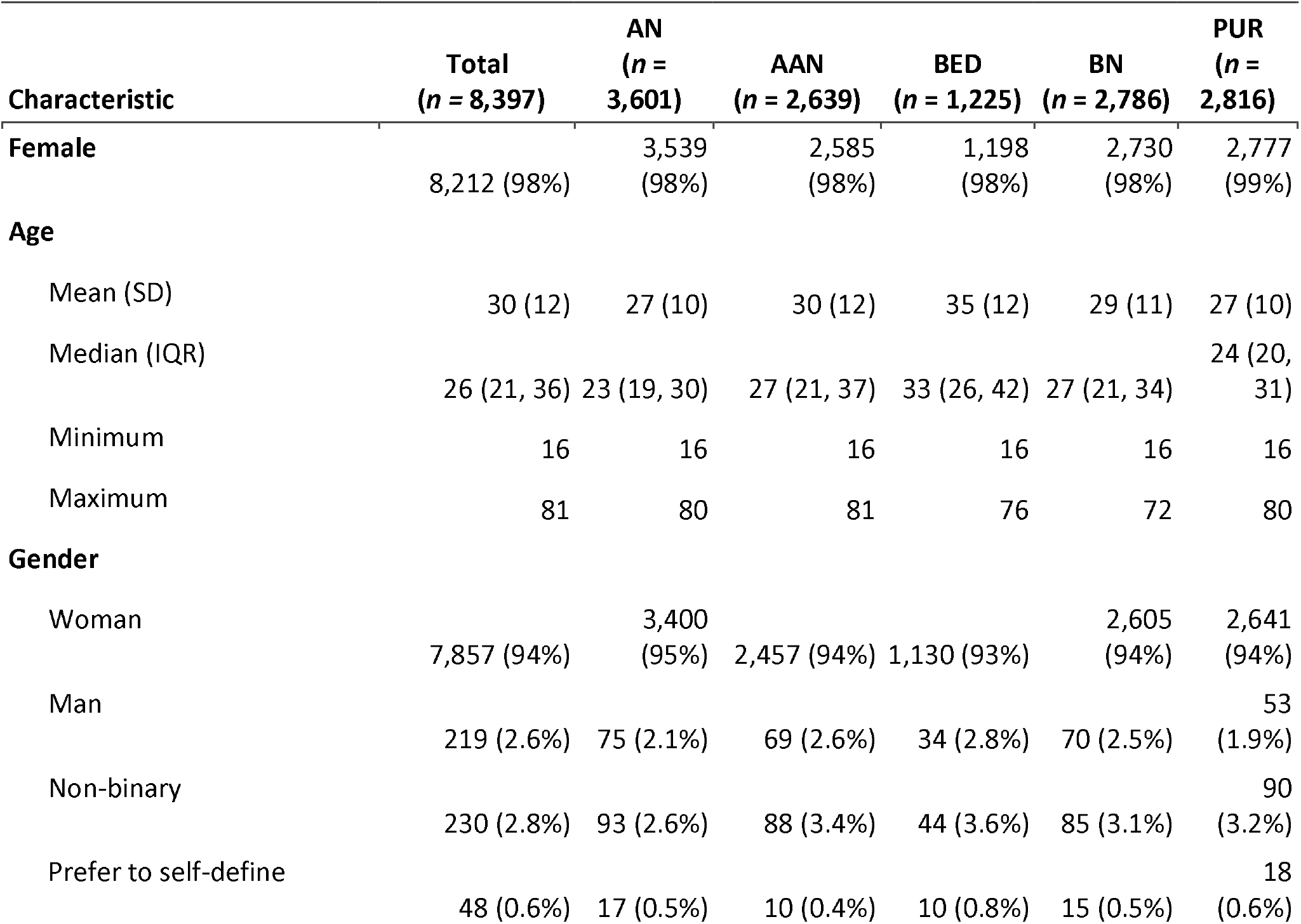
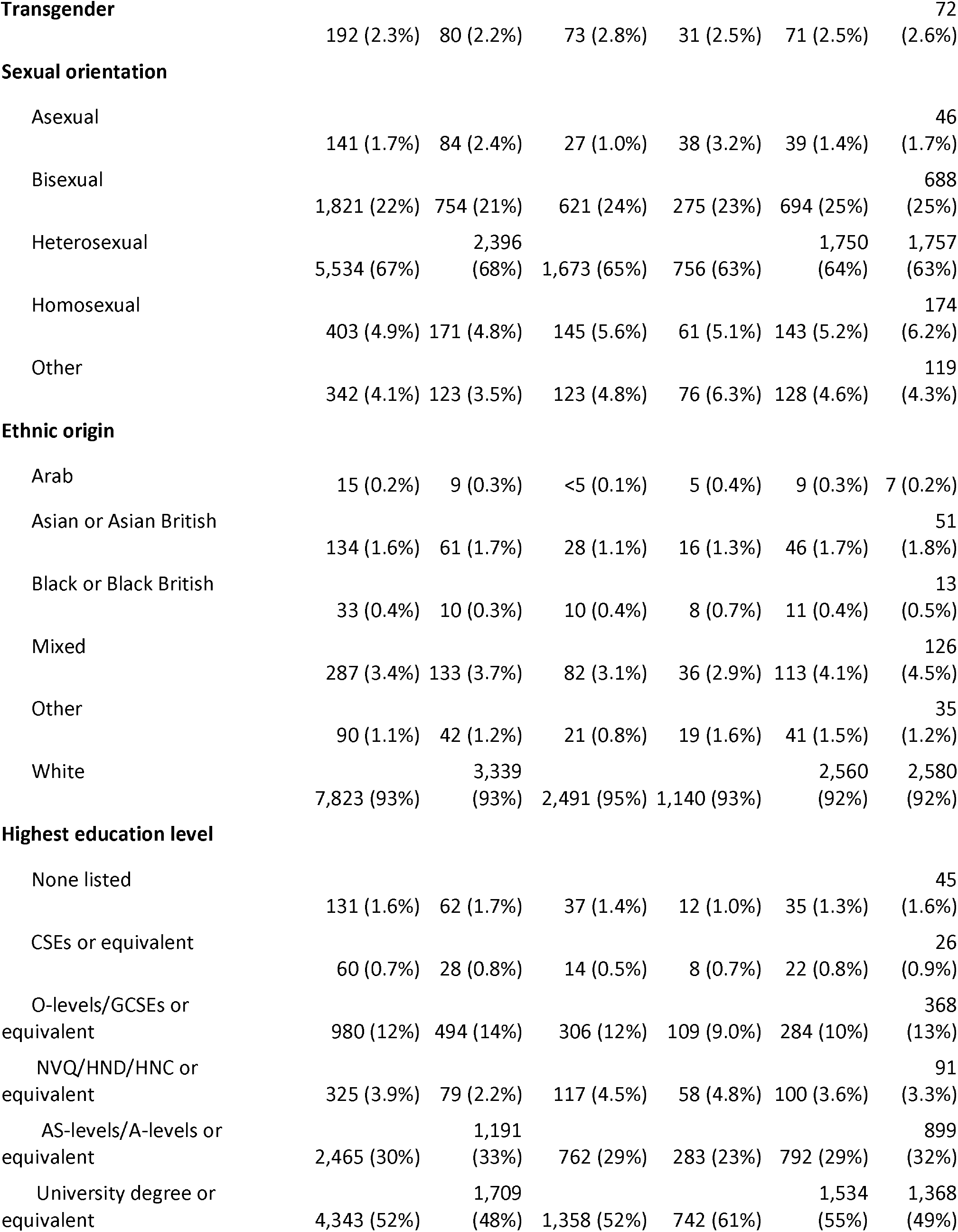
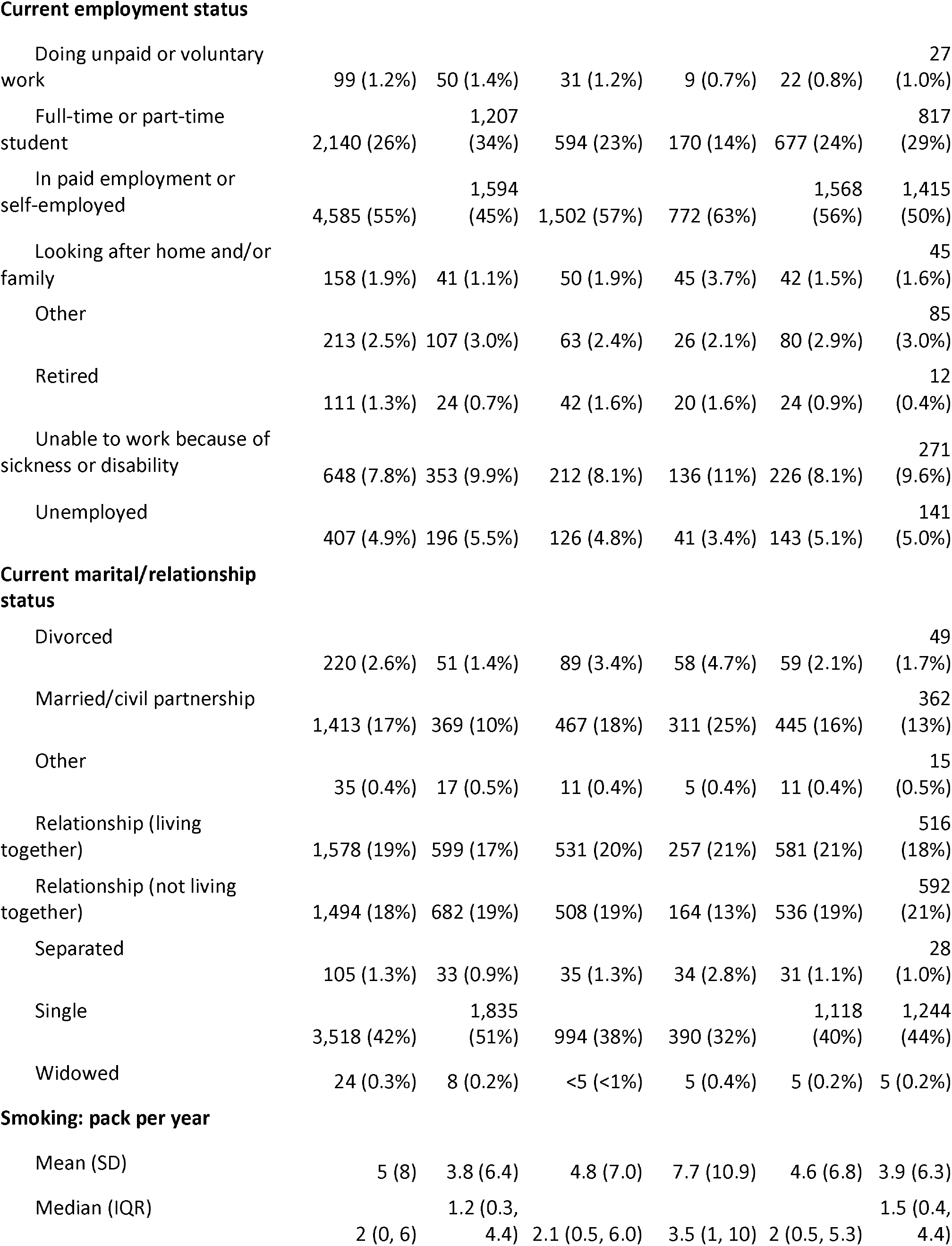
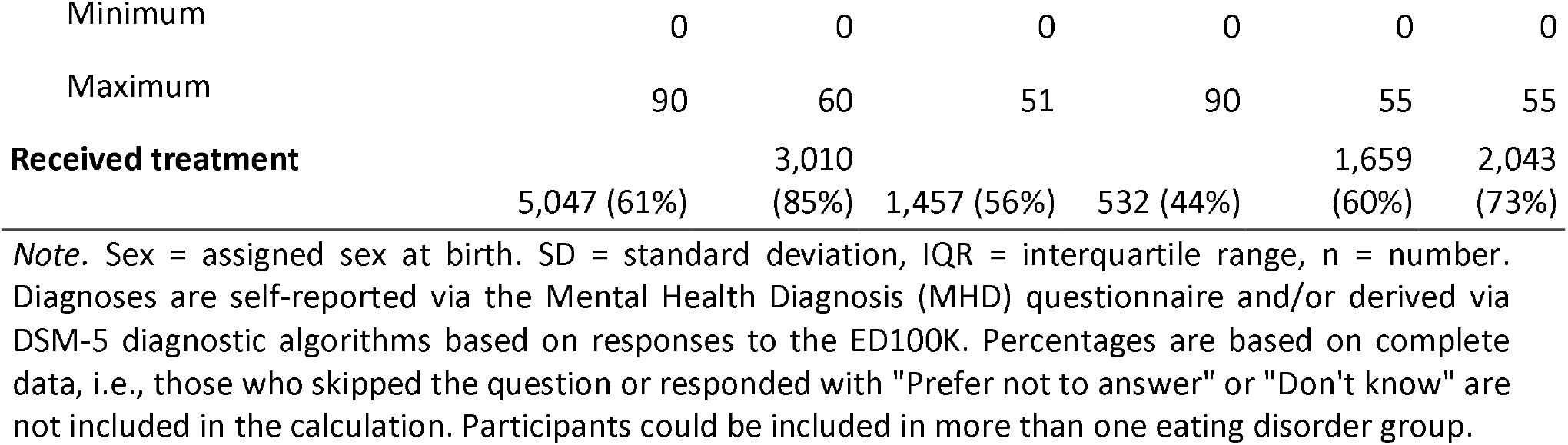
Characteristics of the Eating Disorders Genetics Initiative United Kingdom (EDGI UK) survey participants. The total sample (n = 8,397) and each subsample defined by diagnosis: anorexia nervosa (AN; n = 3,601), atypical anorexia nervosa (AAN; n = 2,639), binge-eating disorder (BED; n = 1,225), bulimia nervosa (BN; n = 2,786), and purging disorder (PUR; n = 2,816), September 2022. Diagnoses are self-reported via the Mental Health Diagnosis (MHD) questionnaire and/or derived via The Diagnostic and Statistical Manual of Mental Disorders, Fifth Edition (DSM-5) diagnostic algorithms based on responses to the ED100K.

**Table 2.**
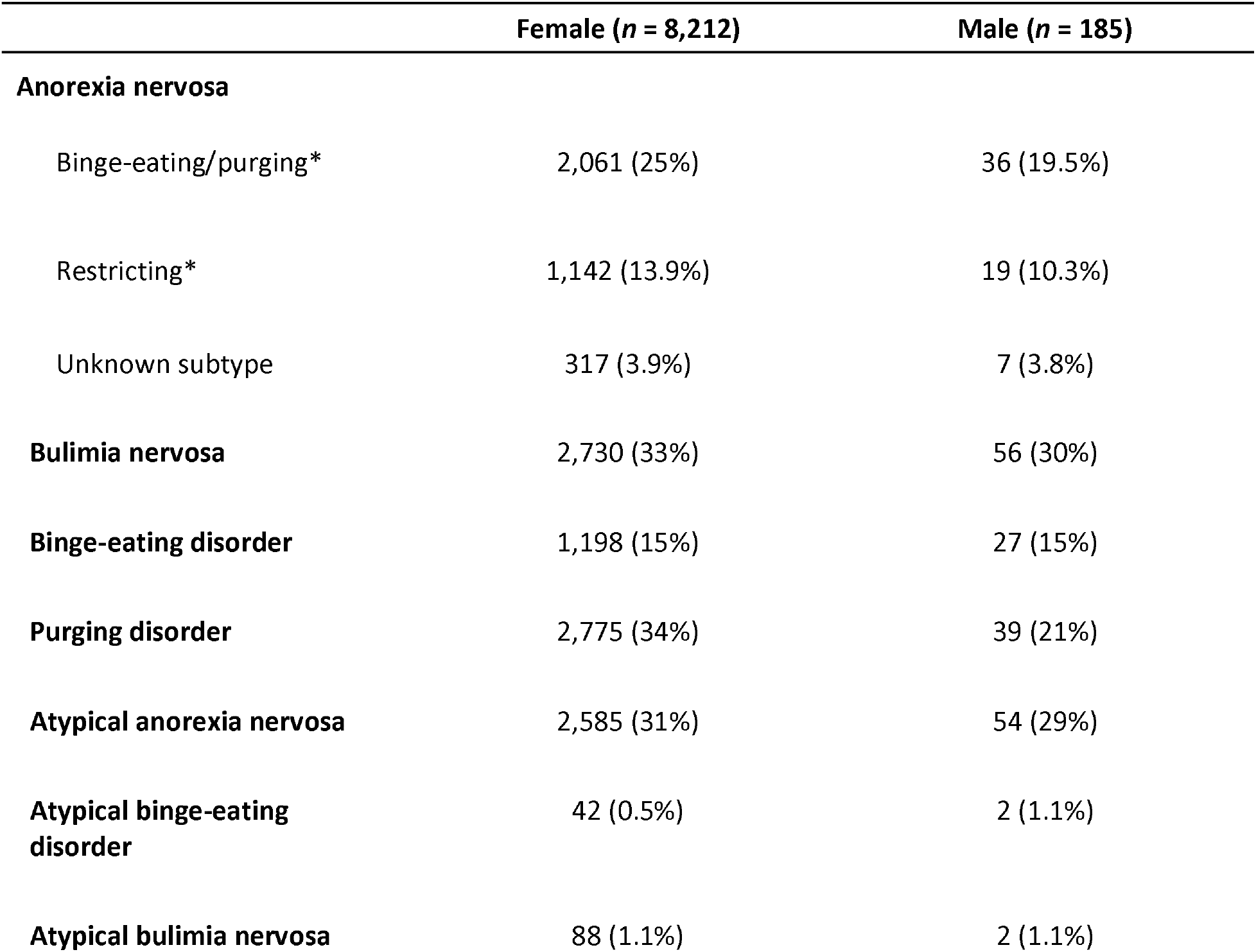
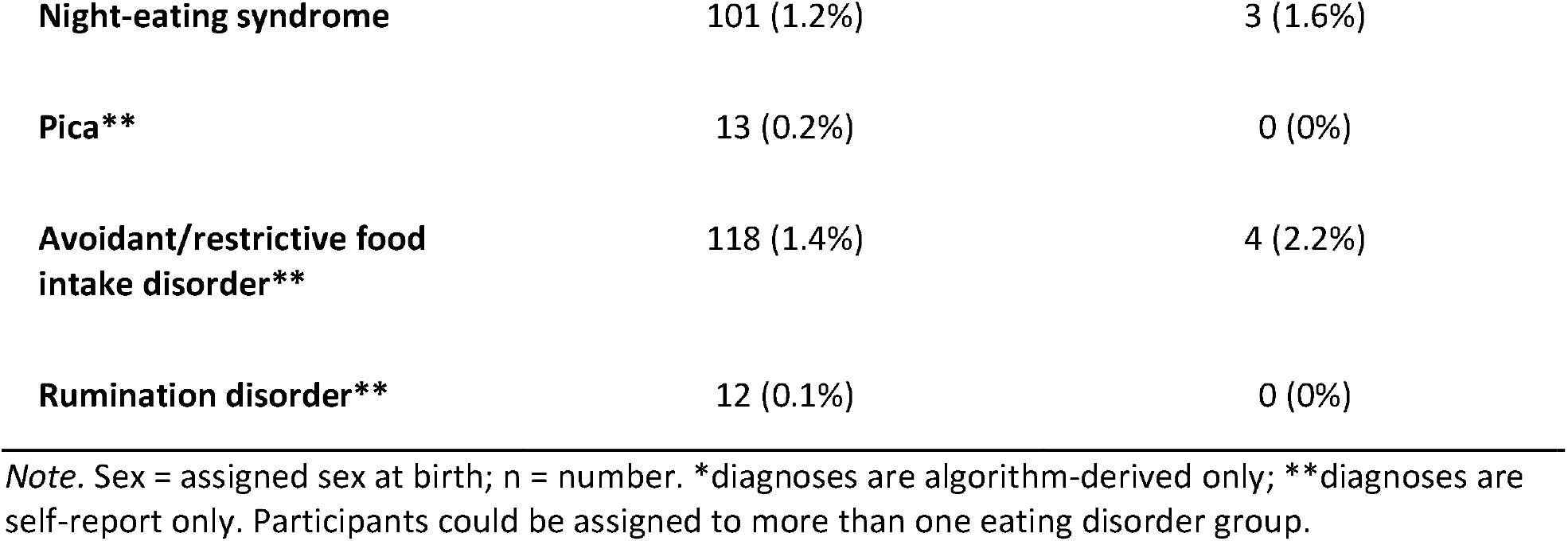
Lifetime self-reported eating disorder diagnoses in the Eating Disorders Genetics Initiative United Kingdom (EDGI UK) survey participants, split by biological sex, as of September 2022. Diagnoses are self-reported via the Mental Health Diagnosis (MHD) questionnaire and/or derived via the Diagnostic and Statistical Manual of Mental Disorders, Fifth Edition (DSM-5) diagnostic algorithms based on responses to the ED100K.

|  | **Female (n = 8,212)** | **Male (n = 185)** |
| --- | --- | --- |
| **Anorexia nervosa** |  |  |
| Binge-eating/purging* | 2,061 (25%) | 36 (19.5%) |
| Restricting* | 1,142 (13.9%) | 19 (10.3%) |
| Unknown subtype | 317 (3.9%) | 7 (3.8%) |
| **Bulimia nervosa** | 2,730 (33%) | 56 (30%) |
| **Binge-eating disorder** | 1,198 (15%) | 27 (15%) |
| **Purging disorder** | 2,775 (34%) | 39 (21%) |
| **Atypical anorexia nervosa** | 2,585 (31%) | 54 (29%) |
| **Atypical binge-eating disorder** | 42 (0.5%) | 2 (1.1%) |
| **Atypical bulimia nervosa** | 88 (1.1%) | 2 (1.1%) |
| **Night-eating syndrome** | 101 (1.2%) | 3 (1.6%) |
| **Pica\*\*** | 13 (0.2%) | 0 (0%) |
| **Avoidant/restrictive food intake disorder\*\*** | 118 (1.4%) | 4 (2.2%) |
| **Rumination disorder\*\*** | 12 (0.1%) | 0 (0%) |

*Note.* Sex = assigned sex at birth; n = number. \*diagnoses are algorithm-derived only; \*\*diagnoses are self-report only. Participants could be assigned to more than one eating disorder group.

**Table 3.** Psychiatric comorbidities in the Eating Disorders Genetics Initiative United Kingdom (EDGI UK) survey participants. Each subsample is defined by diagnosis: anorexia nervosa (AN; n = 3,601), atypical anorexia nervosa (AAN; n = 2,639), binge-eating disorder (BED; n = 1,225), bulimia nervosa (BN; n = 2,786), and purging disorder (PUR; n = 2,816), as of September 2022.

|  | **AN**<br>**($n = 3,601$)** | **AAN**<br>**($n = 2,639$)** | **BED**<br>**($n = 1,225$)** | **BN**<br>**($n = 2,786$)** | **PUR**<br>**($n = 2,816$)** |
| --- | --- | --- | --- | --- | --- |
| **Depression and anxiety** |  |  |  |  |  |
| No depressive or anxiety disorder | 643 (19%) | 417 (17%) | 132 (11%) | 454 (17%) | 395 (15%) |
| Depressive and anxiety disorder | 1,836 (55%) | 1,459 (59%) | 796 (67%) | 1,521 (58%) | 1,619 (61%) |
| Only depressive disorder | 520 (15%) | 370 (15%) | 188 (16%) | 431 (16%) | 403 (15%) |
| Only anxiety disorder | 366 (11%) | 236 (9.5%) | 72 (6.1%) | 228 (8.7%) | 233 (8.8%) |
| **Depressive disorder/s** | 2,357 (70%) | 1,829 (74%) | 985 (83%) | 1,953 (74%) | 2,024 (76%) |
| **Anxiety disorder/s** | 2,202 (65%) | 1,695 (68%) | 868 (73%) | 1,749 (66%) | 1,852 (70%) |
| **PTSD** | 542 (15%) | 412 (16%) | 248 (20%) | 438 (16%) | 508 (18%) |
| **Obsessive compulsive disorder/s** | 867 (61%) | 518 (58%) | 224 (65%) | 626 (61%) | 739 (69%) |
| **Personality disorder** | 465 (13%) | 321 (12%) | 214 (18%) | 416 (15%) | 433 (16%) |
| **Bipolar disorder and schizophrenia** |  |  |  |  |  |
| No psychotic or bipolar disorder | 3,098 (93%) | 2,282 (93%) | 1,047 (89%) | 2,391 (92%) | 2,404 (91%) |
| Psychotic and bipolar disorder | 39 (1.2%) | 24 (1.0%) | 20 (1.7%) | 28 (1.1%) | 38 (1.4%) |
| Only psychotic disorder | 84 (2.5%) | 48 (2.0%) | 28 (2.4%) | 60 (2.3%) | 73 (2.8%) |
| Only bipolar disorder | 110 (3.3%) | 104 (4.2%) | 79 (6.7%) | 123 (4.7%) | 117 (4.4%) |
| **Psychotic disorders** | 125 (3.5%) | 72 (2.8%) | 49 (4.1%) | 88 (3.2%) | 112 (4.0%) |
| **Bipolar disorder** | 150 (4.2%) | 128 (4.9%) | 99 (8.1%) | 152 (5.5%) | 155 (5.5%) |
| **ASD** | 246 (7.0%) | 138 (5.3%) | 71 (5.8%) | 140 (5.1%) | 167 (6.0%) |
| **ADHD** | 185 (5.1%) | 150 (5.7%) | 134 (11%) | 175 (6.3%) | 155 (5.5%) |

*Note.* Psychiatric comorbidities are self-reported via the Mental Health Diagnosis (MHD) questionnaire. Percentages are based on complete data, i.e., those who skipped the question or responded with "Prefer not to answer" or "Don't know" are not included in the calculation. Participants could be assigned to more than one eating disorder group.
Abbreviations: ASD = autism spectrum disorder, ADHD = attention deficit hyperactivity disorder, PTSD = post-traumatic stress disorder.

**Table 4.**
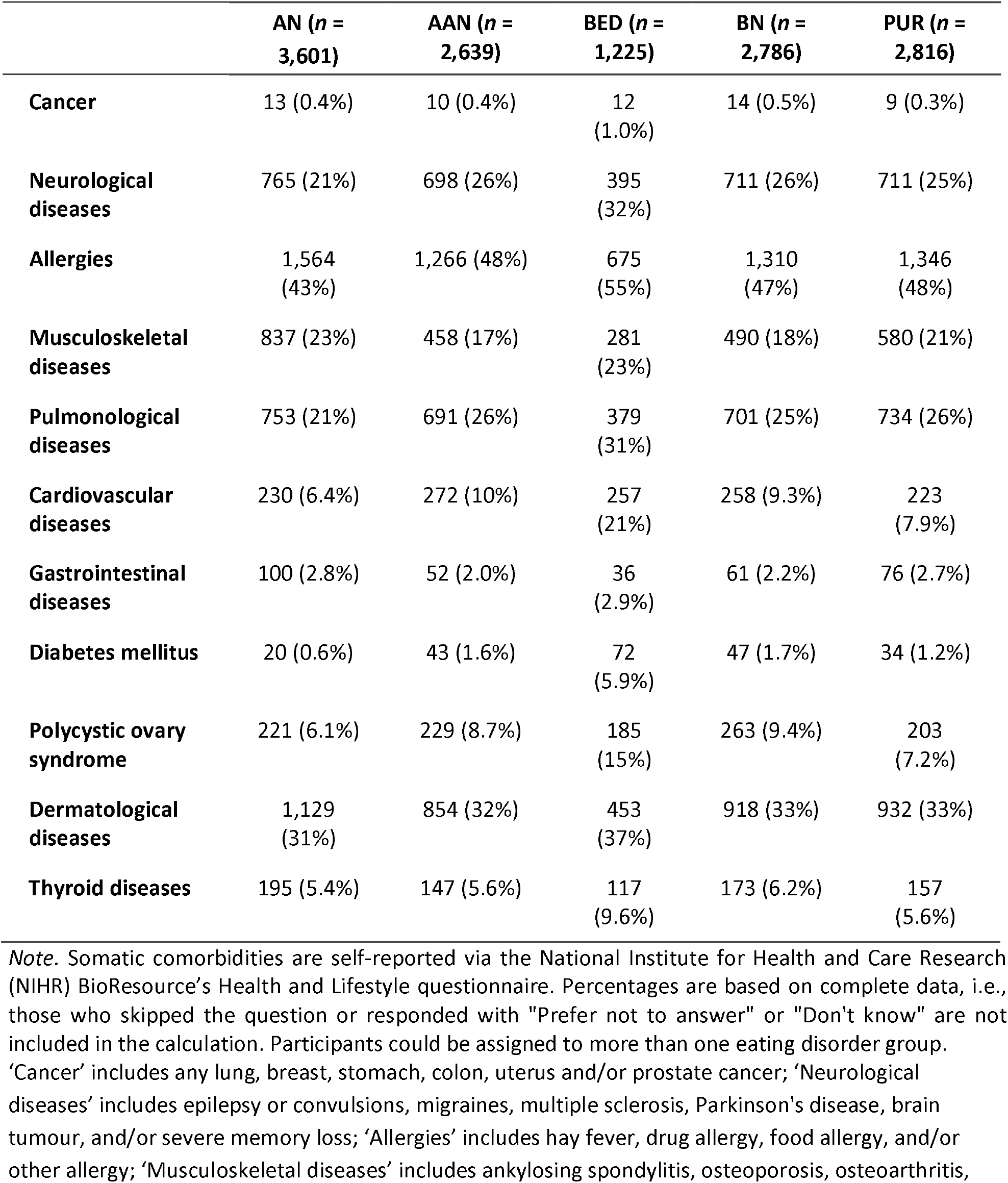
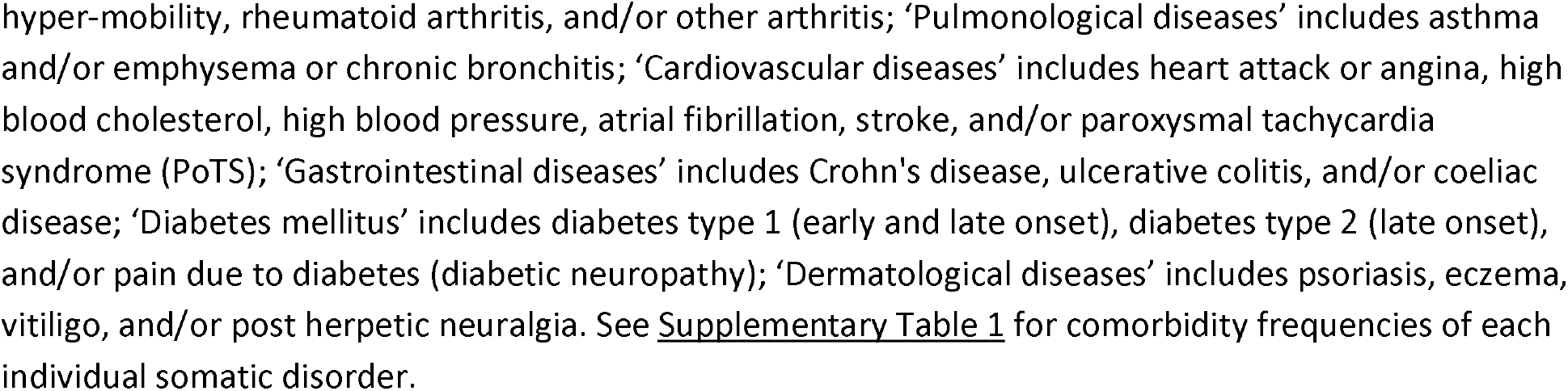
Somatic comorbidities in the Eating Disorders Genetics Initiative United Kingdom (EDGI UK) survey participants. Each subsample is defined by diagnosis: anorexia nervosa (AN; n = 3,601), atypical anorexia nervosa (AAN; n = 2,639), binge-eating disorder (BED; n = 1,225), bulimia nervosa (BN; n = 2,786), and purging disorder (PUR; n = 2,816), as of September 2022.

|  | **AN ($n = 3,601$)** | **AAN ($n = 2,639$)** | **BED ($n = 1,225$)** | **BN ($n = 2,786$)** | **PUR ($n = 2,816$)** |
| --- | --- | --- | --- | --- | --- |
| **Cancer** | 13 (0.4%) | 10 (0.4%) | 12 (1.0%) | 14 (0.5%) | 9 (0.3%) |
| **Neurological diseases** | 765 (21%) | 698 (26%) | 395 (32%) | 711 (26%) | 711 (25%) |
| **Allergies** | 1,564 (43%) | 1,266 (48%) | 675 (55%) | 1,310 (47%) | 1,346 (48%) |
| **Musculoskeletal diseases** | 837 (23%) | 458 (17%) | 281 (23%) | 490 (18%) | 580 (21%) |
| **Pulmonological diseases** | 753 (21%) | 691 (26%) | 379 (31%) | 701 (25%) | 734 (26%) |
| **Cardiovascular diseases** | 230 (6.4%) | 272 (10%) | 257 (21%) | 258 (9.3%) | 223 (7.9%) |
| **Gastrointestinal diseases** | 100 (2.8%) | 52 (2.0%) | 36 (2.9%) | 61 (2.2%) | 76 (2.7%) |
| **Diabetes mellitus** | 20 (0.6%) | 43 (1.6%) | 72 (5.9%) | 47 (1.7%) | 34 (1.2%) |
| **Polycystic ovary syndrome** | 221 (6.1%) | 229 (8.7%) | 185 (15%) | 263 (9.4%) | 203 (7.2%) |
| **Dermatological diseases** | 1,129 (31%) | 854 (32%) | 453 (37%) | 918 (33%) | 932 (33%) |
| **Thyroid diseases** | 195 (5.4%) | 147 (5.6%) | 117 (9.6%) | 173 (6.2%) | 157 (5.6%) |

*Note.* Somatic comorbidities are self-reported via the National Institute for Health and Care Research (NIHR) BioResource's Health and Lifestyle questionnaire. Percentages are based on complete data, i.e., those who skipped the question or responded with "Prefer not to answer" or "Don't know" are not included in the calculation. Participants could be assigned to more than one eating disorder group. 'Cancer' includes any lung, breast, stomach, colon, uterus and/or prostate cancer; 'Neurological diseases' includes epilepsy or convulsions, migraines, multiple sclerosis, Parkinson's disease, brain tumour, and/or severe memory loss; 'Allergies' includes hay fever, drug allergy, food allergy, and/or other allergy; 'Musculoskeletal diseases' includes ankylosing spondylitis, osteoporosis, osteoarthritis,

**Figure 4.**
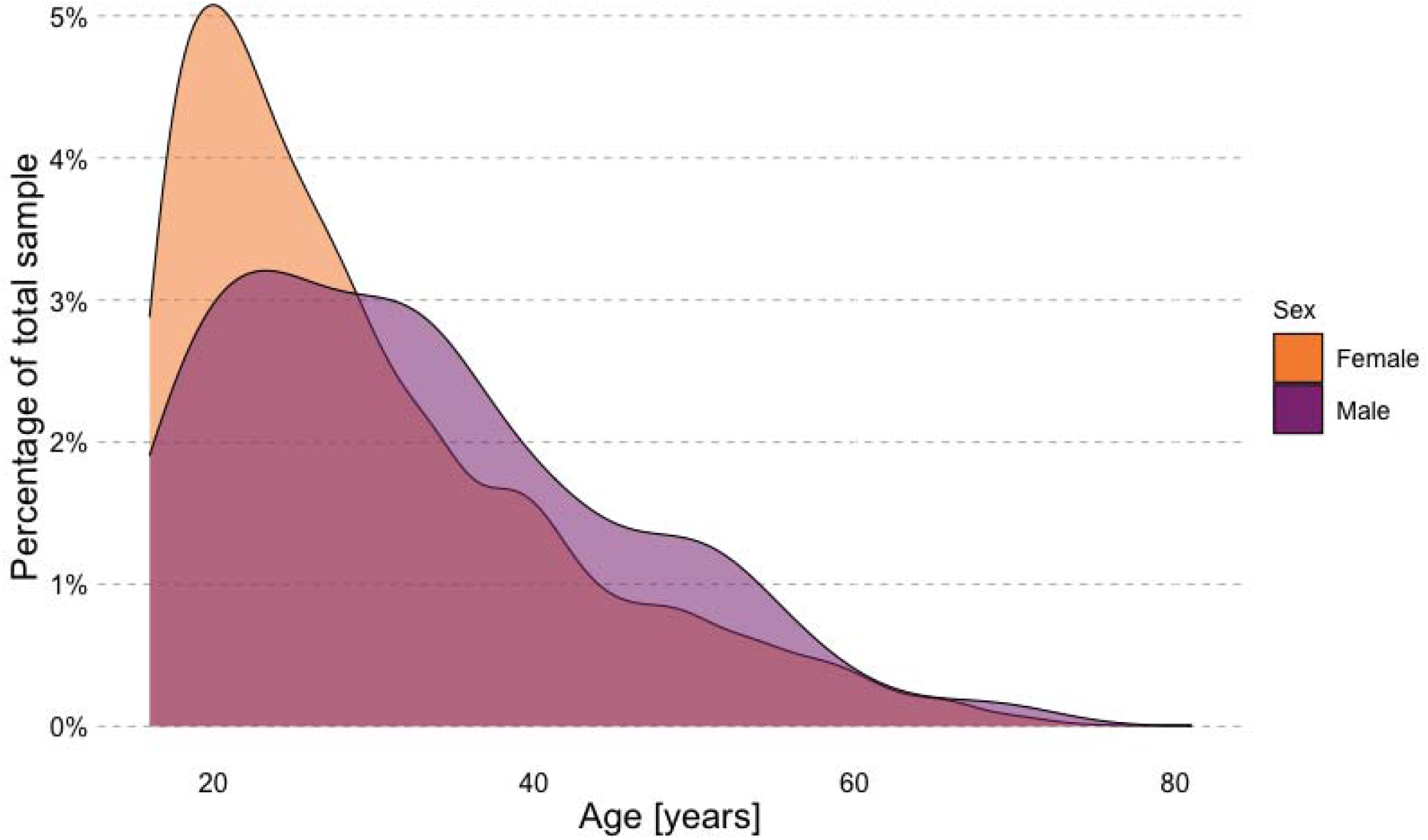
Age distribution of sample. Density plot illustrating the age distribution of the Eating Disorders Genetics Initiative United Kingdom (EDGI UK) survey participants (n = 8,397) stratified by sex assigned at birth (female n = 8,212; male n = 185), as of September 2022.

### 3.2 Psychiatric comorbidities

Over half of EDGI UK survey participants report a lifetime diagnosis of depression and anxiety, with the highest proportion in those with binge-eating disorder (67%). Other highly endorsed psychiatric comorbidities include OCD (58-69%), posttraumatic stress disorder (15-20%), and personality disorder (12-18%). More participants with binge-eating disorder endorsed ADHD (11%), bipolar disorder (8.1%), and a personality disorder (18%), than people with other eating disorders, such as anorexia nervosa, with 5.1%, 4.2%, and 13%, respectively.

### 3.3 Somatic comorbidities

Approximately half of EDGI UK survey participants also report an allergy (43-55%), and approximately one quarter report a lung and/or breathing problem (21-31%). Diabetes (5.9%), polycystic ovary syndrome (15%), and thyroid disease (9.6%) are most highly endorsed in those with binge-eating disorder.

### 3.4 Completed research and policy work

Based on data from EDGI UK, we have already finalised a number of reports. In collaboration with Beat, we published “When do eating disorders start? An investigation into two large UK samples”, showing that low weight, purging, and binge eating start as frequently over the age of 18 as under the age of 18 (*Eating Disorders Are Just as Likely to Start in Adulthood as Childhood, EDGI Report Finds*, n.d.). Professor Gerome Breen and Dave Chawner discussed these findings on Sky News, highlighting the need for investment into adult NHS eating disorder services which receive substantially less funding than child and adolescent services (Parliamentary and Health Service Ombudsman, 2017). During early 2021, Professor Gerome Breen, alongside other experts and volunteers, had several meetings with the UK government to discuss eating disorder research and treatment needs to foster policy changes. Initial results from the COPING study, which included a subset of EDGI UK survey participants, suggested that individuals with psychiatric disorders and those who were more worried about the pandemic were more likely to experience new onset of binge eating or low weight during the pandemic (H. L. Davies et al., 2022). Other findings indicated that being female, younger, or unemployed/a student were linked with experiencing worsening depression, anxiety, or PTSD symptoms during the pandemic (Young et al., 2021).

## 4. Discussion

Despite the impact of COVID-19 throughout 2020-2022 on EDGI UK’s recruitment plans, our targets were largely met. By September 2022, 8,397 eligible participants completed the sign-up questionnaire and of those, 4,294 returned their saliva kits. Most often, volunteers reported a lifetime diagnosis of anorexia nervosa, atypical anorexia nervosa, bulimia nervosa, and/or purging disorder. Comorbid psychiatric and somatic illnesses were highly prevalent. Over half of participants self-reported depression and/or anxiety disorders, and OCD and posttraumatic stress disorder were also common. Regarding somatic comorbidities, participants most often reported suffering from allergies, dermatological, and pulmonological diseases. Social media was a valuable recruitment method, with over half of the participants reporting they heard about the study from social media. As an online study with postal sampling, individuals are able to take part at their convenience from home. This was particularly advantageous during the COVID-19 pandemic. Overall, EDGI UK is optimally equipped to create a large and comprehensive resource for future collaboration in eating disorders research.

Large parts of the assessment battery were based on the clinical Swedish eating disorder quality registers Stepwise and RIKSÄT (Birgegård et al., 2010; Groß et al., 2013), in which most symptoms are assessed in clinical interviews and jointly registered by the patient and medical professional using an online-guided assessment. In addition to an assessment at the beginning of treatment, the registers track long-term clinical outcomes that have been included in genetic studies (Johansson et al., 2022). This will also be possible in EDGI UK. In contrast to EDGI UK, Stepwise only includes individuals for whom an intention to treat has been established, potentially limiting the sample to more severe cases. Whilst EDGI UK does not require individuals to have received treatment or a clinical diagnosis to participate, percentages of registered individuals in Stepwise with anorexia (27-40%) and bulimia nervosa (9-29%) were similar to those in EDGI UK.

The distribution of eating disorders in EDGI UK does not reflect prevalences measured in the underlying population (Micali et al., 2017). Epidemiological studies suggest that binge-eating disorder is more prevalent than anorexia nervosa (Kessler et al., 2013), but anorexia nervosa is most common in EDGI UK. The GLAD Study, our sister project, was more successful in recruiting people with binge-eating disorder than EDGI UK, probably due to the participants’ experience of comorbid depression and binge-eating disorder (M. R. Davies et al., 2019). We hypothesise that participants with comorbid depression and binge-eating disorder identify more with their depression diagnosis, hence tend to sign up for the GLAD Study rather than EDGI UK. A further barrier to those with binge-eating disorder joining EDGI UK may be the stigma and shame surrounding their disorder (Ali et al., 2017; Hepworth & Paxton, 2007), and the lack of recognition of it as a psychiatric disorder amongst both the public (Reas, 2017) and healthcare professionals (Kornstein et al., 2016). The GLAD Study launched 18 months prior to EDGI UK, and this earlier start may have contributed to this recruitment effect. However, as both EDGI UK and the GLAD Study are part of the NIHR BioResource, datasets can be combined for joint analysis due to overlap in phenotypic measures.

### 4.1 Limitations

EDGI UK is a selected sample that comprises far more females than the general English population (98% vs. 51%) (*Population and Household Estimates, England and Wales: Census 2021*, 2021). This in all likelihood reflects a recruitment effect, as it is suggested that ∼25% of individuals with an eating disorder are male (Sweeting et al., 2015). We have been more successful in recruiting white females than other members of the population. A factor complicating recruitment is that males often do not recognise that they have an eating disorder and hence are undiagnosed (Strother et al., 2012). Moreover, EDGI UK is less diverse than the English and Welsh general population (93% vs 84% white) (*Population Estimates by Ethnic Group and Religion, England and Wales: 2019*, 2019), limiting our investigations in minoritised ethnic and racial groups.

Internet access is necessary to take part. This may exclude individuals who cannot afford a device with access or have no experience in using computers. Participants without internet access are advised to visit their local library, ask a trusted family or a friend, or visit a participating NHS Trust. As EDGI UK is mostly advertised via social media content, older individuals may be less exposed to advertising.

Thus far, the ED100K, which assesses eligibility, has only been validated for anorexia nervosa in US-American and Australian samples (Thornton et al., 2018). We have successfully obtained ethical approval at King’s College London to further validate the ED100K questionnaire in inpatients and outpatients. Moreover, the NIAS, used to assess ARFID symptoms, does not include items measuring malnutrition, general practitioner (GP) visits, or social impairment, complicating the assignment of an algorithm-derived ARFID diagnosis. To overcome this limitation, we included the Pica, ARFID, and Rumination Disorder Interview Questionnaire (PARDI-AR-Q; (Bryant-Waugh et al., 2019) for participants who met the cutoff of 24 points on the NIAS.

### 4.2 Future directions

EDGI UK has a clear roadmap for improvement. First, we will work to diversify the racial and ethnic groups represented in the EDGI UK community. With ethical approval to conduct public and patient involvement events including community outreach, we will seek to understand and, ultimately, break down barriers that prevent these underrepresented communities from participating. We will also tailor advertising and recruitment strategies to minoritised ethnic and racial groups. Second, we aim to better understand participation and attrition biases, for example, through polygenic score analyses. Indeed, previous research has shown that polygenic scores for agreeableness, openness, and higher education are associated with continued participation in research studies (Taylor et al., 2018). Third, we will widen the range of participants by recruiting young persons under the age of 16 and by broadening our geographical recruitment area by making EDGI UK available in Northern Ireland, Scotland, and Wales. Fourth, we will seek to motivate more males to take part through targeted advertising and outreach. Fifth, we aim to extend clinical recruitment by collaborating with additional NHS Trusts, including FREED services and eating disorder clinics. Ultimately, EDGI UK is a unique resource contributing substantially to international genetics research in eating disorders. With our goal of recruiting 10,000 participants, we are at the forefront of understanding the genetic and environmental influences on eating disorders.

## Supporting information

EDGI UK Sign-up Questionnaire

EDGI UK Optional Questionnaires

## Data Availability

EDGI UK data are not publicly available however are available via a data request application to the NIHR BioResource (https://bioresource.nihr.ac.uk/using-our-bioresource/academic-and-clinical-researchers/apply-for-bioresource-data/). All code for this study is publicly available: https://github.com/tnggroup/EDGI_protocol.

https://bioresource.nihr.ac.uk/using-our-bioresource/academic-and-clinical-researchers/apply-for-bioresource-data/

https://github.com/tnggroup/EDGI_protocol

## Acknowledgements

We thank NIHR BioResource volunteers for their participation, and gratefully acknowledge NIHR BioResource centres, NHS Trusts and staff for their contribution. We thank the National Institute for Health and Care Research, NHS Blood and Transplant, and Health Data Research UK as part of the Digital Innovation Hub Programme. The views expressed are those of the author(s) and not necessarily those of the NHS, the NIHR or the Department of Health and Social Care. We especially thank all participants of the EDGI UK community and all the people who helped us to promote the study.

