## Supplementary information for "The Eating Disorders Genetics Initiative (EDGI) United Kingdom"

### Supplementary Material

Dina Monssen*[1,2(0000-0003-0080-0799)], Helena L Davies*[1(0000-0002-9419-1009)], Shannon Bristow[1,2(0000-0002-0896-781X)], Saakshi Kakar[1,2(0000-0003-1677-1857)], Susannah C B Curzons[1,2(0000-0001-9445-2910)], Molly R Davies[1,2(0000-0003-3483-9907)], Zain Ahmad[1(0000-0002-6940-8851)], John R Bradley[3(0000-0002-7774-8805)], Steven Bright[1(0000-0002-9408-3171)], Jonathan R I Coleman[1,2(0000-0002-6759-0944)], Kiran Glen[1,2(0000-0002-2831-3266)], Matthew Hotopf[2,4(0000-0002-3980-4466)], Emily J Kelly[1,2(0000-0002-0118-4994)], Abigail R ter Kuile[1,2(0000-0002-7869-3754)], Chelsea Mika Malouf[1,2(0000-0002-5564-7464)], Gursharan Kalsi[1,2(0000-0002-5156-7176)], Nathalie Kingston[3,5(0000-0002-9190-2231)], Monika McAtarsney-Kovacs[1,2(0000-0002-2599-5249)], Jessica Mundy[1(0000-0001-5513-8902)], Alicia J Peel[1(0000-0002-6144-5412)], Alish B Palmos[1,2(0000-0001-5748-6652)], Henry C Rogers[1,2(0000-0003-2531-7496)], Megan Skelton[1,2(0000-0002-7619-349X)], Brett N Adey[1,2(0000-0003-4356-4079)], Sang Hyuck Lee[1,2(0000-0002-5610-033X)], Hope Virgo, Tom Quinn[8], Tom Price[4,12(0000-0002-2716-1475)], Johan Zvrskovec[1,2(0000-0002-8862-0874)], Thalia C Eley[1,2(0000-0001-6458-0700)], Janet Treasure[4(0000-0003-0871-4596)], Christopher Hübel**[1,2,7(0000-0002-1267-8287)], Gerome Breen**[1,2 (0000-0003-2053-1792)]

1. Social, Genetic and Developmental Psychiatry Centre, Institute of Psychiatry, Psychology, and Neuroscience, King's College London, London, UK
2. UK National Institute for Health and Care Research (NIHR) Maudsley Biomedical Research Centre, South London and Maudsley NHS Foundation Trust, London, UK
3. NIHR BioResource, Cambridge University Hospitals NHS Foundation, Cambridge Biomedical Campus, Cambridge, UK
4. Department of Psychological Medicine, Institute of Psychiatry, Psychology and Neuroscience, King's College London, London, United Kingdom
5. Department of Haematology, University of Cambridge, Cambridge Biomedical Campus, Cambridge, UK
6. Section of Eating Disorders, Department of Psychological Medicine, Institute of Psychiatry, Psychology and Neuroscience, King's College London, London, UK
7. National Centre for Register-based Research, Aarhus Business and Social Sciences, Aarhus University, Aarhus, Denmark Beat Eating Disorders, Unit 1, Chalk Hill House, 19 Rosary Road, Norwich, Norfolk, NR1 1SZ
8. Eating Disorders Outpatient Service, South London and Maudsley NHS Foundation Trust, London, UK

*shared first authorship
**shared last authorship

### Table of Contents

Supplementary Material 1. Online news articles **3**

Supplementary Material 2: Sign-up Questionnaire **10**

Supplementary Material 3. Optional Questionnaires **11**

Supplementary Material 4. EDGI UK Saliva Instruction flyer **13**

Supplementary Figure 1. Plot showing the overlap in lifetime eating disorder diagnoses **14**

Supplementary Table 1. Detailed table of somatic diseases **15**

References **19**

**Supplementary Material 1. Online news articles**

Online news articles about the launch of EDGI UK between February, 2020 and March 2020.

**United Kingdom's largest eating disorder charity, Beat:**
https://www.beateatingdisorders.org.uk/news/beat-news/largest-study-eating-disorders/;
Retrieved 24.10.2022

#### Largest ever study of eating disorders launches in England

Researchers at King's College London have launched the largest ever study into eating disorders. Partnering with the National Institute for Health Research (NIHR) BioResource and the eating disorder charity Beat, they aim to recruit at least 10,000 people in England who have experienced an eating disorder at some point in their life to a pioneering new study that aims to unlock the secrets of eating disorders.

The Eating Disorders Genetics Initiative (EDGI) will help researchers better understand these conditions and enable the design of new treatments aimed at improving the lives of patients. EDGI will facilitate the discovery of new genetic and environmental risk factors and by creating a 'bank' of potential study participants who agree to be recontacted for further research, will speed up the pace of research into the most under-researched set of psychiatric disorders.

Geneticist and study lead, Professor Gerome Breen, NIHR Maudsley Biomedical Research Centre, Institute of Psychiatry, Psychology & Neuroscience, King's College London, said, "With EDGI, we hope to discover new genetic and environmental risk factors and provide a platform that will increase the amount of research being done in the field. We want to make research into eating disorders faster, cheaper and more effective to meet desperate need for more effective treatments."

Psychiatrist and clinical lead, Professor Janet Treasure, Institute of Psychiatry, Psychology & Neuroscience, King's College London said "We want to recruit participants across the whole range of eating disorders; we want to understand common risk factors and how to develop both general and specific treatments for these serious and life threatening conditions."

Beat's Chief Executive, Andrew Radford, said: 'It's become increasingly clear that there are genetic factors involved in eating disorders and this crucial study will help to further our understanding and knowledge of these complex mental illnesses. It is particularly heartening that this study will cover all eating disorder diagnoses, including those where there is a serious lack of research. We hope that studies such as these can lead to more tailored treatments for eating disorders, and in time prevent them from developing in the first place."

Hope Virgo, a mental health advocate and campaigner for eating disorder awareness comments, "I am absolutely delighted to be supporting EDGI because research into eating disorders is crucial. Eating disorders completely take over lives, and yet there is still a lack of understanding around treatment, prevention and support. That is why EDGI is so important to take this one step further."

Shanel, an Ambassador for Beat notes: “Having suffered with an eating disorder for 10 years and now fully recovered, I do not wish the experience on anyone. Instead, I support more research into the field and why the EDGI is so valuable. I will be taking part in the study and hope others can join me in this tremendous opportunity for research.”

- Up to 5% of the population will experience an eating disorder. The most well known are bulimia nervosa, anorexia nervosa and binge-eating disorder, but the EDGI project is open to anybody who has experienced an eating disorder.
- Volunteers will be able to sign up online at edgiuk.org, where they'll be asked to complete a 15-20 minute online questionnaire and supply a saliva sample by post, which will be used to analyse their DNA.
- People who enrol will be entered into the NIHR Mental Health BioResource – which is part of the NIHR BioResource, a national resource of research volunteers who can be recontacted up to four times a year to take part in other research projects aimed at developing new treatments for, and understanding the causes of, both mental and physical illness. Crucially, this will speed up research in eating disorders.
- Eating disorders affect an estimated 1.25 million people in the UK. The most common are anorexia nervosa, bulimia nervosa and binge-eating disorder. Eating disorders are serious and have the highest mortality rate amongst all psychiatric disorders. Currently, less than half of individuals reach full recovery.
- Life with an eating disorder can be debilitating. Not only do eating disorders impact social relationships and quality of life, but they can also have devastating physical health effects on an individual. As such, researchers urgently need more people to take part in eating disorder research studies.
- Current research indicates a heritability of between 40-70% for eating disorders depending on the condition. By having a large, diverse group of people available for future studies, researchers hope to find the genetic and environmental risks that increase the risk of having an eating disorder and therefore how to develop more effective treatments.
- EDGI will also be launched in other countries around the world this year, including New Zealand, Australia and the USA but it launches first in England.

EDGI, funded in England by the National Institute for Health Research (NIHR) BioResource. The NIHR BioResource is a national resource of (currently) over 150,000 people - with and without health problems - who are willing to be approached to participate in research studies investigating the link between genes, the environment and health and disease. It is based at centres around England and is funded by the National Institute for Health Research.

The NIHR Mental Health BioResource is an arm of the NIHR BioResource, which aims to increase participation of people with mental health disorders in medical and psychological research. For EDGI, participants will also be recruited into the NIHR Mental Health BioResource.

Visit edgiuk.org to find out more.

**ITV News**
https://www.itv.com/news/2020-02-26/call-for-participants-for-largest-ever-study-of-eating-disorders
Retrieved 24. October 2022

#### Call for participants for largest ever study of eating disorders

- HEALTH
- EATING DISORDERS
- Wednesday 26 February 2020 at 11:00am

The largest ever study of eating disorders has been launched by scientists at King's College London in a bid to better understand the condition.

It's hoped the findings from the Eating Disorders Genetics Initiative (EDGI) will help design new treatments for patients.

Researchers are aiming to recruit at least 10,000 people across England who have experienced an eating disorder at some point in their life.

An estimated 1.25 million people in the UK are affected by eating disorders.

- More children admitted to hospital for eating disorders
- Teenage girls with anxiety potentially at greater risk of developing eating disorders, study finds

*A study by Bristol University found teenage girls with anxiety are potentially at greater risk of developing eating disorders. Credit: PA*

Those behind the project say it will look to identify "genetic and environmental risk factors" around disorders like anorexia nervosa, bulimia nervosa and binge-eating.

Eating disorders currently have the highest mortality rate among all psychiatric disorders and less than half of individuals reach full recovery according to the Anorexia & Bulimia Care organisation.

The study is the joint project of King's College London, the National Institute for Health Research, BioResource, and the eating disorder charity Beat.

The group is appealing for participants willing to join the study.

- Anorexia sufferer reveals how diet firm targeted her as advertising standards issue new guide for influencers

Geneticist and study lead, Professor Gerome Breen, said: "We want to make research into eating disorders faster, cheaper and more effective to meet desperate need for more effective treatments."

Volunteers can sign up online at edgiuk.org.

Potential participants will need to complete a 15-20 minute online questionnaire and supply a saliva sample by post.

*If you think you may be suffering from an eating disorder, the following links can provide support and useful advice:*

- NHS eating disorders information page
- Beat informationpage
- HEALTH
- EATING DISORDERS

**NIHR Bioresource**
https://bioresource.nihr.ac.uk/news/edgi-eating-disorders-genetics-initiative/
Retrieved 24.20.2022

### EDGI, largest ever study of eating disorders, launches in England

A study funded by the National Institute for Health Research aims to unlock the secrets of eating disorders. EDGI, the Eating Disorders Genetics Initiative, is seeking more than 10,000 participants.

A National Institute for Health Research (NIHR) funded study launched on 26 February 2020. This is a pioneering new study that aims to unlock the secrets of eating disorders. It aims to recruit at least 10,000 people in England who have experienced an eating disorder at some point in their life.

Partnering with the NIHR BioResource for Translational Research and the eating disorder charity Beat, the Eating Disorders Genetics Initiative (EDGI) will help researchers better understand these conditions and enable the design of new treatments aimed at improving the lives of patients.

EDGI will facilitate the discovery of new genetic and environmental risk factors. By creating a resource of potential study participants, who agree to be recontacted for further research, it will speed up the pace of research into the most under-researched set of psychiatric disorders.

See EDGI website here

*First published 26 February 2020*

**Pharma Times**
https://www.pharmatimes.com/news/largest_ever_eating_disorders_study_launched_in_uk_1327639
Retrieved 24. October

### Largest ever eating disorders study launched in UK

2nd March 2020

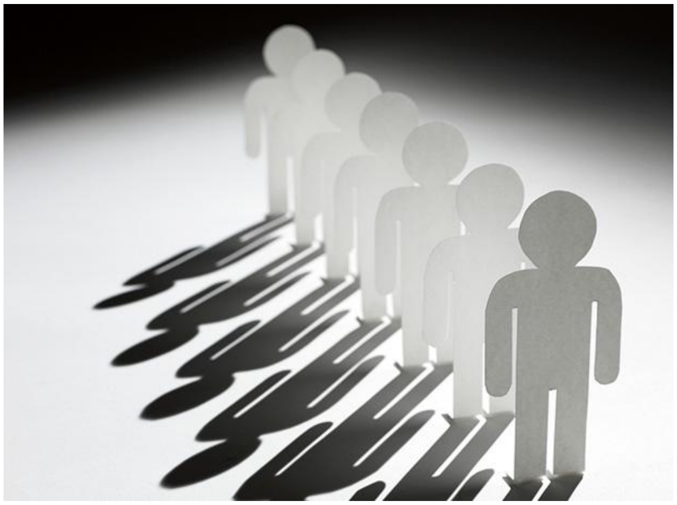

By Anna Smith

The National Institute for Health Research (NIHR) is recruiting 10,000 people in its latest "pioneering" study in to eating disorders.

The recruitment drive, which makes the study the "largest ever" eating disorder survey of its kind, aims to help researchers better understand these conditions and enable the design of new treatments aimed at improving the lives of patients.

Partnering with the NIHR BioResource for Translational Research and the eating disorder charity Beat, the Eating Disorders Genetics Initiative (EDGI) says that it will "facilitate the discovery of new genetic and environmental risk factors and by creating a resource of potential study participants who agree to be re-contacted for further research", in order to speed up the pace of research into the most under-researched set of psychiatric disorders.

Last year it was revealed that a "record number" of children are being hospitalised for eating disorders, with Public Health England reporting that hospital admissions for eating disorders in girls aged just 10 years old had increased by 146% since 2013/14, with a total of 2,196 hospital admissions for eating disorders of children and young people aged 10 to 24 years in 2017/18.

The report also stated that although bulimia is more common among children and young people, it is anorexia which accounts for the larger proportion of hospital admissions.

The new NIHR study hopes "to discover new genetic and environmental risk factors and provide a platform that will increase the amount of research being done in the field", according to Professor Gerome Breen, NIHR Maudsley Biomedical Research Centre.

Gerome continued, “We want to make research into eating disorders faster, cheaper and more effective to meet the desperate need for more effective treatments.”

Up to 5% of the population will experience an eating disorder, with the most well-known being anorexia nervosa, bulimia nervosa and binge-eating disorder.

Eating disorders are serious and have the highest mortality rate amongst all psychiatric disorders. Currently, less than half of individuals reach full recovery.

**Supplementary Material 2: Sign-up Questionnaire**

| EDGI UK Sign-up Questionnaire | |
|---|---|
| | NIHR BioResource Health and Lifestyle Questionnaire |
| | The ED100K Questionnaire (Thornton et al., 2018) |
| | Additional questions covering atypical anorexia nervosa |
| | 7-item Binge-Eating Disorder Screener (BEDS-7) (Herman et al., 2016) |
| | Night Eating Diagnostic Questionnaire (NEQ) (Gluck et al., 2001) |
| | Muscle Dysmorphic Disorder Inventory (MDDI) (Hildebrandt et al., 2004) |
| | Nine-Item Avoidant Restrictive Food Intake Disorder Screen (Zickgraf & Ellis, 2018) |
| | Pica, ARFID, and Rumination Disorder Interview Questionnaire (PARDI-AR-Q) (Bryant-Waugh et al., 2019) |
| | Childhood Trauma Screener (CTS-5) (Glaesmer et al., 2013) |
| | Adverse life experiences including domestic violence and catastrophic trauma (Davis et al., 2020) |
| | Post-Traumatic Stress Disorder Checklist (PCL-6) (Lang et al., 2012 |
| | Study feedback questionnaire |

**Supplementary Material 3. Optional Questionnaires**

Optional questionnaires offered to eligible EDGI UK participants once they have completed the sign up questionnaire, broken down in six categories.

| Module 1: Eating behaviours and physical activity | |
|---|---|
| | Yale Food Addiction Scale (Y-FAS) (Gearhardt et al., 2009) |
| | Adult Eating Behaviours Questionnaire (AEBQ) (Hunot et al., 2016) |
| | Clinical Impairment Assessment (CIA) (Jenkins, 2013) |
| | Eating Disorder Examination Questionnaire (EDE-Q6) (Carey et al., 2019) |
| | Self-Regulation of Eating Behaviours Questionnaire (SREBQ) (Kliemann et al., 2016) |
| | Compulsive Exercise Test (CET) (Taranis et al., 2011) |

| Module 2: Thoughts and Behaviours | |
|---|---|
| | Obsessive Compulsive Inventory-Revised (OCI-R) (Foa et al., 2002) |
| | Adult ADHD Self-Report Scale (ASRS) (Kessler et al., 2005) |
| | Difficulties in Emotion Regulation Scale (DERS-16) (Bjureberg et al., 2016) |
| | Life Events Checklist for DSM-5 (LEC-5) (Weathers et al., n.d.) |
| | Autism Spectrum Quotient(AQ-10) (Allison et al., 2012) |
| | Assessment for lifetime MDD and GAD based on the WHO Composite International Diagnostic Interview short-form (CIDI-SF) (Patten et al., 2000),ICD-11 (Kogan et al., 2016) and other anxiety disorders based on the DSM-V criteria modified for the GLAD Study (Davies et al., 2019) |
| | Generalised Anxiety Disorder 7-item (GAD-7) (Spitzer et al., 2006) |
| | Mood Disorder Questionnaire (MDQ) (Hirschfeld, 2002) |
| | WHO Quality of Life (modified short version for subjective well being) (WHOQOL) (Group, Whoqol, 1994) |
| | Patient Health Questionnaire 9-item (PHQ-9) (Kroenke et al., 2001) |
| | Post-traumatic Stress Disorder Checklist for DSM-5 (PCL-5) (Blevins et al., 2015) |

| Module 3: Lifestyle | |
|---|---|
| | Family Structure (non-validated questionnaire adapted from the Australian Genetics of Depression Study, (Byrne et al., 2019)) |
| | Alcohol Use Disorders Identification Test (AUDIT) (Saunders et al., 1993) |
| | Drug Use Disorders Identification Test (DUDIT) (Bergman et al., 2003) |
| | Drugs and Addiction (non-validated questionnaire from Australian Genetics of Depression Study) (Byrne et al., 2019) |
| | Games and Gambling (non-validated questionnaire adapted from the Australian Genetics of Depression Study) (Byrne et al., 2019) |
| | Headaches and Migraines (non-validated questionnaire adapted from the Australian Genetics of Depression Study) (Byrne et al., 2019) |
| | Work and Sleep (non-validated questionnaire adapted from the Australian Genetics of Depression Study) (Byrne et al., 2019) |

| Module 4: Quality of Life | |
|---|---|
| | 12-Item Short Form Survey (SF-12) (Ware et al., 1996) |
| | Eating Disorder Quality of Life (EDQOL) (Engel et al., 2006) |

| Module 5: Personality | |
|---|---|
| | Personality Inventory for DSM-5 Brief Form (PID-5-BF) (Krueger et al., 2014) |
| | "Personal Standards" from Multidimensional Personality Scale (MPS) (Clavin et al., 1996) |

| Module 6: Treatment | |
|---|---|
| | Eating Disorder Treatment Questionnaire (non-validated based on New Zealand treatment questionnaire) |
| | Medication usage questionnaire (non-validated questionnaire adapted from the Australian Genetics of Depression Study) (Byrne et al., 2019) |

**Supplementary Material 4. EDGI UK Saliva Instruction flyer**

#### What do I do next?

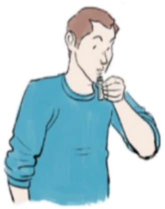

- Please clean your teeth **30 minutes before collection** to reduce food particles mixing with the saliva sample. Please do not use mouthwash. Do not eat, drink, smoke or chew gum in this time.
- There is a DNA stabiliser inside the collection tube, so keep the tube upright to prevent any spillage. Ensure this liquid is at the 2ml line before use. If not then please contact us to arrange a new kit to be sent to you.
- Replace the lid on the tube with the funnel. Please do not overtighten the funnel. Put the cap of the tube somewhere safe.
- **Fill the tube with saliva up to the 4ml line**. This can take a while so do not worry about completing your saliva donation quickly. To encourage saliva production, you can rub the inside of your cheeks with your tongue and/or gently massage your cheeks.
- Once finished, remove and dispose of the funnel, replace the cap, and put these back into the plastic collection box. Please **ensure that the cap is tight** to reduce the risk of leaks.
- Put the completed kit in the transparent safety bag provided. Pull the tab on the top of the bag and seal the bag to ensure it does not leak. Place this transparent bag into the purple mailing bag.
- Seal the return mailing bag and put it in the post.

#### What do I do once I have finished?

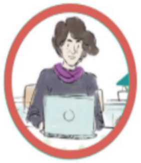

**Once you're done, please place the purple mailing bag into the post box. If you have any concerns, or your contact details have changed since registration, please do not hesitate to CONTACT US on FREEPHONE 0800 917 6016, phone 0207 848 1639, or**

The purple envelope that your saliva kit came in is recyclable if you remove the postage sticker

**THANK YOU FOR TAKING PART—YOU HAVE MADE A VERY IMPORTANT CONTRIBUTION TO RESEARCH.**

EDGI Saliva Instructions flyer, version 1.2 Date: 05/08/2020

NIHR | BioResource

### Providing a Saliva Sample

EATING DISORDERS
EDGI
GENETICS INITIATIVE

Beat
Eating disorders

#### An exciting opportunity to get involved in eating disorder research

Thank you for volunteering to take part in research collaborating with the NIHR BioResource Centre Maudsley, part of the NIHR BioResource, and arranging to give us a saliva sample. This leaflet provides you with some information to **help you give a sample quickly and easily**, and to safely return it to us.

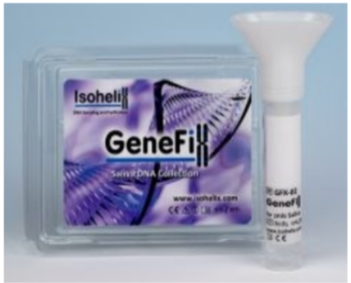


#### Why do I need to provide a saliva sample?

Your saliva contains your **DNA** (your unique genetic blueprint). Researchers can use your anonymised DNA to **investigate the links between mental and physical disorders, the environment, and your genes.**

#### What is in my saliva kit?

- A purple return mailing bag with a tracked 48 return sticker
- A transparent "Safetybag" containing a white absorbent sheet
- 1 x DNA saliva kit

**Supplementary Figure 1.** ***Plot showing the overlap in lifetime eating disorder diagnoses***

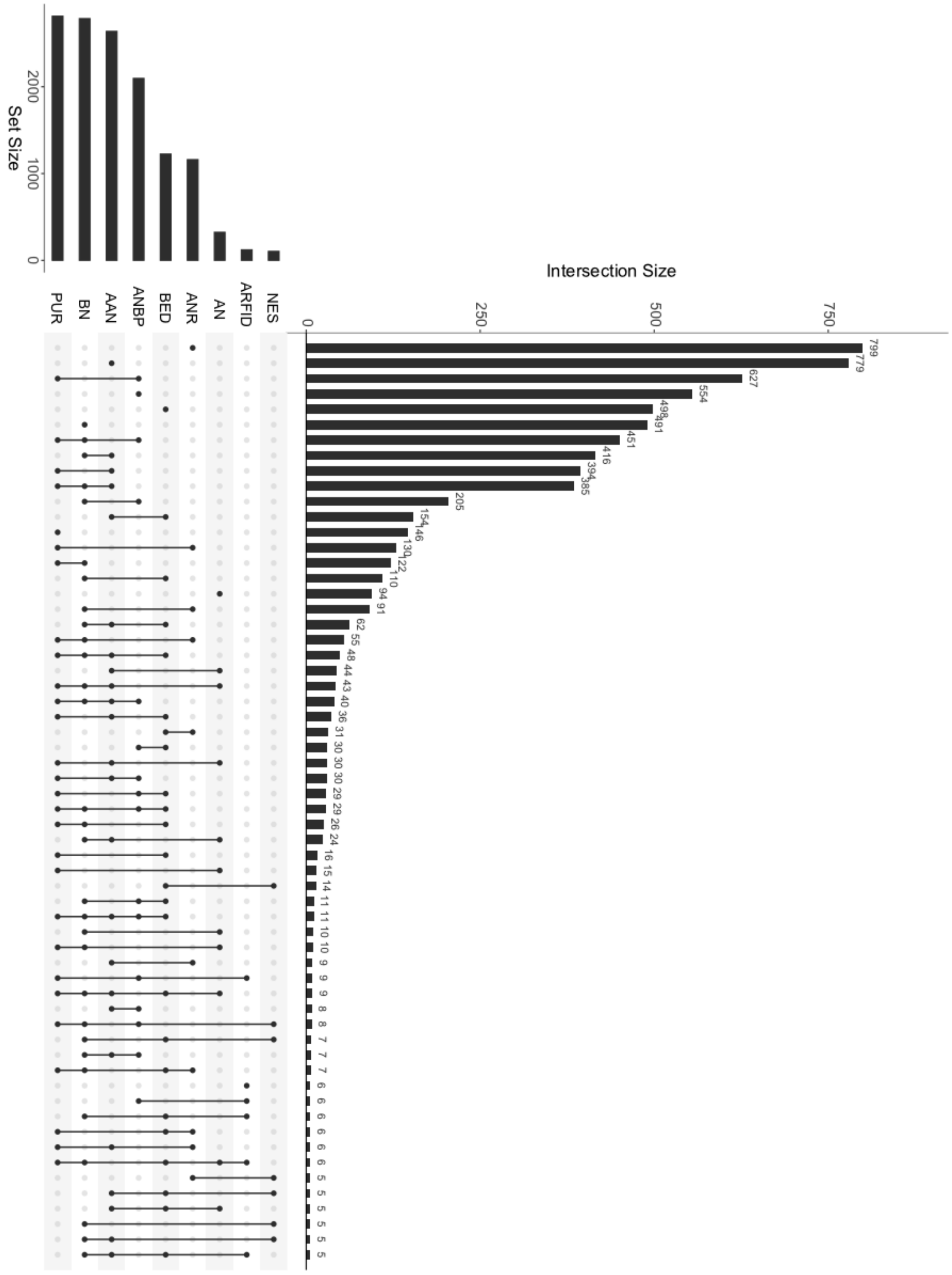


*Diagnoses are algorithm-derived via responses to the ED100K and/or self-report in the Mental Health Diagnosis (MHD) questionnaire. Some eating disorders are algorithm-derived only (anorexia nervosa binge-eating/purging and anorexia nervosa restricting). Set size represents overall size of each group, whilst intersection size indicates size of overlapping groups. Groups smaller than 5 are not presented. Abbreviations: AN = anorexia nervosa (no subtype), ANR = anorexia nervosa restricting, BED = binge-eating disorder, ANBP = anorexia nervosa binge-eating/purging, AAN = atypical anorexia nervosa, BN = bulimia nervosa, PUR = purging disorder.*

#### Supplementary Table 1. Detailed table of somatic diseases

**Supplementary Table 1.** Detailed somatic comorbidities of each subsample defined by diagnosis: anorexia nervosa (AN; n = 3,601), atypical anorexia nervosa (AAN; n = 2,639), binge-eating disorder (BED; n = 1,225), bulimia nervosa (BN; n = 2,786), and purging disorder (PUR; n = 2,816), September 2022.

| **Characteristic** | **AN (*n* = 3,601)** | **AAN (*n* = 2,639)** | **BED (*n* = 1,225)** | **BN (*n* = 2,786)** | **PUR (*n* = 2,816)** |
|---|---|---|---|---|---|
| Epilepsy or convulsions | 91 (2.5%) | 65 (2.5%) | 29 (2.4%) | 70 (2.5%) | 76 (2.7%) |
| Migraines | 703 (20%) | 656 (25%) | 372 (30%) | 666 (24%) | 662 (24%) |
| Multiple sclerosis | 5 (0.1%) | <5 (0.2%) | <5 (0.2%) | <5 (0.1%) | <5 (0.1%) |
| Parkinson's disease | 0 | 0 | 0 | <5 (<0.1%) | 0 |
| Severe memory loss | 7 (0.2%) | 5 (0.2%) | <5 (<0.1%) | <5 (0.1%) | 6 (0.2%) |
| Hay fever | 1,123 (31%) | 941 (36%) | 491 (40%) | 959 (34%) | 981 (35%) |
| Drug allergy | 394 (11%) | 350 (13%) | 208 (17%) | 353 (13%) | 362 (13%) |
| Food allergy | 423 (12%) | 314 (12%) | 142 (12%) | 342 (12%) | 357 (13%) |
| Other allergies | 298 (8.3%) | 268 (10%) | 163 (13%) | 269 (9.7%) | 265 (9.4%) |
| Osteoporosis | 491 (14%) | 111 (4.2%) | 42 (3.4%) | 150 (5.4%) | 239 (8.5%) |

| | | | | | |
|---|---|---|---|---|---|
| Osteoarthriris | 71 (2.0%) | 106 (4.0%) | 105 (8.6%) | 94 (3.4%) | 71 (2.5%) |
| Rheumatoid arthritis | 36 (1.0%) | 34 (1.3%) | 29 (2.4%) | 41 (1.5%) | 32 (1.1%) |
| Any other arthritis | 77 (2.1%) | 62 (2.4%) | 55 (4.5%) | 46 (1.7%) | 57 (2.0%) |
| Asthma | 745 (21%) | 685 (26%) | 373 (31%) | 695 (25%) | 728 (26%) |
| Emphysema or chronic bronchitis | 14 (0.4%) | 8 (0.3%) | 11 (0.9%) | 13 (0.5%) | 10 (0.4%) |
| Heart attack or angina | 10 (0.3%) | 9 (0.3%) | 10 (0.8%) | 8 (0.3%) | 13 (0.5%) |
| High cholesterol | 73 (2.0%) | 109 (4.2%) | 122 (10%) | 96 (3.5%) | 68 (2.4%) |
| High blood pressure | 83 (2.3%) | 144 (5.5%) | 166 (14%) | 137 (4.9%) | 97 (3.5%) |
| Atrial fibrillation | 22 (0.6%) | 18 (0.7%) | 5 (0.4%) | 22 (0.8%) | 21 (0.8%) |
| Stroke | 8 (0.2%) | 9 (0.3%) | 6 (0.5%) | 9 (0.3%) | 8 (0.3%) |
| Crohn's disease | 19 (0.5%) | 12 (0.5%) | 5 (0.4%) | 7 (0.3%) | 17 (0.6%) |
| Ulcerative colitis | 27 (0.8%) | 15 (0.6%) | 13 (1.1%) | 16 (0.6%) | 27 (1.0%) |

| | | | | | |
|---|---|---|---|---|---|
| Coeliac disease | 59 (1.7%) | 30 (1.1%) | 18 (1.5%) | 40 (1.4%) | 38 (1.4%) |
| Diabetes type 1 (early onset) | 6 (0.2%) | 4 (0.2%) | <5 (0.3%) | 9 (0.3%) | 6 (0.2%) |
| Diabetes type 1 (late onset) | 6 (0.2%) | <5 (0.2%) | <5 (0.3%) | 9 (0.3%) | 8 (0.3%) |
| Diabetes type 2 (late onset) | 9 (0.3%) | 31 (1.2%) | 66 (5.4%) | 36 (1.3%) | 17 (0.6%) |
| Diabetic-related pain | 7 (0.2%) | 10 (0.4%) | 10 (0.8%) | 9 (0.3%) | 7 (0.2%) |
| Virus-related pain (post herpetic neuralgia) | 37 (1.0%) | 26 (1.0%) | 18 (1.5%) | 31 (1.1%) | 31 (1.1%) |
| Breast cancer | 12 (0.3%) | 8 (0.3%) | 7 (0.6%) | 8 (0.3%) | 8 (0.3%) |
| Lung cancer | 0 | 0 | 0 | 0 | 0 |
| Stomach cancer | 0 | 0 | 0 | 0 | 0 |
| Colon cancer | <5 (<0.1%) | <5 (<0.1%) | <5 (0.2%) | <5 (<0.1%) | 0 |
| Uterus cancer | 0 | <5 (<0.1%) | <5 (0.2%) | 5 (0.2%) | <5 (<0.1%) |
| Prostate cancer | 0 | 0 | 0 | 0 | 0 |

| | | | | | |
|---|---|---|---|---|---|
| Psoriasis | 183 (5.1%) | 162 (6.2%) | 108 (8.8%) | 173 (6.2%) | 178 (6.3%) |
| Vitiligo | 26 (0.7%) | 23 (0.9%) | 11 (0.9%) | 26 (0.9%) | 26 (0.9%) |
| Eczema | 982 (27%) | 732 (28%) | 375 (31%) | 782 (28%) | 797 (28%) |
| Thyroid disease | 195 (5.4%) | 147 (5.6%) | 117 (9.6%) | 173 (6.2%) | 157 (5.6%) |

*Note.* Somatic comorbidities are self-reported via the MHD. Percentages are based on complete data, i.e., those who skipped the question or responded with "Prefer not to answer" or "Don't know" are not included in the calculation. Participants could be assigned to more than one eating disorder group.
