## Supplementary material for "The Eating Disorders Genetics Initiative (EDGI) United Kingdom": EDGI UK Sign-up Questionnaire

### Eating Disorders Genetics Initiative (EDGI) United Kingdom: Sign-up Questionnaire

#### Contents

| | Page |
|---|---|
| **Introduction** | 3 |
| **Section 1:** Demographics and personal information | 3 |
| **Section 2:** ED100K Questionnaire | 18 |
| **Section 3:** Night Eating Syndrome Questionnaire | 36 |
| **Section 4:** Muscle Dysmorphic Disorder Inventory | 40 |
| **Section 5:** Avoidant/restrictive Food Intake Disorder Screener | 41 |
| **Section 6:** Childhood Trauma and Possible Stresses and Strains of Life | 42 |
| **Section 7:** Study Feedback | 44 |

### 0 - Introduction

#### Questions about your experiences in relation to mental health

Thank you for taking the time to assist us in our research. To make it easier for you to participate, you can start the questionnaire, log off and return to it later.

We are interested in knowing more about your mental health. Some of the questions are sensitive and may be difficult to answer. Please try to answer as many questions as you can and give the most accurate answers possible. However, if you cannot answer a question, you can always choose the "don't know" or "prefer not to answer" option where available, or skip the question and move on.

**If you are concerned about answering these questions, it may be preferable to fill out this survey in the presence of someone who is able to give you support (e.g. a friend or family member).**

Information on where to find help for the issues covered in this questionnaire is given at the end of the survey.

Please note that this questionnaire will not be used to provide you with a diagnosis, your responses will only be used to assess your eligibility to take part in the Eating Disorders Genetics Initiative (EDGI).

If you would like to receive a formal diagnosis, please contact your GP. For more information and support on eating disorders, please visit the Beat website or visit the 'Useful Links' page on the EDGI website.

Please answer each question. If you are unsure, please give us your best guess or estimate.

Please do not use your browser's back button while taking this survey. If you need to return to a previous question, please use the back button at the bottom left corner of this survey.

### 1 – Demographics and personal information (Edited Common Subject Assessment)

#### Questions about you and your medical history

These questions are about you and your medical health. There are no right or wrong answers. You can put "Don't know".

#### Section 1 – Your Information

**1) Date of Birth**

☐☐ ☐☐ ☐☐☐☐

DD MM YYYY

**1.1) How old are you now?**

☐☐

**If over 16, please continue to question 1.2. If under 16 participants will receive this message:**

Thank you for your interest in taking part in the Eating Disorders Genetics Initiative. We appreciate your support and willingness to take part in research.

We are currently only recruiting participants who are **16 or over** and who **live in England.** If you are under the age of 16 and would still like to take part in the BioResource as a volunteer, please contact.

Unfortunately, we are not currently recruiting participants **in other parts of the UK.** However, we will be expanding our research scope in due course.

You have received this message because you have confirmed that you are **under 16** or live **outside of England**. At this time, you are **not eligible** to take part in the EDGI, but if you live in the UK we will contact you when recruitment becomes available in your area. If you are currently under 16, please sign-up again when you have turned 16.

If you live **outside of England** however you attend an **NHS service within England**, please email or call us as you may still be eligible to take part.

**1.2) Please confirm where you are currently living or where your permanent address is based?**

☐England (continue to question 2) ☐Scotland

☐Wales ☐Northern Ireland

☐Outside UK

**If participant selects anything other than England they will receive this message:**

Thank you for your interest in taking part in the Eating Disorders Genetics Initiative. We appreciate your support and willingness to take part in research.

We are currently only recruiting participants who are **16 or over** and who **live in England.**

Unfortunately, we are not currently recruiting participants **in other parts of the UK.** However, we will be expanding our research scope in due course.

You have received this message because you have confirmed that you are **under 16** or live **outside of England**. At this time, you are **not eligible** to take part in the EDGI, but if you live in the UK we will contact you when recruitment becomes available in your area. If you are currently under 16, please sign-up again when you have turned 16.

If you live **outside of England** however you attend an **NHS service within England**, please email or call us as you may still be eligible to take part.

**1.3 Have you previously taken part in the Genetic Links to Anxiety and Depression (GLAD) Study?**

☐Yes ☐No (skip to Q2) ☐Don't know (skip to Q2)

**1.4 Please state which email and phone number and postcode you used in order to sign up to the Genetic Links to Anxiety and Depression (GLAD) Study.**

**Email:_______________**
**Phone number: ________**
**Postcode: _____________**

**1.5 Have you already completed or <u>will</u> complete your Genetic Links to Anxiety and Depression (GLAD) Study saliva sample?**

*Please select "No" to receive a saliva sample in the post*
*Please select "Yes" if you have already provided or will provide a saliva sample, so that we do not send you a second saliva kit (each kit costs the study £10).*

☐Yes ☐No ☐Don't know

**1.4 Did you hear about EDGI through an NHS Trust, GP, talking therapy service, pharmacy, or any other healthcare provider?**

☐Yes (continue to Q1.5) ☐No (skip to Q2)

**1.5 Please specify which NHS Trust, GP or other healthcare provider you hard about EDGI from:**

**Drop down menu with list of all Trusts/GPs, talking therapy services/ pharmacy/ healthcare provider, and Other (with a text box)**

**2) What was your <u>assigned sex at birth</u>? *(Note: This question refers to assigned sex at birth, not gender. Responses to this question are used to select questionnaire items that may be relevant to your medical history.)***

☐Male ☐Female

**2.1) Which gender do you <u>identify</u> with?**

☐Male ☐Female ☐Non-binary

☐Prefer to self-define *(please tell us more)* _____________ ☐Don't know ☐Prefer not to answer

**2.2) Do you identify as transgender?**

☐Yes ☐No

#### 3 is only displayed to those who selected 'female' in 2

**3) Have you ever been pregnant?**

☐Yes ☐No

**4) What is your sexual orientation?**

| | | |
|---|---|---|
| ☐Heterosexual | ☐Homosexual | ☐Bisexual |
| ☐Asexual | ☐ Prefer to self-define *(please tell us more)* ____________________________ ________ | ☐Prefer not to say |

**5) What is your current marital/relationship status?**

| | | |
|---|---|---|
| ☐Single | ☐Relationship (not living together) | ☐Relationship (living together) |
| ☐Married or Civil Partnership | ☐Separated | ☐Divorced |
| ☐Widowed | ☐Other | ☐Prefer not to answer |

**6 Do you have any children?**

☐Yes (Skip to question 6.1) ☐No (Continue to 7)

**6.1) How many children do you have?**

☐☐

**6.2) For each of your children, please list their ages, their genders, their relationship to you (e.g. biological, step, or adopted), and whether or not they live with you in the text box below.**

____________________________________________________________________
__(will be displayed as tick boxes for gender and free text for age and relationship)____________________________________________________________
____________________________________________________________________

**7) How would you describe your vision?**

☐ Normal (i.e. 20/20) ☐Corrected to normal (i.e. glasses)

☐Visually impaired ☐ Prefer not to answer

**8) How would you describe your <u>hearing</u>?**

☐ Normal ☐Corrected to normal (i.e. hearing aid)

☐ Hearing impaired/Deaf ☐Prefer not to answer

**9) Which <u>hand</u> do you usually write with?**

☐Right hand ☐Left hand

☐ I can write with both hands ☐ Prefer not to answer

**10) Do you consider yourself to have a <u>disability</u> (seen or unseen)?**

☐Yes *(continue to 10.1)* ☐No

**10.1 Please tell us more about your disability if you would like to**

___________________________________________________________________________

___________________________________________________________________________

___________________________________________________________________________

**11) How many years did you go to school?** *Please include pre-school (e.g. nursery), primary school, secondary school, college/sixth-form, university and post-university (postgraduate) education.*

☐☐

**11.1) Which of the following qualifications do you have?** *(Please select all that apply)*

☐College or university degree ☐A levels/AS levels or equivalent

☐O levels/GCSEs or equivalent ☐ CSEs or equivalent

☐NVQ or HND or HNC or equivalent ☐Other professional qualifications (e.g. nursing, teaching) _________________

☐None of the above

**12) What is your <u>current employment status?</u>**

☐In paid employment or self-employed ☐Retired

☐Looking after home and/or family ☐Unable to work because of sickness or disability

☐ Unemployed ☐Doing unpaid or voluntary work

☐Full or part-time student ☐None of the above

☐Prefer not to answer

#### Section 2 – Your Origins

**12. You**

| **What is your ethnic origin?** Please tick ONE box: | | | |
|---|---|---|---|
| ☐ | White – British | ☐ | Asian or Asian British - Pakistani |
| ☐ | White - Irish | ☐ | Asian or Asian British - Chinese |
| ☐ | White – Irish Traveler | ☐ | Asian or Asian British - Bangladeshi |
| ☐ | White - German | ☐ | Asian or Asian British - Sri Lanka |
| ☐ | White - French | ☐ | Any other Asian background _____________ |
| ☐ | White - Italian | ☐ | Black or Black British - Caribbean |
| ☐ | White - Polish | ☐ | Black or Black British – South African |
| ☐ | White -Spanish | ☐ | Black or Black British – Kenyan |
| ☐ | White – other ________________ | ☐ | Black or Black British – Nigerian |
| ☐ | Mixed – White and Black Caribbean | ☐ | Black or Black British – Ghanaian |
| ☐ | Mixed – White and Black African | ☐ | Black or Black British – Ugandan |
| ☐ | Mixed – White and Asian | ☐ | Black or Black British – Nigerian |
| ☐ | Other Mixed Background ___________________ | ☐ | Any other black background _______________ |
| ☐ | Asian or Asian British - Indian | ☐ | Arab |

Please answer below:

**12a. Your first language(s)**

__________________________________________________

#### Section 3 –Your Medical History

**The next questions are about height and weight.**

**If you are concerned about answering these questions, it may be preferable to fill out this survey in the presence of someone who is able to give you support (e.g. a friend or family member).**

**Information on where to find help for the issues covered in this questionnaire is given at the end of the survey.**

**13)Please select your preferred units of measure.**

☐Feet/Inches & Pounds ☐Centimeters & Kilograms ☐Body Mass Index (BMI)

**13a) What is your height?**

☐☐☐cm / ☐ft ☐☐ in

**Question 13b will only display to those who selected Feet/Inches & Pounds or Centimeters & Kilograms in question 13**

**13b) How much do you weigh?**

☐☐☐.☐☐ kg / ☐☐ st ☐☐ lb

*If you are pregnant, please provide your weight before you were pregnant.*

**Question 13c will only display to those who selected Body Mass Index in question 13**

**13c) What is your BMI?**

☐☐.☐bmi

**Question 14a will only display to those who selected Feet/Inches & Pounds or Centimeters & Kilograms in question 13**

**14a) What is the <u>most</u> you have <u>ever</u> weighed (except during pregnancy, if female)?**

☐☐☐.☐☐ kg / ☐☐ st ☐☐ lb

*If you were pregnant, please provide your weight before you were pregnant.*

**Question 14b will only display to those who selected Body Mass Index in question 13**

**14b) What is your <u>highest</u> lifetime <u>BMI</u>?**

☐☐.☐bmi

**15) How old were you when you were at your <u>highest</u> weight/BMI?**

☐☐

#### Question 16a will only display to those who selected Feet/Inches & Pounds or Centimeters & Kilograms in question 13

**16a) What is the <u>least</u> you have ever weighed since reaching your <u>adult height</u>? *(When you stopped growing)***

☐☐☐.☐☐ kg / ☐☐ st ☐☐ lb

#### Question 16b will only display to those who selected Body Mass Index in question 13

**16b) What is the <u>lowest</u> your BMI has ever been since reaching your <u>adult height</u>? *(When you stopped growing)***

☐☐.☐bmi

**17) How old were you when you were at your <u>lowest</u> weight/BMI at your adult height? *(When you stopped growing)***

☐☐

#### Question 18 will only display to those who select "female" in question 2

**18) Have you had your first period?**

☐Yes *(continue to 18a)* ☐No *(skip to question 19)*

**18a) How old were you when you had your first period?**

☐☐

**19) Has the <u>doctor</u> ever told you that you were <u>overweight</u>? (Do not include times during pregnancy)**

☐ Yes *(continue to 19b)* ☐ No *(skip to 20)* ☐Don't know *(skip to 20)*

**19a) Please select what ages (in years) that you were overweight. *(Please select all that apply)***

You can tick more than one box.

☐ Young child (10 years or less)
☐Teenager (11-19 years)
☐Young adult (20-30 years)
☐Adult (31-40 years)
☐Adult (41-50 years)
☐Older adult (51-60 years)
☐Older adult (61-70 years)
☐Older adult (71+ years)

**20) Do you have a specific diet?**

☐ No
☐ Vegan
☐Other ______
☐ Vegetarian
☐ Pescatarian

**21) Have you ever been diagnosed with the following illnesses?**

| | | Yes | No | Year diagnosed |
|---|---|---|---|---|
| **Mental Health Disorders** | Depression | ☐ | ☐ | |
| | Depression during or after pregnancy | ☐ | ☐ | |
| | Mania, hypomania, bipolar or manic-depression | ☐ | ☐ | |
| | Anxiety, nerves or generalised anxiety disorder | ☐ | ☐ | |
| | Social anxiety disorder or social phobia | ☐ | ☐ | |
| | Specific phobia (e.g. fear of flying) | ☐ | ☐ | |
| | Agoraphobia | ☐ | ☐ | |
| | Panic attacks or panic disorder | ☐ | ☐ | |
| | Post-traumatic stress disorder (PTSD) | ☐ | ☐ | |
| | Obsessive-compulsive disorder (OCD) | ☐ | ☐ | |
| | Body Dysmorphic disorder (BDD) | ☐ | ☐ | |
| | Other obsessive-compulsive related disorders (e.g. skin-picking) | ☐ | ☐ | |
| | Schizophrenia | ☐ | ☐ | |

| | | | |
|---|---|---|---|
| | Any other type of psychotic illness | ☐ | ☐ |
| | Personality disorder (show Q21a) | ☐ | ☐ |
| | Autism spectrum disorder (ASD) | ☐ | ☐ |
| | Attention deficit or attention deficit and hyperactivity disorder (ADD/ADHD) | ☐ | ☐ |
| **Nervous System Problems** | Epilepsy or convulsions | ☐ | ☐ |
| | Brain tumour | ☐ | ☐ |
| | Migraine | ☐ | ☐ |
| | Multiple sclerosis | ☐ | ☐ |
| | Parkinson's disease | ☐ | ☐ |
| | Severe memory loss (like Alzheimer's disease) | ☐ | ☐ |
| **Allergies** | Hay fever | ☐ | ☐ |
| | Drug allergy<br>If yes, please put which drug:<br>______________________ | ☐ | ☐ |
| | Food allergy<br>If yes, please provide details:<br>________________________ | ☐ | ☐ |
| | Any other allergy<br>If yes, please specify: _______________ | ☐ | ☐ |
| **Bone or Joint Problems** | Osteoporosis or "thin" bones | ☐ | ☐ |
| | Osteoarthritis | ☐ | ☐ |
| | Rheumatoid arthritis | ☐ | ☐ |
| | Hypermobility (Ehlers-Danlos syndrome) | ☐ | ☐ |
| | Ankylosing spondylitis | ☐ | ☐ |
| | Other arthritis | ☐ | ☐ |
| **Lung or Breathing Problems** | Asthma | ☐ | ☐ |
| | Emphysema or chronic bronchitis | ☐ | ☐ |
| **Heart or Circulation Problems** | Heart attack, angina | ☐ | ☐ |
| | High blood cholesterol | ☐ | ☐ |
| | High blood pressure | ☐ | ☐ |
| | Atrial fibrillation | ☐ | ☐ |
| | Paroxysmal tachychardia (POTS) | ☐ | ☐ |
| | Stroke | ☐ | ☐ |

| | | | |
|---|---|---|---|
| **Digestive problems** | Crohn’s disease | ☐ | ☐ |
| | Ulcerative colitis | ☐ | ☐ |
| | Coeliac disease | ☐ | ☐ |
| **Diabetes** | Diabetes type 1 (early onset) | ☐ | ☐ |
| | Diabetes type 2 (late onset) | ☐ | ☐ |
| | Pain due to diabetes (Diabetic neuropathy) | ☐ | ☐ |
| | | ☐ | ☐ |
| **Cancer** | Breast cancer | ☐ | ☐ |
| | Lung cancer | ☐ | ☐ |
| | Stomach cancer | ☐ | ☐ |
| | Colon cancer | ☐ | ☐ |
| | Uterus cancer | ☐ | ☐ |
| | Prostate cancer | ☐ | ☐ |
| **Skin problems** | Psoriasis | ☐ | ☐ |
| | Vitiligo | ☐ | ☐ |
| | Pain due to virus (Post herpetic neuralgia) | | |
| | Eczema | ☐ | ☐ |
| **Endocrine system problems** | Thyroid disease<br>If Yes, please specify___________ | ☐ | ☐ |
| **Put Other Illnesses NOT Listed Above** | 1. | ☐ | ☐ |
| | 2. | ☐ | ☐ |
| | 3. | ☐ | ☐ |

**21a) Which personality disorder have you been diagnosed with?**

☐ Paranoid personality disorder

☐ Schizoid personality disorder

☐ Schizotypal personality disorder

☐ Antisocial personality disorder

☐ Emotionally unstable personality disorder (Borderline personality disorder)

☐ Histrionic personality disorder

☐ Narcissistic personality disorder

☐ Avoidant/anxious personality disorder

☐ Dependent personality disorder

☐ Obsessive-compulsive personality disorder

☐ Don’t know

☐ Prefer not to answer

**22)** Please tell us about any long-term medication that you are taking

| Name of medication | Type (tablets, inhaler etc.) | Reason |
|---|---|---|

**23) Do you have <u>metal implants</u> anywhere in your <u>body</u> (excluding normal dental fillings) e.g. pacemakers, aneurysm clips, cochlear implants or have you suffered an injury involving metal fragments?**

☐ Yes ☐ No

☐ Don't know

**24) Tick the box that best describes you:**

☐ I smoke now. *(Skip to 24c)* ☐ I used to smoke. *(Continue to 24a)*

☐ I have never smoked/vaped. *(skip to 25)* ◻ I vape now. (skip to 24e)

◻ I used to vape (skip to 24f)

**24a) How many years has it been since you <u>stopped smoking?</u>** *(If you are not sure of the exact time, please provide an estimate)*

☐☐

**24b What year did you give up smoking?**

☐☐☐☐

**24c) About how many <u>cigarettes</u> do you or did you smoke per day?**

☐☐

**24d) How many <u>years</u> have you/did you smoke?**

☐☐

**24e.) How many vaping sessions a day do you have? One session is indicated as 5 minutes, or 1 puff every 30 seconds.**

**□ 10 or less** **□ 11-20**
**□21-30** **□31 or more**

**24f. How many years has it been since you stopped vaping? If you are not sure of the exact time, please provide an estimate).**

□□

**24b What year did you give up vaping?**

□□□□

**24c) About how many vaping sessions did you do per day? One session is indicated as 5 minutes, or 1 puff every 30 seconds.**

□□

**25) How many hours per week are you exposed to other people's tobacco smoke? *(passive smoking)***

□□

**26) Do you consume alcohol?**

☐ Yes (continue to 26a) ☐ No (skip to 27)

**26a) Please give an approximate number of units you consume per week (1 pint of beer is 3 units and a small glas of wine is 1.5 units)**

□□

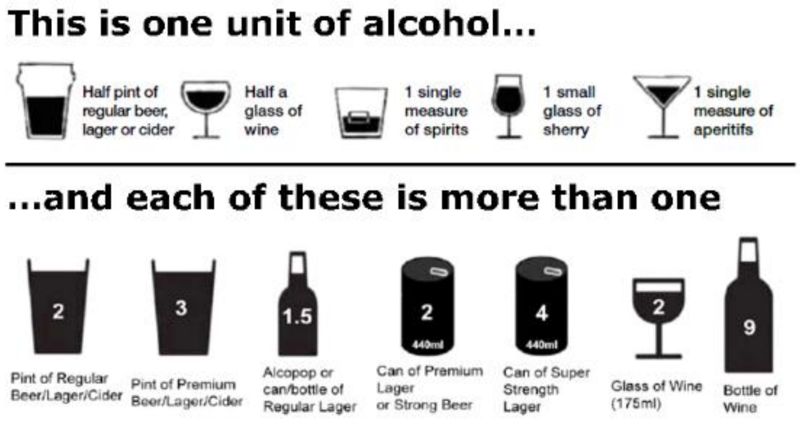

**27) How many cups (or cans) of drinks with <u>caffeine</u> (coffee, tea, caffeinated drinks: e.g. Coke, Diet Coke, Red Bull, etc.) do you have <u>per day</u>?**

☐☐

**28) Do you have <u>trouble</u> falling asleep at night or do you wake up in the middle of the night? *(If this varies a lot, answer this question in relation to the last 4 weeks)***

☐ Never/rarely ☐ Sometimes

☐ Usually ☐ Prefer not to answer

**29) Are you concerned about your memory, because it affects how you work or the way you live from day to day?**

☐Yes *(Continue to 27a)* ☐ No *(skip to question 30)*

**29a) If Yes, has your memory problem got <u>worse</u> in the last year?**

☐ Yes ☐No ☐ Don't know

#### Section 3 –Your Family Medical History

**28) How many brothers and sisters do you have? (Please include all blood relatives, both living and deceased - but not adopted relatives, stepchildren etc.)?**

☐☐ Half brothers ☐☐ Half sisters

☐☐ Full brothers ☐☐ Full sisters

**29) Do you have a twin brother or sister?**

☐ Yes (continue to 29a) ☐No (skip to 30)

**29a) Is your twin…**

☐ Identical ☐Non-identical ☐ Not known

**30.) Have any of your first-degree blood relatives (parents, siblings, children) ever been diagnosed with coronary artery disease or stroke?**

☐ Yes ☐No (skip to Q31) ☐ Don't know (Skip to Q31)

| | | | | |
|---|---|---|---|---|
| Mother | ☐ Coronary artery disease | At age: | ☐ Stroke | At age: |
| Father | ☐ Coronary artery disease | At age: | ☐ Stroke | At age: |
| Brother 1 | ☐ Coronary artery disease | At age: | ☐ Stroke | At age: |
| Brother 2 | ☐ Coronary artery disease | At age: | ☐ Stroke | At age: |
| Sister 1 | ☐ Coronary artery disease | At age: | ☐ Stroke | At age: |
| Sister 2 | ☐ Coronary artery disease | At age: | ☐ Stroke | At age: |
| Child 1 (son/daughter) | ☐ Coronary artery disease | At age: | ☐ Stroke | At age: |
| Child 2 (son/daughter) | ☐ Coronary artery disease | At age: | ☐ Stroke | At age: |

**31. Have any of your first-degree blood relatives (parents, siblings, children) ever been diagnosed with diabetes 1 or/and 2?**

☐ Yes ☐No (skip to Q32) ☐ Don't know (Skip to Q32)

| a. Please indicate which family member | | b. Please indicate approximate age in years at diagnosis | c. Did they receive uninterrupted insulin injections since diagnosis? | | |
|---|---|---|---|---|---|
| Mother | ☐ | At age: | ☐ Yes | ☐ No | ☐ Don't know |
| Father | ☐ | At age: | ☐ Yes | ☐ No | ☐ Don't know |
| Brother 1 | ☐ | At age: | ☐ Yes | ☐ No | ☐ Don't know |
| Brother 2 | ☐ | At age: | ☐ Yes | ☐ No | ☐ Don't know |

| | | | | | |
|---|---|---|---|---|---|
| Sister 1 | ☐ | At age: | ☐ Yes | ☐ No | ☐ Don't know |
| Sister 2 | ☐ | At age: | ☐ Yes | ☐ No | ☐ Don't know |
| Child 1 (son/daughter) | ☐ | At age: | ☐ Yes | ☐ No | ☐ Don't know |
| Child 2 (son/daughter) | ☐ | At age: | ☐ Yes | ☐ No | ☐ Don't know |

**32. Have any of your first-degree blood relatives (parents, siblings, children) ever been diagnosed with cancer?**

☐ Yes ☐No (skip to Section 2) ☐ Don't know (Skip Section 2)

| a. Please indicate which family member | | b. Please indicate approximate age in years at diagnosis | c. Briefly indicate what type of cancer |
|---|---|---|---|
| Mother | ☐ | At age: | Type: |
| Father | ☐ | At age: | Type: |
| Brother 1 | ☐ | At age: | Type: |
| Brother 2 | ☐ | At age: | Type: |
| Sister 1 | ☐ | At age: | Type: |
| Sister 2 | ☐ | At age: | Type: |
| Child 1 (son/daughter) | ☐ | At age: | Type: |
| Child 2 (son/daughter) | ☐ | At age: | Type: |

### 2 – ED100K Questionnaire

#### Section A: Some questions about your mental health

**If you are concerned about answering these questions, it may be preferable to fill out this survey in the presence of someone who is able to give you support (e.g. a friend or family member).**

**Information on where to find help for the issues covered in this questionnaire is given at the end of the survey.**

**A1) Have you ever suspected that you may have an <u>eating disorder</u>, whether or not you were ever diagnosed?**

☐ Yes ☐ No

☐ Not sure

**A2) Have you ever received treatment for an eating disorder?**

☐ Yes ☐ No

**Question A3-4 will only display to those who select either "yes" or "not sure" in question A1 OR those who select "yes" to A2. Those who select "no" to both will skip to the next section**

**A3) Have you ever been diagnosed with one or more of the following eating disorders by a professional, even if you don't have it currently?**

*By professional we mean: any doctor, nurse or person with specialist training (such as a psychologist, psychiatrist etc.) Please include disorders even if you did not need treatment for them or if you did not agree with the diagnosis.*

| | | |
|---|---|---|
| ☐Anorexia nervosa | ☐Atypical anorexia nervosa | ☐Bulimia nervosa |
| ☐Binge-eating disorder | ☐Purging disorder | ☐Night eating syndrome |
| ☐ Atypical bulimia nervosa | ☐Atypical binge-eating disorder | |
| ☐Pica | ☐Avoidant/restrictive food intake disorder | ☐Rumination disorder |
| ☐Other eating disorder ________________ | ☐Don't know | ☐Prefer not to say |

A3a) Are you currently experiencing (answer from QA3) or have you experienced this eating disorder in the past?

| Anorexia nervosa | Current? | Past? |
|---|---|---|
| Atypical anorexia nervosa | | |
| Binge-eating disorder | | |
| Atypical binge-eating disorder | | |
| Bulimia nervosa | | |
| Atypical bulimia nervosa | | |
| Purging disorder | | |
| Night eating disorder | | |
| Pica | | |
| Avoidant/restrictive food intake disorder | | |
| Rumination disorder | | |
| Other eating disorder | | |

**A4) Have you ever had a period of time when you weighed <u>much less</u> than other people thought you ought to weigh?**

☐ Yes ☐ No

☐ Don't know

**Section B:** **The following questions are about the time when you weighed much less than people thought you should weigh, were at a low weight, or when you had anorexia nervosa.**

**(Please note that 'low weight' is considered to be a BMI lower than 18.5)**

**If you are concerned about answering these questions, it may be preferable to fill out this survey in the presence of someone who is able to give you support (e.g. a friend or family member).**

**Information on where to find help for the issues covered in this questionnaire is given at the end of the survey.**

**Section B is only displayed if the participant selected "Yes" to A4, selected "Anorexia nervosa" in A3 or has had a BMI of less than 18.5 (this could be current BMI or lowest ever BMI). If none of this applies participant will be skipped to section C.**

**B1) When you weighed much less than other people thought you ought to weigh or were at your lowest weight, was this due to an illness other than an eating disorder?**

☐ Yes *(continue to B1a)* ☐ No *(skip to B2)*

☐ Don't know *(skip to B2)*

**B1a) What was the illness?**

___________________________________________________________

**B2a is only displayed to those who selected 'Feet, inches and pounds' or 'Centimeters and kilograms' in 13**

**B2a) How low did your weight get during this time?**

☐☐☐.☐☐ kg / ☐☐ st ☐☐ lb

**B2b is only displayed to those who selected 'BMI' in 13**

**B2b) How low did your BMI get during this time?**

☐☐.☐BMI

**B3) How old were you?**

☐☐

**B4) How tall were you then?**

☐☐☐cm / ☐ft ☐☐ in

**B5) During the time when you were at this low weight/BMI, did you still feel fat?**

☐No ☐Slightly ☐Somewhat

☐Very ☐Extremely ☐Don't know/prefer not to say

**B6) During the time when you were at this low weight/BMI, how afraid were you that you might gain weight or become fat?**

☐Not afraid ☐Slightly afraid ☐Somewhat afraid

☐Very afraid ☐Extremely afraid ☐ Don't know/prefer not to say

**B7)For how long did you have anorexia nervosa OR were at this low weight/BMI?**

☐☐years ☐☐months

☐Don’t know

**B8) During the time when you were at this low weight/BMI, how dependent was your self-worth on your body shape or weight?**

| | 1 | 2 | 3 | 4 | 5 | 6 | 7 | |
|---|---|---|---|---|---|---|---|---|
| Not at all dependent | | | | | | | | Completely dependent |

| | **Not at all** | **Somewhat** | **Very much** | **Don’t know** |
|---|---|---|---|---|

| | | | | |
|---|---|---|---|---|
| **B9) Did you ever think your low weight/BMI had negative consequences for your health?** | ☐ | ☐ | ☐ | ☐ |
| **B10) During the time when you were at this low weight/BMI, did you ever experience your body or parts of your body to be larger than they actually were or than other people thought they were?** | ☐ | ☐ | ☐ | ☐ |

**B11 & B12 are only displayed to those who selected "Female" in question 2. Participants who selected "Male" in question 2 will skip to section D.**

**B11) Before this time had your periods already started?**

☐Yes *(continue to B12)* ☐No *(skip to section D)* ☐Don't know *(continue to B12)*

**B12) Did your periods stop at any time during this time of low weight?**

☐Yes *(continue to B13)* ☐No *(skip to section D)* ☐Don't know *(skip to section D)*

**B12.1) For how long (in years and months) did your periods stop?**

☐☐years ☐☐months

☐Don't know

**B12.2) Roughly how old were you (in years) when your periods stopped?**

☐☐

☐Don't know

#### Section C: The following questions are about any significant weight loss you have experienced in your life

**If you are concerned about answering these questions, it may be preferable to fill out this survey in the presence of someone who is able to give you support (e.g. a friend or family member).**

**Information on where to find help for the issues covered in this questionnaire is given at the end of the survey.**

**C1) Has there ever been a period when you have lost what you or others considered to be a significantly large amount of weight?**

☐ Yes *(continue to C1a)* ☐ No *(skip to skip to section D)*

☐ Don't know *(skip to C1a)*

**C1a) How much weight did you lose?**

☐☐☐.☐☐ kg / ☐☐ st ☐☐ lb

**C1b) How much did you weigh prior to this period of significant weight loss?**

☐☐☐.☐☐ kg / ☐☐ st ☐☐ lb

**C1c) Over what time period did this weight loss occur?**

☐ Up to 1 month ☐ Between 1 month and 3 months

☐ Between 3 months and 6 months ☐ Up to 12 months

**If percentage of body weight lost is less than 5% in 1 month, 7.5% in 3 months, 10% in 6 months or 20% in 12 months this is classed as non-severe malnutrition and participants will be skipped to the next block. Participants who meet the criteria for severe malnutrition will continue to Question C2.**

**C2) During this period of significant weight loss was this due to an illness other than an eating disorder?**

☐ Yes *(continue to C2a)* ☐ No *(Skip to C3)*

☐ Don't know *(Skip to C3)*

**C2a) What was the illness?**

_______________________________________________________________

**C3) During this time of significant weight loss, did you still feel fat?**

☐No ☐Slightly ☐Somewhat

☐Very ☐Extremely ☐Don't know/prefer not to say

**C4) During this time of significant weight loss, how afraid were you that you might gain weight or become fat?**

☐Not afraid ☐Slightly afraid ☐Somewhat afraid

☐Very afraid ☐Extremely afraid ☐ Don't know/prefer not to say

**C5) During this time of significant weight loss, how dependent was your self-worth on your body shape or weight?**

| | 1 | 2 | 3 | 4 | 5 | 6 | 7 | |
|---|---|---|---|---|---|---|---|---|
| Not at all dependent | | | | | | | | Completely dependent |

| | Not at all | Somewhat | Very much | Don't know |
|---|---|---|---|---|
| **C6) Did you ever think that this period of significant weight loss had negative consequences for your health?** | ☐ | ☐ | ☐ | ☐ |
| **C7) During this time of significant weight loss, did you ever experience your body or parts of your body to be larger than they actually were or than other people thought they were?** | ☐ | ☐ | ☐ | ☐ |

**C8 & C9 are only displayed to those who selected "Female" in question 2. Participants who selected "Male" in question 2 will skip to section C.**

**C8) Before this time of significant weight loss had your periods already started?**

☐Yes *(continue to C9)* ☐No *(skip to section D)* ☐Don't know *(continue to C9)*

**C9) Did your periods stop at any time during this time of significant weight loss?**

☐Yes *(continue to C9.1)* ☐No *(skip to section D)* ☐Don't know *(skip to section D)*

**C9.1) For how long *(in years and months)* did your periods stop?**

☐☐years ☐☐months

**C9.2) Roughly how old were you *(in years)* when your periods stopped?**

☐☐

#### Section D: The following questions are about your eating habits

**If you are concerned about answering these questions, it may be preferable to fill out this survey in the presence of someone who is able to give you support (e.g. a friend or family member).**

**Information on where to find help for the issues covered in this questionnaire is given at the end of the survey.**

**D1) Have you ever had regular episodes of overeating or eating binges when you ate what most people would regard as an unusually large amount of food in a short period of time (for example, in a 2-hour period)?**

☐Yes *(continue to D2)* ☐No *(skip to section E)* ☐Don't know *(continue to D2)*

**D2) When you were having regularly occurring episodes of binge eating or overeating, did you feel that your eating was out of control such that you felt you could not stop eating, or that you could not control what or how much you were eating?**

☐No *(skip to section E)* ☐Slightly *(Continue to D3)* ☐Somewhat *(Continue to D3)*

☐Very *(Continue to D3)* ☐Extremely *(Continue to D3)* ☐Prefer not to answer *(Continue to D3)*

| D3) During eating binges, did you: | Yes | No | Don't Know |
|---|---|---|---|

| | | | |
|---|---|---|---|
| **a) Eat much more rapidly than usual?** | ☐ | ☐ | ☐ |
| **b) Eat until you felt uncomfortably full?** | ☐ | ☐ | ☐ |
| **c) Eat large amounts of food when you didn't feel physically hungry?** | ☐ | ☐ | ☐ |
| **d) Eat alone because you were embarrassed by what/how much you were eating?** | ☐ | ☐ | ☐ |
| **e) Feel ashamed/disgusted with yourself, depressed, or very guilty after overeating?** | ☐ | ☐ | ☐ |
| **f) Feel like you had no control over your eating (e.g. not being able to stop eating, feeling compelled to eat or going back and forth for more food)?** | ☐ | ☐ | ☐ |
| **g) Make yourself vomit as a means to control your weight and shape?** | ☐ | ☐ | ☐ |

**D4 is only displayed to those who selected "yes" for the corresponding statements in D3. Participants who select "no" for all the statements in D3 will skip to question D5.**

| D4) If yes, how often did you: | Rarely | Sometimes | Often | Always |
|---|---|---|---|---|
| a) Eat much more rapidly than usual? | ☐ | ☐ | ☐ | ☐ |
| b) Eat until you felt uncomfortably full? | ☐ | ☐ | ☐ | ☐ |
| c) Eat large amounts of food when you didn't feel physically hungry? | ☐ | ☐ | ☐ | ☐ |
| d) Eat alone because you were embarrassed by what/how much you were eating? | ☐ | ☐ | ☐ | ☐ |
| e) Feel ashamed/disgusted with yourself, depressed, or very guilty after overeating? | ☐ | ☐ | ☐ | ☐ |
| f) Feel like you had no control over your eating (e.g. not being able to stop eating, feeling compelled to eat or going back and forth for more food)? | ☐ | ☐ | ☐ | ☐ |
| g) Make yourself vomit as a means to control your weight and shape? | ☐ | ☐ | ☐ | ☐ |

**D5) Do you/did you feel distressed about your episodes of overeating?**

☐ Yes *(continue to D5a)* ☐ No *(skip to D6)*

**D5a) How distressed did binge eating usually make you feel?**

☐Slightly ☐Somewhat ☐Very

☐Extremely ☐Don't know/prefer not to say

**D6) When you were having regularly occurring episodes of binge eating, how many episodes would you usually experience in one week?**

☐☐ *(continue to C6a if less than 1, skip to C7 if more than 1)*

**D6a) How many episodes of binge eating would you usually experience in one month?**

☐☐ *(continue to C6b if less than 1, skip to C7 if more than 1)*

**D6b) How many episodes of binge eating would you usually experience in one year?**

☐☐

**D7) For how long did you experience regularly occurring binge eating episodes?**

☐☐years ☐☐months

☐Don't know

**D8) During the time when you were binge eating, how dependent was your self-worth on your body shape or weight?**

| | 1 | 2 | 3 | 4 | 5 | 6 | 7 | |
|---|---|---|---|---|---|---|---|---|
| **Not at all dependent** | | | | | | | | **Completely dependent** |

**D9) Roughly how old were you when you began having regular episodes of binge eating?**

☐☐years

☐Don't know

**D10) How old were you when the regular episodes of binge eating stopped?**

☐☐years OR

☐I still have regularly occurring episodes of binge-eating

☐Don't know

**D11 is only displayed to those who were also asked to answer section B (the low weight section).**

**D11) Did you experience regular episodes of binge eating while at your lowest weight?**

☐ **Yes, only during times of low weight** ☐ **Yes, at times of low weight and at times outside of low weight** ☐ **No**

#### Section E: The following questions are about compensatory behaviors

**If you are concerned about answering these questions, it may be preferable to fill out this survey in the presence of someone who is able to give you support (e.g. a friend or family member).**

**Information on where to find help for the issues covered in this questionnaire is given at the end of the survey.**

**E1) Have you ever used any of the following to control your body shape or weight?**

| | Never | A few times, but it never became a habit | Yes, often |
|---|---|---|---|
| a) Made yourself vomit | ☐ | ☐ | ☐ |
| b) Used laxatives (including pills or liquids meant to stimulate bowel movements) | ☐ | ☐ | ☐ |
| c) Used diuretics (water pills) | ☐ | ☐ | ☐ |
| d) Used diet pills (over the counter or prescription) | ☐ | ☐ | ☐ |
| e) Exercised excessively (e.g. felt compelled to exercise, felt uneasy or distressed if unable to exercise) | ☐ | ☐ | ☐ |
| f) Fasted or did not eat (for 8 waking hours or more) | ☐ | ☐ | ☐ |
| g) Other | ☐ | ☐ | ☐ |

**E2 is only displayed to those who selected “yes” in question E1.**

E2) Have you ever used any of the following to compensate for episodes of binge eating or overeating? *(Mark all that apply)*

| | Yes | No |
|---|---|---|
| a) Made yourself vomit | ☐ | ☐ |
| b) Laxatives *(including pills or liquids meant to stimulate bowel movements)* | ☐ | ☐ |
| c) Diuretics *(water pills)* | ☐ | ☐ |
| d) Weight loss pills *(over the counter or prescription)* | ☐ | ☐ |
| e) Excessive exercise *(e.g. feel compelled to exercise, feel uneasy or distressed if unable to exercise)* | ☐ | ☐ |
| f) Fasting *(not eating for 8 waking hours or more)* | ☐ | ☐ |
| g) Other methods _______(text box)______ | ☐ | ☐ |

**E3 is only displayed to those who select "Yes" to "Other methods" in E2**

E3) Please state any other methods used to compensate for episodes of binge eating or overeating.

_______________________________________________________________

**E4 is only displayed to those who select "Yes" to D1**

E4) Did you ever have periods of time that lasted three months or more when you had binge-eating episodes without regularly engaging in any of the following: making yourself vomit, laxative use, diuretic use, taking weight loss pills, excessively exercising (e.g. feel compelled to exercise, feel uneasy or distressed if unable to exercise), or fasting (not eating for 8 waking hours or more)?

☐ There were periods of time that lasted 3 months or more when I was binge eating when I DID NOT engage in any of the above methods to control my weight or shape.

☐ There were periods of time that lasted 3 months or more when I was binge eating when I DID engage in at least one of the above menthods to control my weight or shape.

☐ Don't know

**E5-6 are only displayed to those who select "Yes" to D1 and selected any of the compensatory behaviours in E2**

E5) Did the binge eating and compensatory behaviours occur at the same time, on average, once a week for 3 months?

☐ Yes, there have been periods when I engaged in binge eating and compensatory behaviors at the same time, on average, once a week for 3 months

☐ No

☐ Don't know

**E6) Was there ever a time when you engaged in compensatory behaviors (making yourself vomit, laxative use, diuretic use, taking weight loss pills, excessively exercising) on average, once a week for 3 months when you were not binge eating?**

☐ **Yes, there have been times when I engaged in compensatory behaviors when I was not binge eating.**

☐ **No, at times when I engaged in any of the compensatory behaviors, I was always also binge eating.**

☐ **Don't know**

#### E7 is only displayed to those who were also asked to answer section B (the low weight section).

**E7) During the period of time when you were at your lowest weight, did you use any of the following as a way to control your weight or shape? (Mark all that apply).**

☐**Making yourself vomit**

☐**Fasting** ***(not eating for 8 waking hours or more)***

☐**Laxatives** **(including pulls or liquids meant to stimulate bowel movements)**

☐**Diuretics** (water pills)

☐**Weight loss pills**

☐**Excessive exercise** ***(e.g. feel compelled to exercise, feel uneasy or distressed if unable to exercise)***

☐**Other methods**

☐**None of the above**

#### E8 is only displayed to those who select “Other methods” in E7

**E8) Please state any other methods used to control your weight or shape.**

______________________________________________________________________

#### E9 is only displayed to those who select “Making yourself vomit” in E1

**E9) How old were you when you began self-inducing vomiting for the first time?**

☐☐

☐Don’t know

**E9.1) How often a week did you usually self-induce vomiting during these periods?**

☐☐

**(E9.2 is only displayed to participants who entered less than 1 in E9.1)**

**E9.2) How often a month did you usually self-induce vomiting during these periods?**

☐☐

**(E9.3 is only displayed to participants who entered less than 1 in E9.2)**

**E9.3) How often a year did you usually self-induce vomiting during these periods?**

☐☐

**E9.4) For how long a period of time were you engaging in self-induced vomiting?**

☐☐years ☐☐months

**E10 will only be displayed to those who selected 'Laxatives' in question E1**

**E10) How old were you when you used laxatives for the first time?**

☐☐

**E10.1) How often a week did you usually use laxatives during these periods?**

☐☐

**(E10.2 is only displayed to participants who entered less than 1 in E10.1)**

**E10.2) How often a month did you usually use laxatives during these periods?**

☐☐

**(E10.3 is only displayed to participants who entered less than 1 in E10.2)**

**E10.3) How often a year did you usually use laxatives during these periods?**

☐☐

**E10.4) For how long a period of time were you using laxatives?**

☐☐years ☐☐months

**E11 will only be displayed to those who selected 'Diuretics' in question E1**

**E11) How old were you when you used diuretics for the first time?**

☐☐

**E11.1) How often a week did you usually use diuretics during these periods?**

☐☐

**(E11.2 is only displayed to participants who entered less than 1 in E11.1)**

**E11.2) How often a month did you usually use diuretics during these periods?**

☐☐

**(E11.3 is only displayed to participants who entered less than 1 in E11.2)**

**E11.3) How often a year did you usually use diuretics during these periods?**

☐☐

**E11.4) For how long a period of time were you using diuretics?**

☐☐years ☐☐months

**E12 will only be displayed to those who selected 'Diet pills' in question E1**

**E12) How old were you when you used diet pills for the first time?**

☐☐

**E12.1) How often a week did you usually use diet pills during these periods?**

☐☐

**(E12.2 is only displayed to participants who entered less than 1 in E12.1)**

**E12.2) <u>How often</u> a <u>month</u> did you usually use <u>diet pills</u> during these periods?**

☐☐

**(E12.3 is only displayed to participants who entered less than 1 in E12.2)**

**E12.3) <u>How often</u> a <u>year</u> did you usually use <u>diet pills</u> during these periods?**

☐☐

**E12.4)For <u>how long</u> a period of time were you using <u>diet pills</u>?**

☐☐years ☐☐months

**E13-E18 will only be displayed to those who selected 'Excessive Exercise' in question E1**

**E13) Have you <u>ever</u> felt <u>uneasy</u> or <u>distressed</u> if unable to exercise?**

☐ Yes

☐ No

☐ Don't know

**E14) Have you <u>ever</u> felt <u>compelled</u> to exercise–-like you had to do it--to control your body shape or weight?**

☐ Yes

☐ No

☐ Don't know

**E15) Have there been times when you <u>declined</u> opportunities to be with friends in order to <u>exercise</u>?**

☐ Yes

☐ No

☐ Don't know

**E16) Have you exercised despite an injury or illness that would have prevented others from exercising?**

☐ Yes

☐ No

☐ Don't know

**E17) Have there been times when you modified your diet/eating habits if you were unable to exercise for any reason?**

☐ Yes

☐ No

☐ Don't know

**E18) How old were you when you first exercised to control your weight and shape and felt either compelled to exercise or distressed if unable to exercise?**

☐☐

**E18.1) For how long a period of time did you feel compelled to exercise or felt distressed if unable to exercise?**

☐☐years ☐☐months

☐Don't know

**E18.2) How often a week did you usually exercise during these periods?**

☐☐

##### (E18.3 is only displayed to participants who entered less than 1 in E18.2)

**E18.3) How often a month did you usually exercise during these periods?**

☐☐

##### (E18.4 is only displayed to participants who entered less than 1 in E18.3)

**E18.4) How often a year did you usually exercise during these periods?**

☐☐

E18.4 How old were you when you first exercised to control your weight and shape AND felt either compelled to exercise or distressed if unable to exercise?

☐☐years ☐☐months

☐Don’t know

E18.5)

**E19 will only be displayed to those who selected ‘Fasting’ in question E1**

**E19) How old were you when you fasted for the first time?** ***(Fasting is not eating for 8 waking hours or more. Do not include fasting for religious purposes.)***

☐☐

☐Don’t know

**E19.1) For what period of time did you engage in periods of fasting? Fasting is not eating for 8 waking hours or more.**

☐☐years ☐☐months

**E19.2) How often a week did you usually fast during these periods?**

☐☐

**(E19.3 is only displayed to participants who entered less than 1 in E19.2)**

**E19.3) How often a month did you usually fast during these periods?**

☐☐

**(E19.4 is only displayed to participants who entered less than 1 in E19.3)**

**E19.4) How often a year did you usually fast during these periods?**

☐☐

**E19.5) How old were you when you stopped fasting?**

☐ I still currently fast

□ Please enter your age when you last fasted if not currently fasting

□□years □□months □Don’t know

### 3 – Night Eating Syndrome Questionnaire (NES-Q)

#### The following questions are about your daily routines and mealtimes

**If you are concerned about answering these questions, it may be preferable to fill out this survey in the presence of someone who is able to give you support (e.g. a friend of family member).**

**Information on where to find help for the issues covered in this questionnaire is given at the end of the survey.**

**1) What time do you usually go to bed in the evening (turn out the lights in order to sleep)?**

___PM
___AM

**2.) What time do you usually get out of bed in the morning?**

___PM
___AM

**3.) On most days, do you experience loss of appetite in the morning?**

☐Yes ☐No

**4.) How often do you typically eat breakfast (after your final morning awakening)?**

________ times a week

**5.) What time do you usually have the first meal of the day?**

___ AM/PM

**6.) What percentage (%) of food do you generally eat after 19:00, as a percentage?**

_______%

**7.) What time do you usually have your evening meal?**

___PM
___AM

**8.) How much food do you generally eat after your evening meal, as a percentage (%)?**

______%

**8a.) For how long have you been consuming at least this much after your evening meal?**

__________years
___________months

**9.) On most days, do you have a strong urge to eat between dinner and sleep onset and/or during the night?**

☐Yes ☐No

**10.) Do you have trouble falling asleep at night?**

☐Yes ☐No

If participant answers YES in Q10, follow to Q10a

**10a.) How many times each week do you have trouble falling asleep at night?**

☐ Drop down menu 1-7, ☐None

**11. Do you have trouble staying asleep at night?**

☐Yes ☐No

If participant answers YES in Q11, continue to question Q11a and 11b

**11a.) How many times each week do you have trouble staying asleep at night?**

_______ times/week

**11b.) How many times each week do you get out of bed during these awakenings?**

_____________times/week

**12. How many times each week do you awake from sleep during the night to use the bathroom?**

_______times/week ☐None

**13. Do you awake from sleep during the night and eat food?**

☐Yes ☐No

If participants answer NO, skip to Q14

**13a.) How many times per week do you awake from sleep during the night and eat food?**

__________times/week

**13b.) For how long have you been getting up at this frequency to eat?**

_____years ______months

**13c.) Do you believe you need to eat in order to fall back to sleep when you wake up at nights?**

☐Yes ☐No

**13d.) How aware are you of your eating during the night?**

☐Not at all ☐Somewhat ☐Extremely

**13e) How often do you recall your eating during the night the next day?**

☐Never ☐Sometimes ☐Always

**14.) Would you consider yourself a night eater?**

☐Yes ☐No

If participant answers NO, skip to Q15.

**14a.) How upset are you about your night eating?**

☐Not at all ☐Somewhat ☐Extremely

**14b.) How much has your eating at night impaired your functioning and/or interfered with your daily life?**

☐Not at all ☐Somewhat ☐Extremely

**14c.) For how long have you been experiencing this night eating behaviour?**

______months
______years

**15.) Do you have sleep apnea?**

☐ Yes ☐No ☐Don't know

**16.) Do you work an evening or night shift?**

▫Yes ▫No

If participant answers YES, continue to Q16a and Q16b.

**16a. What type of shift do you work?**

☐Evening ☐Night ☐Rotating ☐Other

**16b. For how long have you been working this shift?**

______years
______months

**17. Have you been feeling depressed or down nearly every day?**

☐ Yes ☐No

**18. In general, when you are feeling depressed or down, is your mood lower in the:**

☐Morning ☐Afternoon ☐Evening/Nighttime ☐Not applicable

**19. Are you currently dieting to lose weight?**

☐ Yes ☐No

**20. What is your current height and weight (without clothing or shoes)?**

________ height (inch/cm)
________weight (lbs/kgs/st/bmi)

#### 4 – Muscle Dysmorphic Disorder Inventory

##### The following questions relate to exercise and how you feel about your body

**Please read through the following questions and fill in the circle that best indicates how you typically think, feel, or behave.**

**(This section will only be displayed to participants who select 'male' in question 2 or who select 'male' in question 2.1 in the demographics section)**

| | Never | Rarely | Occasionally | Frequently | Always |
|---|---|---|---|---|---|
| 1) I think my body is too small | ☐ | ☐ | ☐ | ☐ | ☐ |
| 2) I wear loose clothing so that people can't see my body | ☐ | ☐ | ☐ | ☐ | ☐ |
| 3) I hate my body | ☐ | ☐ | ☐ | ☐ | ☐ |
| 4) I wish I could get bigger | ☐ | ☐ | ☐ | ☐ | ☐ |
| 5) I think my chest is too small | ☐ | ☐ | ☐ | ☐ | ☐ |
| 6) I think my legs are too thin | ☐ | ☐ | ☐ | ☐ | ☐ |
| 7) I feel like I have too much body fat | ☐ | ☐ | ☐ | ☐ | ☐ |
| 8) I wish my arms were bigger | ☐ | ☐ | ☐ | ☐ | ☐ |
| 9) I am very shy about letting people see me with my shirt off | ☐ | ☐ | ☐ | ☐ | ☐ |
| 10) I feel anxious when I miss one or more workout days. | ☐ | ☐ | ☐ | ☐ | ☐ |
| 11) I pass up social activities with friends because of my workout schedule. | ☐ | ☐ | ☐ | ☐ | ☐ |
| 12) I feel depressed when I miss one or more workout days. | ☐ | ☐ | ☐ | ☐ | ☐ |
| 13) I pass up chances to meet new people because of my workout schedule. | ☐ | ☐ | ☐ | ☐ | ☐ |

#### 5 – Avoidant/restrictive Food Intake Disorder Screener

##### The following questions are about your food intake experiences.

**If you are concerned about answering these questions, it may be preferable to fill out this survey in the presence of someone who is able to give you support (e.g. a friend of family member).**

**Information on where to find help for the issues covered in this questionnaire is given at the end of the survey.**

| | Strongly Disagree | Disagree | Slightly Disagree | Slightly Agree | Agree | Strongly Agree |
|---|---|---|---|---|---|---|
| 1) I am a picky eater | ☐ | ☐ | ☐ | ☐ | ☐ | ☐ |
| 2) I dislike most of the foods that other people eat | ☐ | ☐ | ☐ | ☐ | ☐ | ☐ |
| 3) The list of foods that I like and will eat is shorter than the list of foods I won't eat | ☐ | ☐ | ☐ | ☐ | ☐ | ☐ |
| 4) I am not very interested in eating; I seem to have a smaller appetite than other people | ☐ | ☐ | ☐ | ☐ | ☐ | ☐ |
| 5) I have to push myself to eat regular meals throughout the day, or to eat a large enough amount of food at meals | ☐ | ☐ | ☐ | ☐ | ☐ | ☐ |
| 6) Even when I am eating a food I really like, it is hard for me to eat a large volume at meals | ☐ | ☐ | ☐ | ☐ | ☐ | ☐ |
| 7) I avoid or put off eating because I am afraid of gastrointestinal discomfort, choking, or vomiting | ☐ | ☐ | ☐ | ☐ | ☐ | ☐ |
| 8) I restrict myself to certain foods because I am afraid that other foods will cause gastrointestinal discomfort, chocking or vomiting | ☐ | ☐ | ☐ | ☐ | ☐ | ☐ |
| 9) I eat small portions bevaise I am afraid of gastrointestinal discomfort, chocking or vomiting. | ☐ | ☐ | ☐ | ☐ | ☐ | ☐ |

### 6 – Childhood Trauma and Possible Stresses and Strains of Life

**This section asks about your childhood and some possible stresses and strains of life. If this is too difficult, then *please skip to section 7 on page 47.***

| **F1) When I was growing up…** | **1 Never true** | **2 Rarely true** | **3 Sometimes true** | **4 Often** | **5 Very often true** | **6 Prefer not to answer** |
|---|---|---|---|---|---|---|
| **a.** I felt loved | ☐ | ☐ | ☐ | ☐ | ☐ | ☐ |
| **b.** People in my family hit me so hard that it left me with bruises or marks | ☐ | ☐ | ☐ | ☐ | ☐ | ☐ |
| **c.** I felt that someone in my family hated me | ☐ | ☐ | ☐ | ☐ | ☐ | ☐ |
| **d.** Someone molested me (sexually) | ☐ | ☐ | ☐ | ☐ | ☐ | ☐ |
| **e.** There was someone to take me to the doctor if I needed it | ☐ | ☐ | ☐ | ☐ | ☐ | ☐ |

| **F2) Since I was sixteen…** | **1 Never** | **2 Yes, but not in the last 12 months** | **3 Yes, within the last 12 months** | **4 Prefer not to answer** |
|---|---|---|---|---|
| **a.** I have been in a confiding relationship | ☐ | ☐ | ☐ | ☐ |
| **b.** A partner or ex-partner deliberately hit me or used violence in any other way | ☐ | ☐ | ☐ | ☐ |
| **c.** A partner or ex-partner repeatedly belittled me to the extent that I felt worthless | ☐ | ☐ | ☐ | ☐ |
| **d.** A partner or ex-partner sexually interfered with me, or forced me to have sex against my wishes | ☐ | ☐ | ☐ | ☐ |
| **e.** I have had the money to pay my rent/mortgage payment | ☐ | ☐ | ☐ | ☐ |

**F2a) If so, how often have these statements been true?**

| | **1 Never true** | **2 Rarely true** | **3 Sometimes true** | **4 Often** | **5** | **6** |
|---|---|---|---|---|---|---|

| | | | | | Very often true | Prefer not to answer |
|---|---|---|---|---|---|---|
| **a.** I have been in a confiding relationship | ☐ | ☐ | ☐ | ☐ | ☐ | ☐ |
| **b.** A partner or ex-partner deliberately hit me or used violence in any other way | ☐ | ☐ | ☐ | ☐ | ☐ | ☐ |
| **c.** A partner or ex-partner repeatedly belittled me to the extent that I felt worthless | ☐ | ☐ | ☐ | ☐ | ☐ | ☐ |
| **d.** A partner or ex-partner sexually interfered with me, or forced me to have sex against my wishes | ☐ | ☐ | ☐ | ☐ | ☐ | ☐ |
| **e.** I have had the money to pay my rent/mortgage payment | ☐ | ☐ | ☐ | ☐ | ☐ | ☐ |

| **F3) In your life, have you…?** | 1 Never | 2 Yes, but not in the last 12 months | 3 Yes, within the last 12 months | 4 Prefer not to answer |
|---|---|---|---|---|
| **a.** Been a victim of a sexual assault, whether by a stranger or someone you knew | ☐ | ☐ | ☐ | ☐ |
| **b.** Been attacked, mugged, robbed, or been the victim of a physically violent crime | ☐ | ☐ | ☐ | ☐ |
| **c.** Been in a serious accident that you believed to be life-threatening at the time | ☐ | ☐ | ☐ | ☐ |
| **d.** Witnessed a sudden violent death (e.g., murder, suicide, aftermath of an accident) | ☐ | ☐ | ☐ | ☐ |
| **e.** Been diagnosed with a life-threatening illness | ☐ | ☐ | ☐ | ☐ |
| **f)** Been involved in combat or exposed to a war-zone (either in the military or as a civilian) | ☐ | ☐ | ☐ | ☐ |

**F4) Next is a list of problems and complaints that people sometimes have in response to such extremely stressful experiences. Please indicate how much you have been bothered by that problem in the past month:**

| | 1 Not at all | 2 A little bit | 3 Moderately | 4 Quite a bit | 5 Extremely | 6 Prefer not to answer |
|---|---|---|---|---|---|---|

| | | | | | | |
|---|---|---|---|---|---|---|
| a. Repeated, disturbing memories, thoughts, or images of a stressful experience? | ☐ | ☐ | ☐ | ☐ | ☐ | ☐ |
| b. Feeling very upset when something reminded you of a stressful experience? | ☐ | ☐ | ☐ | ☐ | ☐ | ☐ |
| c. Avoiding activities or situations because they reminded you of a stressful experience? | ☐ | ☐ | ☐ | ☐ | ☐ | ☐ |
| d. Feeling distant or cut off from other people? | ☐ | ☐ | ☐ | ☐ | ☐ | ☐ |
| e. Feeling irritable or having angry outbursts? | ☐ | ☐ | ☐ | ☐ | ☐ | ☐ |
| f. Difficulty concentrating? | ☐ | ☐ | ☐ | ☐ | ☐ | ☐ |

#### 7 – General info/Study Feedback

##### These questions relate to the questionnaire you have just answered

**1) How did you find the <u>length</u> of the questionnaire?**

☐ Too brief ☐ About right

☐ Too long

**2) How did you find <u>completing</u> the questionnaire?**

☐ Not at all enjoyable ☐ Moderately enjoyable

☐ Very enjoyable

**3) Have you <u>previously</u> participated in the UK Biobank?**

☐ Yes *(Continue to 3a)* ☐ No *(Skip to 4)*

☐ Don't know *(Skip to 4)*

**3a) Did you complete the UK Biobank Mental Health Questionnaire?**

☐ Yes ☐ No

☐ Don't know

**4) How did you hear about the <u>EDGI</u>? *(Please select all that apply)***

- ☐ Facebook *(continue to question 4.1)*
- ☐ Twitter *(skip to question 4.2)*
- ☐ Blogger *(skip to question 4.3)*
- ☐ Other social media *(skip to question 4.4)*
- ☐ Search engine *(skip to question 4.5)*
- ☐ Radio *(skip to question 4.6)*
- ☐ TV *(skip to question 4.7)*
- ☐ Newspaper *(skip to question 4.8)*
- ☐ Online tabloid *(skip to question 4.9)*
- ☐ Word of mouth *(skip to question 4.10)*
- ☐ Employer or organisation *(skip to question 4.11)*
- ☐ Beat Eating Disorders *(skip to question 5)*
- ☐ Other Charity *(skip to question 4.12)*
- ☐ Clinician/GP *(skip to question 5)*
- ☐ Other *(skip to question 4.13)*

**4.1) How did you hear about the EDGI through Facebook? *(Please select all that apply)***

- ☐ Friends/family *(skip to question 5)*
- ☐ Promoted posts/advertisements *(skip to question 5)*
- ☐ Celebrity *(continue to question 4.1a)*
- ☐ Charity *(skip to question 4.1b)*
- ☐ EDGI study Facebook page *(skip to question 5)*
- ☐ Online tabloid/news posts *(skip to question 4.1c)*

**4.1.1) Which celebrity did you hear about the EDGI from?**

________________________________________

**4.1.2) Which charity did you hear about the EDGI from?**

________________________________________

**4.1.3) Which online tabloid/newspaper did you hear about the EDGI from?**

________________________________________

**4.2) How did you hear about the EDGI through twitter? *(Please select all that apply)***

- ☐ Friends/family *(skip to question 5)*
- ☐ Promoted posts/advertisements *(skip to question 5)*
- ☐ Celebrity *(continue to question 4.2a)*
- ☐ Charity *(skip to question 4.2b)*
- ☐ EDGI study Facebook page *(skip to question 5)*
- ☐ Online tabloid/news posts *(skip to question 4.2c)*

**4.2.1) Which celebrity did you hear about the EDGI from?**

________________________________________

**4.2.2) Which charity did you hear about the EDGI from?**

________________________________________

**4.2.3) Which online tabloid/newspaper did you hear about the EDGI from?**

________________________________________

**4.3) Which blogger did you hear about the EDGI from?**

___________________________________________________

**4.4) Which other social media did you hear about the EDGI from?**

___________________________________________________

**4.5) On which search engine did you hear about the EDGI from?**

- ☐ Google *(skip to question 5)*
- ☐ Bing *(skip to question 5)*
- ☐ Yahoo *(skip to question 5)*
- ☐ Ask *(skip to question 5)*
- ☐ AOL *(skip to question 5)*
- ☐ MSN *(skip to question 5)*
- ☐ DuckDuckGo *(skip to question 5)*
- ☐ Other *(continue to question 4.5.1)*

**4.5.1) On which other search engine did you hear about the EDGI from?**

___________________________________________________

**4.6) On which radio station/show(s) did you hear about the EDGI from?**

___________________________________________________

**4.7) On which TV channel/show(s) did you hear about the EDGI from?**

___________________________________________________

**4.8) In which newspaper(s) did you hear about the EDGI from?**

___________________________________________________

**4.9) In which online tabloid(s) did you hear about the EDGI from?**

___________________________________________________

**4.10) Who did you hear about the EDGI from?**

- ☐ Friend(s) *(skip to question 5)*
- ☐ Colleague(s) *(skip to question 5)*
- ☐ Family *(skip to question 5)*
- ☐ Other *(skip to question 5)*

**4.11) Which employer/organisation did you hear about the EDGI from?**

___________________________________________________

**4.12) Which charity did you hear about the EDGI from?**

___________________________________________________

**4.13) Please tell us more about how you heard about the EDGI from?**

**5) Is there any other information you would like to share that relates to this study?**

**If you wish to find out more information about this questionnaire, you can contact one of the EDGI researchers on Freephone 0800 917 6016**

If you feel you need any further help with the issues in this questionnaire, we recommend talking it through with someone you trust, including your GP. For more information and support please visit MIND.

If you would like to speak with a trained support worker who can offer help and information on dealing with eating disorders, please contact the Beat helpline at 0808 801 0677 which is open everyday from 12pm-8pm during the week, and 4pm-8pm on weekends and bank holiday.

If you are in distress and need urgent help or advice, please contact someone as soon as possible, such as the Samaritans on 116 123 (or see their website) or the SLaM 24-hour mental health support line on 0800 731 2864 (or visit here).

**Thank you very much for taking the time to complete this questionnaire.**

We are now assessing your eligibility to take part in the EDGI study. If you are eligible, you will receive your saliva kit in the post in the next 10-15 working days.

In the meantime, please feel free to visit the optional projects page where you can complete a variety of additional questionnaires about your experiences with mental and physical health.

Please continue by pressing the next arrow below

**Please note that you will not be automatically directed away from this page, but your survey response has been submitted.**
