## Supplementary material for "The Eating Disorders Genetics Initiative (EDGI) United Kingdom": EDGI UK Optional Questionnaires

### Eating Disorders Genetics Initiative (EDGI) United Kingdom: Optional Questionnaires

### Contents

| Measures | Page |
|---|---|
| **Module 1: Eating Behaviours and Physical Activity** | |
| Yale Food Addiction Scale (Y-FAS) (Gearhardt et al., 2009) | 1-2 |
| Adult Eating Behaviours Questionnaire (AEBQ) (Hunot et al., 2016) | 3-8 |
| Clinical Impairment Assessment (CIA) (Jenkins, 2013) | 9 |
| Eating Disorder Examination Questionnaire (EDE-Q6) (Carey et al., 2019) | 10-12 |
| Self-Regulation of Eating Behaviours Questionnaire (SREBQ) (Kliemann et al., 2016) | 13 |
| Compulsive Exercise Test (CET) (Taranis et al., 2011) | 14-15 |
| **Module 2: Thoughts and Behaviours** | |
| Obsessive Compulsive Inventory-Revised (OCI-R) (Foa et al., 2002) | 16-17 |
| Adult ADHD Self-Report Scale (ASRS) (Kessler et al., 2005) | 18-20 |
| Difficulties in Emotion Regulation Scale (DERS-16) (Bjureberg et al., 2016) | 21 |
| Life Events Checklist for DSM-5 (LEC-5) (Weathers et al., n.d.) | 22-28 |
| Autism Spectrum Quotient (AQ-10) (Allison et al., 2012) | 29 |
| Assessment for lifetime MDD and GAD based on the WHO Composite International Diagnostic Interview short-form (CIDI-SF) (Patten et al., 2000),ICD-11 (Kogan et al., 2016) and other anxiety disorders based on the DSM-V criteria modified for the GLAD Study (Davies et al., 2019) | 30-86 |
| Generalised Anxiety Disorder 7-item (GAD-7) (Spitzer et al., 2006) | 87 |
| Mood Disorder Questionnaire (MDQ) (Hirschfeld, 2002) | 88-89 |
| WHO Quality of Life (modified short version for subjective well being) (WHOQOL) (Group, Whoqol, 1994) | 90-98 |
| Patient Health Questionnaire 9-item (PHQ-9) (Kroenke et al., 2001) | 99 |
| Post-traumatic Stress Disorder Checklist for DSM-5 (PCL-5) (Blevins et al., 2015) | 100-101 |
| **Module 3: Lifestyle** | |
| Family Structure (non-validated questionnaire adapted from the Australian Genetics of Depression Study, (Byrne et al., 2019)) | 102-120 |
| Alcohol Use Disorders Identification Test (AUDIT) (Saunders et al., 1993) | 121-122 |
| Drug Use Disorders Identification Test (DUDIT) (Bergman et al., 2003) | 123-124 |
| Drugs and Addiction (non-validated questionnaire from Australian Genetics of Depression Study) (Byrne et al., 2019) | 125-135 |
| Games and Gambling (non-validated questionnaire adapted from the Australian Genetics of Depression Study) (Byrne et al., 2019) | 136-158 |
| Headaches and Migraines (non-validated questionnaire adapted from the Australian Genetics of Depression Study) (Byrne et al., 2019) | 159-163 |
| Work and Sleep (non-validated questionnaire adapted from the Australian Genetics of Depression Study) (Byrne et al., 2019) | 164-176 |
| **Module 4: Quality of Life** | |
| 12-Item Short Form Survey (SF-12) (Ware et al., 1996) | 177-179 |
| Eating Disorder Quality of Life (EDQOL) (Engel et al., 2006) | 180-184 |
| **Module 5: Personality** | |
| Personality Inventory for DSM-5 Brief Form (PID-5-BF) (Krueger et al., 2014) | 185-187 |

| | |
|---|---|
| "Personal Standards" from Multidimensional Personality Scale (MPS) (Clavin et al., 1996) | 188-198 |
| **Module 6: Treatment** | |
| Eating Disorder Treatment Questionnaire (non-validated based on New Zealand treatment questionnaire) | 199-234 |
| Medication usage questionnaire (non-validated questionnaire adapted from the Australian Genetics of Depression Study) (Byrne et al., 2019) | 235-241 |

**Yale Food Addiction Scale**

**Gearhardt, Corbin, Brownell, 2009**
**Contact:**

**This survey asks about your eating habits in the past year. People sometimes have difficulty controlling their intake of certain foods such as:**

- **Sweets like ice cream, chocolate, doughnuts, cookies, cake, candy, ice cream**
- **Starches like white bread, rolls, pasta, and rice**
- **Salty snacks like chips, pretzels, and crackers**
- **Fatty foods like steak, bacon, hamburgers, cheeseburgers, pizza, and French fries**
- **Sugary drinks like soda pop**

**When the following questions ask about "CERTAIN FOODS" please think of ANY food similar to those listed in the food group or ANY OTHER foods you have had a problem with in the past year**

| IN THE PAST 12 MONTHS: | Never | Once a month | 2-4 times a month | 2-3 times a week | 4 or more times or daily |
|---|---|---|---|---|---|
| 1. I find that when I start eating certain foods, I end up eating much more than planned | 0 | 1 | 2 | 3 | 4 |
| 2. I find myself continuing to consume certain foods even though I am no longer hungry | 0 | 1 | 2 | 3 | 4 |
| 3. I eat to the point where I feel physically ill | 0 | 1 | 2 | 3 | 4 |
| 4. Not eating certain types of food or cutting down on certain types of food is something I worry about | 0 | 1 | 2 | 3 | 4 |
| 5. I spend a lot of time feeling sluggish or fatigued from overeating | 0 | 1 | 2 | 3 | 4 |
| 6. I find myself constantly eating certain foods throughout the day | 0 | 1 | 2 | 3 | 4 |
| 7. I find that when certain foods are not available, I will go out of my way to obtain them. For example, I will drive to the store to purchase certain foods even though I have other options available to me at home. | 0 | 1 | 2 | 3 | 4 |
| 8. There have been times when I consumed certain foods so often or in such large quantities that I started to eat food instead of working, spending time with my family or friends, or engaging in other important activities or recreational activities I enjoy. | 0 | 1 | 2 | 3 | 4 |
| 9. There have been times when I consumed certain foods so often or in such large quantities that I spent time dealing with negative feelings from overeating instead of working, spending time with my family or friends, or engaging in other important activities or recreational activities I enjoy. | 0 | 1 | 2 | 3 | 4 |
| 10. There have been times when I avoided professional or social situations where certain foods were available, because I was afraid I would overeat. | 0 | 1 | 2 | 3 | 4 |
| 11. There have been times when I avoided professional or social situations because I was not able to consume certain foods there. | 0 | 1 | 2 | 3 | 4 |
| 12. I have had withdrawal symptoms such as agitation, anxiety, or other physical symptoms when I cut down or stopped eating certain foods. (Please do NOT include withdrawal symptoms caused by cutting down on caffeinated beverages such as soda pop, coffee, tea, energy drinks, etc.) | 0 | 1 | 2 | 3 | 4 |
| 13. I have consumed certain foods to prevent feelings of anxiety, agitation, or other physical symptoms that were developing. (Please do NOT include consumption of caffeinated beverages such as soda pop, coffee, tea, energy drinks, etc.) | 0 | 1 | 2 | 3 | 4 |
| 14. I have found that I have elevated desire for or urges to consume certain foods when I cut down or stop eating them. | 0 | 1 | 2 | 3 | 4 |
| 15. My behavior with respect to food and eating causes significant distress. | 0 | 1 | 2 | 3 | 4 |
| 16. I experience significant problems in my ability to function effectively (daily routine, job/school, social activities, family activities, health difficulties) because of food and eating. | 0 | 1 | 2 | 3 | 4 |

| IN THE PAST 12 MONTHS: | NO | YES |
|---|---|---|
| 17. My food consumption has caused significant psychological problems such as depression, anxiety, self-loathing, or guilt. | 0 | 1 |
| 18. My food consumption has caused significant physical problems or made a physical problem worse. | 0 | 1 |
| 19. I kept consuming the same types of food or the same amount of food even though I was having emotional and/or physical problems. | 0 | 1 |
| 20. Over time, I have found that I need to eat more and more to get the feeling I want, such as reduced negative emotions or increased pleasure. | 0 | 1 |
| 21. I have found that eating the same amount of food does not reduce my negative emotions or increase pleasurable feelings the way it used to. | 0 | 1 |
| 22. I want to cut down or stop eating certain kinds of food. | 0 | 1 |
| 23. I have tried to cut down or stop eating certain kinds of food. | 0 | 1 |
| 24. I have been successful at cutting down or not eating these kinds of food | 0 | 1 |

| | | | | | |
|---|---|---|---|---|---|
| 25. How many times in the past year did you try to cut down or stop eating certain foods altogether? | 1 or fewer times | 2 times | 3 times | 4 times | 5 or more times |

### Adult Eating Behaviour Questionnaire

#### Please read each statement and tick the box most appropriate to you

| | Strongly disagree | Disagree | Neither agree or disagree | Agree | Strongly agree |
|---|---|---|---|---|---|
| I love food | ☐ | ☐ | ☐ | ☐ | ☐ |
| I often decide that I don't like a food, before tasting it | ☐ | ☐ | ☐ | ☐ | ☐ |
| I enjoy eating | ☐ | ☐ | ☐ | ☐ | ☐ |
| I look forward to mealtimes | ☐ | ☐ | ☐ | ☐ | ☐ |
| I eat more when I'm annoyed | ☐ | ☐ | ☐ | ☐ | ☐ |
| I often notice my stomach rumbling | ☐ | ☐ | ☐ | ☐ | ☐ |
| I refuse new foods at first | ☐ | ☐ | ☐ | ☐ | ☐ |
| I eat more when I'm worried | ☐ | ☐ | ☐ | ☐ | ☐ |
| If I miss a meal I get irritable | ☐ | ☐ | ☐ | ☐ | ☐ |
| I eat more when I'm upset | ☐ | ☐ | ☐ | ☐ | ☐ |
| I often leave food on my plate at the end of a meal | ☐ | ☐ | ☐ | ☐ | ☐ |
| I enjoy tasting new foods | ☐ | ☐ | ☐ | ☐ | ☐ |
| I often feel hungry when I am with someone who is eating | ☐ | ☐ | ☐ | ☐ | ☐ |
| I often finish my meals quickly | ☐ | ☐ | ☐ | ☐ | ☐ |
| I eat less when I'm worried | ☐ | ☐ | ☐ | ☐ | ☐ |
| I eat more when I´m anxious | ☐ | ☐ | ☐ | ☐ | ☐ |
| Given the choice, I would eat most of the time | ☐ | ☐ | ☐ | ☐ | ☐ |

| | Strongly disagree | Disagree | Neither agree or disagree | Agree | Strongly agree |
|---|---|---|---|---|---|
| I eat less when I'm angry | ☐ | ☐ | ☐ | ☐ | ☐ |
| I am interested in tasting new food I haven't tasted before | ☐ | ☐ | ☐ | ☐ | ☐ |
| I eat less when I'm upset | ☐ | ☐ | ☐ | ☐ | ☐ |
| I eat more when I'm angry | ☐ | ☐ | ☐ | ☐ | ☐ |
| I am always thinking about food | ☐ | ☐ | ☐ | ☐ | ☐ |
| I often get full before my meal is finished | ☐ | ☐ | ☐ | ☐ | ☐ |
| I enjoy a wide variety of foods | ☐ | ☐ | ☐ | ☐ | ☐ |
| I am often last at finishing a meal | ☐ | ☐ | ☐ | ☐ | ☐ |
| I eat more and more slowly during the course of a meal | ☐ | ☐ | ☐ | ☐ | ☐ |
| I eat less when I'm annoyed | ☐ | ☐ | ☐ | ☐ | ☐ |
| I often feel so hungry that I have to eat something right away | ☐ | ☐ | ☐ | ☐ | ☐ |
| I eat slowly | ☐ | ☐ | ☐ | ☐ | ☐ |
| I cannot eat a meal if I have had a snack just before | ☐ | ☐ | ☐ | ☐ | ☐ |
| I get full up easily | ☐ | ☐ | ☐ | ☐ | ☐ |
| I often feel hungry | ☐ | ☐ | ☐ | ☐ | ☐ |
| When I see or smell food that I like, it makes me want to eat | ☐ | ☐ | ☐ | ☐ | ☐ |
| If my meals are delayed I get light-headed | ☐ | ☐ | ☐ | ☐ | ☐ |
| I eat less when I'm anxious | ☐ | ☐ | ☐ | ☐ | ☐ |

#### Adult Eating Behaviour Questionnaire - Scoring information

| | | Strongly disagree | Disagree | Neither agree or disagree | Agree | Strongly agree |
|---|---|---|---|---|---|---|
| EF | I love food | ◻ | ◻ | ◻ | ◻ | ◻ |
| FF | I often decide that I don’t like a food, before tasting it | ◻ | ◻ | ◻ | ◻ | ◻ |
| EF | I enjoy eating | ◻ | ◻ | ◻ | ◻ | ◻ |
| EF | I look forward to mealtimes | ◻ | ◻ | ◻ | ◻ | ◻ |
| EOE | I eat more when I'm annoyed | ◻ | ◻ | ◻ | ◻ | ◻ |
| H | I often notice my stomach rumbling | ◻ | ◻ | ◻ | ◻ | ◻ |
| FF | I refuse new foods at first | ◻ | ◻ | ◻ | ◻ | ◻ |
| EOE | I eat more when I'm worried | ◻ | ◻ | ◻ | ◻ | ◻ |
| H | If I miss a meal I get irritable | ◻ | ◻ | ◻ | ◻ | ◻ |
| EOE | I eat more when I'm upset | ◻ | ◻ | ◻ | ◻ | ◻ |
| SR | I often leave food on my plate at the end of a meal | ◻ | ◻ | ◻ | ◻ | ◻ |
| FF* | I enjoy tasting new foods | ◻ | ◻ | ◻ | ◻ | ◻ |
| FR | I often feel hungry when I am with someone who is eating | ◻ | ◻ | ◻ | ◻ | ◻ |
| SE* | I often finish my meals quickly | ◻ | ◻ | ◻ | ◻ | ◻ |
| EUE | I eat less when I'm worried | ◻ | ◻ | ◻ | ◻ | ◻ |
| EOE | I eat more when I´m anxious | ◻ | ◻ | ◻ | ◻ | ◻ |
| FR | Given the choice, I would eat most of the time | ◻ | ◻ | ◻ | ◻ | ◻ |
| EUE | I eat less when I'm angry | ◻ | ◻ | ◻ | ◻ | ◻ |
| FF* | I am interested in tasting new food I haven’t tasted before | ◻ | ◻ | ◻ | ◻ | ◻ |
| EUE | I eat less when I'm upset | ◻ | ◻ | ◻ | ◻ | ◻ |
| EOE | I eat more when I'm angry | ◻ | ◻ | ◻ | ◻ | ◻ |
| FR | I am always thinking about food | ◻ | ◻ | ◻ | ◻ | ◻ |
| SR | I often get full before my meal is finished | ◻ | ◻ | ◻ | ◻ | ◻ |
| FF* | I enjoy a wide variety of foods | ◻ | ◻ | ◻ | ◻ | ◻ |
| SE | I am often last at finishing a meal | ◻ | ◻ | ◻ | ◻ | ◻ |
| SE | I eat more and more slowly during the course of a meal | ◻ | ◻ | ◻ | ◻ | ◻ |

| | | Strongly disagree | Disagree | Neither agree or disagree | Agree | Strongly agree |
|---|---|---|---|---|---|---|
| EUE | I eat less when I'm annoyed | □ | □ | □ | □ | □ |
| H | I often feel so hungry that I have to eat something right away | □ | □ | □ | □ | □ |
| SE | I eat slowly | □ | □ | □ | □ | □ |
| SR | I cannot eat a meal if I have had a snack just before | □ | □ | □ | □ | □ |
| SR | I get full up easily | □ | □ | □ | □ | □ |
| H | I often feel hungry | □ | □ | □ | □ | □ |
| FR | When I see or smell food that I like, it makes me want to eat | □ | □ | □ | □ | □ |
| H | If my meals are delayed I get light-headed | □ | □ | □ | □ | □ |
| EUE | I eat less when I'm anxious | □ | □ | □ | □ | □ |

* Reversed items

| | | |
|---|---|---|
| Enjoyment of food | = | item mean EF |
| Emotional over-eating | = | item mean EOE |
| Emotional under-eating | = | item mean EUE |
| Food fussiness | = | item mean FF |
| Food responsiveness | = | item mean FR |
| Slowness in eating | = | item mean SE |
| Hunger | = | item mean H |
| Satiety responsiveness | = | item mean SR |

Strongly disagree =1, Disagree = 2,Neither agree nor disagree = 3, Agree =4, Strongly agree =5

### Adult Eating Behaviour Questionnaire - Scoring information

| | | Strongly disagree | Disagree | Neither agree or disagree | Agree | Strongly agree |
|---|---|---|---|---|---|---|
| EF | I love food | ☐ | ☐ | ☐ | ☐ | ☐ |
| EF | I enjoy eating | ☐ | ☐ | ☐ | ☐ | ☐ |
| EF | I look forward to mealtimes | ☐ | ☐ | ☐ | ☐ | ☐ |
| EOE | I eat more when I'm annoyed | ☐ | ☐ | ☐ | ☐ | ☐ |
| EOE | I eat more when I'm worried | ☐ | ☐ | ☐ | ☐ | ☐ |
| EOE | I eat more when I'm upset | ☐ | ☐ | ☐ | ☐ | ☐ |
| EOE | I eat more when I´m anxious | ☐ | ☐ | ☐ | ☐ | ☐ |
| EOE | I eat more when I'm angry | ☐ | ☐ | ☐ | ☐ | ☐ |
| EUE | I eat less when I'm worried | ☐ | ☐ | ☐ | ☐ | ☐ |
| EUE | I eat less when I'm angry | ☐ | ☐ | ☐ | ☐ | ☐ |
| EUE | I eat less when I'm upset | ☐ | ☐ | ☐ | ☐ | ☐ |
| EUE | I eat less when I'm annoyed | ☐ | ☐ | ☐ | ☐ | ☐ |
| EUE | I eat less when I'm anxious | ☐ | ☐ | ☐ | ☐ | ☐ |
| FF | I often decide that I don't like a food, before tasting it | ☐ | ☐ | ☐ | ☐ | ☐ |
| FF | I refuse new foods at first | ☐ | ☐ | ☐ | ☐ | ☐ |
| FF* | I enjoy tasting new foods | ☐ | ☐ | ☐ | ☐ | ☐ |
| FF* | I am interested in tasting new food I haven't tasted before | ☐ | ☐ | ☐ | ☐ | ☐ |
| FF* | I enjoy a wide variety of foods | ☐ | ☐ | ☐ | ☐ | ☐ |
| FR | I often feel hungry when I am with someone who is eating | ☐ | ☐ | ☐ | ☐ | ☐ |
| FR | Given the choice, I would eat most of the time | ☐ | ☐ | ☐ | ☐ | ☐ |
| FR | I am always thinking about food | ☐ | ☐ | ☐ | ☐ | ☐ |
| FR | When I see or smell food that I like, it makes me want to eat | ☐ | ☐ | ☐ | ☐ | ☐ |
| H | I often notice my stomach rumbling | ☐ | ☐ | ☐ | ☐ | ☐ |
| H | If I miss a meal I get irritable | ☐ | ☐ | ☐ | ☐ | ☐ |
| H | I often feel so hungry that I have to eat something right away | ☐ | ☐ | ☐ | ☐ | ☐ |
| H | I often feel hungry | ☐ | ☐ | ☐ | ☐ | ☐ |

| | | Strongly disagree | Disagree | Neither agree or disagree | Agree | Strongly agree |
|---|---|---|---|---|---|---|
| H | If my meals are delayed I get light-headed | ☐ | ☐ | ☐ | ☐ | ☐ |
| SE | I am often last at finishing a meal | ☐ | ☐ | ☐ | ☐ | ☐ |
| SE | I eat more and more slowly during the course of a meal | ☐ | ☐ | ☐ | ☐ | ☐ |
| SE | I eat slowly | ☐ | ☐ | ☐ | ☐ | ☐ |
| SE* | I often finish my meals quickly | ☐ | ☐ | ☐ | ☐ | ☐ |
| SR | I often leave food on my plate at the end of a meal | ☐ | ☐ | ☐ | ☐ | ☐ |
| SR | I often get full before my meal is finished | ☐ | ☐ | ☐ | ☐ | ☐ |
| SR | I cannot eat a meal if I have had a snack just before | ☐ | ☐ | ☐ | ☐ | ☐ |
| SR | I get full up easily | ☐ | ☐ | ☐ | ☐ | ☐ |

#### CLINICAL IMPAIRMENT ASSESSMENT QUESTIONNAIRE (CIA 3.0)

**Copyright Bohn and Fairburn, 2008**

**INSTRUCTIONS**
**Please place an 'X' in the column which best describes how your eating habits, exercising or feelings about your eating, shape or weight have affected your life over the past four weeks (28 days). Thank you.**

| | **Over the past 28 days, to what extent have your**<br>**… eating habits**<br>**… exercising**<br>**… or your feelings about your eating, shape or weight …..** | **Not at all** | **A little** | **Quite a bit** | **A lot** |
|---|---|---|---|---|---|
| 1 | ... made it difficult to concentrate? | | | | |
| 2 | ... made you feel critical of yourself? | | | | |
| 3 | ... stopped you going out with others? | | | | |
| 4 | ... affected your work performance (if applicable)? | | | | |
| 5 | ... made you forgetful? | | | | |
| 6 | ... affected your ability to make everyday decisions? | | | | |
| 7 | ... interfered with meals with family or friends? | | | | |
| 8 | ... made you upset? | | | | |
| 9 | ... made you feel ashamed of yourself? | | | | |
| 10 | ... made it difficult to eat out with others? | | | | |
| 11 | ... made you feel guilty? | | | | |
| 12 | ... interfered with you doing things you used to enjoy? | | | | |
| 13 | ... made you absent-minded? | | | | |
| 14 | ... made you feel a failure? | | | | |
| 15 | ... interfered with your relationships with others? | | | | |
| 16 | ... made you worry? | | | | |

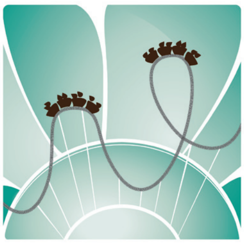

#### *Eating Disorder examination questionnaire (EDE-Q 6.0)*

**Instructions: The following questions are concerned with the past four weeks (28 days) only.**

**Please read each question carefully. Please answer all the questions. Thank you.**

**Questions 1 to 12: Please circle the appropriate number on the right. Remember that the questions only refer to the past four weeks (28 days) only.**

| | ON HOW MANY OF THE PAST 28 DAYS ... | NO DAYS | 1-5 DAYS | 6-12 DAYS | 13-15 DAYS | 16-22 DAYS | 23-27 DAYS | EVERY DAY |
|---|---|---|---|---|---|---|---|---|
| 1 | Have you been deliberately **trying** to limit the amount of food you eat to influence your shape or weight (whether or not you have succeeded)? | 0 | 1 | 2 | 3 | 4 | 5 | 6 |
| 2 | Have you gone for long periods of time (8 waking hours or more) without eating anything at all in order to influence your shape or weight? | 0 | 1 | 2 | 3 | 4 | 5 | 6 |
| 3 | Have you **tried** to exclude from your diet any foods that you like in order to influence your shape or weight (whether or not you have succeeded)? | 0 | 1 | 2 | 3 | 4 | 5 | 6 |
| 4 | Have you **tried** to follow definite rules regarding your eating (for example, a calorie limit) in order to influence your shape or weight (whether or not you have succeeded)? | 0 | 1 | 2 | 3 | 4 | 5 | 6 |
| 5 | Have you had a definite desire to have an **empty** stomach with the aim of influencing your shape or weight? | 0 | 1 | 2 | 3 | 4 | 5 | 6 |
| 6 | Have you had a definite desire to have a **totally flat** stomach? | 0 | 1 | 2 | 3 | 4 | 5 | 6 |
| 7 | Has thinking about **food, eating or calories** made it very difficult to concentrate on things you are interested in (for example, working, following a conversation, or reading)? | 0 | 1 | 2 | 3 | 4 | 5 | 6 |
| 8 | Has thinking about **shape or weight** made it very difficult to concentrate on things you are interested in (for example, working, following a conversation, or reading)? | 0 | 1 | 2 | 3 | 4 | 5 | 6 |
| 9 | Have you had a definite fear of losing control over eating? | 0 | 1 | 2 | 3 | 4 | 5 | 6 |
| 10 | Have you had a definite fear that you might gain weight? | 0 | 1 | 2 | 3 | 4 | 5 | 6 |
| 11 | Have you felt fat? | 0 | 1 | 2 | 3 | 4 | 5 | 6 |
| 12 | Have you had a strong desire to lose weight? | 0 | 1 | 2 | 3 | 4 | 5 | 6 |

**PAGE 1/3 PLEASE GO TO THE NEXT PAGE**

© 2008 Christopher G Fairburn and Sarah Beglin

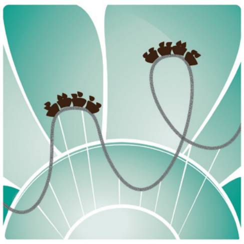

### *Eating Disorder examination questionnaire (EDE-Q 6.0)*

**Questions 13-18: Please fill in the appropriate number in the boxes on the right. Remember that the questions only refer to th past four weeks (28 days).**

**Over the past four weeks (28 days)….**

| | |
|---|---|
| 13 | Over the past 28 days, how many **times** have you eaten what other people would regards as an **unusually large amount of food** (given the circumstances)? |
| 14 | … On how many of these times did you have a sense of having lost control over your eating (at the time you were eating)? |
| 15 | Over the past 28 days, on how many **DAYS** have such episodes of overeating occurred (i.e. you have eaten an unusually large amount of food **and** have had a sense of loss of control at the time)? |
| 16 | Over the past 28 days, how many **times** have you made yourself sick (vomit) as a means of controlling your shape or weight? |
| 17 | Over the past 28 days, how many **times** have you taken laxatives as a means of controlling your shape or weight? |
| 18 | Over the past 28 days, how many **times** have you exercised in a "driven" or "compulsive" way as a means of controlling your weight, shape or amount of fat, or to burn off calories? |

**Questions 19 to 21: Please circle the appropriate number. <u>Please note that for these questions the term "binge eating" means</u> eating what others would regard as an unusually large amount of food for the circumstances, accompanied by a sense of having lost control over eating.**

| | | NO DAYS | 1-5 DAYS | 6-12 DAYS | 13-15 DAYS | 16-22 DAYS | 23-27 DAYS | EVERY DAY |
|---|---|---|---|---|---|---|---|---|
| 19 | Over the past 28 days, on how many days have you eaten in secret (ie, furtively)? … Do not count episodes of binge eating. | 0 | 1 | 2 | 3 | 4 | 5 | 6 |
| | | None of the times | A few of the times | Less than half | Half of the times | More than half | Most of the time | Every time |
| 20 | On what proportion of the times that you have eaten have you felt guilty (felt that you've done wrong) because of its effect on your shape or weight? … Do not count episodes of binge eating. | 0 | 1 | 2 | 3 | 4 | 5 | 6 |
| | | | Not at all | Slightly | Moderately | | Markedly | |
| 21 | Over the past 28 days, how concerned have you been about other people seeing you eat? … Do not count episodes of binge eating. | 0 | 1 | 2 | 3 | 4 | 5 | 6 |

**PAGE 2/3 PLEASE GO TO THE NEXT PAGE**

© 2008 Christopher G Fairburn and Sarah Beglin

*Eating Disorder examination questionnaire (EDE-Q 6.0)*

**Questions 22 to 28: Please circle the appropriate number on the right. Remember that the questions only refer to the past four weeks (28 days).**

| | **ON HOW MANY OVER THE PAST 28 DAYS ...** | **NOT AT ALL** | **SLIGHTLY** | | **MODERATELY** | | **MARKEDLY** | |
|---|---|---|---|---|---|---|---|---|
| 22 | Has your **weight** influenced how you think about (judge) yourself as a person? | 0 | 1 | 2 | 3 | 4 | 5 | 6 |
| 23 | Has your **shape** influenced how you think about (judge) yourself as a person? | 0 | 1 | 2 | 3 | 4 | 5 | 6 |
| 24 | How much would it have upset you if you had been asked to weigh yourself once a week (no more, or less, often) for the next four weeks? | 0 | 1 | 2 | 3 | 4 | 5 | 6 |
| 25 | How dissatisfied have you been with your **weight**? | 0 | 1 | 2 | 3 | 4 | 5 | 6 |
| 26 | How dissatisfied have you been with your **shape**? | 0 | 1 | 2 | 3 | 4 | 5 | 6 |
| 27 | How uncomfortable have you felt seeing your body (for example, seeing your shape in the mirror, in a shop window reflection, while undressing or taking a bath or shower)? | 0 | 1 | 2 | 3 | 4 | 5 | 6 |
| 28 | How uncomfortable have you felt about **others** seeing your shape or figure (for example, in communal changing rooms, when swimming, or wearing tight clothes)? | 0 | 1 | 2 | 3 | 4 | 5 | 6 |

**What is your weight at present? (Please give your best estimate.):** ..............................

**What is your height? (Please give your best estimate.):** ..............................

**If female: Over the past three to four months have you missed any menstrual periods?:** **YES** ○ **NO** ○

**If so, how many?:** ○

**Have you been taking the "pill"?:** **YES** ○ **NO** ○

**PAGE 3/3**

**THANK YOU**

© 2008 Christopher G Fairburn and Sarah Beglin

| ID: |
|---|

### Self-Regulation of Eating Behaviour Questionnaire (SREBQ)

**1. Do you find any of these foods tempting (that is, do you want to eat more of them than you think you should)? (Tick all those that you find tempting)**

| | | | | | |
|---|---|---|---|---|---|
| Chocolate | ☐ | Fizzy drinks | ☐ | Pizza | ☐ |
| Crisps | ☐ | Biscuits | ☐ | Fried foods | ☐ |
| Cakes | ☐ | Sweets | ☐ | Chips | ☐ |
| Ice cream | ☐ | Popcorn | ☐ | Other foods | ☐ |
| Bread/toast | ☐ | Pastries | ☐ | I don't find any food tempting | ☐ |

**If you have ticked 'Other foods', please specify:**

**2. Do you intend NOT to eat too much of the foods you find tempting in the previous question?**

| | |
|---|---|
| Yes | ☐ |
| No | ☐ |

**3. Do you intend to have a healthy diet?**

| | |
|---|---|
| Yes | ☐ |
| No | ☐ |

**4. Please read the following statements and tick the boxes most appropriate to you.**

For the next few questions, please, understand that:

- 'Tempting foods' are any food you want to eat more of than you think your should.
- 'Eating intentions' refer to the way you are aiming to eat, for example you may intend to avoid tempting foods or eat healthy foods.

| | **Never** | **Rarely** | **Sometimes** | **Often** | **Always** |
|---|---|---|---|---|---|
| **A**. I give up too easily on my eating intentions | ☐ | ☐ | ☐ | ☐ | ☐ |
| **B**. I'm good at resisting tempting food | ☐ | ☐ | ☐ | ☐ | ☐ |
| **C**. I easily get distracted from the way I intend to eat | ☐ | ☐ | ☐ | ☐ | ☐ |
| **D**. If I am not eating in the way I intend to, I make changes | ☐ | ☐ | ☐ | ☐ | ☐ |
| **E**. I find it hard to remember what I have eaten throughout the day | ☐ | ☐ | ☐ | ☐ | ☐ |

#### The Compulsive Exercise Test (CET)

The CET is the first measure of problematic exercise that has been developed specifically for use in eating disorders research and assessment, and within a cognitive-behavioural framework. The following references are the key publications:

1. *Taranis, L., Touyz, S., & Meyer, C. (2011). Disordered eating and exercise: Development and preliminary validation of the Compulsive Exercise Test (CET.) European Eating Disorders Review, 19, 256-268*
2. *Goodwin, H., Haycraft, E., Taranis, L. & Meyer, C. (2011). Psychometric evaluation of the Compulsive Exercise Test (CET) in an adolescent population: Links with eating psychopathology. European Eating Disorders Review.19, 269-279.*
3. *Meyer, C., Taranis, L., Goodwin, H., & Haycraft, E. (2011). Compulsive exercise and eating disorders. European Eating Disorders Review, 19 174-189.*

The CET is a 24-item self-report measure designed to assess the core features of excessive exercise in the eating disorders; *compulsivity* (e.g. continuing to exercise despite illness or injury, lack of exercise enjoyment, the experience of extreme guilt when unable to exercise, making up for missed exercise sessions), *affect regulation* (e.g. the positive and negative reinforcement properties of exercise), *weight and shape driven exercise* (e.g. exercising solely to burn calories, compensatory exercise such as debting), and *exercise rigidity* (rigid adherence to a strict and repetitive exercise routine). Items were generated from a comprehensive appraisal of the eating disorder and excessive exercise literature, consultation with clinical eating disorder specialists, interviews with eating disorder patients, and a critical review of existing scales, and were included based on theoretical relevance and clinical specificity.

The CET uses a 6-point Likert scale anchored by 0 (never true) and 5 (always true) with higher scores indicative of greater pathology. Factor analysis revealed 5 factors explaining 63.5% of the variance. These were used to construct the 5 subscales of: avoidance and rule-driven behaviour, weight control exercise, mood improvement, lack of exercise enjoyment, and exercise rigidity. Initial validation results are encouraging with good internal consistency, content validity, and concurrent validity of the CET. The CET also demonstrates strong positive associations with measures of eating pathology and known correlates of disordered eating. It is concluded that the CET could be a reliable and valid instrument for use in both clinical and research settings.

**CET Scoring Criteria:**

- Items 8 and 12 are reverse scored.
- Subscale scores are obtained by summing the scores for all items in the subscale and dividing by the number of items (mean score).
- CET total score is calculated by summing the mean scores for all subscales.

| *Subscale Items* | |
|---|---|
| Avoidance and rule-driven behaviour | 9, 10, 11, 15, 16, 20, 22, 23 |
| Weight control exercise | 2, 6, 8, 13, 18 |
| Mood improvement | 1, 4, 14, 17, 24 |
| Lack of exercise enjoyment | 5, 12, 21 |
| Exercise rigidity | 3, 7, 19 |

**CET**

**Instructions**

Listed below are a series of statements regarding exercise. Please read each statement carefully and circle the number that best indicates how true each statement is of you. Please answer <u>all</u> the questions as honestly as you can.

| Never true | Rarely true | Sometimes true | Often true | Usually true | Always true |
|---|---|---|---|---|---|
| **0** | **1** | **2** | **3** | **4** | **5** |

| | | | | | | | |
|---|---|---|---|---|---|---|---|
| 1) | I feel happier and/or more positive after I exercise. | 0 | 1 | 2 | 3 | 4 | 5 |
| 2) | I exercise to improve my appearance. | 0 | 1 | 2 | 3 | 4 | 5 |
| 3) | I like my days to be organised and structured of which exercise is just one part. | 0 | 1 | 2 | 3 | 4 | 5 |
| 4) | I feel less anxious after I exercise. | 0 | 1 | 2 | 3 | 4 | 5 |
| 5) | I find exercise a chore. | 0 | 1 | 2 | 3 | 4 | 5 |
| 6) | If I feel I have eaten too much, I will do more exercise. | 0 | 1 | 2 | 3 | 4 | 5 |
| 7) | My weekly pattern of exercise is repetitive. | 0 | 1 | 2 | 3 | 4 | 5 |
| 8) | I do not exercise to be slim. | 0 | 1 | 2 | 3 | 4 | 5 |
| 9) | If I cannot exercise I feel low or depressed. | 0 | 1 | 2 | 3 | 4 | 5 |
| 10) | I feel extremely guilty if I miss an exercise session. | 0 | 1 | 2 | 3 | 4 | 5 |
| 11) | I usually continue to exercise despite injury or illness, unless I am very ill or too injured. | 0 | 1 | 2 | 3 | 4 | 5 |
| 12) | I enjoy exercising. | 0 | 1 | 2 | 3 | 4 | 5 |
| 13) | I exercise to burn calories and lose weight. | 0 | 1 | 2 | 3 | 4 | 5 |
| 14) | I feel less stressed and/or tense after I exercise. | 0 | 1 | 2 | 3 | 4 | 5 |
| 15) | If I miss an exercise session, I will try and make up for it when I next exercise. | 0 | 1 | 2 | 3 | 4 | 5 |
| 16) | If I cannot exercise I feel agitated and/or irritable. | 0 | 1 | 2 | 3 | 4 | 5 |
| 17) | Exercise improves my mood. | 0 | 1 | 2 | 3 | 4 | 5 |
| 18) | If I cannot exercise, I worry that I will gain weight. | 0 | 1 | 2 | 3 | 4 | 5 |
| 19) | I follow a set routine for my exercise sessions e.g. walk or run the same route, particular exercises, same amount of time, and so on. | 0 | 1 | 2 | 3 | 4 | 5 |
| 20) | If I cannot exercise I feel angry and/or frustrated. | 0 | 1 | 2 | 3 | 4 | 5 |
| 21) | I do not enjoy exercising. | 0 | 1 | 2 | 3 | 4 | 5 |
| 22) | I feel like I've let myself down if I miss an exercise session. | 0 | 1 | 2 | 3 | 4 | 5 |
| 23) | If I cannot exercise I feel anxious. | 0 | 1 | 2 | 3 | 4 | 5 |
| 24) | I feel less depressed or low after I exercise. | 0 | 1 | 2 | 3 | 4 | 5 |

### OCI-R

The following statements refer to experiences that many people have in their everyday lives. Circle the number that best describes **HOW MUCH** that experience has **DISTRESSED or BOTHERED you during the PAST MONTH.** The numbers refer to the following verbal labels:

| 0<br>Not at all | 1<br>A little | 2<br>Moderately | 3<br>A lot | 4<br>Extremely |
|---|---|---|---|---|

| | | | | | | |
|---|---|---|---|---|---|---|
| 1. | I have saved up so many things that they get in the way. | 0 | 1 | 2 | 3 | 4 |
| 2. | I check things more often than necessary. | 0 | 1 | 2 | 3 | 4 |
| 3. | I get upset if objects are not arranged properly. | 0 | 1 | 2 | 3 | 4 |
| 4. | I feel compelled to count while I am doing things. | 0 | 1 | 2 | 3 | 4 |
| 5. | I find it difficult to touch an object when I know it has been touched by strangers or certain people. | 0 | 1 | 2 | 3 | 4 |
| 6. | I find it difficult to control my own thoughts. | 0 | 1 | 2 | 3 | 4 |
| 7. | I collect things I don’t need. | 0 | 1 | 2 | 3 | 4 |
| 8. | I repeatedly check doors, windows, drawers, etc. | 0 | 1 | 2 | 3 | 4 |
| 9. | I get upset if others change the way I have arranged things. | 0 | 1 | 2 | 3 | 4 |
| 10. | I feel I have to repeat certain numbers. | 0 | 1 | 2 | 3 | 4 |
| 11. | I sometimes have to wash or clean myself simply because I feel contaminated. | 0 | 1 | 2 | 3 | 4 |
| 12. | I am upset by unpleasant thoughts that come into my mind against my will. | 0 | 1 | 2 | 3 | 4 |
| 13. | I avoid throwing things away because I am afraid I might need them later. | 0 | 1 | 2 | 3 | 4 |
| 14. | I repeatedly check gas and water taps and light switches after turning them off. | 0 | 1 | 2 | 3 | 4 |
| 15. | I need things to be arranged in a particular way. | 0 | 1 | 2 | 3 | 4 |
| 16. | I feel that there are good and bad numbers. | 0 | 1 | 2 | 3 | 4 |
| 17. | I wash my hands more often and longer than necessary. | 0 | 1 | 2 | 3 | 4 |
| 18. | I frequently get nasty thoughts and have difficulty in getting rid of them. | 0 | 1 | 2 | 3 | 4 |

#### Obsessive-Compulsive Inventory – Revised (OCI-R)

Administration & Scoring

The OCI-R is a short version of the OCD (Foa, Kozak, Salkovskis, Coles, & Amir, 1998) and is a self-report scale for assessing symptoms of Obsessive-Compulsive Disorder (OCD). It consists of 18 questions that a person endorses on a 5-point Likert scale.

Scores are generated by adding the item scores. The possible range of scores is 0-72. Mean score for persons with OCD is 28.0 (*SD* = 13.53). Recommended cutoff score is 21, with scores at or above this level indicating the likely presence of OCD.

### Adult ADHD Self-Report Scale (ASRS-v1.1) Symptom Checklist Instructions

*The questions on the back page are designed to stimulate dialogue between you and your patients and to help confirm if they may be suffering from the symptoms of attention-deficit/hyperactivity disorder (ADHD).*

Description: The Symptom Checklist is an instrument consisting of the eighteen DSM-IV-TR criteria. Six of the eighteen questions were found to be the most predictive of symptoms consistent with ADHD. These six questions are the basis for the ASRS v1.1 Screener and are also Part A of the Symptom Checklist. Part B of the Symptom Checklist contains the remaining twelve questions.

**Instructions:**

**Symptoms**

1. Ask the patient to complete both Part A and Part B of the Symptom Checklist by marking an X in the box that most closely represents the frequency of occurrence of each of the symptoms.

2. Score Part A. If four or more marks appear in the darkly shaded boxes within Part A then the patient has symptoms highly consistent with ADHD in adults and further investigation is warranted.

3. The frequency scores on Part B provide additional cues and can serve as further probes into the patient's symptoms. Pay particular attention to marks appearing in the dark shaded boxes. The frequency-based response is more sensitive with certain questions. No total score or diagnostic likelihood is utilized for the twelve questions. It has been found that the six questions in Part A are the most predictive of the disorder and are best for use as a screening instrument.

**Impairments**

1. Review the entire Symptom Checklist with your patients and evaluate the level of impairment associated with the symptom.

2. Consider work/school, social and family settings.

3. Symptom frequency is often associated with symptom severity, therefore the Symptom Checklist may also aid in the assessment of impairments. If your patients have frequent symptoms, you may want to ask them to describe how these problems have affected the ability to work, take care of things at home, or get along with other people such as their spouse/significant other.

**History**

1. Assess the presence of these symptoms or similar symptoms in childhood. Adults who have ADHD need not have been formally diagnosed in childhood. In evaluating a patient's history, look for evidence of early-appearing and long-standing problems with attention or self-control. Some significant symptoms should have been present in childhood, but full symptomology is not necessary.

### Adult ADHD Self-Report Scale (ASRS-v1.1) Symptom Checklist

| Patient Name | | Today's Date |
|---|---|---|

| Please answer the questions below, rating yourself on each of the criteria shown using the scale on the right side of the page. As you answer each question, place an X in the box that best describes how you have felt and conducted yourself over the past 6 months. Please give this completed checklist to your healthcare professional to discuss during today's appointment. | Never | Rarely | Sometimes | Often | Very Often |
|---|---|---|---|---|---|
| 1. How often do you have trouble wrapping up the final details of a project, once the challenging parts have been done? | | | | | |
| 2. How often do you have difficulty getting things in order when you have to do a task that requires organization? | | | | | |
| 3. How often do you have problems remembering appointments or obligations? | | | | | |
| 4. When you have a task that requires a lot of thought, how often do you avoid or delay getting started? | | | | | |
| 5. How often do you fidget or squirm with your hands or feet when you have to sit down for a long time? | | | | | |
| 6. How often do you feel overly active and compelled to do things, like you were driven by a motor? | | | | | |
| | | | | | Part A |
| 7. How often do you make careless mistakes when you have to work on a boring or difficult project? | | | | | |
| 8. How often do you have difficulty keeping your attention when you are doing boring or repetitive work? | | | | | |
| 9. How often do you have difficulty concentrating on what people say to you, even when they are speaking to you directly? | | | | | |
| 10. How often do you misplace or have difficulty finding things at home or at work? | | | | | |
| 11. How often are you distracted by activity or noise around you? | | | | | |
| 12. How often do you leave your seat in meetings or other situations in which you are expected to remain seated? | | | | | |
| 13. How often do you feel restless or fidgety? | | | | | |
| 14. How often do you have difficulty unwinding and relaxing when you have time to yourself? | | | | | |
| 15. How often do you find yourself talking too much when you are in social situations? | | | | | |
| 16. When you're in a conversation, how often do you find yourself finishing the sentences of the people you are talking to, before they can finish them themselves? | | | | | |
| 17. How often do you have difficulty waiting your turn in situations when turn taking is required? | | | | | |
| 18. How often do you interrupt others when they are busy? | | | | | |
| | | | | | Part B |

### The Value of Screening for Adults With ADHD

Research suggests that the symptoms of ADHD can persist into adulthood, having a significant impact on the relationships, careers, and even the personal safety of your patients who may suffer from it.[1-4] Because this disorder is often misunderstood, many people who have it do not receive appropriate treatment and, as a result, may never reach their full potential. Part of the problem is that it can be difficult to diagnose, particularly in adults.

The Adult ADHD Self-Report Scale (ASRS-v1.1) Symptom Checklist was developed in conjunction with the World Health Organization (WHO), and the Workgroup on Adult ADHD that included the following team of psychiatrists and researchers:

- **Lenard Adler, MD**
  Associate Professor of Psychiatry and Neurology
  New York University Medical School

- **Ronald C. Kessler, PhD**
  Professor, Department of Health Care Policy
  Harvard Medical School

- **Thomas Spencer, MD**
  Associate Professor of Psychiatry
  Harvard Medical School

As a healthcare professional, you can use the ASRS v1.1 as a tool to help screen for ADHD in adult patients. Insights gained through this screening may suggest the need for a more in-depth clinician interview. The questions in the ASRS v1.1 are consistent with DSM-IV criteria and address the manifestations of ADHD symptoms in adults. Content of the questionnaire also reflects the importance that DSM-IV places on symptoms, impairments, and history for a correct diagnosis.[4]

The checklist takes about 5 minutes to complete and can provide information that is critical to supplement the diagnostic process.

#### Difficulties in Emotion Regulation Scale – 16 Item Version (DERS-16)

| 1 | 2 | 3 | 4 | 5 |
|---|---|---|---|---|
| Almost never | Sometimes | About half the time | Most of the time | Almost always |
| 0–10 % | 11–35 % | 36–65 % | 66–90 % | 91–100 % |
| Please indicate how often the following statements apply to you by writing the appropriate number from the scale above (1–5) on the line beside each item. | | | | |
| 1. I have difficulty making sense out of my feelings. [CLARITY*] | | | | |
| 2. I am confused about how I feel. [CLARITY] | | | | |
| 3. When I am upset, I have difficulty getting work done. [GOALS] | | | | |
| 4. When I am upset, I become out of control. [IMPULSE] | | | | |
| 5. When I am upset, I believe that I will remain that way for a long time. [STRATEGIES] | | | | |
| 6. When I am upset, I believe that I'll end up feeling very depressed. [STRATEGIES] | | | | |
| 7. When I am upset, I have difficulty focusing on other things. [GOALS] | | | | |
| 8. When I am upset, I feel out of control. [IMPULSE] | | | | |
| 9. When I am upset, I feel ashamed with myself for feeling that way. [NONACCEPTANCE] | | | | |
| 10. When I am upset, I feel like I am weak. [NONACCEPTANCE] | | | | |
| 11. When I am upset, I have difficulty controlling my behaviors. [IMPULSE] | | | | |
| 12. When I am upset, I believe that there is nothing I can do to make myself feel better. [STRATEGIES] | | | | |
| 13. When I am upset, I become irritated with myself for feeling that way. [NONACCEPTANCE] | | | | |
| 14. When I am upset, I start to feel very bad about myself. [STRATEGIES] | | | | |
| 15. When I am upset, I have difficulty thinking about anything else. [GOALS] | | | | |
| 16. When I am upset, my emotions feel overwhelming. [STRATEGIES] | | | | |

*Factor as described in the original study of the 36-item DERS (Gratz & Roemer, 2004). This information was not presented to participants.

#### Life events

This questionnaire contains a number of questions about events you may have experienced during your life, including some that may have been traumatic or that may be upsetting to think about.

If you find any of these questions upsetting, please feel free to skip them.

**1) Do you have a romantic partner now?**

☐**Yes *(Skip to 3a)*** ☐**No *(Continue to 2)***

**2) Have you had a romantic partner in the last 12 months?**

☐**Yes *(Continue to 3a)*** ☐**No *(Skip to 3b)***

☐**Don't know *(Skip to 3b)***

**3) Have you had any serious problems getting along with any of the following individuals <u>during the past 12 months</u>?**

| | Yes | No |
|---|---|---|
| a) Your partner | ☐ | ☐ |
| b) Family member (other than partner) | ☐ | ☐ |
| c) A close friend | ☐ | ☐ |
| d) A neighbour | ☐ | ☐ |
| e) Someone living with you (e.g. child, flatmate or elderly parent) | ☐ | ☐ |
| f) A workmate/co-worker | ☐ | ☐ |

**Question 4 will only display to participants who selected "yes" in Question 1**

**4) Please judge your partner's attitudes and behaviour towards you in recent times.**

| | Very true | Moderately true | Somewhat true | Not true at all |
|---|---|---|---|---|
| a) Is very loving to me | ☐ | ☐ | ☐ | ☐ |
| b) Is a good companion | ☐ | ☐ | ☐ | ☐ |
| c) Is affectionate to me | ☐ | ☐ | ☐ | ☐ |
| d) Is very considerate of me | ☐ | ☐ | ☐ | ☐ |
| e) Is fun to be with | ☐ | ☐ | ☐ | ☐ |
| f) Shows his/her appreciation of me | ☐ | ☐ | ☐ | ☐ |
| g) Understands my problems and worries | ☐ | ☐ | ☐ | ☐ |
| h) Confides closely in me | ☐ | ☐ | ☐ | ☐ |
| i) Makes me feel needed | ☐ | ☐ | ☐ | ☐ |
| j) Is physically gentle and considerate | ☐ | ☐ | ☐ | ☐ |

| | No | Yes, with one or two people | Yes, with more than two people |
|---|---|---|---|
| 5) If you get angry or upset do you have people you can tell just how you feel? | ☐ | ☐ | ☐ |
| 6) Recently have you had any fights or arguments with people close to you? | ☐ | ☐ | ☐ |

| | No | Yes | Yes, sort of |
|---|---|---|---|
| 7) Are you a member of any social club or sporting group? | ☐ | ☐ | ☐ |
| 8) Do you have someone you can trust with your private thoughts and feelings? | ☐ | ☐ | ☐ |
| 9) If you're having a tough time, do you have someone you can really depend on? | ☐ | ☐ | ☐ |
| 10) Is there anyone who really knows you very well (e.g. understands how you think and feel)? | ☐ | ☐ | ☐ |
| 11) Is there anyone you feel close to that understands your concerns / difficulties? | ☐ | ☐ | ☐ |
| 12) Is there anyone you feel you can turn to, if in trouble or a crisis? | ☐ | ☐ | ☐ |
| 13) When you feel happy do you have someone you can share this with? | ☐ | ☐ | ☐ |

| | Hardly ever | Some of the time | Most of the time |
|---|---|---|---|
| 14) Does it seem that your family and friends (people who are important to you) understand you? | ☐ | ☐ | ☐ |
| 15) Do you feel useful to your family and friends (people important to you)? | ☐ | ☐ | ☐ |
| 16) Do you know what is going on with your family and friends? | ☐ | ☐ | ☐ |
| 17) When you are talking with your family and friends, do you feel you are being listened to? | ☐ | ☐ | ☐ |
| 18) Do you feel you have a definite role or place in your family and among your friends? | ☐ | ☐ | ☐ |
| 19) Can you talk about your deepest problems with at least some of your family and friends? | ☐ | ☐ | ☐ |

**20) How often do friends and or family ...**

| | Never | Rarely | Sometimes | Often |
|---|---|---|---|---|
| a) Create tensions or arguments with you? | ☐ | ☐ | ☐ | ☐ |
| b) Criticise you? | ☐ | ☐ | ☐ | ☐ |
| c) Express interest in how you are doing? | ☐ | ☐ | ☐ | ☐ |
| d) Make too many demands on you? | ☐ | ☐ | ☐ | ☐ |
| e) Make you feel cared for? | ☐ | ☐ | ☐ | ☐ |

**21) Other than members of your family, how many people do you feel you can depend on or feel very close to?**

☐ **None** ☐ **1-2 people**

☐ **More than 2 people**

**22) And, thinking specifically about your family and friends, about how many times in the past week *(excluding time spent at school or work):***

| | Number of times |
|---|---|
| a) Did you spend time with someone who doesn't live with you (e.g. went to see them or they came to visit you, or you went out together)? | |
| b) Did you talk to someone (friends, relatives or others) on the telephone? | |
| c) Did you go to meetings of clubs, religious meetings, or other groups of which you're a member? | |
| d) Did you use the internet to spend time with someone, talk with someone, or attend club / group meetings? | |

**23) During the past 12 months have you had any of these events occur?**

| | Yes | No |
|---|---|---|
| **a)** Divorce | ☐ | ☐ |
| **b)** Marital separation | ☐ | ☐ |
| **c)** Broken engagement or steady relationship | ☐ | ☐ |
| **d)** Separation from other loved one or close friend | ☐ | ☐ |
| **e)** Serious illness or injury | ☐ | ☐ |
| **f)** Serious accident (not involving personal injury) | ☐ | ☐ |
| **g)** Burgled or robbed | ☐ | ☐ |
| **h)** Laid off or sacked from job | ☐ | ☐ |
| **i)** Other serious difficulties at work | ☐ | ☐ |
| **j)** Major financial problems | ☐ | ☐ |
| **k)** Legal troubles or involvement with police | ☐ | ☐ |
| **l)** Living in unpleasant surroundings | ☐ | ☐ |

The next series of questions will ask you about events you may have experienced during your life, including some that may have been traumatic or that may be upsetting to think about.

*If you find any of these questions upsetting, please feel free to skip them.*

**24) Listed below are a number of difficult or stressful things that sometimes happen to people. For each event mark one or more of the boxes to the right to indicate that:**

(a) It **happened to you** personally;
(b) You **witnessed it** happen to someone else;
(c) You **learned about it** happening to a close family member or close friend;
(d) You were exposed to it as **part of your job** (for example, paramedic, police, military or other first responder);
(e) You're **not sure** if it fits; or
(f) It **doesn't apply** to you.

Be sure to consider your **entire life** (growing up as well as adulthood) as you go through the list of events.

| | Happened to me | Witnessed it | Learned about it | Part of my job | Not sure | N/A |
|---|---|---|---|---|---|---|
| **a)** Natural disaster (e.g. flood, cyclone, tornado, earthquake) | ☐ | ☐ | ☐ | ☐ | ☐ | ☐ |
| **b)** Fire or explosion | ☐ | ☐ | ☐ | ☐ | ☐ | ☐ |
| **c)** Transportation accident (e.g. car accident, boat accident, train wreck, plane crash) | ☐ | ☐ | ☐ | ☐ | ☐ | ☐ |
| **d)** Serious accident at work, home or during recreational activity | ☐ | ☐ | ☐ | ☐ | ☐ | ☐ |
| **e)** Exposure to toxic substances (e.g. dangerous chemicals, radiation) | ☐ | ☐ | ☐ | ☐ | ☐ | ☐ |
| **f)** Physical assault (e.g. being attacked, hit, slapped, kicked, beaten up) | ☐ | ☐ | ☐ | ☐ | ☐ | ☐ |
| **g)** Assault with a weapon (e.g. being shot, stabbed, threatened with a knife, gun, bomb) | ☐ | ☐ | ☐ | ☐ | ☐ | ☐ |
| **h)** Sexual assault (rape, attempted rape, made to perform any type of sexual act through force or threat of harm) | ☐ | ☐ | ☐ | ☐ | ☐ | ☐ |
| **i)** Other unwanted or uncomfortable sexual experience | ☐ | ☐ | ☐ | ☐ | ☐ | ☐ |
| **k)** Combat or exposure to a war-zone (in the military or as a civilian) | ☐ | ☐ | ☐ | ☐ | ☐ | ☐ |
| **l)** Captivity (e.g. being kidnapped, abducted, held hostage, prisoner of war) | ☐ | ☐ | ☐ | ☐ | ☐ | ☐ |
| **m)** Life-threatening illness or injury | ☐ | ☐ | ☐ | ☐ | ☐ | ☐ |
| **n)** Severe human suffering | ☐ | ☐ | ☐ | ☐ | ☐ | ☐ |
| **o)** Sudden violent death (e.g. homicide, suicide) | ☐ | ☐ | ☐ | ☐ | ☐ | ☐ |
| **p)** Sudden accidental death | ☐ | ☐ | ☐ | ☐ | ☐ | ☐ |
| **q)** Serious injury, harm or death you caused to someone else | ☐ | ☐ | ☐ | ☐ | ☐ | ☐ |
| **r)** Any other very stressful event or experience | ☐ | ☐ | ☐ | ☐ | ☐ | ☐ |

**Questions 25-28 will only display to participants who selected "happened to me," "witnessed it" or "part of my job" for at least one of the corresponding activities in Question 24**

| | Yes | No |
|---|---|---|
| 25) Sometimes images or strong memories of traumatic events keep coming back in flashbacks, thoughts that you can't get rid of, or repeated nightmares. Has that ever happened to you? | ☐ | ☐ |
| 26) Did you make a special effort to avoid thinking or talking about what happened or deliberately stayed away from things or people that reminded you of the terrible experience? | ☐ | ☐ |
| 27) Did you make a special effort to avoid thinking or talking about what happened or deliberately stayed away from things or people that reminded you of the terrible experience? | ☐ | ☐ |
| 28) After this experience did you have trouble sleeping, have difficulty concentrating, have outbursts of anger, feel overly watchful or on guard, or were you unusually irritable, very jumpy or easily startled? | ☐ | ☐ |

**29) People may experience stressful situations in childhood which may affect their future health and well-being. Please indicate if you experienced any of these situations during your childhood.**

| | Yes | No | Don't know |
|---|---|---|---|
| a) Emotional abuse (e.g. often being told you were no good, yelled at in a scary way, threatened, ignored, or stopped from making friends) | ☐ | ☐ | ☐ |
| b) Emotional neglect (e.g. often not being shown affection, or not being given encouragement or support) | ☐ | ☐ | ☐ |
| c) Physical neglect (e.g. often not being given enough to eat or drink, appropriate clothing, shelter, medical care, education, supervision or a safe home environment) | ☐ | ☐ | ☐ |

**Question 30 will only display to participants who selected "happened to me," "witnessed it," "part of my job" or "learned about it" for at least one of the corresponding activities in Question 24 OR selected "yes" at least once in question 29**

**30) How old were you the first and last time these things happened?**

***This includes if it happened to you personally, you witnessed it happening to someone else, you learned about it happening to a close family member or close friend, or you were exposed to it as part of your job.***

***If something happened only once, please enter the same age for the first and last time.***

| | First time *(age in years)* | Last time *(age in years)* |
|---|---|---|
| **a)** Natural disaster | ☐☐ | ☐☐ |
| **b)** Fire or explosion | ☐☐ | ☐☐ |
| **c)** Transportation accident | ☐☐ | ☐☐ |
| **d)** Serious accident | ☐☐ | ☐☐ |
| **e)** Exposure to toxic substance | ☐☐ | ☐☐ |
| **f)** Physical assault | ☐☐ | ☐☐ |
| **g)** Assault with a weapon | ☐☐ | ☐☐ |
| **h)** Sexual assault | ☐☐ | ☐☐ |
| **i)** Other unwanted or uncomfortable sexual experience | ☐☐ | ☐☐ |
| **j)** Combat or exposure to a war-zone | ☐☐ | ☐☐ |
| **k)** Captivity | ☐☐ | ☐☐ |
| **l)** Life-threatening illness or injury | ☐☐ | ☐☐ |
| **m)**Severe human suffering | ☐☐ | ☐☐ |
| **n)** Sudden violent death | ☐☐ | ☐☐ |
| **o)** Sudden accidental death | ☐☐ | ☐☐ |
| Serious injury, harm or death you caused to someone else | ☐☐ | ☐☐ |
| Other stressful event or experience | ☐☐ | ☐☐ |
| Emotional abuse | ☐☐ | ☐☐ |
| Emotional neglect | ☐☐ | ☐☐ |
| Physical neglect | ☐☐ | ☐☐ |

31) How old were you when you first had sexual intercourse with your consent?

☐☐ Age in years OR

☐ I have not had sexual intercourse with consent

National Institute for Health Research

# AQ-10

#### Autism Spectrum Quotient (AQ)

*A quick referral guide for adults with suspected autism who do not have a learning disability.*

| | ***Please tick one option per question only:*** | **Definitely Agree** | **Slightly Agree** | **Slightly Disagree** | **Definitely Disagree** |
|---|---|---|---|---|---|
| 1 | I often notice small sounds when others do not | | | | |
| 2 | I usually concentrate more on the whole picture, rather than the small details | | | | |
| 3 | I find it easy to do more than one thing at once | | | | |
| 4 | If there is an interruption, I can switch back to what I was doing very quickly | | | | |
| 5 | I find it easy to 'read between the lines' when someone is talking to me | | | | |
| 6 | I know how to tell if someone listening to me is getting bored | | | | |
| 7 | When I'm reading a story I find it difficult to work out the characters' intentions | | | | |
| 8 | I like to collect information about categories of things (e.g. types of car, types of bird, types of train, types of plant etc) | | | | |
| 9 | I find it easy to work out what someone is thinking or feeling just by looking at their face | | | | |
| 10 | I find it difficult to work out people's intentions | | | | |

**SCORING:** Only 1 point can be scored for each question. *Score 1 point for Definitely or Slightly Agree on each of items 1, 7, 8, and 10. Score 1 point for Definitely or Slightly Disagree on each of items 2, 3, 4, 5, 6, and 9.* If the individual scores **more than 6 out of 10**, consider referring them for a specialist diagnostic assessment.

This test is recommended in 'Autism: recognition, referral, diagnosis and management of adults on the autism spectrum' (NICE clinical guideline CG142). www.nice.org.uk/CG142

**Key reference:** Allison C, Auyeung B, and Baron-Cohen S, (2012) *Journal of the American Academy of Child and Adolescent Psychiatry* 51(2):202-12.

UNIVERSITY OF CAMBRIDGE

autism research centre arc

© SBC/CA/BA/ARC/Cambridge University 1/5/12

<u>Lifetime Depression and Anxiety Questionnaire</u>

CIDID.SYM.1.0 Have you ever had a time in your life when you have felt sad, blue, or depressed for **two weeks or more** in a row?

- ◯ Yes (1)
- ◯ No (0)
- ◯ Prefer not to answer (-99)

CIDID.SYM.2.0 Have you ever had a time in your life lasting **two weeks or more** when you lost interest in most things like hobbies, work, or activities that usually give you pleasure?

- ◯ Yes (1)
- ◯ No (0)
- ◯ Prefer not to answer (-99)

*Display This Question:*

*If Anhedonia2Weeks = Yes*

*Or LowMood2Weeks = Yes*

HEAD Please think of the **two-week period** in your life when your feelings of depression or loss of interest were worst.

---

*Display This Question:*

*If Header Is Displayed*

CIDID.SYM.3.0 **How much** of the day did these feelings **usually** last?

- All day long (4)
- Most of the day (3)
- About half of the day (2)
- Less than half of the day (1)
- Don't know (-88)
- Prefer not to answer (-99)

*Display This Question:*

*If Header Is Displayed*

HEAD Please think of the **two-week period** in your life when your feelings of depression or loss of interest were worst.

---

*Display This Question:*

*If Header Is Displayed*

CIDID.SYM.4.0 Did you feel this way:

- Every day (3)
- Almost every day (2)
- Less often (1)
- Don't know (-88)
- Prefer not to answer (-99)

---

*Display This Question:*

*If Header Is Displayed*

CIDID.SYM.5.0 Did you feel more **tired** out or **low on energy** than is usual for you?

- ○ Yes (1)
- ○ No (0)
- ○ Don't know (-88)
- ○ Prefer not to answer (-99)

*Display This Question:*

HEAD Please think of the **two-week period** in your life when your feelings of depression or loss of interest were worst.

---

*Display This Question:*

*If Header Is Displayed*

CIDID.ATY.1.0 Did your **weight** change?

(do **not** include weight change as a side-effect of medication you were taking)

- ○ Gained weight (1)
- ○ Lost weight (2)
- ○ Both gained and lost some weight during the episode (3)
- ○ Stayed about the same or was on a diet (0)
- ○ Don't know (-88)
- ○ Prefer not to answer (-99)

*Display This Question:*

*If Did your weight change? = Gained weight*

*Or Did your weight change? = Lost weight*

*Or Did your weight change? = Both gained and lost some weight during the episode*

CIDID.ATY.2.0 Did your **weight** change by about **10lbs (4kg)** or more?

- ◯ Yes (1)
- ◯ No (0)
- ◯ Don't know (-88)
- ◯ Prefer not to answer (-99)

*Display This Question:*

*If Header Is Displayed*

HEAD Please think of the **two-week period** in your life when your feelings of depression or loss of interest were worst.

---

*Display This Question:*

*If Header Is Displayed*

CIDID.SYM.6.0 Did your **sleep** change?

*(do **not** include sleep change as a side-effect of medication you were taking)*

- ◯ Yes (1)
- ◯ No (0)
- ◯ Don't know (-88)
- ◯ Prefer not to answer (-99)

*Display This Question:*

*If Did your sleep change? = Yes*

CIDID.SYM Was that:

| | Yes (1) | No (0) |
|---|---|---|
| Trouble falling asleep *(sleeping too little)* (CIDID.SYM.7.0) | ○ | ○ |
| Waking **too early** *(sleeping too little)* (CIDID.SYM.8.0) | ○ | ○ |
| Sleeping **too much** (CIDID.SYM.9.0) | ○ | ○ |
| Both sleeping **too much** and **too little** during the **same** depression episode (CIDID.SYM.10.0) | ○ | ○ |

*Display This Question:*

*If SleepChangeMoreInfo = Trouble falling asleep <i>(sleeping too little)</i> [ Yes ]*

*Or SleepChangeMoreInfo = Waking <b>too early</b> <i>(sleeping too little)</i> [ Yes ]*

*Or SleepChangeMoreInfo = Sleeping <b>too much</b> [ Yes ]*

*Or SleepChangeMoreInfo = Both sleeping <b>too much</b> and <b>too little</b> during the <b>same</b> depression episode [ Yes ]*

CIDID.ATY.3.0 How many **hours per day** did you **sleep** on average **during the depression** episode, including nighttime sleep and daytime naps?

________________________________________________________________

*Display This Question:*

*If SleepChangeMoreInfo = Trouble falling asleep <i>(sleeping too little)</i> [ Yes ]*

*Or SleepChangeMoreInfo = Waking <b>too early</b> <i>(sleeping too little)</i> [ Yes ]*

*Or SleepChangeMoreInfo = Sleeping <b>too much</b> [ Yes ]*

*Or SleepChangeMoreInfo = Both sleeping <b>too much</b> and <b>too little</b> during the <b>same</b> depression episode [ Yes ]*

CIDID.ATY.4.0 How many **hours per day** did you used to **sleep** on average when you were **not depressed**?

________________________________________________________________

*Display This Question:*

*If Header Is Displayed*

HEAD Please think of the **two-week period** in your life when your feelings of depression or loss of interest were worst.

---

*Display This Question:*

*If Header Is Displayed*

CIDID.SYM.11.0 Did you experience a **change** in your **appetite**?

- ◯ No changes in appetite (0)
- ◯ Increased appetite (1)
- ◯ Decreased appetite (2)
- ◯ Experienced both increased an decreased appetite during the same depression episode (3)
- ◯ Don’t know (-88)
- ◯ Prefer not to answer (-99)

---

*Display This Question:*

*If Header Is Displayed*

CIDID.ATY.5.0 Did your **mood brighten** in response to **positive events?**

- ◯ Yes (1)
- ◯ No (0)
- ◯ Don't know (-88)
- ◯ Prefer not to answer (-99)

*Display This Question:*

*If Header Is Displayed*

CIDID.ATY.6.0 Did you experience **heavy feelings** in your arms or legs? *(Did your arms or legs feel **"heavy"**?)*

◯ Yes (1)

◯ No (0)

◯ Don't know (-88)

◯ Prefer not to answer (-99)

---

*Display This Question:*

*If Did you experience heavy feelings in your arms or legs? = Yes*

CIDID.ATY.7.0 For how many **hours per day** was the **heaviness** feeling present?

________________________________________________________________

*Display This Question:*

*If Header Is Displayed*

HEAD Please think of the **two-week period** in your life when your feelings of depression or loss of interest were worst.

---

*Display This Question:*

*If Header Is Displayed*

CIDID.ATY.8.0 Were you **overly sensitive** to interpersonal **rejection**?

◯ No (0)

◯ Yes, and this **significantly** impaired your social or work relationships. (1)

◯ Yes, but this did **not** significantly impair your social or work relationships (2)

◯ Don't know (-88)

◯ Prefer not to answer (-99)

*Display This Question:*

*If Header Is Displayed*

HEAD Please think of the **two-week period** in your life when your feelings of depression or loss of interest were worst.

---

*Display This Question:*

*If Header Is Displayed*

CIDID.SYM.12.0 Was your mood **worse:**

- In the **morning** (1)
- In the **afternoon** (2)
- At **night** (3)
- My mood did **not** vary (0)
- Don't know (-88)
- Prefer not to answer (-99)

---

*Display This Question:*

*If Header Is Displayed*

CIDID.SYM.13.0 Did you have a lot more **trouble concentrating than usual?**

- Yes (1)
- No (0)
- Don't know (-88)
- Prefer not to answer (-99)

*Display This Question:*

*If Header Is Displayed*

HEAD Please think of the **two-week period** in your life when your feelings of depression or loss of interest were worst.

*Display This Question:*

*If Header Is Displayed*

CIDID.SYM.14.0 People sometimes **feel down** on themselves, no good, worthless. Did you feel this way?

- Yes (1)
- No (0)
- Don't know (-88)
- Prefer not to answer (-99)

*Display This Question:*

*If Header Is Displayed*

CIDID.SYM.15.0 Did you **think** a lot about **death** - either your own, someone else's, or death in general?

- Yes (1)
- No (0)
- Don't know (-88)
- Prefer not to answer (-99)

*Display This Question:*

*If Header Is Displayed*

HEAD Please think of the **two-week period** in your life when your feelings of depression or loss of interest were worst.

*Display This Question:*

*If  Header Is Displayed*

CIDID.SYM.16.0 About **how long** altogether did you feel this way?

- Less than a **month** (1)
- Between **one** and **three months** (2)
- **Over** three months, but **less** than six months (3)
- **Over** six months, but **less** than 12 months (4)
- One to two **years** (5)
- **Over** two years (6)
- Don't know (-88)
- Prefer not to answer (-99)

---

*Display This Question:*

*If About how long altogether did you feel this way? , <B>Less</b> than a <b>month</b> Is Displayed*

CIDID.SYM.24.0 Was **this** your **longest** episode of depression or low mood?

- Yes (1)
- No (2)
- Don't know (-88)
- Prefer not to answer (-99)

*isplay This Question:*

*If Was this your longest episode of depression or low mood? = No*

CIDID.SYM.25.0 What is the **longest** period of time that you have experienced **depression** or **low mood**?

- Less than 6 months (1)
- Over 6 months but less than 12 months (2)
- Over 1 year but less than 5 years (3)
- More than 5 years (4)
- All of my life / as long as I can remember (5)

*Display This Question:*

*If Header Is Displayed*

HEAD Please think of the **two-week period** in your life when your feelings of depression or loss of interest were worst.

---

*Display This Question:*

*If Header Is Displayed*

CIDID.SYM.17.0 Think about your **roles** at the time of this episode, including study/employment, childcare and housework, leisure pursuits. How much did these problems **interfere** with your life or activities?

- A lot (3)
- Some (2)
- A little (1)
- Not at all (0)
- Prefer not to answer (-99)

*Display This Question:*

*If Header Is Displayed*

CIDID.SYM.18.0 How many periods of depression or low mood have you had in your life **lasting two or more weeks?**

- ◯ One (1)
- ◯ Two - three (2)
- ◯ Several (3)
- ◯ All of my life / as long as I can remember (4)
- ◯ Prefer not to answer (-99)

---

*Display This Question:*

*If How many periods of depression or low mood have you had? = Two - three*

*Or How many periods of depression or low mood have you had? = Several*

*Or How many periods of depression or low mood have you had? = All of my life / as long as I can remember*

CIDID.SYM.19.0 Please estimate the number of times you have had periods of depression or low mood in your life lasting two or more weeks:

▼ 1 (1) ... 13+ (13)

*Display This Question:*

*If LowMood2Weeks = Yes*

*Or Anhedonia2Weeks = Yes*

CIDID.SYM.20.0 About how old were you the **first** time you had a **period of two weeks** like this? (Whether or not you received any help for it.)

*Please put your age in years. An approximate age is fine.*

________________________________________________________________

*Display This Question:*

*If LowMood2Weeks = Yes*

*Or Anhedonia2Weeks = Yes*

*And If*

*NumberOfDepressiveEpisodes != 1*

*Or How many periods of depression or low mood have you had? != One*

CIDID.SYM.21.0 About how old were you the **last** time you had a **period of two weeks** like this? (Whether or not you received any help for it.)

*Please put your age in years. An approximate age is fine.*

________________________________________________________________

*Display This Question:*

*If What was your biological sex at birth?Note: This question refers to biological sex, not gender. R... = Female*

*And If*

*LowMood2Weeks = Yes*

*Or Anhedonia2Weeks = Yes*

CIDID.SYM.22.0 Did any of these episodes occur within the **first year of giving birth?** Or has it been suggested you had post-natal depression?

- ◯ Yes (1)
- ◯ No (0)
- ◯ Don't know (-88)
- ◯ Prefer not to answer (-99)
- ◯ Not applicable to me (2)

*Display This Question:*

*If Header Is Displayed*

CIDID.SYM.23.0 Did any of these episodes occur following a **significant** or **traumatic event** such as **death/serious illness** of a close relative or friend, or following **a distressing event** or **illness** that happened to you?

- ◯ Most/all (3)
- ◯ More than once (2)
- ◯ Once (1)
- ◯ Not at all (0)
- ◯ Prefer not to answer (-99)

*Display This Question:*

*If Header Is Displayed*

CIDID.TRE.1.0 Did you **ever** tell a professional about these problems? *(Medical doctor, psychologist, social worker, counsellor, nurse, clergy, or other helping professional)*

- ◯ Yes (1)
- ◯ No (0)
- ◯ Don't know (-88)
- ◯ Prefer not to answer (-99)

*Display This Question:*

*If Header Is Displayed*

CIDID.TRE.2.0 Did you **ever** try or are **currently** trying the following for these problems?

*(Please select all that apply)*

☐ Medication prescribed to you for at least two weeks (1)

☐ Unprescribed medication more than once (2)

☐ Drugs or alcohol more than once (3)

☐ Psychotherapy or other talking therapy more than once (including internet-based CBT) (5)

☐ Structured wellbeing activity (e.g. mindfulness, meditation, self-help) (6)

☐ Regular physical exercise (e.g. yoga, running, walking) (7)

☐ ⊗None of the above (0)

☐ ⊗Prefer not to answer (-99)

*Display This Question:*

*If Did you ever try the following for depression? = Psychotherapy or other talking therapy more than once (including internet-based CBT)*

CIDID.IAP.1.0 Are you **currently** enrolled in an **NHS** funded **talking therapy** or **psychotherapy** (IAPT)?

○ Yes (1)

○ No (0)

○ Don't know (-88)

*Display This Question:*

*If Did you ever try the following for depression? = Medication prescribed to you for at least two weeks*

CIDID.TRE.3.0 Did you take your medication as **advised?**

- ◯ Yes (1)
- ◯ No (0)
- ◯ Don't know (-88)
- ◯ Prefer not to answer (-99)

---

*Display This Question:*

*If Did you ever try the following for depression? = Medication prescribed to you for at least two weeks*

CIDID.TRE.4.0 Did you find the medication **helpful?**

- ◯ Yes (1)
- ◯ No (0)
- ◯ Don't know (-88)
- ◯ Prefer not to answer (-99)

*Display This Question:*

*If Did you ever try the following for depression? = Psychotherapy or other talking therapy more than once (including internet-based CBT)*

*Or Did you ever try the following for depression? = Structured wellbeing activity (e.g. mindfulness, meditation, self-help)*

CIDID.THE.1.0 You previously mentioned that you have tried psychotherapy, another talking therapy, or a structured wellbeing activity for **depression**. Please **select all** that you attended **more than once**.

- ☐ Counselling (1)
- ☐ Mindfulness (2)
- ☐ Relationship therapy (3)
- ☐ Group therapy (4)
- ☐ Guided self-help (5)
- ☐ Family therapy (6)
- ☐ Cognitive Behavioral Therapy (CBT) (7)
- ☐ Workshops (8)
- ☐ Online therapy (9)
- ☐ Other (10) ________________________________________________
- ☐ ⊗Never tried psychotherapy or other talking therapy (0)
- ☐ ⊗Don't know (-88)
- ☐ ⊗Prefer not to answer (-99)

*Display This Question:*

*If please select all therapy for depression that you attended , Counselling Is Displayed*

*And please select all therapy for depression that you attended != Never tried psychotherapy or other talking therapy*

*And please select all therapy for depression that you attended != Don't know*

*And please select all therapy for depression that you attended != Prefer not to answer*

CIDID.THE.2.0 Did you **complete** your **course** of psychotherapy or other talking therapy?

◯ Yes (1)

◯ No (2)

◯ Don't know (3)

◯ Prefer not to answer (5)

---

*Display This Question:*

*If please select all therapy for depression that you attended != Never tried psychotherapy or other talking therapy*

*And please select all therapy for depression that you attended != Don't know*

*And please select all therapy for depression that you attended != Prefer not to answer*

*And please select all therapy for depression that you attended , Counselling Is Displayed*

CIDID.THE.3.0 Did you find psychotherapy or other talking therapy **helpful**?

◯ Yes (1)

◯ No (2)

◯ Don't know (3)

◯ Prefer not to answer (4)

HEAD Over the last **2 weeks**, how often have you been bothered by any of the following problems?

Select ONE for each of the following statements:

| | Not at all (0) | Several days (1) | More than half the days (2) | Nearly every day (3) |
|---|---|---|---|---|
| Feeling nervous, anxious, or on edge (GAD.1.0) | ○ | ○ | ○ | ○ |
| Not being able to stop or control worrying (GAD.2.0) | ○ | ○ | ○ | ○ |
| Worrying too much about different things (GAD.3.0) | ○ | ○ | ○ | ○ |
| Trouble relaxing (GAD.4.0) | ○ | ○ | ○ | ○ |
| Being so restless that it is hard to sit still (GAD.5.0) | ○ | ○ | ○ | ○ |
| Becoming easily annoyed or irritable (GAD.6.0) | ○ | ○ | ○ | ○ |
| Feeling afraid as if something awful might happen (GAD.7.0) | ○ | ○ | ○ | ○ |

CIDIA.SYM.1.0 Have you ever had a period lasting **one month or longer** when most of the time you felt worried, tense, or anxious?

○ Yes (1)

○ No (0)

○ Don't know (-88)

○ Prefer not to answer (-99)

CIDIA.SYM.2.0 People differ a lot in how much they worry about things. Did you ever have a time when you worried a lot **more than most people** would in your situation?

- Yes (1)
- No (0)
- Don't know (-88)
- Prefer not to answer (-99)

*Display This Question:*

*If Have you ever had a period when most of the time you felt worried, tense = Yes*

*Or Have you ever had a period when most of the time you felt worried, tense = Don't know*

*Or Did you worry more than most people? = Yes*

*Or Did you worry more than most people? = Don't know*

CIDIA.SYM.3.0 What is the **longest period of time** that this kind of worrying has ever continued? *(If you are not sure of the exact amount of time, please give an estimate)*

- Less than 6 months (1)
- Over 6 months but less than 12 months (2)
- Over 1 year but less than 5 years (3)
- More than 5 years (4)
- All of my life / as long as I can remember (5)

*Display This Question:*

*If What is the longest period of time that this kind of worrying has ever continued?(If you are not... = Less than 6 months*

CIDIA.SYM.3.0.2 What is the **longest period of time (in months)** that this kind of worrying has ever continued for?

▼ Less than 1 month (0) ... 6 (6)

*Display This Question:*

*If What is the longest period of time (in months) that this kind of worrying has ever continued for? = Less than 1 month*

CIDIA.SYM.3.0.2 What is the longest period of time (in weeks) that this kind of worrying has ever continued for?

▼ Less than 1 week (1) ... 4 (5)

*Display This Question:*

*If What is the longest period of time (in months) that this kind of worrying has ever continued for? != Less than 1 month*

*And What is the longest period of time (in months) that this kind of worrying has ever continued for? , Less than 1 month Is Displayed*

CIDIA.SYM.20.0.5 **How many periods** of this kind of worry have you had in your life lasting **1 month or longer**?

- One (1)
- Two-three (2)
- Several (3)
- All of my life / as long as I can remember (4)
- Prefer not to answer (-99)

*Display This Question:*

*If What is the longest period of time that this kind of worrying has ever continued?(If you are not... != Less than 6 months*

*And What is the longest period of time that this kind of worrying has ever continued?(If you are not... , Less than 6 months Is Displayed*

CIDIA.SYM.20.0 **How many periods** of this kind of worry have you had in your life lasting **6 or more months**?

- One (1)
- Two-three (2)
- Several (3)
- All of my life / as long as I can remember (4)
- Prefer not to answer (-99)

*Display This Question:*

*If How many periods of this kind of worry have you had in your life lasting 6 or more months? = Two-three*

*Or How many periods of this kind of worry have you had in your life lasting 6 or more months? = Several*

*Or How many periods of this kind of worry have you had in your life lasting 6 or more months? = All of my life / as long as I can remember*

CIDIA.SYM.21.0 Please estimate the **number of times** you have had periods of this kind of worry in your life **lasting 6 or more months**:

▼ 1 (1) ... 13+ (13)

---

*Display This Question:*

*If How many periods of this kind of worry have you had in your life lasting 1 month or longer? = Two-three*

*Or How many periods of this kind of worry have you had in your life lasting 1 month or longer? = Several*

*Or How many periods of this kind of worry have you had in your life lasting 1 month or longer? = All of my life / as long as I can remember*

CIDIA.SYM.21.0.5 Please estimate the **number of times** you have had periods of this kind of worry in your life lasting **1 month or longer**:

▼ 1 (1) ... 13+ (13)

*Display This Question:*

*If What is the longest period of time that this kind of worrying has ever continued?(If you are not... != Less than 6 months*

*And If*

*Have you ever had a period when most of the time you felt worried, tense = Yes*

*Or Did you worry more than most people? = Yes*

CIDIA.SYM.22.0 About **how old were you** the **first time** you had a period of **6 months** like this? (Whether or not you received any help for it.)

*Please put your age in years, an approximate age is fine.*

________________________________________________________________

---

*Display This Question:*

*If What is the longest period of time (in months) that this kind of worrying has ever continued for? != Less than 1 month*

*And What is the longest period of time (in months) that this kind of worrying has ever continued for? , Less than 1 month Is Displayed*

*And If*

*Have you ever had a period when most of the time you felt worried, tense = Yes*

*Or Did you worry more than most people? = Yes*

CIDIA.SYM.22.0.5 About **how old were you** the **first time** you had a period of **1 month or longer** like this? (Whether or not you received any help for it.)

*Please put your age in years, an approximate age is fine.*

________________________________________________________________

*Display This Question:*

*If How many periods of this kind of worry have you had in your life lasting 6 or more months? != One*

*Or Please estimate the number of times you have had periods of this kind of worry in your life lasti... != 1*

*And If*

*What is the longest period of time that this kind of worrying has ever continued?(If you are not... != Less than 6 months*

*And If*

*Have you ever had a period when most of the time you felt worried, tense = Yes*

*Or Have you ever had a period when most of the time you felt worried, tense = Don't know*

*Or Did you worry more than most people? = Yes*

*Or Did you worry more than most people? = Don't know*

CIDIA.SYM.23.0 About **how old** were you the **last time** you had a period of **6 months** like this? (whether or not you received any help for it.)

*Please put your age in years, an approximate age is fine.*

________________________________________________________________

*Display This Question:*

*If How many periods of this kind of worry have you had in your life lasting 1 month or longer? != One*

*Or Please estimate the number of times you have had periods of this kind of worry in your life lasti... != 1*

*And If*

*What is the longest period of time (in months) that this kind of worrying has ever continued for? != Less than 1 month*

*And What is the longest period of time (in months) that this kind of worrying has ever continued for? , Less than 1 month Is Displayed*

*And If*

*Have you ever had a period when most of the time you felt worried, tense = Yes*

*Or Have you ever had a period when most of the time you felt worried, tense = Don't know*

*Or Did you worry more than most people? = Yes*

*Or Did you worry more than most people? = Don't know*

CIDIA.SYM.23.0.5 About **how old** were you the **last time** you had a period of **1 month or longer** like this? (whether or not you received any help for it.)

*Please put your age in years, an approximate age is fine.*

________________________________________________________________

*Display This Question:*

*If What is the longest period of time that this kind of worrying has ever continued?(If you are not... = Over 6 months but less than 12 months*

*Or What is the longest period of time that this kind of worrying has ever continued?(If you are not... = Over 1 year but less than 5 years*

*Or What is the longest period of time that this kind of worrying has ever continued?(If you are not... = More than 5 years*

*Or What is the longest period of time that this kind of worrying has ever continued?(If you are not... = All of my life / as long as I can remember*

*Or What is the longest period of time (in months) that this kind of worrying has ever continued for? = 1*

*Or What is the longest period of time (in months) that this kind of worrying has ever continued for? = 2*

*Or What is the longest period of time (in months) that this kind of worrying has ever continued for? = 3*

*Or What is the longest period of time (in months) that this kind of worrying has ever continued for? = 4*

*Or What is the longest period of time (in months) that this kind of worrying has ever continued for? = 5*

*Or What is the longest period of time (in months) that this kind of worrying has ever continued for? = 6*

HEAD Please think of the period in your life when you have felt **worried, tense, anxious,** or **more worried** than most people would in your situation. This could be in the past, or it could be continuing

now.
The following questions refer to this period of time.

*Display This Question:*

*If Header Is Displayed*

CIDIA.SYM.5.0 During that period, was your worry **stronger** than in other people?

- Yes (1)
- No (0)
- Don't know (-88)
- Prefer not to answer (-99)

*Display This Question:*

*If Header Is Displayed*

CIDIA.SYM.6.0 Did you worry **most days**?

- Yes (1)
- No (0)
- Don't know (-88)
- Prefer not to answer (-99)

*Display This Question:*

*If Header Is Displayed*

HEAD Please think of the period in your life when you have felt **worried, tense, anxious,** or **more worried** than most people would in your situation. This could be in the past, or it could be continuing now.

*Display This Question:*

*If Header Is Displayed*

CIDIA.SYM.7.0 Did you usually worry about **one particular thing**, such as your job security or the failing health of a loved one, or **more than one thing**?

- One thing (0)
- More than one thing (1)
- Don't know (-88)
- Prefer not to answer (-99)

---

*Display This Question:*

*If Header Is Displayed*

CIDIA.SYM.8.0 Did you find it **difficult to stop** worrying?

- Yes (1)
- No (0)
- Don't know (-88)
- Prefer not to answer (-99)

*Display This Question:*

*If Header Is Displayed*

HEAD Please think of the period in your life when you have felt **worried, tense, anxious,** or **more worried** than most people would in your situation. This could be in the past, or it could be continuing now.

---

*Display This Question:*

*If Header Is Displayed*

CIDIA.SYM.9.0 Did you ever have different worries on your mind **at the same time**?

- Yes (1)
- No (0)
- Don't know (-88)
- Prefer not to answer (-99)

---

*Display This Question:*

*If Header Is Displayed*

CIDIA.SYM.10.0 How often was your worry so strong that you **couldn't** put it out of your mind no matter how hard you tried?

- Often (3)
- Sometimes (2)
- Rarely (1)
- Never (0)
- Don't know (-88)
- Prefer not to answer (-99)

*Display This Question:*

*If Header Is Displayed*

HEAD Please think of the period in your life when you have felt **worried, tense, anxious,** or **more worried** than most people would in your situation. This could be in the past, or it could be continuing now.

---

*Display This Question:*

*If Header Is Displayed*

CIDIA.SYM.11.0 How often did you find it **difficult to control** your worry?

- Often (3)
- Sometimes (2)
- Rarely (1)
- Never (0)
- Don't know (-88)
- Prefer not to say (-99)

*Display This Question:*

*If Header Is Displayed*

HEAD When you were **worried or anxious**, were you also:

| | Yes (1) | No (0) | Don't know (-88) |
|---|---|---|---|
| Restless? (CIDIA.SYM.12.0) | ○ | ○ | ○ |
| Keyed up or on edge? (CIDIA.SYM.13.0) | ○ | ○ | ○ |
| Easily tired? (CIDIA.SYM.14.0) | ○ | ○ | ○ |
| Having difficulty keeping your mind on what you were doing? (CIDIA.SYM.15.0) | ○ | ○ | ○ |
| More irritable than usual? (CIDIA.SYM.16.0) | ○ | ○ | ○ |
| Having tense, sore, or aching muscles? (CIDIA.SYM.17.0) | ○ | ○ | ○ |
| Often having trouble falling or staying asleep? (CIDIA.SYM.18.0) | ○ | ○ | ○ |

*Display This Question:*

*If Header Is Displayed*

CIDIA.TRE.1.0 Did you **ever** tell a **professional** about these problems? *(medical doctor, psychologist, social worker, counselor, nurse, clergy, or other helping professional)*

- ○ Yes (1)
- ○ No (0)
- ○ Don't know (-88)
- ○ Prefer no to answer (-99)

*Display This Question:*

*If Header Is Displayed*

CIDIA.TRE.2.0 Regarding times in your life when you have felt worried, tense or anxious: Did you **ever** try or are **currently** trying the following for the worry or the problems it caused? Please include any treatments that you have already told us about under 'depression' if they were also for anxiety. Select ALL that apply.

- ☐ Medication prescribed to you (for at least two weeks) (1)
- ☐ Specific anti-anxiety medication prescribed to you for at least one week (2)
- ☐ Unprescribed medication (more than once) (3)
- ☐ Drugs or alcohol (more than once) (4)
- ☐ Psychotherapy or other talking therapy more than once (including internet-based CBT) (5)
- ☐ Structured wellbeing activity (e.g. mindfulness, meditation, self-help book) (6)
- ☐ Regular physical exercise (e.g. yoga, running, walking) (7)
- ☐ ⊗None of the above (0)
- ☐ ⊗Prefer not to answer (-99)

*Display This Question:*

*If Did you ever use the following treatment for the anxiety? = Psychotherapy or other talking therapy more than once (including internet-based CBT)*

CIDIA.IAP.1.0 Are you **currently** enrolled in an **NHS** funded **talking therapy** or **psychotherapy** (IAPT)?

- Yes (1)
- No (0)
- Don't know (-88)

*Display This Question:*

*If Did you ever use the following treatment for the anxiety? = Medication prescribed to you (for at least two weeks)*

*Or Did you ever use the following treatment for the anxiety? = Specific anti-anxiety medication prescribed to you for at least one week*

CIDIA.TRE.3.0 Did you take your medication **as advised**?

- Yes (1)
- No (0)
- Don't know (-88)
- Prefer not to answer (-99)

---

*Display This Question:*

*If Did you ever use the following treatment for the anxiety? = Medication prescribed to you (for at least two weeks)*

*Or Did you ever use the following treatment for the anxiety? = Specific anti-anxiety medication prescribed to you for at least one week*

CIDIA.TRE.4.0 Did you find the medication **helpful**?

- Yes (1)
- No (0)
- Don't know (-88)
- Prefer not to answer (-99)

*Display This Question:*

*If Header Is Displayed*

*And Did you ever use the following treatment for the anxiety? = Psychotherapy or other talking therapy more than once (including internet-based CBT)*

*Or Did you ever use the following treatment for the anxiety? = Structured wellbeing activity (e.g. mindfulness, meditation, self-help book)*

CIDIA.THE.1.0 You previously mentioned that you have tried psychotherapy, another talking therapy, or a structured well-being activity for **anxiety**. Please **select all** that you attended **more than once**.

- ▢ Counselling (1)
- ▢ Group therapy (2)
- ▢ Cognitive Behavioral Therapy (CBT) (3)
- ▢ Mindfulness (4)
- ▢ Guided self-help (5)
- ▢ Workshops (6)
- ▢ Online therapy (7)
- ▢ Family therapy (9)
- ▢ Relationship therapy (10)
- ▢ Other: (8) ________________________________________________
- ▢ ⊗Never tried psychotherapy or other talking therapies (0)
- ▢ ⊗Don't know (-88)
- ▢ ⊗Prefer not to answer (-99)

*Display This Question:*

*If Did you ever use the following treatment for the anxiety? = Psychotherapy or other talking therapy more than once (including internet-based CBT)*

*And Which talking therapy or psychotherapy did you try for anxiety? != Never tried psychotherapy or other talking therapies*

CIDIA.THE.2.0 Did you **complete** your **course** of psychotherapy or other talking therapy?

- Yes (1)
- No (0)
- Don't know (-88)
- Prefer not to answer (-99)

*Display This Question:*

*If Header Is Displayed*

CIDIA.SYM.19.0
Regarding times in your life when you have felt **worried, tense or anxious:**

Think about your **roles** at the time of this episode, including study/employment, childcare and housework, leisure pursuits. How much did these problems **interfere** with your life or activities?

- A lot (3)
- Some (2)
- A little (1)
- Not at all (0)
- Prefer not to answer (-99)

HEAD The next set of questions is about **unusual experiences** that you may have had, like **seeing visions** or **hearing voices**.

We believe that these things may be quite common, but we don't know for sure, so please take your time and think carefully before answering.

CIDIP.1.0 Did you **ever** see something that **wasn't** really there or that other people could not see?

*(Please do not include any times when you were dreaming or half-asleep or under the influence of alcohol or drugs.)*

- Yes (1)
- No (0)
- Don't know (-88)
- Prefer not to answer (-99)

*Display This Question:*

*If Did you ever see something that wasn't really there? = Yes*

CIDIP.2.0 About **how many times** in your lifetime did you see things in this manner?

▼ 1 (1) ... 13+ (13)

CIDIP.3.0 Did you **ever** hear things that other people said did **not** exist, like **strange voices** coming from **inside your head** talking to you or about you, or voices coming **out of the air** when there was no one around?

*(Please do not include any times when you were dreaming or half-asleep or under the influence of alcohol or drugs.)*

- Yes (1)
- No (0)
- Don't know (-88)
- Prefer not to answer (-99)

*Display This Question:*

*If Did you ever hear voices that did not exist? = Yes*

CIDIP.4.0 About **how many times** in your life did you hear things in this manner?

▼ 1 (1) ... 13+ (13)

CIDIP.5.0 Did you **ever** believe that a **strange force** was trying to communicate directly with you by sending **special signs or signals** that you could understand but no one else could *(for example through the radio or television)?*

*(Please do not include any times when you were dreaming or half-asleep or under the influence of alcohol or drugs.)*

- Yes (1)
- No (0)
- Don't know (-88)
- Prefer not to answer (-99)

*Display This Question:*

*If Did you ever believe that a strange force was trying to communicate with you? = Yes*

CIDIP.6.0 About **how many times** in your life did this happen?

▼ 1 (1) ... 13+ (13)

CIDIP.7.0 Did you **ever** believe that there was an **unjust plot** going on to **harm you** or to have people **follow you**, and which your family and friends did **not** believe existed?

*(Please do not include any times when you were dreaming or half-asleep or under the influence of alcohol or drugs.)*

- Yes (1)
- No (0)
- Don't know (-88)
- Prefer not to answer (-99)

*Display This Question:*

*If Did you ever believe there was an unjust plot? = Yes*

CIDIP.8.0 About **how many times** in your life did this happen?

▼ 1 (1) ... 13+ (13)

*Display This Question:*

*If Did you ever see something that wasn't really there? = Yes*

*Or Did you ever hear voices that did not exist? = Yes*

*Or Did you ever believe that a strange force was trying to communicate with you? = Yes*

*Or Did you ever believe there was an unjust plot? = Yes*

CIDIP.9.0 How **often** did any of these experiences happen in the **past 1 year** (*seeing a vision, hearing a voice, or believing that something strange was trying to communicate with you, or there was a plot against you)?*

- ○ Not at all (0)
- ○ Once or twice (1)
- ○ **Less** than once a month (2)
- ○ **More** than once a month (3)
- ○ Nearly every day or daily (4)
- ○ Prefer not to answer (-99)

*Display This Question:*

*If How often did any of these experiences happen in the past 1 year? , Not at all Is Displayed*

CIDIP.10.0 **How old** were you *(approximately)* when you **first** had one of these experiences *(seeing a vision, hearing a voice, or believing something strange was trying to communicate with you, or there was a plot against you)?*

Please put your age in **years.** An **approximate** age is fine.

________________________________________________________________

*Display This Question:*

*If How often did any of these experiences happen in the past 1 year? , Not at all Is Displayed*

CIDIP.10.0 **Or select one of the options below:**

- ☐ ⊗ As long as I can remember (1)
- ☐ ⊗ Don't know (-88)
- ☐ ⊗ Prefer not to answer (-99)

*Display This Question:*

*If If How old were you when you first had one of these experiences? Text Response Is Displayed*

CIDIP.11.0 How **distressing** did you find having any of these experiences *(seeing a vision, hearing a voice, or believing that something strange was trying to communicate with you, or there was a plot against you)?*

- ○ **Not** distressing at all, it was a **positive** experience (1)
- ○ **Not** distressing at all, a **neutral** experience (2)
- ○ **A bit** distressing (3)
- ○ **Quite** distressing (4)
- ○ **Very** distressing (5)
- ○ Don't know (-88)
- ○ Prefer not to answer (-99)

*Display This Question:*

*If How distressing did you find having any of these experiences? , <b>Not</b> distressing at all, it was a <b>positive</b> experience Is Displayed*

CIDIP.12.0 Did you **ever** talk to a doctor, counsellor, psychiatrist, or other health professional about any of these experiences *(seeing a vision, hearing a voice, or believing that something strange was trying to communicate with you, or there was a plot against you)?*

◯ Yes (1)

◯ No (0)

◯ Don't know (-88)

◯ Prefer not to answer (-99)

*Display This Question:*

*If Did you ever talk to a health professional about this psychosis? , Yes Is Displayed*

CIDIP.13.0 Were you **ever** prescribed a medication by a **health professional** for any of these experiences *(seeing a vision, hearing a voice, or believing that something is strange was trying to communicate with you, or there was a plot against you)?*

◯ Yes (1)

◯ No (0)

◯ Don't know (-88)

◯ Prefer not to answer (-99)

---

*Display This Question:*

*If Were you ever prescribed a medication for psychosis? = Yes*

CIDIP.13txt.0 **Which** medication(s) were you prescribed?

________________________________________________________________

### Edited UK BioBank MHQ Questionnaire

#### Section A: Some questions about your mental health

**Choose an answer for EACH statement below:**
Please tick the appropriate boxes

| | 1<br>Yes | 2<br>No | 3<br>Don't Know | 4<br>Prefer not to answer |
|---|---|---|---|---|
| **A1)** In your life, have you suffered from a period of mental distress that prevented you from doing your usual activities? | ☐ | ☐ | ☐ | ☐ |
| **A2)** In your life, did you seek or receive help from a professional (medical doctor, psychologist, social worker, counsellor, nurse, clergy, or other helping professional) for mental distress or illness, psychological problems or unusual experiences? | ☐ | ☐ | ☐ | ☐ |

#### Section B: We next want to ask a few questions about your mood and feelings recently

**B1) Over the last 2 weeks, how often have you been bothered by any of the following problems?**

| | 1<br>Not at all | 2<br>Several days | 3<br>More than half the days | 4<br>Nearly every day |
|---|---|---|---|---|
| **a.** Little interest or pleasure in doing things | ☐ | ☐ | ☐ | ☐ |
| **b.** Feeling down, depressed, or hopeless | ☐ | ☐ | ☐ | ☐ |
| **c.** Trouble falling or staying asleep, or sleeping too much | ☐ | ☐ | ☐ | ☐ |
| **d.** Feeling tired or having little energy | ☐ | ☐ | ☐ | ☐ |
| **e.** Poor appetite or overeating | ☐ | ☐ | ☐ | ☐ |
| **f.** Feeling bad about yourself or that you are a failure or have let yourself or your family down | ☐ | ☐ | ☐ | ☐ |
| **g.** Trouble concentrating on things, such as reading the newspaper or watching television | ☐ | ☐ | ☐ | ☐ |
| **h.** Moving or speaking so slowly that other people could have noticed? Or the opposite, being so fidgety or restless that you have been moving around a lot more than usual | ☐ | ☐ | ☐ | ☐ |
| **i.** Thoughts that you would be better off dead or of hurting yourself in some way | ☐ | ☐ | ☐ | ☐ |

**B2) Have you ever had a time in your life when you felt sad, blue, or depressed for two weeks or more in a row?**

☐ Yes

☐ No
☐ Prefer not to answer

**B3) Have you ever had a time in your life lasting <u>two weeks or more</u> when you lost interest in most things like hobbies, work, or activities that usually give you pleasure?**

☐ Yes
☐ No
☐ Prefer not to answer

#### If 'Yes' to either B2 or B3, continue to B4. If 'No'/'Prefer not to answer' to both B2 or B3, skip to B25

**B4) Please think of the <u>two-week period</u> in your life when your feelings of depression or loss of interest were worst:**

**B4a) How much of the day did these feelings usually last?**

☐ All day long
☐ Most of the day
☐ About half of the day
☐ Less than half of the day
☐ Don't know
☐ Prefer not to answer

**B4b) Did you feel this way:**

☐ Every day
☐ Almost every day
☐ Less often
☐ Don't know
☐ Prefer not to answer

**B5) Did you feel more <u>tired</u> out or <u>low on energy</u> than is usual for you?**

☐ Yes
☐ No
☐ Don't know
☐ Prefer not to answer

**In the next section (B6-B19), we would like to know more about your depression or low mood. *If you find this is too difficult, you can skip to B20 on pg.5.***

**B6) Did your <u>weight</u> change? (*do not include weight change as a side-effect of medication you were taking*)**

☐ Gained weight *(continue to B6a)*
☐ Lost weight *(continue to B6a)*
☐ Both gained weight and lost some weight during the episode (*continue to B6a*)
☐ Stayed about the same or was on a diet (skip to B7)
☐ Don't know (skip to B7)
☐ Prefer not to answer (skip to B7)

**B6a) Did your weight change by about 10lbs (4kg) or more?**

- ☐ Yes
- ☐ No
- ☐ Don't know
- ☐ Prefer not to answer

**B7) Did your sleep change? (*do not include sleep change as a side-effect of medication you were taking*)**

- ☐ Yes *(continue to B7a)*
- ☐ No *(skip to B8)*
- ☐ Don't know *(skip to B8)*
- ☐ Prefer not to answer *(skip to B8)*

| **B7a) Was that:** *(please select all that apply)* | **1 Yes** | **2 No** |
|---|---|---|
| Trouble falling asleep (sleeping too little) | ☐ | ☐ |
| Waking too early (sleeping too little) | ☐ | ☐ |
| Sleeping too much | ☐ | ☐ |
| Both sleeping too much and too little during the same depression episode | ☐ | ☐ |

**Any 'Yes' selections made in B7a lead to display of B7b and B7c**

**B7b) How many hours per day did you sleep on average during the depression episode, including nighttime sleep and daytime naps?**

☐☐

**B7c) How many hours per day did you used to sleep on average when you were not depressed?**

☐☐

**B8) Did you experience a change in your appetite?**

- ☐ No changes in appetite
- ☐ Increased appetite
- ☐ Decreased appetite
- ☐ Experienced both increased an decreased appetite during the same depression episode
- ☐ Don't know
- ☐ Prefer not to answer

**B9) Did your mood brighten in response to positive events?**

- ☐ Yes
- ☐ No
- ☐ Don't know
- ☐ Prefer not to answer

**B10) Did you experience heavy feelings in your arms or legs? (Did your arms or legs feel "heavy"?)**

☐ Yes *(continue to B10.1)*
☐ No *(skip to B11)*
☐ Don't know *(skip to B11)*
☐ Prefer not to answer *(skip to B11)*

**B10.1 For how many hours per day was the heaviness feeling present?**

☐☐

**B11) Where you overly sensitive to interpersonal rejection?**

☐ No
☐ Yes, and this significantly impaired your social or work relationships
☐ Yes, but this did not significantly impair your social or work relationships
☐ Don't know
☐ Prefer not to answer

**B12) Was your mood worse:**

☐ In the morning
☐ In the afternoon
☐ At night
☐ My mood did not vary
☐ Don't know
☐ Prefer not to answer

| | 1<br>Yes | 2<br>No | 3<br>Don't Know | 4<br>Prefer not to answer |
|---|---|---|---|---|
| **B13)** Did you have a lot more trouble concentrating than usual? | ☐ | ☐ | ☐ | ☐ |
| **B14)** People sometimes feel down on themselves, no good, worthless. Did you feel this way? | ☐ | ☐ | ☐ | ☐ |
| **B15)** Did you think a lot about death – either your own, someone else's or death in general? | ☐ | ☐ | ☐ | ☐ |

**B16) Roughly how long altogether did you feel this way?**

☐ Less than a month
☐ Between one and three months
☐ Over three months, but less than six months
☐ Over six months, but less than 12 months
☐ One to two years
☐ Over two years
☐ Don't know
☐ Prefer not to answer

**B17 Was this your longest episode of depression or low mood?**

☐ Yes (skip to B18) ☐ No ☐ Don't know (skip to B18) ☐ Prefer not to answer (skip to B18)

**B17a) What is the longest period of time that you have experienced depression or low mood?**

☐ Less than 6 months ☐ Between 6 and 12 months
☐ Between 1 and 5 years ☐ More than 5 years
☐ All of my life / as long as I can remember

**B18) Think about your roles at the time of this episode, including study/employment, childcare and housework, leisure pursuits. How much did these problems interfere with your life or activities?**

☐ A lot ☐ Some ☐ A little
☐ Not at all ☐ Prefer not to answer

**B19) How many periods of depression or low mood have you had in your life lasting two or more weeks?**

☐ One *(skip to B20)* ☐ two – three (continue to B19a)

☐ Several *(continue to B19a)* ☐ All of my life / as long as I can remember (skip to B20)

☐ Prefer not to answer *(skip to B20)*

**B19a) If you have had several, please estimate the number of times you have had periods of depression or low mood in your life lasting two or more weeks:**

☐☐

**B20) About how old were you the first time you had a period of two weeks like this? (Whether or not you received any help for it)**

☐☐

B21 is not displayed to those who answered "all my life/ as long as I can remember" in B19 or B17a

**B21) About how old were you the last time you had a period of two weeks like this? (Whether or not you received any help for it)**

☐☐

**B22) Did any of these episodes occur within the first year of giving birth? Or has it been suggested you had post-natal depression?**

☐ Yes ☐ No ☐ Don't know

☐ Prefer not to answer ☐ Not applicable

**B21.1) Did any of these episodes occur following a significant or traumatic event such as death/serious illness of close relative or friend, or following an distressing event or illness that happened to you?**

☐ Most/All ☐ More than once ☐ Once
☐ Not at all ☐ Prefer not to answer

**B22) Did you ever tell a professional about these problems?** *(Medical doctor, psychologist, social worker, counsellor, nurse, clergy, or other helping professional)*

☐ Yes ☐ No
☐ Don't know ☐ Prefer not to answer

**B23) Did you ever try the following for these problems?**

Tick ALL that apply:

☐ Medication prescribed to you for at least two weeks
☐ Unprescribed medication more than once
☐ Drugs or alcohol more than once
☐ Psychotherapy or other talking therapy more than once (including internet-based CBT)
☐ Structured wellbeing activity (e.g. mindfulness, meditation, self-help book)
☐ Prefer not to answer
☐ None of the above
☐ Regular physical exercise (e.g. yoga, running, walking)

**B23a) Are you currently enrolled in an NHS funded talking therapy or psychotherapy (IAPT)?**

☐ Yes ☐ No
☐ Don't know

#### B23b and B23c are only displayed to those who selected 'prescribed medication' in B23

**B23b) Did you take your medication as advised?**

☐ Yes ☐ No
☐ Don't know ☐ Prefer not to answer

**B23c) Did you find the medication useful?**

☐ Yes ☐ No
☐ Don't know ☐ Prefer not to answer

**B24 is only displayed to those who selected 'psychotherapy or other talking therapy' in B23**

**B24) You previously mentioned that you have tried psychotherapy or another talking therapy, please select all that you attended more than once.**

- ☐ Counselling
- ☐ Group therapy
- ☐ Cognitive Behavioral Therapy (CBT)
- ☐ Mindfulness
- ☐ Guided self-help
- ☐ Workshops
- ☐ Relationship therapy
- ☐ Family therapy
- ☐ Online therapy
- ☐ Never tried psychotherapy or other talking therapies
- ☐ Prefer not to answer
- ☐ Other

**24a) Did you complete your course of psychotherapy or other talking therapy?**

- ☐ Yes
- ☐ No
- ☐ Don't know
- ☐ Prefer not to answer

**24b) Did you find psychotherapy or other talking therapy useful?**

- ☐ Yes
- ☐ No
- ☐ Don't know
- ☐ Prefer not to answer

**Questions B25.1-B25.4 will only display to participants with a score of 5 or more on B1 and selected 'yes' to either B2 or 'yes' to B3**

**B25.1) How long ago did your current or most recent episode of depression or low mood begin?**

- ☐ Less than 1 year ago *(continue to B1.2)*
- ☐ 1 to 2 years ago *(continue to B1.2)*
- ☐ More than 2 years ago *(continue to B1.2)*

**B25.2) During the current or most recent episode of depression or low mood, how many antidepressant medications have you taken for 6 weeks or longer?**

- ☐ I have not taken medication during my current episode of depression *(continue to B1.3)*
- ☐ 1-2 medications *(continue to B1.3)*
- ☐ 3-4 medications *(continue to B1.3)*
- ☐ 5-6 medications *(continue to B1.3)*
- ☐ 7-10 medications *(continue to B1.3)*
- ☐ >10 medications *(continue to B1.3)*

**B25.3) If people don't respond fully to antidepressants, doctors sometime prescribe "add-on" or "augmentation" medications in addition to the antidepressant (such as lithium, quetiapine or aripiprazole).**

**During the current or most recent episode of depression or low mood, have you taken an add-on medication for 6 weeks or longer?**

- ☐ Yes *(continue to B1.4)*
- ☐ No *(continue to B1.4)*
- ☐ Prefer not to answer *(continue to B1.4)*

**B25.4) Have you received electroconvulsive therapy (ECT) in this current or most recent episode of depression or low mood? (Please only answer yes if your course of ECT included 8 treatment sessions or more.)**

- ☐ Yes *(continue to B2*
- ☐ No *(continue to B2)*
- ☐ Prefer not to answer *(continue to B2)*

**Now we want to know about some different symptoms in your lifetime.**

**B25) Has there ever been a period of time when you were not your usual self and experienced any of the following?**

Tick ALL that apply.

- ☐ ...you felt so good or so hyper that other people thought you were not your normal self or you were so hyper that you got into trouble?
- ☐ ...you were so irritable that you shouted at people or start fights or arguments?
- ☐ ...you felt much more self-confident than usual?
- ☐ ...you got much less sleep than usual and found you didn't really miss it?
- ☐ ...you were much more talkative or spoke much faster than usual?
- ☐ …thoughts raced through your head or you couldn't slow your mind down?
- ☐ ...you were so easily distracted by things around you that you had trouble concentrating or staying on track?
- ☐ ...you had much more energy than usual?
- ☐ ...you were much more active or did many more things than usual?
- ☐ ...you were much more social or outgoing than usual, for example, you telephoned friends in the middle of the night?
- ☐ ...you were much more interested in sex than usual?
- ☐ ...you did things that were unusual for you or that other people might have thought were excessive, foolish, or risky?
- ☐ ...spending money got you or your family into trouble?
- ☐ None of these (skip to B30)
- ☐ Prefer not to answer

**B26) If you ticked any of the above, have several of these ever happened during the same period of time?**

- ☐ Yes
- ☐ No (skip to B28)
- ☐ Don't know
- ☐ Prefer not to answer

**B27) Please tick all that occurred during the same period of time.**

**Tick ALL that apply.**

- ☐ ...you felt so good or so hyper that other people thought you were not your normal self or you were so hyper that you got into trouble?
- ☐ ...you were so irritable that you shouted at people or start fights or arguments?
- ☐ ...you felt much more self-confident than usual?
- ☐ ...you got much less sleep than usual and found you didn't really miss it?
- ☐ ...you were much more talkative or spoke much faster than usual?
- ☐ ...thoughts raced through your head or you couldn't slow your mind down?
- ☐ ...you were so easily distracted by things around you that you had trouble concentrating or staying on track?
- ☐ ...you had much more energy than usual?
- ☐ ...you were much more active or did many more things than usual?
- ☐ ...you were much more social or outgoing than usual, for example, you telephoned friends in the middle of the night?
- ☐ ...you were much more interested in sex than usual?
- ☐ ...you did things that were unusual for you or that other people might have thought were excessive, foolish, or risky?
- ☐ ...spending money got you or your family into trouble?

**B28) What is the longest time that these "high" or "irritable" periods have lasted?**

- ☐ Less than 24 hours
- ☐ At least a day
- ☐ Four days to one week
- ☐ A week or more
- ☐ Don't know
- ☐ Prefer not to answer

**B29) How much of a problem did any of these cause you - like being unable to work; having family, money or legal troubles; getting into arguments or fights?**

- ☐ No problem
- ☐ Minor problem
- ☐ Moderate problem
- ☐ Serious problem
- ☐ Don't know
- ☐ Prefer not to answer

**B30) Has a health professional ever told you that you have manic-depressive illness or bipolar disorder?**

☐ Yes ☐ No
☐ Don't know ☐ Prefer not to answer

**B31) Have any of your blood relatives (i.e. children, siblings, parents, grandparents, aunts, uncles) had manic-depressive illness or bipolar disorder?**

☐ Yes ☐ No
☐ Don't know ☐ Prefer not to answer

**B32) During any of your episodes of depression or mania, were you also diagnosed with psychosis?**

***(hearing voices or seeing things that other people said did not exist or believing that you had special powers, that you were in danger, that others were trying to communicate with you in unusual ways or that a catastrophe was imminent)***

☐ Yes ☐ No
☐ Don't know ☐ Prefer not to answer

**B33) Have you ever had an episode where psychosis was the primary symptom or diagnosis?**

☐ Yes ☐ No
☐ Don't know ☐ Prefer not to answer

#### Section C: Some questions about anxiety or nerves

**C1) Over the last 2 weeks, how often have you been bothered by any of the following problems?**

Tick ONE for each of the following statements:

| LAST 2 WEEKS | 1 Not at all | 2 Several days | 3 More than half the days | 4 Nearly every day |
|---|---|---|---|---|
| **a.** Feeling nervous, anxious or on edge | ☐ | ☐ | ☐ | ☐ |
| **b.** Not being able to stop or control worrying | ☐ | ☐ | ☐ | ☐ |
| **c.** Worrying too much about different things | ☐ | ☐ | ☐ | ☐ |
| **d.** Trouble relaxing | ☐ | ☐ | ☐ | ☐ |
| **e.** Being so restless that it is hard to sit still | ☐ | ☐ | ☐ | ☐ |
| **f.** Becoming easily annoyed or irritable | ☐ | ☐ | ☐ | ☐ |
| **g.** Feeling afraid as if something awful might happen | ☐ | ☐ | ☐ | ☐ |

**C2a) Have you ever had a period lasting one month or longer when most of the time you felt worried, tense, or anxious?**

☐ Yes *(continue to C2b)*
☐ No *(skip to C3)*
☐ Don't know *(skip to C3)*
☐ Prefer not to answer *(skip to C3)*

**C2b) What is the longest period of time that this kind of worrying has ever continued?**

☐ Less than 6 months (skip to C2e)
☐ Between 6 and 12 months
☐ Between 1 and 5 years
☐ More than 5 years
☐ All of my life / as long as I can remember (skip to C2e)

**C2c) How many periods of this kind of worry have you had in your life lasting 6 or more months?**

☐ One (skip to C2e)
☐ Two-three (skip to C2d)
☐ Several (continue to C2d)
☐ All of my life / as long as I can remember (skip to C2e)
☐ Prefer not to answer (skip to C2e)

**C2d) Please estimate the number of times you have had periods of this kind of worry in your life lasting 6 or more months:**

☐☐

**C2e) About how old were you the first time you had a period of six months like this? (Whether or not you received any help for it.) Please put your age in years. An approximate age is fine.**

☐☐

**C2f is not displayed to those who answered "all my life/ as long as I can remember" in C2b or in C2c**

**C2f) About how old were you the last time you had a period of six months like this? (Whether or not you received any help for it.) Please put your age in years. An approximate age is fine.**

☐☐

**C3) People differ a lot in how much they worry about things. Did you ever have a time when you worried a lot <u>more than most people</u> would in your situation?**

- ☐ Yes *(continue to C4)*
- ☐ No *(skip to Section D on pg. 12)*
- ☐ Don't know *(skip to Section D on pg. 12)*
- ☐ Prefer not to answer *(skip to Section D on pg. 12)*

**Please think of the period in your life when you have felt worried, tense, anxious, or more worried than most people would in your situation. This could be in the past, or it could be continuing now.**
*If this lasted for 6 months or greater, please continue to C4.*
*If this lasted less than 6 months, please skip to Section D.*

Tick an answer for each statement:

| | **1**<br>**Yes** | **2**<br>**No** | **3**<br>**Don't Know** | **4**<br>**Prefer not to answer** |
|---|---|---|---|---|
| **C4) During that period, was your worry <u>stronger</u> than in <u>other people</u>?** | ☐ | ☐ | ☐ | ☐ |
| **C5) Did you worry <u>most days</u>?** | ☐ | ☐ | ☐ | ☐ |

| | **1**<br>**One thing** | **2** | **3**<br>**Don't Know** | **4** |
|---|---|---|---|---|

| | | More than one thing | | Prefer not to answer |
|---|---|---|---|---|
| **C6) Did you usually worry about <u>one particular thing</u>, such as your job security or the failing health of a loved one, or <u>more than one thing</u>?** | ☐ | ☐ | ☐ | ☐ |

| | 1 Yes | 2 No | 3 Don't Know | 4 Prefer not to answer |
|---|---|---|---|---|
| **C7) Did you find it <u>difficult to stop</u> worrying?** | ☐ | ☐ | ☐ | ☐ |
| **C8) Did you ever have different worries on your mind <u>at the same time?</u>** | ☐ | ☐ | ☐ | ☐ |

| | 1 Often | 2 Sometimes | 3 Rarely | 4 Never | 5 Don't know | 6 Prefer not to say |
|---|---|---|---|---|---|---|
| **C9) How often was your worry so strong that you <u>couldn't</u> put it out of your mind no matter how hard you tried?** | ☐ | ☐ | ☐ | ☐ | ☐ | ☐ |
| **C10) How often did you find it <u>difficult to control</u> your worry?** | ☐ | ☐ | ☐ | ☐ | ☐ | ☐ |

**C11) When you were worried or anxious, were you also:**

Tick an answer for each statement:

| | 1 Yes | 2 No | 3 Don't Know |
|---|---|---|---|
| **a.** Restless? | ☐ | ☐ | ☐ |
| **b.** Keyed up or on edge? | ☐ | ☐ | ☐ |
| **c.** Easily tired? | ☐ | ☐ | ☐ |
| **d.** Having difficulty keeping your mind on what you were doing? | ☐ | ☐ | ☐ |
| **e.** More irritable than usual? | ☐ | ☐ | ☐ |
| **f.** Having tense, sore, or aching muscles? | ☐ | ☐ | ☐ |
| **g.** Often have trouble falling or staying asleep? | ☐ | ☐ | ☐ |

**C12) Did you <u>ever</u> tell a <u>professional</u> about these problems (medical doctor, psychologist, social worker, counsellor, nurse, clergy, or other helping professional)?**

☐ Yes ☐ No

☐ Don't know ☐ Prefer not to answer

**C13) Did you ever use the following for the worry or the problems it caused? Please include any treatments that you have already told us about under 'depression' if they were also for anxiety:**

**Tick ALL that apply**

- ☐ Medication prescribed to you for at least two weeks
- ☐ Specific anti-anxiety medication prescribed to you for at least one week
- ☐ Unprescribed medication more than once
- ☐ Drugs or alcohol more than once
- ☐ Psychotherapy or other talking therapy more than once (including internet based CBT)
- ☐ Structured wellbeing activity (e.g. mindfulness, meditation, self-help book)_
- ☐ Regular physical exercise (e.g. yoga, running, walking)
- ☐ Prefer not to answer
- ☐ None of the above

**C13a) Are you currently enrolled in an NHS funded talking therapy or psychotherapy (IAPT)?**

- ☐ Yes
- ☐ No
- ☐ Don't know

**C13b and C13c is only displayed to those who selected 'prescribed medication' in B13**

**C13b) Did you take your medication as advised?**

- ☐ Yes
- ☐ No
- ☐ Don't know
- ☐ Prefer not to answer

**C13cb) Did you find the medication useful?**

- ☐ Yes
- ☐ No
- ☐ Don't know
- ☐ Prefer not to answer

**C14 is only displayed to those who selected 'psychotherapy or other talking therapy' in C13**

**C14) You previously mentioned that you have tried psychotherapy or another talking therapy, please select all that you attended more than once.**

- ☐ Counselling
- ☐ Group therapy
- ☐ Cognitive Behavioral Therapy (CBT)
- ☐ Psychotherapy
- ☐ Mindfulness
- ☐ Guided self-help
- ☐ Workshops
- ☐ Online therapy
- ☐ Psychodynamic
- ☐ Psychoanalysis
- ☐ Family therapy
- ☐ Relationship therapy
- ☐ Never tried psychotherapy or other talking therapies
- ☐ Prefer not to answer
- ☐ Other

**C14a) Did you complete your course of psychotherapy or other talking therapy?**

- ☐ Yes
- ☐ No
- ☐ Don’t know
- ☐ Prefer not to answer

**C14b) Did you find psychotherapy or other talking therapy useful?**

- ☐ Yes
- ☐ No
- ☐ Don’t know
- ☐ Prefer not to answer

**C15) Think about your roles at the time of this episode, including study/employment, childcare and housework, leisure pursuits. How much did these problems interfere with your life or activities?**

- ☐ A lot
- ☐ Some
- ☐ A little
- ☐ Not at all
- ☐ Prefer not to answer

#### Section D: The next set of questions is about unusual experiences that you may have had, like seeing visions or hearing voices.

**We believe that these things may be quite common, so please take your time and think carefully before answering.**

**E1) Did you ever see something that wasn't really there that other people could not see?**
Please do not include any times when you were dreaming or half-asleep or under the influence of alcohol or drugs.

- ☐ Yes *(continue to E1a)*
- ☐ No *(skip to E2)*
- ☐ Don't know *(skip to E2)*
- ☐ Prefer not to answer *(skip to E2)*

**E1a) About how many times in your lifetime did you see things in this manner?**

☐☐

**E2) Did you ever hear things that other people said did not exist, like strange voices coming from inside your head talking to you or about you, or voices coming out of the air when there was no one around?**
Please do not include any times when you were dreaming or half-asleep or under the influence of alcohol or drugs.

- ☐ Yes *(continue to E2a)*
- ☐ No *(skip to E3)*
- ☐ Don't know *(skip to E3)*
- ☐ Prefer not to answer *(skip to E3)*

**E2a) About how many times in your lifetime did you hear things in this manner?**

☐☐

**E3) Did you ever believe that a strange force was trying to communicate directly with you by sending special signs or signals that you could understand but that no one else could understand (for example through the radio or television)?**
Please do not include any times when you were dreaming or half-asleep or under the influence of alcohol or drugs.

- ☐ Yes *(continue to E3a)*
- ☐ No *(skip to E4)*
- ☐ Don't know *(skip to E4)*
- ☐ Prefer not to answer *(skip to E4)*

**E3a) About how many times in your life did this happen?**

☐☐

**E4) Did you ever believe that that there was an unjust plot going on to harm you or others, or to have people follow you, and which your family and friends did not believe existed?**
Please do not include any times when you were dreaming or half-asleep or under the influence of alcohol or drugs.

- ☐ Yes *(continue to E4a)*
- ☐ No *(skip to E5)*
- ☐ Don't know *(skip to E5)*
- ☐ Prefer not to answer *(skip to E5)*

**E4a) About how many times in your life did this happen?**

☐☐

**If you have answered 'yes' to E1, E2, E3 or E4, please continue.**
**If you have not answered 'yes' to E1, E2, E3 or E4 please skip to Section F**

**E5) How often did any of these experiences happen in the past 1 year (seeing a vision, hearing a voice, or believing that something strange was trying to communicate with you, or there was a plot against you)?**

- ☐ Not at all
- ☐ Once or twice
- ☐ Less than once a month
- ☐ More than once a month
- ☐ Nearly every day or daily
- ☐ Prefer not to answer

**E6) How old were you (approximately) when you first had one of these experiences (seeing a vision, hearing a voice, or believing that something strange was trying to communicate with you, or there was a plot against you)?**

- ☐ Please specify age (in years): ______
- ☐ As long as I can remember
- ☐ Don't remember
- ☐ Prefer not to answer

**E7) How distressing did you find having any of these experiences (seeing a vision, hearing a voice, or believing that something strange was trying to communicate with you, or there was a plot against you)?**

- ☐ Not distressing at all, it was a positive experience
- ☐ Not distressing, a neutral experience
- ☐ A bit distressing
- ☐ Quite distressing
- ☐ Very distressing
- ☐ Don't know
- ☐ Prefer not to answer

**E8) Did you ever talk to a doctor, counsellor, psychiatrist or other health professional about any of these experiences (seeing a vision, hearing a voice, or believing that something strange was trying to communicate with you, or there was a plot against you)?**

- ☐ Yes
- ☐ No
- ☐ Don't know
- ☐ Prefer not to answer

**E9) Were you ever prescribed a medication by a health professional for any of these experiences (seeing a vision, hearing a voice, or believing that something strange was trying to communicate with you, or there was a plot against you)?**

- ☐ Yes
- ☐ No
- ☐ Don't know
- ☐ Prefer not to answer

**E9a) If yes, which medication were you prescribed? _________________________**

**Section E: This section asks about your childhood and some possible stresses and strains of life. If this is too difficult, then *please skip to section G on pg. 24.***

| F1) When I was growing up… | 1 Never true | 2 Rarely true | 3 Sometimes true | 4 Often | 5 Very often true | 6 Prefer not to answer |
|---|---|---|---|---|---|---|
| **a.** I felt loved | ☐ | ☐ | ☐ | ☐ | ☐ | ☐ |
| **b.** People in my family hit me so hard that it left me with bruises or marks | ☐ | ☐ | ☐ | ☐ | ☐ | ☐ |
| **c.** I felt that someone in my family hated me | ☐ | ☐ | ☐ | ☐ | ☐ | ☐ |
| **d.** Someone molested me (sexually) | ☐ | ☐ | ☐ | ☐ | ☐ | ☐ |
| **e.** There was someone to take me to the doctor if I needed it | ☐ | ☐ | ☐ | ☐ | ☐ | ☐ |

| F2) Since I was sixteen… | 1 Never | 2 Yes, but not in the last 12 months | 3 Yes, within the last 12 months | 4 Prefer not to answer |
|---|---|---|---|---|
| **a.** I have been in a confiding relationship | ☐ | ☐ | ☐ | ☐ |
| **b.** A partner or ex-partner deliberately hit me or used violence in any other way | ☐ | ☐ | ☐ | ☐ |
| **c.** A partner or ex-partner repeatedly belittled me to the extent that I felt worthless | ☐ | ☐ | ☐ | ☐ |
| **d.** A partner or ex-partner sexually interfered with me, or forced me to have sex against my wishes | ☐ | ☐ | ☐ | ☐ |
| **e.** I have had the money to pay my rent/mortgage payment | ☐ | ☐ | ☐ | ☐ |

**F2a) If so, how often have these statements been true?**

| | 1 Never true | 2 Rarely true | 3 Sometimes true | 4 Often | 5 Very often true | 6 Prefer not to answer |
|---|---|---|---|---|---|---|
| **a.** I have been in a confiding relationship | ☐ | ☐ | ☐ | ☐ | ☐ | ☐ |
| **b.** A partner or ex-partner deliberately hit me or used violence in any other way | ☐ | ☐ | ☐ | ☐ | ☐ | ☐ |
| **c.** A partner or ex-partner repeatedly belittled me to the extent that I felt worthless | ☐ | ☐ | ☐ | ☐ | ☐ | ☐ |

| | | | | | | |
|---|---|---|---|---|---|---|
| **d.** A partner or ex-partner sexually interfered with me, or forced me to have sex against my wishes | ☐ | ☐ | ☐ | ☐ | ☐ | ☐ |
| **e.** I have had the money to pay my rent/mortgage payment | ☐ | ☐ | ☐ | ☐ | ☐ | ☐ |

| **F3) In your life, have you…?** | 1<br>Never | 2<br>Yes, but not in the last 12 months | 3<br>Yes, within the last 12 months | 4<br>Prefer not to answer |
|---|---|---|---|---|
| **a.** Been a victim of a sexual assault, whether by a stranger or someone you knew | ☐ | ☐ | ☐ | ☐ |
| **b.** Been attacked, mugged, robbed, or been the victim of a physically violent crime | ☐ | ☐ | ☐ | ☐ |
| **c.** Been in a serious accident that you believed to be life-threatening at the time | ☐ | ☐ | ☐ | ☐ |
| **d.** Witnessed a sudden violent death (eg. murder, suicide, aftermath of an accident) | ☐ | ☐ | ☐ | ☐ |
| **e.** Been diagnosed with a life-threatening illness | ☐ | ☐ | ☐ | ☐ |
| **f)** Been involved in combat or exposed to a war-zone (either in the military or as a civilian) | ☐ | ☐ | ☐ | ☐ |

**F4) Next is a list of problems and complaints that people sometimes have in response to such extremely stressful experiences. Please indicate how much you have been bothered by that problem in the past month:**

| | 1<br>Not at all | 2<br>A little bit | 3<br>Moderately | 4<br>Quite a bit | 5<br>Extremely | 6<br>Prefer not to answer |
|---|---|---|---|---|---|---|
| a. Repeated, disturbing memories, thoughts, or images of a stressful experience? | ☐ | ☐ | ☐ | ☐ | ☐ | ☐ |
| b. Feeling very upset when something reminded you of a stressful experience? | ☐ | ☐ | ☐ | ☐ | ☐ | ☐ |
| c. Avoiding activities or situations because they reminded you of a stressful experience? | ☐ | ☐ | ☐ | ☐ | ☐ | ☐ |
| d. Feeling distant or cut off from other people? | ☐ | ☐ | ☐ | ☐ | ☐ | ☐ |
| e. Feeling irritable or having angry outbursts? | ☐ | ☐ | ☐ | ☐ | ☐ | ☐ |
| f. Difficulty concentrating? | ☐ | ☐ | ☐ | ☐ | ☐ | ☐ |

#### Section F: This section relates to how you feel about things in general

**G1. In general, how happy are you?**

- ☐ Extremely happy
- ☐ Very happy
- ☐ Moderately happy
- ☐ Moderately unhappy
- ☐ Very unhappy
- ☐ Extremely unhappy
- ☐ Don't know
- ☐ Prefer not to answer

**G2. In general, how happy are you with your health?**

- ☐ Extremely happy
- ☐ Very happy
- ☐ Moderately happy
- ☐ Moderately unhappy
- ☐ Very unhappy
- ☐ Extremely unhappy
- ☐ Don't know
- ☐ Prefer not to answer

**G3. To what extent do you feel your life to be meaningful?**

- ☐ Not at all
- ☐ A little
- ☐ A moderate amount
- ☐ Very much
- ☐ An extreme amount
- ☐ Don't know
- ☐ Prefer not to answer

### GAD-7 Anxiety

| Over the last two weeks, how often have you been bothered by the following problems? | Not at all | Several days | More than half the days | Nearly every day |
|---|---|---|---|---|
| 1. Feeling nervous, anxious, or on edge | 0 | 1 | 2 | 3 |
| 2. Not being able to stop or control worrying | 0 | 1 | 2 | 3 |
| 3. Worrying too much about different things | 0 | 1 | 2 | 3 |
| 4. Trouble relaxing | 0 | 1 | 2 | 3 |
| 5. Being so restless that it is hard to sit still | 0 | 1 | 2 | 3 |
| 6. Becoming easily annoyed or irritable | 0 | 1 | 2 | 3 |
| 7. Feeling afraid, as if something awful might happen | 0 | 1 | 2 | 3 |

Column totals _____ + _____ + _____ + _____ =

*Total score* ________

If you checked any problems, how difficult have they made it for you to do your work, take care of things at home, or get along with other people?

| Not difficult at all | Somewhat difficult | Very difficult | Extremely difficult |
|---|---|---|---|
| ☐ | ☐ | ☐ | ☐ |

Source: Primary Care Evaluation of Mental Disorders Patient Health Questionnaire (PRIME-MD-PHQ). The PHQ was developed by Drs. Robert L. Spitzer, Janet B.W. Williams, Kurt Kroenke, and colleagues. For research information, contact Dr. Spitzer at. PRIME-MD® is a trademark of Pfizer Inc. Copyright© 1999 Pfizer Inc. All rights reserved. Reproduced with permission

### Scoring GAD-7 Anxiety Severity

This is calculated by assigning scores of 0, 1, 2, and 3 to the response categories, respectively, of "not at all," "several days," "more than half the days," and "nearly every day."
GAD-7 total score for the seven items ranges from 0 to 21.

0–4: minimal anxiety

5–9: mild anxiety

10–14: moderate anxiety

15–21: severe anxiety

### THE MOOD DISORDER QUESTIONNAIRE

**Instructions:** Please answer each question to the best of your ability.

| | YES | NO |
|---|---|---|
| 1. Has there ever been a period of time when you were not your usual self and... | | |
| ...you felt so good or so hyper that other people thought you were not your normal self or you were so hyper that you got into trouble? | ❍ | ❍ |
| ...you were so irritable that you shouted at people or started fights or arguments? | ❍ | ❍ |
| ...you felt much more self-confident than usual? | ❍ | ❍ |
| ...you got much less sleep than usual and found you didn't really miss it? | ❍ | ❍ |
| ...you were much more talkative or spoke much faster than usual? | ❍ | ❍ |
| ...thoughts raced through your head or you couldn't slow your mind down? | ❍ | ❍ |
| ...you were so easily distracted by things around you that you had trouble concentrating or staying on track? | ❍ | ❍ |
| ...you had much more energy than usual? | ❍ | ❍ |
| ...you were much more active or did many more things than usual? | ❍ | ❍ |
| ...you were much more social or outgoing than usual, for example, you telephoned friends in the middle of the night? | ❍ | ❍ |
| ...you were much more interested in sex than usual? | ❍ | ❍ |
| ...you did things that were unusual for you or that other people might have thought were excessive, foolish, or risky? | ❍ | ❍ |
| ...spending money got you or your family into trouble? | ❍ | ❍ |
| 2. If you checked YES to more than one of the above, have several of these ever happened during the same period of time? | ❍ | ❍ |

3. How much of a problem did any of these cause you – like being unable to work; having family, money or legal troubles; getting into arguments or fights? *Please circle one response only.*

No Problem     Minor Problem     Moderate Problem     Serious Problem

| | YES | NO |
|---|---|---|
| 4. Have any of your blood relatives (i.e. children, siblings, parents, grandparents, aunts, uncles) had manic-depressive illness or bipolar disorder? | ❍ | ❍ |
| 5. Has a health professional ever told you that you have manic-depressive illness or bipolar disorder? | ❍ | ❍ |

© 2000 by The University of Texas Medical Branch. Reprinted with permission. This instrument is designed for screening purposes only and is not to be used as a diagnostic tool.

### SCORING THE MOOD DISORDER QUESTIONNAIRE (MDQ)

The MDQ was developed by a team of psychiatrists, researchers and consumer advocates to address a critical need for timely and accurate diagnosis of bipolar disorder, which can be fatal if left untreated. The questionnaire takes about five minutes to complete, and can provide important insights into diagnosis and treatment. Clinical trials have indicated that the MDQ has a high rate of accuracy; it is able to identify seven out of ten people who have bipolar disorder and screen out nine out of ten people who do not.[1]

A recent National DMDA survey revealed that nearly 70% of people with bipolar disorder had received at least one misdiagnosis and many had waited more than 10 years from the onset of their symptoms before receiving a correct diagnosis. National DMDA hopes that the MDQ will shorten this delay and help more people to get the treatment they need, when they need it.

The MDQ screens for Bipolar Spectrum Disorder, (which includes Bipolar I, Bipolar II and Bipolar NOS).

**If the patient answers:**

1. **"Yes"** to seven or more of the 13 items in question number 1;

AND

2. **"Yes"** to question number 2;

AND

3. **"Moderate"** or **"Serious"** to question number 3;

you have a positive screen. All three of the criteria above should be met. A positive screen should be followed by a comprehensive medical evaluation for Bipolar Spectrum Disorder.

**ACKNOWLEDGEMENT:** This instrument was developed by a committee composed of the following individuals: Chairman, Robert M.A. Hirschfeld, MD – University of Texas Medical Branch; Joseph R. Calabrese, MD – Case Western Reserve School of Medicine; Laurie Flynn – National Alliance for the Mentally Ill; Paul E. Keck, Jr., MD – University of Cincinnati College of Medicine; Lydia Lewis – National Depressive and Manic-Depressive Association; Robert M. Post, MD – National Institute of Mental Health; Gary S. Sachs, MD – Harvard University School of Medicine; Robert L. Spitzer, MD – Columbia University; Janet Williams, DSW – Columbia University and John M. Zajecka, MD – Rush Presbyterian-St. Luke's Medical Center.

[1] Hirschfeld, Robert M.A., M.D., Janet B.W. Williams, D.S.W., Robert L. Spitzer, M.D., Joseph R. Calabrese, M.D., Laurie Flynn, Paul E. Keck, Jr., M.D., Lydia Lewis, Susan L. McElroy, M.D., Robert M. Post, M.D., Daniel J. Rapport, M.D., James M. Russell, M.D., Gary S. Sachs, M.D., John Zajecka, M.D., "Development and Validation of a Screening Instrument for Bipolar Spectrum Disorder: The Mood Disorder Questionnaire." *American Journal of Psychiatry* 157:11 (November 2000) 1873-1875.

Identity Number_____

### The WHOQOL-Bref
### UK Version

**Department of Mental Health**

**World Health Organisation**

**Geneva**

This document is not issued to the general public and all rights are reserved by the World Health Organisation (WHO). This document may not be reviewed, abstracted, quoted, reproduced, translated, referred to in bibliographic matter or cited in part or in whole without prior written permission of the WHO. No part of this document may be stored in a retrieval system or transmitted in any form by any means – electronic, mechanical or other – without the prior written permission of the WHO. The WHOQOL Group, Department of Mental Health, WHO, CH-1211, Geneva 27, Switzerland.

Permission to use the UK instrument must be obtained from Professor Suzanne Skevington, WHO Centre for the Study of Quality of Life, University of Bath, Bath, BA2 7AY, UK.

### The UK WHOQOL-Bref

#### Instructions

**Please read this carefully**

This questionnaire asks how you feel about your quality of life, health and other areas of your life. Please answer all the questions. If you are unsure about which response to give to a question, please choose the best one you can. There are no right or wrong answers. Your answer will be kept strictly confidential. Please keep in mind your standards, hopes, pleasures and concerns. We ask that you think about your life in the **last two weeks**.

For example, thinking about the **last two weeks**, a question might ask:

How much do you worry about your health?

| Not at all | Not much | A moderate amount | Very much | An extreme amount |
|---|---|---|---|---|
| 1 | 2 | 3 | 4 | 5 |

You should circle the number that best fits how much you have worries about your health over the last two weeks. So you would circle the number 4 if you worried about your health “very much”, or circle number 1 if you have worried “not at all” about your health. Please read each question, assess your feelings, and circle the number on the scale for each question that gives the best answer for you.

**Thank you for your help, please turn over page**

The following questions ask about **how much** you have experienced certain things in the last two weeks, for example, positive feelings such as happiness or contentment. If you have experienced these things an extreme amount, circle the number next to "An extreme amount". If you have not experienced these things at all, circle the number next to "Not at all". You should circle one of the numbers in between if you wish to show that your answer lies somewhere between "Not at all" and "Extremely". **Questions refer to the last two weeks**.

1. **How much do you feel that pain prevents you from doing what you need to do?** (F1.4)

| Not at all | Not much | A moderate amount | Very much | An extreme amount |
|---|---|---|---|---|
| 1 | 2 | 3 | 4 | 5 |

2. **How much do you enjoy life?** (F4.1)

| Not at all | Not much | A moderate amount | Very much | An extreme amount |
|---|---|---|---|---|
| 1 | 2 | 3 | 4 | 5 |

3. **How well are you able to concentrate?** (F5.3)

| Not at all | Not much | Moderately | Very well | Extremely |
|---|---|---|---|---|
| 1 | 2 | 3 | 4 | 5 |

4. **How much do you need medical treatment to function in your daily life?** (F11.3)

| Not at all | Not much | A moderate amount | Very much | An extreme amount |
|---|---|---|---|---|
| 1 | 2 | 3 | 4 | 5 |

5. **How safe do you feel in your daily life?** (F16.1)

| Not at all | Not much | Moderately | Very much | Extremely |
|---|---|---|---|---|
| 1 | 2 | 3 | 4 | 5 |

6. **How healthy is your physical environment?** (F22.1)

| Not at all | Not much | Moderately | Very much | Extremely |
|---|---|---|---|---|
| 1 | 2 | 3 | 4 | 5 |

The following questions ask about **how completely** you experienced, or were able to do certain things in the last two weeks, for example activities of daily living like washing, dressing or eating. If you have been able to do these things completely, circle the number next to "Completely". If you have not been able to do these things at all, circle the number next to "Not at all". You should circle one of the numbers in between if you wish to show that your answer lies somewhere between "Not at all" and "Completely". **Questions refer to the last two weeks**.

7. **Do you have enough energy for everyday life?** (F2.1)

| Not at all | Not much | Moderately | A great deal | Completely |
|---|---|---|---|---|
| 1 | 2 | 3 | 4 | 5 |

8. **How much are you able to accept your bodily appearance?** (F7.1)

| Not at all | Not much | Moderately | A great deal | Completely |
|---|---|---|---|---|
| 1 | 2 | 3 | 4 | 5 |

9. **To what extent do you have enough money to meet your needs?** (F18.1)

| Not at all | Not much | Moderately | A great deal | Completely |
|---|---|---|---|---|
| 1 | 2 | 3 | 4 | 5 |

10. **How available to you is the information that you need in your day-to-day life?** (F20.1)

| Not at all | Not much | Moderately | A great deal | Completely |
|---|---|---|---|---|
| 1 | 2 | 3 | 4 | 5 |

11. **To what extent do you have the opportunity for leisure activities?** (F21.1)

| Not at all | Not much | Moderately | A great deal | Completely |
|---|---|---|---|---|
| 1 | 2 | 3 | 4 | 5 |

The following questions ask you to say how **satisfied, happy or good** you have felt about various aspects of your life over the last two weeks, for example, about your family life or you energy level. Decide how satisfied or dissatisfied you are with each aspect of your life and then circle the number that best fits how you feel about this. **Questions refer to the last two weeks.**

12. **How satisfied are you with your health?** (G4)

| Very dissatisfied | Dissatisfied | Neither satisfied nor dissatisfied | Satisfied | Very satisfied |
|---|---|---|---|---|
| 1 | 2 | 3 | 4 | 5 |

13. **How satisfied are you with your sleep?** (F3.3)

| Very dissatisfied | Dissatisfied | Neither satisfied nor dissatisfied | Satisfied | Very satisfied |
|---|---|---|---|---|
| 1 | 2 | 3 | 4 | 5 |

14. **How satisfied are you with yourself?** (F6.3)

| Very dissatisfied | Dissatisfied | Neither satisfied nor dissatisfied | Satisfied | Very satisfied |
|---|---|---|---|---|
| 1 | 2 | 3 | 4 | 5 |

15. **How satisfied are you with your ability to perform daily living activities?** (F10.3)

| Very dissatisfied | Dissatisfied | Neither satisfied nor dissatisfied | Satisfied | Very satisfied |
|---|---|---|---|---|
| 1 | 2 | 3 | 4 | 5 |

16. **How satisfied are you with your personal relationships?** (F13.3)

| Very dissatisfied | Dissatisfied | Neither satisfied nor dissatisfied | Satisfied | Very satisfied |
|---|---|---|---|---|
| 1 | 2 | 3 | 4 | 5 |

17. **How satisfied are you with your sex life?** (F15.3)

| Very dissatisfied | Dissatisfied | Neither satisfied nor dissatisfied | Satisfied | Very satisfied |
|---|---|---|---|---|
| 1 | 2 | 3 | 4 | 5 |

18. **How satisfied are you with the support you get from your friends?** (F14.4)

| Very dissatisfied | Dissatisfied | Neither satisfied nor dissatisfied | Satisfied | Very satisfied |
|---|---|---|---|---|
| 1 | 2 | 3 | 4 | 5 |

19. **How satisfied are you with the conditions of your living place?** (F17.3)

| Very dissatisfied | Dissatisfied | Neither satisfied nor dissatisfied | Satisfied | Very satisfied |
|---|---|---|---|---|
| 1 | 2 | 3 | 4 | 5 |

20. **How satisfied are you with your access to health services?** (F19.3)

| Very dissatisfied | Dissatisfied | Neither satisfied nor dissatisfied | Satisfied | Very satisfied |
|---|---|---|---|---|
| 1 | 2 | 3 | 4 | 5 |

21 **How satisfied are you with your transport?** (F23.3)

| Very dissatisfied | Dissatisfied | Neither satisfied nor dissatisfied | Satisfied | Very satisfied |
|---|---|---|---|---|
| 1 | 2 | 3 | 4 | 5 |

22. **How would you rate your quality of life?** (G1)

| Very poor | Poor | Neither poor nor good | Good | Very good |
|---|---|---|---|---|
| 1 | 2 | 3 | 4 | 5 |

The following questions refer to **how often** you have felt or experienced certain things, for example the support of your family or friends, or negative experiences such as feeling unsafe. If you have not experienced these things at all in the last two weeks, circle the response "never". If you have experienced these things, decide how often and circle the appropriate number. So for example if you have experienced pain all the time in the last two weeks, circle the number next to "Always". **Questions refer to the last two weeks**.

23. **How often do you have negative feelings, such as blue mood, despair, anxiety, depression?** (F8.1)

| Never | Seldom | Quite often | Very often | Always |
|---|---|---|---|---|
| 1 | 2 | 3 | 4 | 5 |

The following questions refer to any **work** that you do. **Work here means any major activity that you do. This includes voluntary work, studying full-time, taking care of the home, taking care of children, paid work, or unpaid work. So work, as it is used here, means the activities you feel take up a major part of your time and energy. Questions refer to the last two weeks**.

24. **How satisfied are you with your capacity for work?** (F12.4)

| Very dissatisfied | Dissatisfied | Neither satisfied nor dissatisfied | Satisfied | Very satisfied |
|---|---|---|---|---|
| 1 | 2 | 3 | 4 | 5 |

The next few questions ask about **how well you were able to move around** in the last two weeks. This refers to your physical ability to move your body in such a way as to allow you to move about and do the things you would like to do, as well as the things that you need to do. **Questions refer to the last two weeks.**

25. **How well are you able to get around?** (F9.1)

| Very poor | Poor | Neither good nor poor | Good | Very good |
|---|---|---|---|---|
| 1 | 2 | 3 | 4 | 5 |

The following questions are concerned with **your personal beliefs** and how these affect your quality of life. These questions refer to religion, spirituality and any other personal beliefs you may hold. Once again these questions refer to the **last two weeks.**

26. **To what extent do you feel life to be meaningful?** (F24.2)

| Not at all | Not much | Moderately | Very much | Extremely |
|---|---|---|---|---|
| 1 | 2 | 3 | 4 | 5 |

**ABOUT YOU**

We would like you to answer a few general questions about yourself: by **circling** the correct answer or by **filling in the space provided**.

What is your gender? **MALE / FEMALE**

What is your date of birth? ____/____/____ (day / month / year)

What is the highest education you have received?
**None at all**
**Primary School**
**Secondary School**
**Further Education e.g. Technical/Clerical**
**University**

What is your marital status?
**Single** **Separated**
**Married** **Divorced**
**Living as married** **Widowed**

How is your health?

F9.1

| Very poor | Poor | Neither good nor poor | Good | Very good |
|---|---|---|---|---|
| 1 | 2 | 3 | 4 | 5 |

Are you currently ill? **YES / NO**

If something is wrong with your health, what do you think it is? Please write your illness(s) or problems here________________________________________________________

____________________________________________________________________

Are you currently in paid work? **YES / NO**

What is your occupation?__________________________________________________

**THANK YOU VERY MUCH FOR YOUR HELP**

### PATIENT HEALTH QUESTIONNAIRE-9 (PHQ-9)

| **Over the last 2 weeks, how often have you been bothered by any of the following problems?** *(Use "✔" to indicate your answer)* | **Not at all** | **Several days** | **More than half the days** | **Nearly every day** |
|---|---|---|---|---|
| **1.** Little interest or pleasure in doing things | 0 | 1 | 2 | 3 |
| **2.** Feeling down, depressed, or hopeless | 0 | 1 | 2 | 3 |
| **3.** Trouble falling or staying asleep, or sleeping too much | 0 | 1 | 2 | 3 |
| **4.** Feeling tired or having little energy | 0 | 1 | 2 | 3 |
| **5.** Poor appetite or overeating | 0 | 1 | 2 | 3 |
| **6.** Feeling bad about yourself — or that you are a failure or have let yourself or your family down | 0 | 1 | 2 | 3 |
| **7.** Trouble concentrating on things, such as reading the newspaper or watching television | 0 | 1 | 2 | 3 |
| **8.** Moving or speaking so slowly that other people could have noticed? Or the opposite — being so fidgety or restless that you have been moving around a lot more than usual | 0 | 1 | 2 | 3 |
| **9.** Thoughts that you would be better off dead or of hurting yourself in some way | 0 | 1 | 2 | 3 |

FOR OFFICE CODING ___*0*___ + ______ + ______ + ______

**=Total Score:** ______

**If you checked off any problems, how difficult have these problems made it for you to do your work, take care of things at home, or get along with other people?**

| **Not difficult at all** | **Somewhat difficult** | **Very difficult** | **Extremely difficult** |
|---|---|---|---|
| ☐ | ☐ | ☐ | ☐ |

Developed by Drs. Robert L. Spitzer, Janet B.W. Williams, Kurt Kroenke and colleagues, with an educational grant from Pfizer Inc. No permission required to reproduce, translate, display or distribute.

National Center for PTSD

POSTTRAUMATIC STRESS DISORDER

### PTSD Checklist for *DSM-5* (PCL-5)

**Version date:** 11 April 2018

**Reference:** Weathers, F. W., Litz, B. T., Keane, T. M., Palmieri, P. A., Marx, B. P., & Schnurr, P. P. (2013). *The PTSD Checklist for DSM-5 (PCL-5) – Standard* [Measurement instrument]. Available from https://www.ptsd.va.gov/

**URL:** https://www.ptsd.va.gov/professional/assessment/adult-sr/ptsd-checklist.asp

**Note:** This is a fillable form. You may complete it electronically.

**PCL-5**

**Instructions:** Below is a list of problems that people sometimes have in response to a very stressful experience. Please read each problem carefully and then circle one of the numbers to the right to indicate how much you have been bothered by that problem in the past month.

| In the past month, how much were you bothered by: | Not at all | A little bit | Moderately | Quite a bit | Extremely |
|---|---|---|---|---|---|
| 1. Repeated, disturbing, and unwanted memories of the stressful experience? | 0 | 1 | 2 | 3 | 4 |
| 2. Repeated, disturbing dreams of the stressful experience? | 0 | 1 | 2 | 3 | 4 |
| 3. Suddenly feeling or acting as if the stressful experience were actually happening again (as if you were actually back there reliving it)? | 0 | 1 | 2 | 3 | 4 |
| 4. Feeling very upset when something reminded you of the stressful experience? | 0 | 1 | 2 | 3 | 4 |
| 5. Having strong physical reactions when something reminded you of the stressful experience (for example, heart pounding, trouble breathing, sweating)? | 0 | 1 | 2 | 3 | 4 |
| 6. Avoiding memories, thoughts, or feelings related to the stressful experience? | 0 | 1 | 2 | 3 | 4 |
| 7. Avoiding external reminders of the stressful experience (for example, people, places, conversations, activities, objects, or situations)? | 0 | 1 | 2 | 3 | 4 |
| 8. Trouble remembering important parts of the stressful experience? | 0 | 1 | 2 | 3 | 4 |
| 9. Having strong negative beliefs about yourself, other people, or the world (for example, having thoughts such as: I am bad, there is something seriously wrong with me, no one can be trusted, the world is completely dangerous)? | 0 | 1 | 2 | 3 | 4 |
| 10. Blaming yourself or someone else for the stressful experience or what happened after it? | 0 | 1 | 2 | 3 | 4 |
| 11. Having strong negative feelings such as fear, horror, anger, guilt, or shame? | 0 | 1 | 2 | 3 | 4 |
| 12. Loss of interest in activities that you used to enjoy? | 0 | 1 | 2 | 3 | 4 |
| 13. Feeling distant or cut off from other people? | 0 | 1 | 2 | 3 | 4 |
| 14. Trouble experiencing positive feelings (for example, being unable to feel happiness or have loving feelings for people close to you)? | 0 | 1 | 2 | 3 | 4 |
| 15. Irritable behavior, angry outbursts, or acting aggressively? | 0 | 1 | 2 | 3 | 4 |
| 16. Taking too many risks or doing things that could cause you harm? | 0 | 1 | 2 | 3 | 4 |
| 17. Being "superalert" or watchful or on guard? | 0 | 1 | 2 | 3 | 4 |
| 18. Feeling jumpy or easily startled? | 0 | 1 | 2 | 3 | 4 |
| 19. Having difficulty concentrating? | 0 | 1 | 2 | 3 | 4 |
| 20. Trouble falling or staying asleep? | 0 | 1 | 2 | 3 | 4 |

#### Family Structure Optional Questionnaire

HEAD.FAM1 The following section asks a little bit about your **biological parents.**

FAM.MOT.1.0 What is your mother's country of origin?

▼ Afghanistan (1) ... Zimbabwe (1357)

---

FAM.MOT.2.0 Which of the following **qualifications** does your mother have? (Please select all that apply)

- ▢ College or university degree (1)
- ▢ A levels/AS levels or equivalent (2)
- ▢ O levels/GCSEs or equivalent (3)
- ▢ CSEs or equivalent (4)
- ▢ NVQ or HND or HNC or equivalent (5)
- ▢ Other professional qualifications (e.g. nursing, teaching) (6)
- ▢ ⊗None of the above (0)
- ▢ ⊗Don't Know (-88)
- ▢ ⊗Prefer not to answer (-99)

FAM.MOT.3.0 Is your biological mother still **alive**?

- ◯ Yes (1)
- ◯ No (0)
- ◯ Don't know (-88)
- ◯ Prefer not to answer (-99)

*Display This Question:*

*If Is your biological mother still alive? = Yes*

FAM.MOT.4age.0 **How old** is your biological mother **now**?

________________________________________________________________

*Display This Question:*

*If Is your biological mother still alive? = No*

FAM.MOT.4pass.0 **How old** was your biological mother when she passed away?

________________________________________________________________

*Display This Question:*

*If Is your biological mother still alive? = No*

FAM.MOT.4yr.0 In what **year** did she pass away?

________________________________________________________________

*Display This Question:*

*If Is your biological mother still alive? = No*

FAM.MOT.4cau.0 What was her **cause** of death?

________________________________________________________________

FAM.FAT.1.0 What is your father's country of origin?

▼ Afghanistan (1) ... Zimbabwe (1357)

---

FAM.FAT.2.0 Which of the following **qualifications** does your father have? (Please select all that apply)

- ▢ College or university degree (1)
- ▢ A levels/AS levels or equivalent (2)
- ▢ O levels/GCSEs or equivalent (3)
- ▢ CSEs or equivalent (4)
- ▢ NVQ or HND or HNC or equivalent (5)
- ▢ Other professional qualifications (e.g. nursing, teaching) (6)
- ▢ ⊗None of the above (0)
- ▢ ⊗Don't Know (-88)
- ▢ ⊗Prefer not to answer (-99)

FAM.FAT.3.0 Is your biological father still **alive?**

- ○ Yes (1)
- ○ No (0)
- ○ Don't know (-88)
- ○ Prefer not to answer (-99)

*Display This Question:*

*If Is your biological father still alive? = Yes*

FAM.FAT.4age.0 **How old** is your biological father **now?**

____________________________________________________________

*Display This Question:*

*If Is your biological father still alive? = No*

FAM.FAT.4pass.0 **How old** was your biological father when he passed away?

________________________________________________________________

*Display This Question:*

*If Is your biological father still alive? = No*

FAM.FAT.4yr.0 In what **year** did he pass away?

________________________________________________________________

*Display This Question:*

*If Is your biological father still alive? = No*

FAM.FAT.4cau.0 What was his **cause** of death?

________________________________________________________________

HEAD.SIB This section refers to any **siblings** or **children** which you have.

FAM.ADO.1.0 Are you **adopted?**

- ○ Yes (1)
- ○ No (0)
- ○ Don't know (-88)
- ○ Prefer not to answer (-99)

FAM.SIB.1.0 Do you have any **siblings?**

*(Including full, half, step and adopted siblings, including any who have passed away)*

- ◯ Yes (1)
- ◯ No (0)
- ◯ Don't know (-88)
- ◯ Prefer not to answer (-99)

*Skip To: HEAD.MH1 If Do you have any siblings? (Including full, half, step and adopted siblings, including any who hav... = No*

*Skip To: HEAD.MH1 If Do you have any siblings? (Including full, half, step and adopted siblings, including any who hav... = No*

*Skip To: HEAD.MH1 If Do you have any siblings? (Including full, half, step and adopted siblings, including any who hav... = No*

FAM.SIB.2.0 How **many** siblings do you have?

▼ 1 (1) ... 7+ (7)

FAM.BRO.0.0 How many brothers do you have?

*(Please include full, half, step and adopted brothers, including any who have passed away)*

________________________________________________________________

*Display This Question:*

*If If How many brothers do you have? (Please include full, half, step and adopted brothers, including any who have passed away) Text Response Is Greater Than or Equal to 1*

HEAD.FAM.BRO For each of your **brothers**, please list their **relationship to you** *(e.g. full, half, step, adopted)* and their age.

*(If they are deceased, please enter their age at death)*

| | Age | Relationship |
|---|---|---|

| | (1) | |
|---|---|---|
| Brother 1 (FAM.BRO.1.0) | | ▼ Full (1 ... Adopted (4) |
| Brother 2 (FAM.BRO.2.0) | | ▼ Full (1 ... Adopted (4) |
| Brother 3 (FAM.BRO.3.0) | | ▼ Full (1 ... Adopted (4) |
| Brother 4 (FAM.BRO.4.0) | | ▼ Full (1 ... Adopted (4) |
| Brother 5 (FAM.BRO.5.0) | | ▼ Full (1 ... Adopted (4) |
| Brother 6 (FAM.BRO.6.0) | | ▼ Full (1 ... Adopted (4) |
| Brother 7 (FAM.BRO.7.0) | | ▼ Full (1 ... Adopted (4) |

*Display This Question:*

*If If How many brothers do you have? (Please include full, half, step and adopted brothers, including any who have passed away) Text Response Is Greater Than or Equal to 7*

FAM.BRO.8.0 Do you have any more brothers?

○ Yes (1)

○ No (0)

*Display This Question:*

*If Do you have any more brothers? = Yes*

FAM.BRO.8.0.TEXT For each of your other **brothers,** please list their **current age** (or age of death) and their relationship to you in the text box below.

________________________________________________________________

________________________________________________________________

________________________________________________________________

________________________________________________________________

________________________________________________________________

FAM.SIS.0.0 How many **sisters** do you have?

*(Please include full, half, step and adopted sisters, including any who have passed away)*

________________________________________________________________

*Skip To: HEAD.MH1 If Condition: How many sisters do you hav... Is Equal to 0. Skip To: The following section asks about any ....*

*Display This Question:*

*If If How many sisters do you have? (Please include full, half, step and adopted sisters, including... Text Response Is Greater Than or Equal to 1*

HEAD.FAM.SIS For each of your **sisters**, please list their **relationship to you** *(e.g. full, half, step, adopted)* and their age.

*(If they are deceased, please enter their age at death)*

| | Age | Relationship |
|---|---|---|
| | (1) | |

| | | |
|---|---|---|
| Sister 1 (FAM.SIS.1.0) | | ▼ Full (1 ... Adopted (4) |
| Sister 2 (FAM.SIS.2.0) | | ▼ Full (1 ... Adopted (4) |
| Sister 3 (FAM.SIS.3.0) | | ▼ Full (1 ... Adopted (4) |
| Sister 4 (FAM.SIS.4.0) | | ▼ Full (1 ... Adopted (4) |
| Sister 5 (FAM.SIS.5.0) | | ▼ Full (1 ... Adopted (4) |
| Sister 6 (FAM.SIS.6.0) | | ▼ Full (1 ... Adopted (4) |
| Sister 7 (FAM.SIS.7.0) | | ▼ Full (1 ... Adopted (4) |

*Display This Question:*

*If If How many sisters do you have? (Please include full, half, step and adopted sisters, including... Text Response Is Greater Than or Equal to 7*

FAM.SIS.8.0 Do you have any more sisters?

◯ Yes (1)

◯ No (0)

*Display This Question:*

*If Do you have any more sisters? = Yes*

FAM.SIS.8.0.TEXT For each of your other **sisters**, please list their **current age** (or age of death) and their **relationship to you** in the text box below.

________________________________________________________________

________________________________________________________________

________________________________________________________________

________________________________________________________________

________________________________________________________________

HEAD.MH1

The following section asks about any **illnesses** that your **immediate family** may have been diagnosed with throughout their lifetime.

HEAD.FMH1 Thinking about your **immediate** family *(parents, brothers, sisters or children)*, have they ever been diagnosed with the following mental health disorders?

Please tick the appropriate boxes.

| | Yes (1) | No (0) | Don't know (-88) |
|---|---|---|---|
| Depression (FAM.MH.1.0) | ○ | ○ | ○ |
| Depression during or after pregnancy *(antenatal/postnatal depression)* (FAM.MH.2.0) | ○ | ○ | ○ |
| Mania, hypomania bipolar or manic depression (FAM.MH.3.0) | ○ | ○ | ○ |
| Anxiety, nerves or generalised anxiety disorder (FAM.MH.4.0) | ○ | ○ | ○ |
| Social anxiety or social phobia (FAM.MH.5.0) | ○ | ○ | ○ |
| Specific phobia *(e.g. phobia of flying)* (FAM.MH.6.0) | ○ | ○ | ○ |
| Agoraphobia (FAM.MH.7.0) | ○ | ○ | ○ |
| Panic attacks (FAM.MH.8.0) | ○ | ○ | ○ |
| Post-traumatic stress disorder (PTSD) (FAM.MH.9.0) | ○ | ○ | ○ |
| Obsessive compulsive disorder (OCD) (FAM.MH.10.0) | ○ | ○ | ○ |
| Body dysmorphic disorder (BDD) (FAM.MH.11.0) | ○ | ○ | ○ |
| Other obsessive-compulsive related disorder *(e.g. skin-picking)* (FAM.MH.12.0) | ○ | ○ | ○ |

FAM.MH.1num.0 How **many** of your immediate family **(parents, brothers, sisters or children)**, have ever been diagnosed with **${lm://Field/1}?**

________________________________________________________________

HEAD.FMH2 Thinking about your **immediate** family *(parents, brothers, sisters or children*), have they ever been diagnosed with the following mental health disorders?

Please tick the appropriate boxes.

| | Yes (1) | No (0) | Don't know (-88) |
|---|---|---|---|
| Anorexia nervosa (13_FAM.MH.2.0) | ○ | ○ | ○ |
| Atypical anorexia nervosa (14_FAM.MH.2.0) | ○ | ○ | ○ |
| Bulimia nervosa (15_FAM.MH.2.0) | ○ | ○ | ○ |
| Psychological over-eating or binge-eating (16_FAM.MH.2.0) | ○ | ○ | ○ |
| Schizophrenia (17_FAM.MH.2.0) | ○ | ○ | ○ |
| Any other type of psychosis or psychotic illness (18_FAM.MH.2.0) | ○ | ○ | ○ |
| Personality disorder (19_FAM.MH.2.0) | ○ | ○ | ○ |
| Autism, asperger's or autistic spectrum disorder (20_FAM.MH.2.0) | ○ | ○ | ○ |
| Attention deficit or attention deficit and hyperactivity disorder (ADD/ADHD) (21_FAM.MH.2.0) | ○ | ○ | ○ |
| Other *(Please tell us more)* (22_FAM.MH.2.0) | ○ | ○ | ○ |

*Display This Question:*

*If Thinking about your immediate family (parents, brothers, sisters or children), have they ever bee... = Personality disorder [ Yes ]*

FAM.PD.1.0 **Which** personality disorder(s) has a member of your **immediate** family been diagnosed with?

*(Please select all that apply)*

- ☐ Paranoid (1)
- ☐ Schizoid (2)
- ☐ Schizotypal (3)
- ☐ Antisocial (4)
- ☐ Borderline (5)
- ☐ Histrionic (6)
- ☐ Narcissistic (7)
- ☐ Avoidant/anxious (8)
- ☐ Dependent (9)
- ☐ Obsessive-compulsive (10)
- ☐ ⊗Don't know (-88)
- ☐ ⊗Prefer not to answer (-99)

FAM.MH.2num.0 How many of your immediate family (parents, brothers, sisters or children), have ever been diagnosed with ${lm://Field/1}?

________________________________________________________________

HEAD.FNS1 Thinking about your **immediate** family *(parents, brothers, sisters or children*), have they ever been diagnosed with the following illnesses?

Please tick the appropriate boxes.

| | Yes (1) | No (0) | Don't know (-88) |
|---|---|---|---|
| Epilepsy or convulsions (FAM.NS.1.0) | ○ | ○ | ○ |
| Migraines (FAM.NS.2.0) | ○ | ○ | ○ |
| Multiple sclerosis (FAM.NS.3.0) | ○ | ○ | ○ |
| Parkinson's disease (FAM.NS.4.0) | ○ | ○ | ○ |
| Severe memory loss (like Alzheimer's) (FAM.NS.5.0) | ○ | ○ | ○ |

FAM.NS.1num.0 How many of your immediate family (parents, brothers, sisters or children), have ever been diagnosed with ${lm://Field/1}?

________________________________________________________________

HEAD.FALL1 Thinking about your **immediate** family *(parents, brothers, sisters or children)*, have they ever been diagnosed with the following illnesses?

Please tick the appropriate boxes.

| | Yes (1) | No (0) | Don't know (-88) |
|---|---|---|---|
| Hay fever (FAM.ALL.1.0) | ○ | ○ | ○ |
| Drug allergy *(if yes, which drug?)* (FAM.ALL.2.0) | ○ | ○ | ○ |
| Food allergy *(if yes, which food?)* (FAM.ALL.3.0) | ○ | ○ | ○ |
| Any other allergy *(if yes, which allergy?)* (FAM.ALL.4.0) | ○ | ○ | ○ |

FAM.ALL.1num.0 How many of your immediate family (parents, brothers, sisters or children), have ever been diagnosed with ${lm://Field/1}?

________________________________________________________________

HEAD.FBON1 Thinking about your **immediate** family *(parents, brothers, sisters or children)*, have they ever been diagnosed with the following illnesses?

Please tick the appropriate boxes.

| | Yes (1) | No (0) | Don't Know (-88) |
|---|---|---|---|
| Osteoporosis or "thin" bones (FAM.BON.1.0) | ○ | ○ | ○ |
| Osteoarthritis (FAM.BON.2.0) | ○ | ○ | ○ |
| Rheumatoid arthritis (FAM.BON.3.0) | ○ | ○ | ○ |
| Other arthritis (FAM.BON.4.0) | ○ | ○ | ○ |

FAM.BON.1num.0 How many of your immediate family (parents, brothers, sisters or children), have ever been diagnosed with ${lm://Field/1}?

________________________________________________________________

HEAD.FRES1 Thinking about your **immediate** family *(parents, brothers, sisters or children)*, have they ever been diagnosed with the following illnesses?

Please tick the appropriate boxes.

| | Yes (1) | No (0) | Don't know (-88) |
|---|---|---|---|
| Asthma (FAM.LUN.1.0) | ○ | ○ | ○ |
| Emphysema or chronic bronchitis (FAM.LUN.2.0) | ○ | ○ | ○ |
| Heart attack or angina (FAM.HRT.1.0) | ○ | ○ | ○ |
| High blood cholesterol (FAM.HRT.2.0) | ○ | ○ | ○ |
| High blood pressure (FAM.HRT.3.0) | ○ | ○ | ○ |
| Atrial fibrillation (FAM.HRT.4.0) | ○ | ○ | ○ |
| Stroke (FAM.HRT.5.0) | ○ | ○ | ○ |

FAM.LHT.1.num.0 How **many** of your immediate family **(parents, brothers, sisters or children)**, have ever been diagnosed with **${lm://Field/1}?**

________________________________________________________________

HEAD.FDIG1 Thinking about your **immediate** family *(parents, brothers, sisters or children)*, have they ever been diagnosed with the following illnesses?

Please tick the appropriate boxes.

| | Yes (1) | No (0) | Don't know (-88) |
|---|---|---|---|
| Crohn's disease (FAM.DIG.1.0) | ◯ | ◯ | ◯ |
| Ulcerative colitis (FAM.DIG.2.0) | ◯ | ◯ | ◯ |
| Coeliac disease (FAM.DIG.3.0) | ◯ | ◯ | ◯ |
| Pain due to diabetes (diabetic neuropathy) (FAM.DIA.3.0) | ◯ | ◯ | ◯ |
| Polycystic ovary syndrome (PCOS) (FAM.PCOS.1.0) | ◯ | ◯ | ◯ |
| Pain due to virus (post herpetic neuralgia) (FAM.DIA.4.0) | ◯ | ◯ | ◯ |

FAM.DIG/DIA.1num.0 How many of your immediate family (parents, brothers, sisters or children), have ever been diagnosed with ${lm://Field/1}?

________________________________________________________________

HEAD.FOTH1 Thinking about your **immediate** family *(parents, brothers, sisters or children)*, have they ever been diagnosed with the following illnesses?

Please tick the appropriate boxes.

| | Yes (1) | No (0) | Don't know (-88) |
|---|---|---|---|
| Psoriasis (FAM.SKN.1.0) | ○ | ○ | ○ |
| Vitiligo (FAM.SKN.2.0) | ○ | ○ | ○ |
| Eczema (FAM.SKN.3.0) | ○ | ○ | ○ |
| Thyroid disease *(if yes, please specify) (FAM.THY.1.0)* | ○ | ○ | ○ |
| Any other illnesses not listed? *(Please specify) (FAM.OTH.1.0)* | ○ | ○ | ○ |

FAM.OTH.1num.0 How many of your immediate family (parents, brothers, sisters or children), have ever been diagnosed with ${lm://Field/1}?

________________________________________________________________

### Alcohol use disorders identification test (AUDIT)

AUDIT is a comprehensive 10 question alcohol harm screening tool. It was developed by the World Health Organisation (WHO) and modified for use in the UK and has been used in a variety of health and social care settings.

| Questions | Scoring system | | | | | Your score |
|---|---|---|---|---|---|---|
| | 0 | 1 | 2 | 3 | 4 | |
| How often do you have a drink containing alcohol? | Never | Monthly or less | 2 to 4 times per month | 2 to 3 times per week | 4 times or more per week | |
| How many units of alcohol do you drink on a typical day when you are drinking? | 0 to 2 | 3 to 4 | 5 to 6 | 7 to 9 | 10 or more | |
| How often have you had 6 or more units if female, or 8 or more if male, on a single occasion in the last year? | Never | Less than monthly | Monthl y | Weekly | Daily or almost daily | |
| How often during the last year have you found that you were not able to stop drinking once you had started? | Never | Less than monthly | Monthl y | Weekly | Daily or almost daily | |
| How often during the last year have you failed to do what was normally expected from you because of your drinking? | Never | Less than monthly | Monthl y | Weekly | Daily or almost daily | |
| How often during the last year have you needed an alcoholic drink in the morning to get yourself going after a heavy drinking session? | Never | Less than monthly | Monthl y | Weekly | Daily or almost daily | |
| How often during the last year have you had a feeling of guilt or remorse after drinking? | Never | Less than monthly | Monthl y | Weekly | Daily or almost daily | |
| How often during the last year have you been unable to remember what happened the night before because you had been drinking? | Never | Less than monthly | Monthl y | Weekly | Daily or almost daily | |
| Have you or somebody else been injured as a result of your drinking? | No | | Yes, but not in the last year | | Yes, during the last year | |
| Has a relative or friend, doctor or other health worker been concerned about your drinking or suggested that you cut down? | No | | Yes, but not in the last year | | Yes, during the last year | |

| **Total AUDIT score** |
|---|

#### Scoring:

- 0 to 7 indicates low risk
- 8 to 15 indicates increasing risk
- 16 to 19 indicates higher risk,
- 20 or more indicates possible dependence

### Giving feedback and advice

If the score is lower

If the score is 8 or above, give brief advice to reduce risk for alcohol harm. If the score is 20 or above, consider referral to specialist alcohol harm assessment.

### Alcohol unit reference

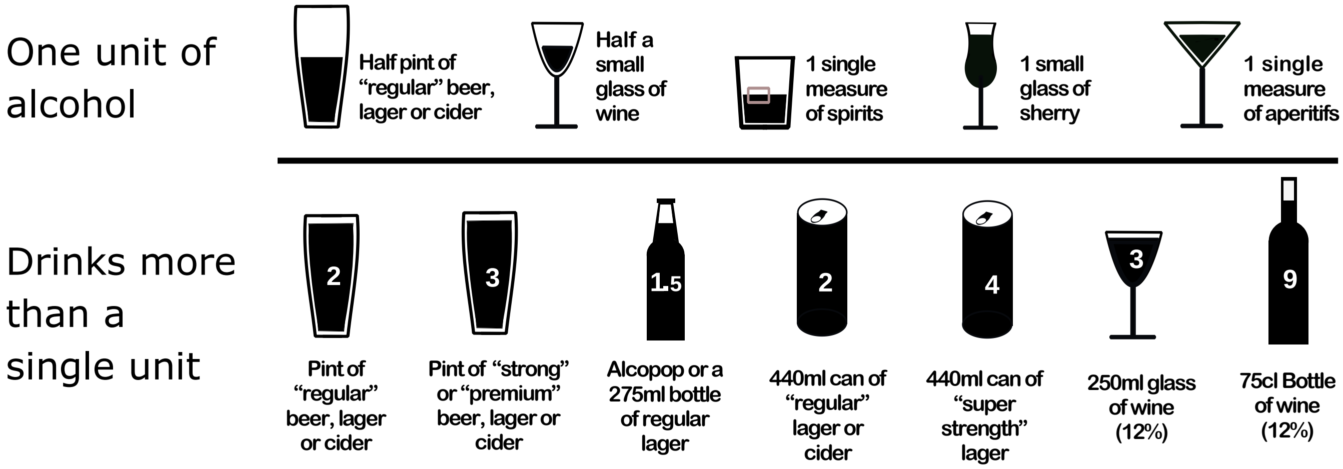

Id. nr. ☐☐☐☐☐☐

### DUDIT

Drug Use Disorders Identification Test

**Here are a few questions about drugs.** Please answer as correctly and honestly as possible by indicating which answer is right for you.

☐ **Man** ☐ **Woman** **Age** ☐☐

| Question | | | | | |
|---|---|---|---|---|---|
| 1. How often do you use drugs other than alcohol? (See list of drugs on back side.) | Never ☐ | Once a month or less often ☐ | 2-4 times a month ☐ | 2-3 times a week ☐ | 4 times a week or more often ☐ |
| 2. Do you use more than one type of drug on the same occasion? | Never ☐ | Once a month or less often ☐ | 2-4 times a month ☐ | 2-3 times a week ☐ | 4 times a week or more often ☐ |
| 3. How many times do you take drugs on a typical day when you use drugs? | 0 ☐ | 1-2 ☐ | 3-4 ☐ | 5-6 ☐ | 7 or more ☐ |
| 4. How often are you influenced heavily by drugs? | Never ☐ | Less often than once a month ☐ | Every month ☐ | Every week ☐ | Daily or almost every day ☐ |
| 5. Over the past year, have you felt that your longing for drugs was so strong that you could not resist it? | Never ☐ | Less often than once a month ☐ | Every month ☐ | Every week ☐ | Daily or almost every day ☐ |
| 6. Has it happened, over the past year, that you have not been able to stop taking drugs once you started? | Never ☐ | Less often than once a month ☐ | Every month ☐ | Every week ☐ | Daily or almost every day ☐ |
| 7. How often over the past year have you taken drugs and then neglected to do something you should have done? | Never ☐ | Less often than once a month ☐ | Every month ☐ | Every week ☐ | Daily or almost every day ☐ |
| 8. How often over the past year have you needed to take a drug the morning after heavy drug use the day before? | Never ☐ | Less often than once a month ☐ | Every month ☐ | Every week ☐ | Daily or almost every day ☐ |
| 9. How often over the past year have you had guilt feelings or a bad conscience because you used drugs? | Never ☐ | Less often than once a month ☐ | Every month ☐ | Every week ☐ | Daily or almost every day ☐ |
| 10. Have you or anyone else been hurt (mentally or physically) because you used drugs? | No ☐ | Yes, but not over the past year ☐ | Yes, over the past year ☐ | | |
| 11. Has a relative or a friend, a doctor or a nurse, or anyone else, been worried about your drug use or said to you that you should stop using drugs? | No ☐ | Yes, but not over the past year ☐ | Yes, over the past year ☐ | | |

© 2002 Anne H. Berman, Hans Bergman, Tom Palmstierna & Frans Schlyter Europé English version 1

**Turn the page to see the list of drugs**

### LIST OF DRUGS

**(Note! Not alcohol!)**

| Cannabis | Amphetamines | Cocaine | Opiates | Hallucinogens | Solvents/inhalants | GHB and others |
|---|---|---|---|---|---|---|
| Marijuana<br>Hash<br>Hash oil | Methamphetamine<br>Phenmetraline<br>Khat<br>Betel nut<br>Ritaline<br>(Methylphenidate) | Crack<br>Freebase<br>Coca leaves | Smoked heroin<br>Heroin<br>Opium | Ecstasy<br>LSD (Lisergic acid)<br>Mescaline<br>Peyote<br>PCP, angel dust<br>(Phencyclidine)<br>Psilocybin<br>DMT<br>(Dimethyltryptamine) | Thinner<br>Trichlorethylene<br>Gasoline/petrol<br>Gas<br>Solution<br>Glue | GHB<br>Anabolic steroids<br>Laughing gas<br>(Halothane)<br>Amyl nitrate<br>(Poppers)<br>Anticholinergic compounds |

### PILLS – MEDICINES

Pills count as drugs when you take

- more of them or take them more often than the doctor has prescribed for you
- pills because you want to have fun, feel good, get "high", or wonder what sort of effect they have on you
- pills that you have received from a relative or a friend
- pills that you have bought on the "black market" or stolen

**SLEEPING PILLS/SEDATIVES**

| | | |
|---|---|---|
| Alprazolam | Glutethimide | Rohypnol |
| Amobarbital | Halcion | Secobarbital |
| Apodorm | Heminevrin | Sobril |
| Apozepam | Iktorivil | Sonata |
| Aprobarbital | Imovane | Stesolid |
| Butabarbital | Mephobarbital | Stilnoct |
| Butalbital | Meprobamate | Talbutal |
| Chloral hydrate | Methaqualone | Temesta |
| Diazepam | Methohexital | Thiamyal |
| Dormicum | Mogadon | Thiopental |
| Ethcholorvynol | Nitrazepam | Triazolam |
| Fenemal | Oxascand | Xanor |
| Flunitrazepam | Pentobarbital | Zopiklon |
| Fluscand | Phenobarbital | |

**PAINKILLERS**

| | | |
|---|---|---|
| Actiq | Durogesic | OxyNorm |
| Coccilana-Etyfin | Fentanyl | Panocod |
| Citodon | Ketodur | Panocod forte |
| Citodon forte | Ketogan | Paraflex comp |
| Dexodon | Kodein | Somadril |
| Depolan | Maxidon | Spasmofen |
| Dexofen | Metadon | Subutex |
| Dilaudid | Morfin | Temgesic |
| Distalgesic | Nobligan | Tiparol |
| Dolcontin | Norflex | Tradolan |
| Doleron | Norgesic | Tramadul |
| Dolotard | Opidol | Treo comp |
| Doloxene | OxyContin | |

Pills do NOT count as drugs if they have been prescribed by a doctor and you take them in the prescribed dosage.

© 2002 Anne H. Berman, Hans Bergman, Tom Palmstierna & Frans Schlyter Europé English version 1

Drugs and Addiction Optional Questionnaire

SUB.1.0 Do you **currently** take any substances *(for example, cannabis, cocaine, speed, amphetamines, ecstasy, khat, LSD, heroin or methadone)* or legal "highs"?

- ○ Yes (1)
- ○ No (0)
- ○ Prefer not to answer (-99)

*Display This Question:*

*If Do you currently take any substances? = Yes*

SUB.1txt.0 Please give the **name** of the substance:

- ☐ Amphetamine (e.g. Speed) (4)
- ☐ Cocaine (5)
- ☐ Heroin (6)
- ☐ Ketamine (7)
- ☐ Khat (8)
- ☐ MDMA (e.g. Ecstasy) (9)
- ☐ Mephedrone (10)
- ☐ Solvents (e.g. glue, aerosols) (11)
- ☐ Other (12) ________________________________________________
- ☐ None of the above (13)
- ☐ Prefer not to answer (14)

*Display This Question:*

*If Do you currently take any substances? = Yes*

SUB.2.0 How **often** do you take it?

- ◯ Monthly or less (1)
- ◯ 2 to 4 times a month (2)
- ◯ 2 to 3 times a week (3)
- ◯ 4 or more times a week (4)
- ◯ Prefer not to answer (-99)

HEAD.DRUG1 This questionnaire asks about your past and/or current experiences with **drugs, addictions,** and **dependence**.

**Your answers will remain confidential, so please be honest.**

Click next below to continue to the questionnaire.

DRUG.1.0 Have you **ever** been **addicted** to or **dependent** on something? *(not including cigarettes or caffeine)*

- ◯ Yes (1)
- ◯ No (0)
- ◯ Don't know (-88)
- ◯ Prefer not to answer (-99)

*Skip To: DRUG.CAN.1.0 If Have you ever been addicted to or dependent on something? (not including cigarettes or caffeine) = No*

*Skip To: DRUG.CAN.1.0 If Have you ever been addicted to or dependent on something? (not including cigarettes or caffeine) = Don't know*

*Skip To: DRUG.CAN.1.0 If Have you ever been addicted to or dependent on something? (not including cigarettes or caffeine) = Prefer not to answer*

DRUG.2.0 Have you **ever** been **addicted to alcohol?**

- ◯ Yes (1)
- ◯ No (0)
- ◯ Don't know (-88)
- ◯ Prefer not to answer (-99)

---

*Display This Question:*

*If Have you ever been addicted to alcohol? = Yes*

DRUG.3.0 Is this addiction **ongoing?**

- ◯ Yes (1)
- ◯ No (0)
- ◯ Prefer not to answer (-99)

*Display This Question:*

*If Have you ever been addicted to alcohol? = Yes*

DRUG.4.0

Have you been **physically dependent** on **alcohol**? For example, have you experienced sweating, shaking, or nausea, which stop once you drink alcohol?

- ◯ Yes (1)
- ◯ No (0)
- ◯ Don't know (-88)
- ◯ Prefer not to answer (-99)

DRUG.MED.1.0 Have you ever been **addicted** to or **dependent** on **prescription** or over-the-counter **medication**?

- Yes (1)
- No (0)
- Don't know (-88)
- Prefer not to answer (-99)

---

*Display This Question:*

*If Have you ever been addicted to or dependent on prescription or over-the-counter medication? = Yes*

DRUG.DEP.1.0 Was this **addiction or dependence** to one of the following? *(Please select all that apply)*

- A sedative, benzodiazepine or sleeping tablet (1)
- A painkiller (2)
- Something else (3)
- ⊗Don't know (-88)
- ⊗Prefer not to answer (-99)

*Display This Question:*

*If Was this addiction or dependence to one of the following? (Please select all that apply) = A sedative, benzodiazepine or sleeping tablet*

DRUG.DEP.2.0 Which of the following was this **addiction or dependence** to? *(Please select all that apply)*

- ☐ Sedative (1)
- ☐ Benzodiazepine (2)
- ☐ Sleeping tablet (3)
- ☐ ⊗Prefer not to answer (-99)

---

*Display This Question:*

*If Was this addiction or dependence to one of the following? (Please select all that apply) = A sedative, benzodiazepine or sleeping tablet*

DRUG.DEP.3.0 Is this addiction or dependence **ongoing?**

- ○ Yes (1)
- ○ No (0)
- ○ Prefer not to answer (-99)

*Display This Question:*

*If Was this addiction or dependence to one of the following? (Please select all that apply) = A painkiller*

DRUG.PAI.2.0 Was this **addiction or dependence** to an **opioid** painkiller? *(e.g. oxycodone, codeine)*

- ○ Yes (1)
- ○ No (0)
- ○ Prefer not to answer (-99)

---

*Display This Question:*

*If Was this addiction or dependence to one of the following? (Please select all that apply) = A painkiller*

DRUG.PAI.3.0 Is this addiction or dependence **ongoing?**

○ Yes (1)

○ No (0)

○ Prefer not to answer (-99)

*Display This Question:*

*If Was this addiction or dependence to one of the following? (Please select all that apply) = Something else*

DRUG.OTH.1txt.0 Which **other** prescription or over-the-counter medication **not previously mentioned** was this addiction or dependence to?

________________________________________________________________

*Display This Question:*

*If Was this addiction or dependence to one of the following? (Please select all that apply) = Something else*

DRUG.OTH.2.0 Is this addiction or dependence **ongoing?**

○ Yes (1)

○ No (0)

○ Prefer not to answer (-99)

DRUG.ILL.1.0 Have you **ever** been addicted to **illicit** recreational drugs?

○ Yes (1)

○ No (0)

○ Don't know (-88)

○ Prefer not to answer (-99)

*Display This Question:*

*If Have you ever been addicted to illicit recreational drugs? = Yes*

DRUG.ILL.2.0 What **type** of illicit drug(s) have you **ever** been addicted to? *(Please select all that apply)*

- ▢ Amphetamine (e.g. Speed) (1)
- ▢ Cocaine (2)
- ▢ Heroin (4)
- ▢ Ketamine (5)
- ▢ Khat (6)
- ▢ MDMA (e.g. Ecstasy) (3)
- ▢ Mephedrone (7)
- ▢ Solvents (e.g. glue, aerosols) (9)
- ▢ Other (10)
- ▢ ⊗None of the above (0)
- ▢ ⊗Prefer not to answer (-99)

---

*Display This Question:*

*If What type of illicit drug(s) have you ever been addicted to? (Please select all that apply) = Other*

DRUG.ILL.2txt.0 Please specify what **other drug** you have been addicted to:

________________________________________________________________

*Display This Question:*

*If Have you ever been addicted to illicit recreational drugs? = Yes*

DRUG.ILL.3.0 Is this addiction or dependence **ongoing?**

*(Please select **yes** if the addiction is ongoing for **any** of the drugs selected in the previous question)*

- ◯ Yes (1)
- ◯ No (0)
- ◯ Prefer not to answer (-99)

DRUG.BEH.1.0 Have you **ever** been addicted to a behaviour *(such as gambling)?*

- ◯ Yes (1)
- ◯ No (0)
- ◯ Don't know (-88)
- ◯ ⊗ Prefer not to answer (-99)

---

*Display This Question:*

*If Have you ever been addicted to a behaviour (such as gambling)? = Yes*

DRUG.BEH.2.0 What **behaviour** were you addicted to? *(please specify)*

________________________________________________________________

*Display This Question:*

*If Have you ever been addicted to a behaviour (such as gambling)? = Yes*

DRUG.BEH.3.0 Is this addiction or dependence **ongoing?**

- ◯ Yes (1)
- ◯ No (0)
- ◯ Prefer not to answer (-99)

DRUG.ELS.1.0 Have you **ever** been addicted to anything else that has not been mentioned already?

- Yes (1)
- No (0)
- Don't know (-88)
- Prefer not to answer (-99)

---

*Display This Question:*

*If Have you ever been addicted to anything else that has not been mentioned already? = Yes*

DRUG.ELS.2.0 What **else** have you been addicted to? *(please specify)*

________________________________________________________________

*Display This Question:*

*If Have you ever been addicted to anything else that has not been mentioned already? = Yes*

DRUG.ELS.3.0 Is this addiction or dependence **ongoing?**

- Yes (1)
- No (0)
- Prefer not to answer (-99)

DRUG.CAN.1.0 Have you **ever** taken cannabis *(marijuana, grass, hash, ganja, blow, draw, skunk, weed, spliff, dope)*, even if it was a long time ago?

NO SKIP TO HEAD.DUDIT1 NEXT SECTION

- No (0)
- Yes, 1-2 times (1)
- Yes, 3-10 times (2)
- Yes, 11-100 times (3)
- Yes, more than 100 times (4)
- Don't know (-88)
- Prefer not to answer (-99)

*Skip To: HEAD.DUDIT1 If Have you ever taken cannabis (marijuana, grass, hash, ganja, blow, draw, skunk, weed, spliff, dop... = No*

*Skip To: HEAD.DUDIT1 If Have you ever taken cannabis (marijuana, grass, hash, ganja, blow, draw, skunk, weed, spliff, dop... = Don't know*

*Skip To: HEAD.DUDIT1 If Have you ever taken cannabis (marijuana, grass, hash, ganja, blow, draw, skunk, weed, spliff, dop... = Prefer not to answer*

DRUG.CAN.2.0 Considering when you were taking cannabis **most regularly**, how often did you take it?

- ○ Less than once a month (1)
- ○ Once a month or more, but not every week (2)
- ○ Once a week or more, but not every day (3)
- ○ Every day (4)
- ○ Don't know (-88)
- ○ Prefer not to answer (-99)

DRUG.CAN.3.0 About **how old** were you when you **first** had **cannabis**?

________________________________________________________________

DRUG.CAN.4.0 About **how old** were you when you **last** had **cannabis**?

________________________________________________________________

<u>**Games and Gambling Questionnaire**</u>

HEAD.GAM1 We would like to ask you about your experiences with various kinds of gambling. By gambling we mean **placing a bet on the outcome of a race** or a **game of skill or chance**, or **playing a game**, including for charity, in which you might win or lose your money.
**Do not count** any gambling that you may have done for a **prize other than money**, such as a car raffle.

GAM.ACT.1.0 Have you **ever** participated in any of the following activities? *(Please select all that apply)*
Played **electronic gaming machines** such as slot machines, poker machines, video draw poker, or blackjack (1)

- ☐ Bet on **horse or greyhound races excluding** sweeps (2)
- ☐ Bought **instant scratch tickets** (3)
- ☐ Played **lotto** or any other **lottery game** (4)
- ☐ Played **Keno** at a club, hotel or casino (5)
- ☐ Played **poker for money** against other individuals (6)
- ☐ Played **table games** at a casino (not including poker), such as Blackjack or Roulette (7)
- ☐ Played **casino games** on the **internet** (8)
- ☐ Played **bingo** at a club or hall (for cash prizes) (9)
- ☐ Bet on a **sporting event** like football, cricket or tennis (10)
- ☐ Played games like **cards or mahjong** for money at home or any other place (11)
- ☐ Bet on other **games of skill** like billiards (pool) (12)
- ☐ Played any other gambling activity **excluding** raffles or sweeps (13)
- ☐ ⊗ None of the above (0)

*Carry Forward Selected Choices from "Have you ever participated in any of the following activities?  (Please select all that apply)SKIP TO END OF BLOCK (HEAD Section) IF COUNT = 0 "*

HEAD.GAM2 During the **last 12 months**, on how many days have you participated in the following activities?

| | Not at all in the last 12 months (0) | 1 - 10 days (1) | 11 - 100 days (2) | More than 100 days (3) |
|---|---|---|---|---|
| Played **electronic gaming machines** such as slot machines, poker machines, video draw poker, or blackjack (HEAD.GAM2_x1) | ○ | ○ | ○ | ○ |
| Bet on **horse or greyhound races excluding** sweeps (HEAD.GAM2_x2) | ○ | ○ | ○ | ○ |
| Bought **instant scratch tickets** (HEAD.GAM2_x3) | ○ | ○ | ○ | ○ |
| Played **lotto** or any other **lottery game** (HEAD.GAM2_x4) | ○ | ○ | ○ | ○ |
| Played **Keno** at a club, hotel or casino (HEAD.GAM2_x5) | ○ | ○ | ○ | ○ |
| Played **poker for money** against other individuals (HEAD.GAM2_x6) | ○ | ○ | ○ | ○ |
| Played **table games** at a casino (not including poker), such as Blackjack or Roulette (HEAD.GAM2_x7) | ○ | ○ | ○ | ○ |
| Played **casino games** on the **internet** (HEAD.GAM2_x8) | ○ | ○ | ○ | ○ |
| Played **bingo** at a club or hall (for cash prizes) (HEAD.GAM2_x9) | ○ | ○ | ○ | ○ |

| | | | | |
|---|---|---|---|---|
| Bet on a **sporting event** like football, cricket or tennis (HEAD.GAM2_x10) | ○ | ○ | ○ | ○ |
| Played games like **cards or mahjong** for money at home or any other place (HEAD.GAM2_x11) | ○ | ○ | ○ | ○ |
| Bet on other **games of skill** like billiards (pool) (HEAD.GAM2_x12) | ○ | ○ | ○ | ○ |
| Played any other gambling activity **excluding** raffles or sweeps (HEAD.GAM2_x13) | ○ | ○ | ○ | ○ |
| ⊗ None of the above (HEAD.GAM2_x14) | ○ | ○ | ○ | ○ |

*Carry Forward Selected Choices from "Have you ever participated in any of the following activities?  (Please select all that apply)SKIP TO END OF BLOCK (HEAD Section) IF COUNT = 0 "*

HEAD.GAM3 In your **entire life**, on how many days have you participated in the following activities?

| | 1 - 10 days (1) | 11 - 100 days (2) | More than 100 days (3) |
|---|---|---|---|
| Played **electronic gaming machines** such as slot machines, poker machines, video draw poker, or blackjack (HEAD.GAM3_x1) | ○ | ○ | ○ |
| Bet on **horse or greyhound races excluding** sweeps (HEAD.GAM3_x2) | ○ | ○ | ○ |
| Bought **instant scratch tickets** (HEAD.GAM3_x3) | ○ | ○ | ○ |
| Played **lotto** or any other **lottery game** (HEAD.GAM3_x4) | ○ | ○ | ○ |
| Played **Keno** at a club, hotel or casino (HEAD.GAM3_x5) | ○ | ○ | ○ |
| Played **poker for money** against other individuals (HEAD.GAM3_x6) | ○ | ○ | ○ |
| Played **table games** at a casino (not including poker), such as Blackjack or Roulette (HEAD.GAM3_x7) | ○ | ○ | ○ |
| Played **casino games** on the **internet** (HEAD.GAM3_x8) | ○ | ○ | ○ |
| Played **bingo** at a club or hall (for cash prizes) (HEAD.GAM3_x9) | ○ | ○ | ○ |
| Bet on a **sporting event** like football, cricket or tennis (HEAD.GAM3_x10) | ○ | ○ | ○ |
| Played games like **cards or mahjong** for money at home or any other place (HEAD.GAM3_x11) | ○ | ○ | ○ |

| | | | |
|---|---|---|---|
| Bet on other **games of skill** like billiards (pool) (HEAD.GAM3_x12) | ○ | ○ | ○ |
| Played any other gambling activity **excluding** raffles or sweeps (HEAD.GAM3_x13) | ○ | ○ | ○ |
| ⊗ None of the above (HEAD.GAM3_x14) | ○ | ○ | ○ |

*Carry Forward Selected Choices from "Have you ever participated in any of the following activities?  (Please select all that apply)SKIP TO END OF BLOCK (HEAD Section) IF COUNT = 0 "*

HEAD.GAM4 **How old** were you *(in years)* when you **first** participated in the following activities?

| | Age in years (1) |
|---|---|
| Played **electronic gaming machines** such as slot machines, poker machines, video draw poker, or blackjack (GAM.AGE.1.0) | |
| Bet on **horse or greyhound races excluding** sweeps (GAM.AGE.2.0) | |
| Bought **instant scratch tickets** (GAM.AGE.3.0) | |
| Played **lotto** or any other **lottery game** (GAM.AGE.4.0) | |
| Played **Keno** at a club, hotel or casino (GAM.AGE.5.0) | |
| Played **poker for money** against other individuals (GAM.AGE.6.0) | |
| Played **table games** at a casino (not including poker), such as Blackjack or Roulette (GAM.AGE.7.0) | |
| Played **casino games** on the **internet** (GAM.AGE.8.0) | |

| |
|---|
| Played **bingo** at a club or hall (for cash prizes) (GAM.AGE.9.0) |
| Bet on a **sporting event** like football, cricket or tennis (GAM.AGE.10.0) |
| Played games like **cards or mahjong** for money at home or any other place (GAM.AGE.11.0) |
| Bet on other **games of skill** like billiards (pool) (GAM.AGE.12.0) |
| Played any other gambling activity **excluding** raffles or sweeps (GAM.AGE.13.0) |
| ⊗ None of the above (x14) |

GAM.TEN.1.0 Have you ever gambled **at least** 10 times in a single year?

○ No (0)

○ Yes (1)

GAM.SIX.1.0 Have you ever gambled **at least** once a week for **at least** 6 months in a row? *(This does not have to be in the same gambling activity.)*

- ◯ No (0)
- ◯ Yes (1)

*Display This Question:*

*If During the last 12 months, on how many days have you participated in the following activities? [ 1 - 10 days] (Count) > 0*

*Or During the last 12 months, on how many days have you participated in the following activities? [ 11 - 100 days] (Count) > 0*

*Or During the last 12 months, on how many days have you participated in the following activities? [ More than 100 days] (Count) > 0*

GAM.ONL.1.0 In terms of your gambling over the last **12 months**, which of the following statements is **most accurate** for you?

- ◯ I have only **gambled online** in the last 12 months (1)
- ◯ I have **mostly gambled online**, but I have sometimes gambled offline (2)
- ◯ About **half** of my gambling has been **online** and half has been offline (3)
- ◯ I have **mostly gambled offline**, but I have sometimes gambled online (4)
- ◯ I have **never gambled on the internet** in the last 12 months (0)

*Display This Question:*

*If In terms of your gambling over the last 12 months, which of the following statements is most accu... = I have only <strong>gambled online</strong> in the last 12 months*

*Or In terms of your gambling over the last 12 months, which of the following statements is most accu... = I have <strong>mostly gambled online</strong>, but I have sometimes gambled offline*

*Or In terms of your gambling over the last 12 months, which of the following statements is most accu... = About <strong>half </strong>of my gambling has been <strong>online </strong>and half has been offline*

*Or In terms of your gambling over the last 12 months, which of the following statements is most accu... = I have <strong>mostly gambled offline</strong>, but I have sometimes gambled online*

GAM.ONL.2.0 What **year** did you **first** start using the **internet** for gambling purposes?

▼ Before 1995 (1) ... 2020 (27)

*Display This Question:*

*If In terms of your gambling over the last 12 months, which of the following statements is most accu... = I have only <strong>gambled online</strong> in the last 12 months*

*Or In terms of your gambling over the last 12 months, which of the following statements is most accu... = I have <strong>mostly gambled online</strong>, but I have sometimes gambled offline*

*Or In terms of your gambling over the last 12 months, which of the following statements is most accu... = About <strong>half </strong>of my gambling has been <strong>online </strong>and half has been offline*

*Or In terms of your gambling over the last 12 months, which of the following statements is most accu... = I have <strong>mostly gambled offline</strong>, but I have sometimes gambled online*

GAM.ONL.3.0 Thinking about the past **12 months**, what percentage of the **total** amount of **money** you have wagered on all types of gambling has been **online?** *Enter a number between 1 (for 1 % of the total amount of money) and 100 (for 100 % of the total amount of money). Do not enter decimals.*

________________________________________________________________

*Display This Question:*

*If In terms of your gambling over the last 12 months, which of the following statements is most accu... = I have only <strong>gambled online</strong> in the last 12 months*

*Or In terms of your gambling over the last 12 months, which of the following statements is most accu... = I have <strong>mostly gambled online</strong>, but I have sometimes gambled offline*

*Or In terms of your gambling over the last 12 months, which of the following statements is most accu... = About <strong>half </strong>of my gambling has been <strong>online </strong>and half has been offline*

*Or In terms of your gambling over the last 12 months, which of the following statements is most accu... = I have <strong>mostly gambled offline</strong>, but I have sometimes gambled online*

GAM.ONL.4.0 Thinking about the past **12 months**, what percentage of your **total time** spent gambling has been **online?** *Enter a number between 1 (for 1 % of the total amount of time) and 100 (for 100 % of the total amount of time). Do not enter decimals.*

________________________________________________________________

HEAD.GAM5 These next questions ask you about **experiences** people sometimes have with gambling.

| | Never (0) | 1 - 2 times (1) | 3 - 5 times (2) | More than 5 times (3) |
|---|---|---|---|---|
| Have you **ever** bet **more** than you could really **afford** to lose? (GAM.EXP.1.0) | ○ | ○ | ○ | ○ |
| Have you **ever** needed to gamble with **larger amounts of money** to get the **same feeling** of excitement? (GAM.EXP.2.0) | ○ | ○ | ○ | ○ |
| When you gambled, did you **ever** go back another day to try to **win back** the money you **lost?** (GAM.EXP.3.0) | ○ | ○ | ○ | ○ |
| Have you **ever borrowed money** or sold anything to **get** money to gamble? (GAM.EXP.4.0) | ○ | ○ | ○ | ○ |
| Have you **ever** felt that you might have a **problem** with gambling? (GAM.EXP.5.0) | ○ | ○ | ○ | ○ |
| Has gambling **ever** caused you **any** health problems, including **stress or anxiety?** (GAM.EXP.6.0) | ○ | ○ | ○ | ○ |

| | | | | |
|---|---|---|---|---|
| Have people **criticised** your betting or told you that you had a **gambling problem**, regardless of whether or not you thought it was true? (GAM.EXP.7.0) | ○ | ○ | ○ | ○ |
| Has your gambling caused any **financial problems** for you or your household? (GAM.EXP.8.0) | ○ | ○ | ○ | ○ |
| Have you felt **guilty** about the way you gamble or what happens when you gamble? (GAM.EXP.9.0) | ○ | ○ | ○ | ○ |

HEAD.GAM6 Some of these questions may seem **similar** to ones that have already been asked, but there are some slight **differences** in the wording that may change the meaning. Because experts **don't** always agree on the **best** way to measure gambling experiences, we are trying **several** different approaches. Your answers will help us to **better understand** the correct ways to ask such questions.

Remember that the following questions are about your **entire lifetime**, so please indicate if you have **ever** had any of these experiences.

HEAD.GAM7 Has there ever been a period lasting **two weeks or longer** when you ...

| | No (0) | Yes (1) |
|---|---|---|
| Spent a lot of time **thinking** about your gambling experiences? (GAM.FTN.1.0) | ○ | ○ |
| Spent a lot of time **planning** future gambling ventures or bets, or **thinking about ways** of getting money with which to gamble? (GAM.FTN.2.0) | ○ | ○ |
| **Needed** to gamble with **increasing** amounts of money or with **larger** bets than before in order to get the **same feeling** of excitement? (GAM.FTN.3.0) | ○ | ○ |
| Have you ever **tried to stop**, **cut down**, or **control** your gambling? (GAM.FTN.4.0) | ○ | ○ |

*Display This Question:*

*If Has there ever been a period lasting two weeks or longer when you ... = Have you ever <strong>tried to stop</strong>, <strong>cut down</strong>, or <strong>control </strong>your gambling? [ Yes ]*

GAM.STP.1.0 How many times have you **tried to stop, cut down, or control** your gambling?

________________________________________________________________

*Display This Question:*

*If Has there ever been a period lasting two weeks or longer when you ... = Have you ever <strong>tried to stop</strong>, <strong>cut down</strong>, or <strong>control </strong>your gambling? [ Yes ]*

GAM.STP.2.0 Have you ever tried to **stop, cut down, or control** your gambling, but were **unable** to?

- No (0)
- Yes, once or twice (1)
- Yes, 3 or more times (2)

*Display This Question:*

*If Has there ever been a period lasting two weeks or longer when you ... = Have you ever <strong>tried to stop</strong>, <strong>cut down</strong>, or <strong>control </strong>your gambling? [ Yes ]*

GAM.STP.3.0 On one or more of the times when you **tried to stop, cut down, or control** your gambling, were you **restless and irritable?**

- No (0)
- Yes (1)

*Display This Question:*

*If Has there ever been a period lasting two weeks or longer when you ... = Have you ever <strong>tried to stop</strong>, <strong>cut down</strong>, or <strong>control </strong>your gambling? [ Yes ]*

HEAD.GAM8 On one or more of the times when you **tried to stop, cut down, or control** your gambling, did you ever experience any of the following **more than usual**?

| | No (0) | Yes (1) |
|---|---|---|
| Cravings or urges to gamble? (GAM.STP.4.0) | ○ | ○ |
| Sadness or depressed mood? (GAM.STP.5.0) | ○ | ○ |
| Anger (GAM.STP.6.0) | ○ | ○ |
| Difficulty sleeping (GAM.STP.7.0) | ○ | ○ |
| Difficulty concentrating (GAM.STP.8.0) | ○ | ○ |

GAM.PRB.1.0 Have you **ever** gambled as a way to **escape** from personal problems?

○ No (0)

○ Yes (1)

GAM.UNC.1.0 Have you ever gambled to **relieve** uncomfortable feelings such as **guilt, anxiety, helplessness or depression?**

○ No (0)

○ Yes (1)

GAM.RTN.1.0 Has there ever been a period when, if you **lost** money gambling one day, you would often **return** another day to get even?

○ No (0)

○ Yes, once or twice (1)

○ Yes, 3 or more times (2)

GAM.LIE.1.0 Have you **ever lied to family members, friends or others** about how much you gambled or how much money you lost gambling?

- ◯ No (0)
- ◯ Yes, once or twice (1)
- ◯ Yes, 3 or more times (2)

GAM.CHQ.1.0 Have you ever **deliberately** written a **cheque that bounced,** or **stolen** or **taken things** that didn't belong to you in order to gamble?

- ◯ No (0)
- ◯ Yes (1)

GAM.REL.1.0 Has your gambling ever caused **serious or repeated problems** in your **relationships** with any of your family members or friends?

- ◯ No (0)
- ◯ Yes (1)

GAM.JOB.1.0 Has your gambling ever caused you **problems in school**, or to **lose a job**, have **trouble with your job**, or **interfered with your career**?

○ No (0)

○ Yes (1)

GAM.LND.1.0 Have you ever needed to ask family members or anyone else to **lend you money** or otherwise bail you out of a desperate money situation that was largely caused by your gambling?

○ No (0)

○ Yes (1)

HEAD.GAM9 Please think about the **12-month** period in your life when you experienced the **most** problems related to gambling.

Which experiences did you have then?

| | No (0) | Yes (1) |
|---|---|---|
| Spent a lot of time **thinking** about gambling experiences (GAM.MST.1.0) | ◯ | ◯ |
| Spent a lot of time **planning** future gambling or thinking about ways of getting money to gamble (GAM.MST.2.0) | ◯ | ◯ |
| Needed to gamble with **increasing amounts of money** to get the **same feeling** of excitement (GAM.MST.3.0) | ◯ | ◯ |
| **Unable** to stop, cut down or control gambling (GAM.MST.4.0) | ◯ | ◯ |
| **Restless or irritable** when you tried to stop, cut down or control gambling (GAM.MST.5.0) | ◯ | ◯ |
| Gambled to **escape** from personal problems (GAM.MST.6.0) | ◯ | ◯ |
| Gambled to relieve feelings of **guilt, anxiety, helplessness or depression** (GAM.MST.7.0) | ◯ | ◯ |
| After losing money, you would often return another day to get even (GAM.MST.8.0) | ◯ | ◯ |
| **Lied** to family members, friends or others about gambling or money lost gambling (GAM.MST.9.0) | ◯ | ◯ |
| Wrote a cheque that bounced, or took something that didn't belong to you to pay for gambling (GAM.MST.10.0) | ◯ | ◯ |
| Gambling caused **serious or repeated problems** in relationships with family or friends (GAM.MST.11.0) | ◯ | ◯ |
| Gambling caused **problems** in school, or work, or loss of a job, or interfered with your career (GAM.MST.12.0) | ◯ | ◯ |

| | | |
|---|---|---|
| **Needed** family members or anyone else to provide money to get out of a desperate situation caused by gambling (GAM.MST.13.0) | ○ | ○ |

#### Headaches and Migraines

We are interested in knowing more about any experience you may have had with either headache or migraines.

Please try to answer as many questions as you can and give the most accurate answers possible. However, if you cannot answer a question, you can always choose the "don't know" or "prefer not to answer" option where available, or skip the question and move on.

**1) Have you ever had migraine or recurrent attacks of headaches?**

☐Yes *(Continue to 2)* ☐No*(Skip to the end of questionnaire)*

**2) Associated with your headaches, have you ever had recurrent attacks of any of the following?**

| | Yes | No |
|---|---|---|
| **a) Stomach or intestinal pain/dysfunction** | ☐ | ☐ |
| **b) Nausea, vomiting or diarrhoea** | ☐ | ☐ |
| **c) Visual problems such as blurring, showers of light, blind spots, or double vision** | ☐ | ☐ |

**3) Would you describe the pain associated with your headaches as:**

☐**Mild** ☐**Moderate**
☐**Severe** ☐**Unbearable**

**4) How much do your headaches impair your daily activities?**

☐**Not at all** ☐**Interfere with work or social life**
☐**Must stay home from work or school** ☐**Must remain in a dark room *(i.e. go to bed)***

**5) Would you describe the headache pain you usually experience as:**

| | Yes | No |
|---|---|---|
| **a) Throbbing, pulsating or pounding - like being stabbed with a sharp knife** | ☐ | ☐ |
| **b) Pressing - like a weight pushing down on your head** | ☐ | ☐ |
| **c) Squeezing - like a tight band around your head** | ☐ | ☐ |

**6) Do the headaches usually occur on <u>one</u> side of the head?**

☐ **No** ***(pain on both sides)*** ☐ **Left**

☐ **Right** ☐ **Either** ***(pain is sometimes on the left and other times on the right side)***

**7) Associated with your headaches, do you experience enhanced sensitivity to:**

| | Yes | No |
|---|---|---|
| a) Light | ☐ | ☐ |
| b) Smell – such as perfume, petrol or smoke | ☐ | ☐ |
| c) Noise | ☐ | ☐ |

**8) Do these headaches occur in an attack-like manner or are they continuous?**

☐ **Attack-like** ☐ **Continuous**

**9) <u>How old</u> were you the <u>first</u> time you had these headaches?**

☐☐

**10) <u>How old</u> were you the <u>last</u> time you had these headaches?**

☐☐

**11) <u>How many</u> of these headaches have you had <u>during</u> your lifetime?**

☐ **1-2** ☐ **3-4**

☐ **5-10** ☐ **11-50**

☐ **51-100** ☐ **More than 100**

**12) On average, <u>how long</u> does/did a typical untreated or unsuccessfully treated migraine/headache episode last?**

☐ **Days** ***(continue to 12a)*** ☐ **Hours** ***(skip to 12b)***

☐ **Minutes** ***(skip to 12c)***

**12a) Please tell us how many <u>days?</u>**

☐☐

**12b) Please tell us how many hours?**

☐☐

**12c) Please tell us how many minutes?**

☐☐

**13) On average, how often do / did you have these headaches?**

☐**Every day** ☐**5-6 days per week** ☐**3-4 days per week**
☐**2 days per week** ☐**1 day per week** ☐**2-3 days per month**
☐**1 day per month** ☐**3-11 days per year** ☐**Less often**

**14) Are your headaches aggravated by walking up or down stairs or similar routine physical activity?**

☐**Yes** ☐**No**

**15) Associated with your headaches, have you ever had:**

| | Yes | No |
|---|---|---|
| **a) Difficulties speaking** | ☐ | ☐ |
| **b) One-sided numbness or weakness** | ☐ | ☐ |

**16) With your headaches, have you ever had visual disturbances lasting several minutes (e.g. deficiency in your visual fields, scintillating zigzag pattern, sparks or stars in your visual field, blurred or double vision, or some other visual disturbance)?**

☐**Yes** ☐**No**

**17) The following six questions are only relevant if you are female. What was your biological sex at birth?**

☐**Male *(skip to question 24)*** ☐**Female *(continue to question 18)***

**18) When you experience your headaches, do they occur between 2 days before and 2 days after your period starts?** ***If you no longer menstruate, please answer according to how your headaches were when you did menstruate.***

☐**No** ***(skip to question 21)***

☐ **Yes, I have/had these headaches exclusively around this period of menstruation in at least two out of three menstrual cycles and at no other times of the cycle** ***(skip to question 21)***

☐ **Yes, I have/had these headaches around this period of menstruation in at least two out of three menstrual cycles and additionally at other times of the cycle** ***(continue to question 19)***

☐**Yes, I have/had these headaches around this period of menstruation, but in less than two out of three menstrual cycles** ***(continue to question 19)***

☐**Don't know** ***(skip to question 21)***

**19) Approximately what percentage of your headaches occur around menstruation?**

☐☐%

**20) Do / did your headaches around menstruation <u>differ</u> from your headaches at other times?**

☐**Yes** ☐**No**

**21) Do / did you get your headaches with <u>oral contraceptive (Pill)</u> use?**

☐**Yes**

☐**No**

☐ **Not applicable - I've never used oral contraceptives**

☐ **Don't know**

**22) Have you reached <u>menopause</u>?**

☐**Yes** ***(continue to question 23)*** ☐**No** ***(skip to question 24)***

**23) After you reached menopause, did the frequency of your headaches ... ?**

☐**Remain constant**

☐**Increase, but only the headaches occurring around**

☐ **Increase, both the headaches occurring around menstruation and at other**

☐ **Decrease, but only the headaches occurring around**

☐**Decrease, both the headaches around menstruation and at other times**

☐**Not applicable - I haven't reached menopause**

☐**Don't know**

**24) Which of the following medications have you ever taken for your migraine or headaches?** ***Please select all that apply.***

- ☐ **Sumatriptan (e.g. Imigran, Iptam, Sumatab, Sumagran, Sumatran)**
- ☐ **Rizatriptan (e.g. Maxalt)**
- ☐ **Eletriptan (e.g. Relpax)**
- ☐ **Cyproheptadine (e.g. Periactin)**
- ☐ **Botulinum toxin type A (Botox)**
- ☐ **I have never taken medication for migraine or headaches**
- ☐ **Zolmitriptan (e.g. Zomig, Zoltrip)**
- ☐ **Naratriptan (e.g. Naramig)**
- ☐ **Pizotifen (e.g. Sandomigran)**
- ☐ **Topiramate (e.g. Topamax, Epiramax, Tamate)**
- ☐ **Other (specify): ________________________**

### <u>Work and Sleep Questionnaire</u>

HEAD.SLEEP1 This module will ask questions about your **work** and **sleep patterns**. Please answer the following questions as best you can. **If you are retired**, please skip any questions that aren't relevant to you. Click "Next" to continue to the questionnaire.

SLEEP.WRK.1.0 Do you have a **regular** work schedule *(i.e. work the same hours every day on the same days each week)*? This includes being a housewife or househusband.

○ Yes (1)

○ No (0)

*Display This Question:*

*If Do you have a regular work schedule (i.e. work the same hours every day on the same days each wee... = No*

SLEEP.WRK.2.0 Which of the following best describes your **current** work arrangements? *(Please select all that apply)*

☐ Shiftwork with rotating shifts (1)

☐ Shiftwork with irregular shifts (2)

☐ On-call or standby (3)

☐ Overtime or extra hours (paid or unpaid) (4)

☐ Fly-in fly-out (FIFO), drive-in drive-out (DIDO) or equivalent (5)

SLEEP.WRK.7.0 How many **days per week** do you work on **average**?

▼ 0 (0) ... 7 (7)

HEAD.SLEEP2 The following questions relate to your **usual sleep habits** during the **past month only**.

Your answers should indicate the most accurate reply for the **majority** of days and nights in the past month.

HEAD.SLEEP3 During the **past month,** when have you usually gone to bed at night?

| | |
|---|---|
| On work days (SLEEP.HAB.1.0) | ▼ Earlier than 8:00 pm (1) ... Don't know (18) |
| On free days (e.g. weekend) (SLEEP.HAB.2.0) | ▼ Earlier than 8:00 pm (1) ... Don't know (18) |
| In an ideal situation (i.e. you have no responsibilities such as work, children, or engagements the next day) (SLEEP.HAB.3.0) | ▼ Earlier than 8:00 pm (1) ... Don't know (18) |

HEAD.SLEEP4 During the **past month**, when have you usually gotten **up** in the morning?

| | |
|---|---|
| On work days (SLEEP.HAB.6.0) | ▼ Before 4:30 am (1) ... Don't know (21) |
| On free days (e.g. weekend) (SLEEP.HAB.7.0) | ▼ Before 4:30 am (1) ... Don't know (21) |
| In an ideal situation (i.e. you have no responsibilities such as work, children, or engagements the next day) (SLEEP.HAB.8.0) | ▼ Before 4:30 am (1) ... Don't know (21) |

SLEEP.HAB.9.0 During the **past month,** how many hours of **actual sleep** did you get at night? Please give us a rough estimate of the average number of hours sleep per night.

*(This may be different than the number of hours you spend in bed)*

________________________________________________________________

SLEEP.HAB.10.0 Do you have young children who **disrupt** your sleep or who have changed your usual sleep pattern?

○ Yes (1)

○ No (0)

HEAD.SLEEP5 How likely are you to **doze off** or **fall asleep** in the following situations, in contrast to feeling just tired? This refers to your usual way of life in **recent times.** Even if you have not done some of these things recently, try to work out how they would have affected you. Choose the most appropriate option for each situation.

| | Would never doze (0) | Slight chance of dozing (1) | Moderate chance of dozing (2) | High chance of dozing (3) |
|---|---|---|---|---|
| Sitting and reading (SLEEP.SIT.1.0) | ○ | ○ | ○ | ○ |
| Watching TV (SLEEP.SIT.2.0) | ○ | ○ | ○ | ○ |
| Sitting, inactive in a public place (e.g. a theatre or a meeting) (SLEEP.SIT.3.0) | ○ | ○ | ○ | ○ |
| As a passenger in a car for an hour without a break (SLEEP.SIT.4.0) | ○ | ○ | ○ | ○ |
| Lying down to rest in the afternoon when circumstances permit (SLEEP.SIT.5.0) | ○ | ○ | ○ | ○ |
| Sitting and talking to someone (SLEEP.SIT.6.0) | ○ | ○ | ○ | ○ |
| Sitting quietly after lunch without alcohol (SLEEP.SIT.7.0) | ○ | ○ | ○ | ○ |
| In a car, while stopped for a few minutes in traffic (SLEEP.SIT.8.0) | ○ | ○ | ○ | ○ |

SLEEP.HAB.11.0 If you usually have to get up at a specific time in the morning, how much do you **depend** on an alarm clock?

- ○ Not at all (0)
- ○ Slightly (1)
- ○ Somewhat (2)
- ○ Very much (3)

SLEEP.HAB.12.0 During the first half hour **after** you wake up in the morning, how do you feel?

- ○ Very tired (1)
- ○ Fairly tired (2)
- ○ Fairly refreshed (3)
- ○ Very refreshed (4)

SLEEP.HAB.13.0 If you had no commitments the next day, what time would you go to bed **compared** to your usual bedtime?

- ○ Seldom or never later (0)
- ○ Less than 1 hour later (1)
- ○ 1-2 hours later (2)
- ○ More than 2 hours later (3)

SLEEP.HAB.14.0 At **approximately** what time in the evening do you feel tired, and, as a result, in need of sleep?

- 8:00 pm - 9:00 pm (1)
- 9:00 pm - 10:15 pm (2)
- 10:15 pm - 12:45 am (3)
- 12:45 am - 2:00 am (4)
- 2:00 am - 3:00 am (5)

SLEEP.HAB.15.0 At **approximately** what time of day do you usually feel your best?

- 5:00 am - 8:00 am (1)
- 8:00 am - 10:00 am (2)
- 10:00 am - 5:00 pm (3)
- 5:00 pm - 10:00 pm (4)
- 10:00 pm - 5:00 am (5)

SLEEP.HAB.16.0 One hears about **"morning types"** and **"evening types."** Which one of these types do you consider yourself to be?

- Definitely a morning type (1)
- Rather more a morning type than an evening type (2)
- Rather more an evening type than a morning type (3)
- Definitely an evening type (4)

SLEEP.DIF.1.0 Over the **last 2 weeks,** have you had problems with falling asleep, staying asleep or waking up too early?

- Yes (1)
- No (0)

*Display This Question:*

*If Over the last 2 weeks, have you had problems with falling asleep, staying asleep or waking up too... = Yes*

HEAD.SLEEP6 Please rate the **current** *(i.e. last 2 weeks)* **severity** of your insomnia problem(s).

| | None (0) | Mild (1) | Moderate (2) | Severe (3) | Very severe (4) |
|---|---|---|---|---|---|
| Difficulty **falling** asleep (SLEEP.DIF.2.0) | ○ | ○ | ○ | ○ | ○ |
| Difficulty **staying** asleep (SLEEP.DIF.3.0) | ○ | ○ | ○ | ○ | ○ |
| Problem **waking up** too early (SLEEP.DIF.4.0) | ○ | ○ | ○ | ○ | ○ |

SLEEP.DIF.5.0 How **satisfied/dissatisfied** are you with your **current** sleep pattern?

- Very dissatisfied (0)
- Dissatisfied (1)
- Moderately satisfied (2)
- Satisfied (3)
- Very satisfied (4)

*Display This Question:*

*If How satisfied/dissatisfied are you with your current sleep pattern? = Very dissatisfied*

*Or How satisfied/dissatisfied are you with your current sleep pattern? = Dissatisfied*

*Or How satisfied/dissatisfied are you with your current sleep pattern? = Moderately satisfied*

*Or Please rate the current (i.e. last 2 weeks) severity of your insomnia problem(s). [ Mild] (Count) > 0*

*Or Please rate the current (i.e. last 2 weeks) severity of your insomnia problem(s). [ Moderate] (Count) > 0*

*Or Please rate the current (i.e. last 2 weeks) severity of your insomnia problem(s). [ Severe] (Count) > 0*

*Or Please rate the current (i.e. last 2 weeks) severity of your insomnia problem(s). [ Very severe] (Count) > 0*

SLEEP.DIF.6.0 How **noticeable** to others do you think your poor sleep is in terms of impairing the quality of your life?

- ◯ Not at all noticeable (0)
- ◯ A little (1)
- ◯ Somewhat (2)
- ◯ Much (3)
- ◯ Very much noticeable (4)

*Display This Question:*

*If How satisfied/dissatisfied are you with your current sleep pattern? = Very dissatisfied*

*Or How satisfied/dissatisfied are you with your current sleep pattern? = Dissatisfied*

*Or How satisfied/dissatisfied are you with your current sleep pattern? = Moderately satisfied*

*Or Please rate the current (i.e. last 2 weeks) severity of your insomnia problem(s). [ Mild] (Count) > 0*

*Or Please rate the current (i.e. last 2 weeks) severity of your insomnia problem(s). [ Moderate] (Count) > 0*

*Or Please rate the current (i.e. last 2 weeks) severity of your insomnia problem(s). [ Severe] (Count) > 0*

*Or Please rate the current (i.e. last 2 weeks) severity of your insomnia problem(s). [ Very severe] (Count) > 0*

SLEEP.DIF.7.0 How **worried/distressed** are you about your current sleep problem?

- ◯ Not at all worried (0)
- ◯ A little (1)
- ◯ Somewhat (2)
- ◯ Much (3)
- ◯ Very much worried (4)

*Display This Question:*

*If How satisfied/dissatisfied are you with your current sleep pattern? = Very dissatisfied*

*Or How satisfied/dissatisfied are you with your current sleep pattern? = Dissatisfied*

*Or How satisfied/dissatisfied are you with your current sleep pattern? = Moderately satisfied*

*Or Please rate the current (i.e. last 2 weeks) severity of your insomnia problem(s). [ Mild] (Count) > 0*

*Or Please rate the current (i.e. last 2 weeks) severity of your insomnia problem(s). [ Moderate] (Count) > 0*

*Or Please rate the current (i.e. last 2 weeks) severity of your insomnia problem(s). [ Severe] (Count) > 0*

*Or Please rate the current (i.e. last 2 weeks) severity of your insomnia problem(s). [ Very severe] (Count) > 0*

SLEEP.DIF.8.0 To what extent do you consider your sleep problem to **interfere** with your daily functioning (e.g, daytime fatigue, ability to function at work/daily chores, concentration, memory, mood, etc.) **currently**?

- ◯ Not at all (0)
- ◯ A little (1)
- ◯ Somewhat (2)
- ◯ Much (3)
- ◯ Very much (4)

SLEEP.DIF.9.0 During the past month, how often have you taken **medicine** to help you sleep? *(Please include both prescribed or "over the counter")*

- ◯ Not during the past month (0)
- ◯ Less than once a week (1)
- ◯ Once or twice per week (2)
- ◯ Three or more times per week (3)

SLEEP.DIF.10.0 During the past month, how often have you had **trouble staying awake** while driving, eating meals, or engaging in social activity?

- ◯ Not during the past month (0)
- ◯ Less than once a week (1)
- ◯ Once or twice per week (2)
- ◯ Three or more times per week (3)

SLEEP.CAF.1.0 If you were to drink coffee in the evening, would it **stop** you from getting to sleep?

- ◯ Yes (1)
- ◯ No (0)
- ◯ I don't drink coffee (2)

HEAD.CAF1 How many cups/cans/bottles of the following **caffeinated beverages** do you drink **per day?** *Please note that decaffeinated coffee or caffeine-free cola do not count towards this total.)*

Please click or tap on the dropdown list to register your response, even if the answer is "0".

| | |
|---|---|
| Coffee (SLEEP.CAF.2.0) | ▼ 0 (1) ... 10+ (11) |
| Tea (SLEEP.CAF.3.0) | ▼ 0 (1) ... 10+ (11) |
| Fizzy drinks (e.g. Coca-Cola, Pepsi, Mountain Dew, etc) (SLEEP.CAF.4.0) | ▼ 0 (1) ... 10+ (11) |
| Energy drinks (e.g. Red Bull, Mother, Rockstar) (SLEEP.CAF.5.0) | ▼ 0 (1) ... 10+ (11) |

HEAD.CAF2 On average, how much time do you **spend outdoors** in natural light **per day**?

| | |
|---|---|
| On work days (SLEEP.OUT.1.0) | ▼ Less than 1 hour (1) ... 15+ hours (16) |
| On free days (e.g. weekend) (SLEEP.OUT.2.0) | ▼ Less than 1 hour (1) ... 15+ hours (16) |
| In an ideal situation (i.e. you have no responsibilities such as work, children, or engagements the next day) (SLEEP.OUT.3.0) | ▼ Less than 1 hour (1) ... 15+ hours (16) |

HEAD.CAF3 During the last month, on **how many** nights or days per week have you had or been told you had the following:

| | Never (0) | Rarely, less than once a week (1) | 1-2 times per week (2) | 3-4 times per week (3) | 5-7 times per week (4) | Don't know (-88) |
|---|---|---|---|---|---|---|
| Loud snoring (SLEEP.BRE.1.0) | ○ | ○ | ○ | ○ | ○ | ○ |
| Snorting or gasping (SLEEP.BRE.2.0) | ○ | ○ | ○ | ○ | ○ | ○ |
| Your breathing stops or you choke or struggle for breath (SLEEP.BRE.3.0) | ○ | ○ | ○ | ○ | ○ | ○ |

HEAD.LIV1 The purpose of the following questions is to find out how your mood and behaviour **change over time.**

*We are interested in **your** experience, not others you may have observed.*

---

SLEEP.LIV.1.0 For **how long** have you **lived** in your **current** town or in the surrounding area?

*You may enter number of years and/or number of months. Number of months must not exceed 12.*

▢ Years (1) ________________________________________________

▢ Months (2) ________________________________________________

HEAD.SC1 To what degree do the following change **with the seasons**?

| | No change (0) | Slight change (1) | Moderate change (2) | Marked change (3) | Extremely marked change (4) |
|---|---|---|---|---|---|
| Sleep length (SLEEP.SC.1.0) | ○ | ○ | ○ | ○ | ○ |
| Social activity (SLEEP.SC.2.0) | ○ | ○ | ○ | ○ | ○ |
| Mood (overall feeling of well being) (SLEEP.SC.3.0) | ○ | ○ | ○ | ○ | ○ |
| Weight (SLEEP.SC.4.0) | ○ | ○ | ○ | ○ | ○ |
| Appetite (SLEEP.SC.5.0) | ○ | ○ | ○ | ○ | ○ |
| Energy level (SLEEP.SC.6.0) | ○ | ○ | ○ | ○ | ○ |

HEAD.SC2 In the following question, please select **all applicable months.** This may be a single month, a cluster of months, or any other grouping. *(If there are no effects, select N/A)*

At what **time of year** do you....?

| | Jan (1) | Feb (2) | Mar (3) | Apr (4) | May (5) | Jun (6) | Jul (7) | Aug (8) | Sep (9) | Oct (10) | Nov (11) | Dec (12) | No particular months (0) | N/A (0) |
|---|---|---|---|---|---|---|---|---|---|---|---|---|---|---|
| Feel best (SLEEP.FB.1.0) | | | | | | | | | | | | | ☐ | |
| Tend to gain most weight (SLEEP.WG.1.0) | | | | | | | | | | | | | ☐ | |
| Socialise most (SLEEP.SM.1.0) | | | | | | | | | | | | | ☐ | |
| Sleep least (SLEEP.SLL.1.0) | | | | | | | | | | | | | ☐ | |
| Eat most (SLEEP.EM.1.0) | | | | | | | | | | | | | ☐ | |
| Lose most weight (SLEEP.LW.1.0) | | | | | | | | | | | | | ☐ | |
| Socialise least (SLEEP.SL.1.0) | | | | | | | | | | | | | ☐ | |
| Feel worst (SLEEP.FW.1.0) | | | | | | | | | | | | | ☐ | |
| Eat least (SLEEP.EL.1.0) | | | | | | | | | | | | | ☐ | |
| Sleep most (SLEEP.SLM.1.0) | | | | | | | | | | | | | ☐ | |

*Display This Question:*

*If To what degree do the following change with the seasons? [ Slight change] (Count) > 0*

*And To what degree do the following change with the seasons? [ Moderate change] (Count) > 0*

*And To what degree do the following change with the seasons? [ Marked change] (Count) > 0*

*And To what degree do the following change with the seasons? [ Extremely marked change] (Count) > 0*

SLEEP.SEA.1.0 If you experience changes with the seasons (in energy, mood, sleep etc), do you feel that they are a **problem** for you?

- Yes (1)
- No (0)

*Display This Question:*

*If If you experience changes with the seasons (in energy, mood, sleep etc), do you feel that they ar... = Yes*

SLEEP.SEA.2.0 Is the problem...?

- Mild (1)
- Moderate (2)
- Marked (3)
- Severe (4)
- Disabling (5)

HEAD.SEA1 Approximately how many **hours** of each 24-hour day do you **sleep** during each **season**? *(Include naps)*

| | |
|---|---|
| Winter (SLEEP.24H.1.0) | ▼ 0 (1) ... Over 18 hours (20) |
| Spring (SLEEP.24H.2.0) | ▼ 0 (1) ... Over 18 hours (20) |
| Summer (SLEEP.24H.3.0) | ▼ 0 (1) ... Over 18 hours (20) |
| Autumn (SLEEP.24H.4.0) | ▼ 0 (1) ... Over 18 hours (20) |

#### SF-12®  Patient Questionnaire

**Patient Initials _____ _____ ______ Date of Birth: ___/___/___ Patkey: ______**

**Surgeon Name:** ______________________________________________ **Date:** ____________

**Examination Period:**
_____ Preop (1)
_____ Immediate Postop (2)
_____ 1 Year (3)
_____ 3 Year (4)
_____ 5 Year (5)
_____ Other (specify) (6): _____________

---

**SF-12®:**
This information will help your doctors keep track of how you feel and how well you are able to do your usual activities. Answer every question by placing a check mark on the line in front of the appropriate answer. It is not specific for arthritis. If you are unsure about how to answer a question, please give the best answer you can and make a written comment beside your answer.

1. In general, would you say your health is:
_____ Excellent (1)
_____ Very Good (2)
_____ Good (3)
_____ Fair (4)
_____ Poor (5)

The following two questions are about activities you might do during a typical day. Does YOUR HEALTH NOW LIMIT YOU in these activities? If so, how much?

2. MODERATE ACTIVITIES, such as moving a table, pushing a vacuum cleaner, bowling, or playing golf:
_____ Yes, Limited A Lot (1)
_____ Yes, Limited A Little (2)
_____ No, Not Limited At All (3)

3. Climbing SEVERAL flights of stairs:
_____ Yes, Limited A Lot (1)
_____ Yes, Limited A Little (2)
_____ No, Not Limited At All (3)

During the PAST 4 WEEKS have you had any of the following problems with your work or other regular activities AS A RESULT OF YOUR PHYSICAL HEALTH?

4. ACCOMPLISHED LESS than you would like:
_____ Yes (1)
_____ No (2)

5. Were limited in the KIND of work or other activities:
_____ Yes (1)
_____ No (2)

**Surgeon Initials __________ Date: _____________**

**Patient Initials** _____ _____ ______ **Date of Birth:** ___/___/___ **Patkey:** ______

**Surgeon Name:** ____________________________________________ **Date:** ____________

**Examination Period:** _____ Preop (1) _____ 3 Year (4)
_____ Immediate Postop (2) _____ 5 Year (5)
_____ 1 Year (3) _____ Other (specify) (6): ____________

---

**SF-12® Cont'd:**

During the PAST 4 WEEKS, were you limited in the kind of work you do or other regular activities AS A RESULT OF ANY EMOTIONAL PROBLEMS (such as feeling depressed or anxious)?

6. ACCOMPLISHED LESS than you would like:
   _____ Yes (1)
   _____ No (2)

7. Didn't do work or other activities as CAREFULLY as usual:
   _____ Yes (1)
   _____ No (2)

8. During the PAST 4 WEEKS, how much did PAIN interfere with your normal work (including both work outside the home and housework)?
   _____ Not At All (1)
   _____ A Little Bit (2)
   _____ Moderately (3)
   _____ Quite A Bit (4)
   _____ Extremely (5)

The next three questions are about how you feel and how things have been DURING THE PAST 4 WEEKS. For each question, please give the one answer that comes closest to the way you have been feeling. How much of the time during the PAST 4 WEEKS –

9. Have you felt calm and peaceful?
   _____ All of the Time (1)
   _____ Most of the Time (2)
   _____ A Good Bit of the Time (3)
   _____ Some of the Time (4)
   _____ A Little of the Time (5)
   _____ None of the Time (6)

**Surgeon Initials** __________ **Date:** _____________

**Patient Initials** _____ _____ ______ **Date of Birth:** ___/___/___ **Patkey:** ______

**Surgeon Name:** ____________________________________________ **Date:** ___________

**Examination Period:**
_____ Preop (1)
_____ Immediate Postop (2)
_____ 1 Year (3)
_____ 3 Year (4)
_____ 5 Year (5)
_____ Other (specify) (6): ___________

---

**SF-12® Cont'd:**

10. Did you have a lot of energy?
    _____ All of the Time (1)
    _____ Most of the Time (2)
    _____ A Good Bit of the Time (3)
    _____ Some of the Time (4)
    _____ A Little of the Time (5)
    _____ None of the Time (6)

11. Have you felt downhearted and blue?
    _____ All of the Time (1)
    _____ Most of the Time (2)
    _____ A Good Bit of the Time (3)
    _____ Some of the Time (4)
    _____ A Little of the Time (5)
    _____ None of the Time (6)

12. During the PAST 4 WEEKS, how much of the time has your PHYSICAL HEALTH OR EMOTIONAL PROBLEMS interfered with your social activities (like visiting with friends, relatives, etc.)?
    _____ All of the Time (1)
    _____ Most of the Time (2)
    _____ A Good Bit of the Time (3)
    _____ Some of the Time (4)
    _____ A Little of the Time (5)
    _____ None of the Time (6)

**Surgeon Signature**_____________________________________________ **Date**________________________

SF-12® Health Survey © 1994, 2002 by Medical Outcomes Trust and QualityMetric Incorporated. All Rights Reserved
SF-12® is a registered trademark of Medical Outcomes Trust

<u>**Eating Disorder Quality of Life Questionnaire**</u>

Q6 *The following section relates to the negative impact eating behaviour and weight can have in a person's life.*

Please answer the following statements according to how well they describe you in the **last 30 days**. Please be as open as possible. There are no right or wrong answers.

For those items that do not apply to you, please leave them blank.

*If you feel uncomfortable and do not wish to continue answering, please feel free to skip any of the questions.*

Q1 In the **last 30 days**, how often has your eating/weight...

| | Never (0) | Rarely (1) | Sometimes (2) | Often (3) | Always (4) |
|---|---|---|---|---|---|
| Resulted in you **feeling embarrassed** or **"different"**? (1) | ○ | ○ | ○ | ○ | ○ |
| Made you feel **worse** about yourself? (2) | ○ | ○ | ○ | ○ | ○ |
| Made you **not want** to be with people? (3) | ○ | ○ | ○ | ○ | ○ |
| Resulted in you believing that you will **never** get better? (4) | ○ | ○ | ○ | ○ | ○ |

Q2 In the last 30 days, how often has your eating/weight...

| | Never (1) | Rarely (2) | Sometimes (3) | Often (4) | Always (5) |
|---|---|---|---|---|---|
| Made you feel **lonely**? (1) | ○ | ○ | ○ | ○ | ○ |
| Resulted in **less interest** or **pleasure** in activities? (2) | ○ | ○ | ○ | ○ | ○ |
| Led you to not **care** about yourself? (3) | ○ | ○ | ○ | ○ | ○ |
| Made you feel **odd**, **weird**, or **unusual**? (4) | ○ | ○ | ○ | ○ | ○ |
| Resulted in **avoiding** eating in front of others? (5) | ○ | ○ | ○ | ○ | ○ |

Q3 In the last 30 days, how often has your eating/weight...

| | Never (1) | Rarely (2) | Sometimes (3) | Often (4) | Always (5) |
|---|---|---|---|---|---|
| Caused **cold** hands and feet? (1) | ○ | ○ | ○ | ○ | ○ |
| Caused **frequent** headaches? (2) | ○ | ○ | ○ | ○ | ○ |
| Caused **weakness**? (3) | ○ | ○ | ○ | ○ | ○ |
| Affected your ability to **pay attention** when you wanted to? (4) | ○ | ○ | ○ | ○ | ○ |
| Affected your ability to **comprehend** some verbal and written information? (5) | ○ | ○ | ○ | ○ | ○ |
| Reduced your ability to **concentrate**? (6) | ○ | ○ | ○ | ○ | ○ |

Q4 In the last 30 days, how often has your eating/weight...

| | Never (1) | Rarely (2) | Sometimes (3) | Often (4) | Always (5) |
|---|---|---|---|---|---|
| Led to problems with treatment provider(s) regarding **cost of treatment**? (1) | ○ | ○ | ○ | ○ | ○ |
| Led to you having **difficulties paying** monthly bills? (2) | ○ | ○ | ○ | ○ | ○ |
| Resulted in significant financial **debt**? (3) | ○ | ○ | ○ | ○ | ○ |
| Led to the need to **spend money** from savings or used your credit card frequently? (4) | ○ | ○ | ○ | ○ | ○ |
| Resulted in the need to **borrow** money? (5) | ○ | ○ | ○ | ○ | ○ |

Q5 In the last 30 days, how often has your eating/weight...

| | Never (1) | Rarely (2) | Sometimes (3) | Often (4) | Always (5) |
|---|---|---|---|---|---|
| Led to a **leave of absence** from work? (1) | ○ | ○ | ○ | ○ | ○ |
| Led to **low grades**? (2) | ○ | ○ | ○ | ○ | ○ |
| Resulted in **reduced hours** at work? (3) | ○ | ○ | ○ | ○ | ○ |
| Resulted in you **losing a job** or **dropping out** of school? (4) | ○ | ○ | ○ | ○ | ○ |
| Led to **failure** in a class or classes? (5) | ○ | ○ | ○ | ○ | ○ |

**The APA is offering a number of "emerging measures" for further research and clinical evaluation. These patient assessment measures were developed to be administered at the initial patient interview and to monitor treatment progress. They should be used in research and evaluation as potentially useful tools to enhance clinical decision-making and not as the sole basis for making a clinical diagnosis. Instructions, scoring information, and interpretation guidelines are provided; further background information can be found in DSM-5. The APA requests that clinicians and researchers provide further data on the instruments' usefulness in characterizing patient status and improving patient care at http://www.dsm5.org/Pages/Feedback-Form.aspx.**

**Measure:** The Personality Inventory for DSM-5—Brief Form (PID-5-BF)—Adult
**Rights granted:** This measure can be reproduced without permission by researchers and by clinicians for use with their patients.
**Rights holder:** American Psychiatric Association
**To request permission for any other use beyond what is stipulated above, contact:** http://www.appi.org/CustomerService/Pages/Permissions.aspx

### The Personality Inventory for DSM-5—Brief Form (PID-5-BF)—Adult

**Name:** ________________________ **Age:** ____ **Sex:** ❑ **Male** ❑ **Female** **Date:**_____________

| **Instructions:** This is a list of things different people might say about themselves. We are interested in how you would describe yourself. There are no right or wrong answers. So you can describe yourself as honestly as possible, we will keep your responses confidential. We'd like you to take your time and read each statement carefully, selecting the response that best describes you. | | | | | | **Clinician Use** |
|---|---|---|---|---|---|---|
| | | **Very False or Often False** | **Sometimes or Somewhat False** | **Sometimes or Somewhat True** | **Very True or Often True** | **Item score** |
| 1 | People would describe me as reckless. | 0 | 1 | 2 | 3 | |
| 2 | I feel like I act totally on impulse. | 0 | 1 | 2 | 3 | |
| 3 | Even though I know better, I can't stop making rash decisions. | 0 | 1 | 2 | 3 | |
| 4 | I often feel like nothing I do really matters. | 0 | 1 | 2 | 3 | |
| 5 | Others see me as irresponsible. | 0 | 1 | 2 | 3 | |
| 6 | I'm not good at planning ahead. | 0 | 1 | 2 | 3 | |
| 7 | My thoughts often don't make sense to others. | 0 | 1 | 2 | 3 | |
| 8 | I worry about almost everything. | 0 | 1 | 2 | 3 | |
| 9 | I get emotional easily, often for very little reason. | 0 | 1 | 2 | 3 | |
| 10 | I fear being alone in life more than anything else. | 0 | 1 | 2 | 3 | |
| 11 | I get stuck on one way of doing things, even when it's clear it won't work. | 0 | 1 | 2 | 3 | |
| 12 | I have seen things that weren't really there. | 0 | 1 | 2 | 3 | |
| 13 | I steer clear of romantic relationships. | 0 | 1 | 2 | 3 | |
| 14 | I'm not interested in making friends. | 0 | 1 | 2 | 3 | |
| 15 | I get irritated easily by all sorts of things. | 0 | 1 | 2 | 3 | |
| 16 | I don't like to get too close to people. | 0 | 1 | 2 | 3 | |
| 17 | It's no big deal if I hurt other peoples' feelings. | 0 | 1 | 2 | 3 | |
| 18 | I rarely get enthusiastic about anything. | 0 | 1 | 2 | 3 | |
| 19 | I crave attention. | 0 | 1 | 2 | 3 | |
| 20 | I often have to deal with people who are less important than me. | 0 | 1 | 2 | 3 | |
| 21 | I often have thoughts that make sense to me but that other people say are strange. | 0 | 1 | 2 | 3 | |
| 22 | I use people to get what I want. | 0 | 1 | 2 | 3 | |
| 23 | I often "zone out" and then suddenly come to and realize that a lot of time has passed. | 0 | 1 | 2 | 3 | |
| 24 | Things around me often feel unreal, or more real than usual. | 0 | 1 | 2 | 3 | |
| 25 | It is easy for me to take advantage of others. | 0 | 1 | 2 | 3 | |
| | | | | | Total/Partial Raw Score: | |
| | | | | | Prorated Total Score: (if 1-6 items left unanswered) | |
| | | | | | Average Total Score: | |

Krueger RF, Derringer J, Markon KE, Watson D, Skodol AE.

Copyright © 2013 American Psychiatric Association. All Rights Reserved.
This material can be reproduced without permission by researchers and by clinicians for use with their patients.

**Personality Trait Domain Scoring**

| FOR CLINICIAN USE ONLY | Personality Trait Domain | PID-5 BF items | Total/Partial Raw Domain Score | Prorated Domain Score | Average Domain Score |
|---|---|---|---|---|---|
| | Negative Affect | 8, 9, 10, 11, 15 | | | |
| | Detachment | 4, 13, 14, 16, 18 | | | |
| | Antagonism | 17, 19, 20, 22, 25 | | | |
| | Disinhibition | 1, 2, 3, 5, 6 | | | |
| | Psychoticism | 7, 12, 21, 23, 24 | | | |

**Instructions to Clinicians**

This Personality Inventory for DSM-5—Brief Form (PID-5-BF)—Adult is a 25-item self-rated personality trait assessment scale for adults age 18 and older. It assesses 5 personality trait domains including negative affect, detachment, antagonism, disinhibition, and psychoticism, with each trait domain consisting of 5 items. The measure is completed by the individual prior to a visit with the clinician. If the individual receiving care is an adult age 18 and older with impaired capacity and unable to complete the form, a knowledgeable informant may complete the informant version of the measure (the PID-5-IRF). Each item on the PID-5-BF asks the individual receiving care to rate how well the item describes him or her generally.

**Scoring and Interpretation**

Each item on the measure is rated on a 4-point scale (i.e., 0=very false or often false; 1=sometimes or somewhat false; 2=sometimes or somewhat true; 3=very true or often true). The overall measure has a range of scores from 0 to 75, with higher scores indicating greater overall personality dysfunction. Each trait domain ranges in score from 0 to 15, with higher scores indicating greater dysfunction in the specific personality trait domain. The clinician is asked to review the score on each item on the measure during the clinical interview and indicate the raw score for each item in the section provided for "Clinician Use." The raw scores on the 25 items should be summed to obtain a total raw score. The scores on the items within each trait domain should be summed and entered in the appropriate raw domain score box. In addition, the clinician is asked to calculate and use **average scores for each domain and for the overall measure**. **The average scores** reduce the overall score as well as the scores for each domain to a 4-point scale, which allows the clinician to think of the patient's personality dysfunction relative to observed norms.[1] The **average domain score** is calculated by dividing the raw domain score by the number of items in the domain (e.g., if all the items within the "negative affect " domain are rated as being "sometimes or somewhat true" then the average domain score would be 10/5 = 2, indicating moderate negative affect). The **average total score** is calculated by dividing the raw overall score by the total number of items in the measure (i.e., 25). The average domain and overall personality dysfunction scores were found to be reliable, easy to use, and clinically useful to the clinicians in the DSM-5 Field Trials.

**Note:** If 7 or more items are left unanswered on the measure (i.e., more than 25% of the total items are missing), the scores should not be calculated. Similarly, if 2 or more items are left unanswered on any one domain, the domain score should not be calculated. Therefore, the individual should be encouraged to complete all of the items on the measure. However, if 7 or more of the total items on the measure are left unanswered but 4 or 5 items for some of the domains are completed, the raw or average domain scores may be used for those domains. If for the overall measure 1 to 6 items are left unanswered, or for any domain only one item is left unanswered, you may prorate the total raw score or domain score by first summing the number of items that were answered to get a **partial raw score**. Next, multiply the partial raw score by the total number of items on the measure (i.e., 25) or in the domain (i.e., 5). Finally, divide the value by the number of items that were actually answered to obtain the prorated total or domain raw score.

$$\text{Prorated Score} = \frac{\text{(Partial Raw Score x number of items on the PID-5 BF)}}{\text{Number of items that were actually answered}}$$

If the result is a fraction, round to the nearest whole number.

**Frequency of Use**

To track change in the severity of the individual's personality dysfunction over time, it is recommended that the measure be completed at regular intervals as clinically indicated, depending on the stability of the individual's symptoms and treatment status. Consistently high scores on a particular domain may indicate significant and problematic areas for the individual receiving care that might warrant further assessment, treatment, and follow-up. Your clinical judgment should guide your decision.

[1]Krueger RF, Derringer J, Markon KE, Watson D, Skodol AE. (2013). *The Personality Inventory for DSM-5 Brief Form (PID-5-BF).* Manuscript in preparation.

Copyright © 2013 American Psychiatric Association. All Rights Reserved.
This material can be reproduced without permission by researchers and by clinicians for use with their patients

**Personal Standards Questionnaire**

HEAD.MPS.1.0 The following pages contain a number of statements concerning personal characteristics and traits. Read each item and decide whether you agree or disagree & to what extent.

| Q1 | Disagree (1) (1) | 2 (2) | 3 (3) | 4 (4) | 5 (5) | 6 (6) | Agree (7) (7) |
|---|---|---|---|---|---|---|---|
| When I am working on something, I **cannot relax** until it is perfect (1) | ○ | ○ | ○ | ○ | ○ | ○ | ○ |
| I am not likely to criticise someone for **giving up** too easily (2) | ○ | ○ | ○ | ○ | ○ | ○ | ○ |
| It is **not important** that people I am close to are successful (3) | ○ | ○ | ○ | ○ | ○ | ○ | ○ |
| I seldom criticise my friends for accepting **second best** (4) | ○ | ○ | ○ | ○ | ○ | ○ | ○ |
| I find it **difficult** to meet others' expectations of me (5) | ○ | ○ | ○ | ○ | ○ | ○ | ○ |
| One of my goals is to be **perfect** in everything I do (6) | ○ | ○ | ○ | ○ | ○ | ○ | ○ |
| Everything that **others** do must be of top-notch quality (7) | ○ | ○ | ○ | ○ | ○ | ○ | ○ |

| Q2 | Disagree (1) (1) | 2 (2) | 3 (3) | 4 (4) | 5 (5) | 6 (6) | Agree (7) (7) |
|---|---|---|---|---|---|---|---|
| I **never** aim for perfection on my work (1) | ○ | ○ | ○ | ○ | ○ | ○ | ○ |
| Those around me readily **accept** that I can make mistakes too (2) | ○ | ○ | ○ | ○ | ○ | ○ | ○ |
| It **doesn't matter** when someone close to me does not do their absolute best (3) | ○ | ○ | ○ | ○ | ○ | ○ | ○ |
| The better I do, the better I am **expected** to do (4) | ○ | ○ | ○ | ○ | ○ | ○ | ○ |
| I **seldom** feel the need to be **perfect** (5) | ○ | ○ | ○ | ○ | ○ | ○ | ○ |
| Anything that I do that is **less** than excellent will be seen as poor work by those around me (6) | ○ | ○ | ○ | ○ | ○ | ○ | ○ |

| | | | | | | | |
|---|---|---|---|---|---|---|---|
| I **strive** to be as **perfect** as I can be (7) | ○ | ○ | ○ | ○ | ○ | ○ | ○ |

| Q3 | Disagree (1) (1) | 2 (2) | 3 (3) | 4 (4) | 5 (5) | 6 (6) | Agree (7) (7) |
|---|---|---|---|---|---|---|---|
| It is very **important** that I am perfect in everything I attempt (1) | ○ | ○ | ○ | ○ | ○ | ○ | ○ |
| I have **high expectations** for the people who are important to me (2) | ○ | ○ | ○ | ○ | ○ | ○ | ○ |
| I **strive** to be the **best** at everything I do (3) | ○ | ○ | ○ | ○ | ○ | ○ | ○ |
| The people around me expect me to **succeed** at everything I do (4) | ○ | ○ | ○ | ○ | ○ | ○ | ○ |
| I do **not** have very high standards for those around me (5) | ○ | ○ | ○ | ○ | ○ | ○ | ○ |
| I demand **nothing less** than perfection of myself (6) | ○ | ○ | ○ | ○ | ○ | ○ | ○ |
| Others will like me **even if I don't** excel at everything (7) | ○ | ○ | ○ | ○ | ○ | ○ | ○ |

| Q4 | Disagree (1) (1) | 2 (2) | 3 (3) | 4 (4) | 5 (5) | 6 (6) | Agree (7) (7) |
|---|---|---|---|---|---|---|---|
| I **can't be bothered** with people who won't strive to better themselves (1) | ○ | ○ | ○ | ○ | ○ | ○ | ○ |
| It makes me **uneasy** to see an error in my work (2) | ○ | ○ | ○ | ○ | ○ | ○ | ○ |
| I **do not** expect a lot from my friends (3) | ○ | ○ | ○ | ○ | ○ | ○ | ○ |
| Success means that I must work **even harder** to please others (4) | ○ | ○ | ○ | ○ | ○ | ○ | ○ |
| If I ask someone to do something, I expect it to be done **flawlessly** (5) | ○ | ○ | ○ | ○ | ○ | ○ | ○ |
| I **cannot stand** to see people close to me make mistakes (6) | ○ | ○ | ○ | ○ | ○ | ○ | ○ |
| I am **perfectionistic** in setting my goals (7) | ○ | ○ | ○ | ○ | ○ | ○ | ○ |

| Q5 | Disagree (1) (1) | 2 (2) | 3 (3) | 4 (4) | 5 (5) | 6 (6) | Agree (7) (7) |
|---|---|---|---|---|---|---|---|
| The people who matter to me should **never** let me down (1) | ○ | ○ | ○ | ○ | ○ | ○ | ○ |
| Others think I am okay, **even** when I do not succeed (2) | ○ | ○ | ○ | ○ | ○ | ○ | ○ |
| I feel that people are **too demanding** of me (3) | ○ | ○ | ○ | ○ | ○ | ○ | ○ |
| I **must** work to my **full potential** at all times (4) | ○ | ○ | ○ | ○ | ○ | ○ | ○ |
| Although they **may not say it**, other people get very upset with me when I slip up (5) | ○ | ○ | ○ | ○ | ○ | ○ | ○ |
| I do **not** have to be the best at whatever I am doing (6) | ○ | ○ | ○ | ○ | ○ | ○ | ○ |
| My **family** expects me to be perfect (7) | ○ | ○ | ○ | ○ | ○ | ○ | ○ |

| Q6 | Disagree (1) (1) | 2 (2) | 3 (3) | 4 (4) | 5 (5) | 6 (6) | Agree (7) (7) |
|---|---|---|---|---|---|---|---|
| I do not have very high **goals** for myself (1) | ○ | ○ | ○ | ○ | ○ | ○ | ○ |
| My parent **rarely** expected me to excel in all aspects of my life (2) | ○ | ○ | ○ | ○ | ○ | ○ | ○ |
| I **respect** people who are average (3) | ○ | ○ | ○ | ○ | ○ | ○ | ○ |
| People expect **nothing less** than perfection from me (4) | ○ | ○ | ○ | ○ | ○ | ○ | ○ |
| I set very **high standards** for myself (5) | ○ | ○ | ○ | ○ | ○ | ○ | ○ |
| People expect **more** from me than I am capable of giving (6) | ○ | ○ | ○ | ○ | ○ | ○ | ○ |
| I must **always** be successful at school or work (7) | ○ | ○ | ○ | ○ | ○ | ○ | ○ |

| Q1620 | Disagree (1) (1) | 2 (2) | 3 (3) | 4 (4) | 5 (5) | 6 (6) | Agree (7) (7) |
|---|---|---|---|---|---|---|---|
| It **does not matter** to me when a close friend does not try their hardest (1) | ○ | ○ | ○ | ○ | ○ | ○ | ○ |
| People around me think I am **still competent** even if I **make a mistake** (2) | ○ | ○ | ○ | ○ | ○ | ○ | ○ |
| I **seldom expect** others to excel at whatever they do (3) | ○ | ○ | ○ | ○ | ○ | ○ | ○ |

Q2
Please indicate **how much** you are like the following statements:

| | Very much like me (5) | Pretty much like me (4) | Moderately like me (3) | Somewhat like me (2) | Not at all like me (1) |
|---|---|---|---|---|---|
| If I do not set the **highest standards** for myself, I am likely to end up a second rate person (MPS4 PS) | ○ | ○ | ○ | ○ | ○ |
| It is important to me that I be thoroughly competent in **everything I do** (MPS6 PS) | ○ | ○ | ○ | ○ | ○ |
| If I fail at work/school, I am a **failure** as a person (MPS9 CM) | ○ | ○ | ○ | ○ | ○ |
| If I fail **partly**, it is as bad as being a complete failure (MPS14 CM) | ○ | ○ | ○ | ○ | ○ |
| I am very good at focusing my efforts on **attaining a goal** (MPS16 PS) | ○ | ○ | ○ | ○ | ○ |
| Even when I do something very carefully, I often feel that it is **not quite** done right (MPS17 DA) | ○ | ○ | ○ | ○ | ○ |

Q1621
Please indicate **how much** you are like the following statements:

| | Very much like me (5) | Pretty much like me (4) | Moderately like me (3) | Somewhat like me (2) | Not at all like me (1) |
|---|---|---|---|---|---|
| I hate being **less than the best** at things (MPS4 PS) | ○ | ○ | ○ | ○ | ○ |
| I have extremely high **goals** (MPS6 PS) | ○ | ○ | ○ | ○ | ○ |
| People will probably **think less of me** if I make a mistake (MPS9 CM) | ○ | ○ | ○ | ○ | ○ |
| I usually have **doubts** about the simple everyday things I do (MPS14 CM) | ○ | ○ | ○ | ○ | ○ |
| I tend to get **behind** in my work because I **repeat** things over and over (MPS16 PS) | ○ | ○ | ○ | ○ | ○ |
| It takes me a long time to do something 'right' (MPS17 DA) | ○ | ○ | ○ | ○ | ○ |

**Treatment for Eating Disorders Questionnaire**

These next questions are about your past experience with treatment or healthcare.

Please answer the questions as best you can, and try to be as accurate as possible.

If you cannot answer a question, you can always choose the "don't know" or "prefer not to answer" option where available, or skip the question and move on.

Click "Next" below to continue to the questionnaire.

This section asks questions about you, your pathway into treatment, types of treatment, the impact on aspects of your life, various direct and indirect financial costs you may have experienced in relation to treatment, the type of therapy you received, your views about possible reasons why the eating disorder has developed, and your views on what is helpful in recovery from the eating disorder.

Treat1 What, if any of the following patterns, would best describe your treatment pathway?

○ High frequency at first, then gradually tapering off (1)

○ Low frequency at first, followed by a peak and then a general tapering off (2)

○ Reasonably constant during the eating disorder (3)

○ Intensive treatment when problems were severe or during relapses, but less frequent when eating disorder was manageable or in recovery (4)

○ Other (please comment) (5) ________________________________________________

○ No treatment (6)

Treat1b Have you ever experienced **binge eating**?

*Please note that the term "binge eating" means eating what others would regard as an* ***unusually***

***large amount of food** for the circumstances, accompanied by a sense of having **lost control** over eating.*

◯ Yes (1)

◯ No (2)

◯ Don't know (3)

*Display This Question:*

*If What, if any of the following patterns, would best describe your treatment pathway? != No treatment*

Treat2 How long after becoming aware of the symptoms of the eating disorder did you first seek help?

◯ Immediately (1)

◯ Within 1 month (2)

◯ Between 1 month and 6 months (3)

◯ Between 6 months and 1 year (4)

◯ More than 1 year (5)

*Display This Question:*

*If What, if any of the following patterns, would best describe your treatment pathway? != No treatment*

Treat3 Who first diagnosed you with an eating disorder?

- ◯ General practitioner/primary care physician/family medicine physician (1)
- ◯ Paediatrician/adolescent medicine physician (2)
- ◯ Psychologist (3)
- ◯ Psychiatrist (4)
- ◯ Other health professional (5) ________________________________________________
- ◯ I never received a formal diagnosis (6)
- ◯ Prefer not to answer (-99)

*Display This Question:*

*If What, if any of the following patterns, would best describe your treatment pathway? != No treatment*

Treat4 What was the nature of the treatment you received for your eating disorder? Check all that apply.

- ▢ Inpatient treatment (general hospital) (1)
- ▢ Inpatient treatment (general psychiatric unit) (2)
- ▢ Inpatient treatment (specialist eating disorders unit) (3)
- ▢ Residential treatment (4)
- ▢ Partial hospitalisation (residing at home, but attending a treatment centre up to 7 days per week) (5)
- ▢ Intensive outpatient treatment (6)
- ▢ Outpatient treatment (7)
- ▢ Emergency room visits (8)
- ▢ Another form of treatment or support (please specify) (9) ________________________________________________

*Display This Question:*

*If What was the nature of the treatment you received for your eating disorder? Check all that apply. = Inpatient treatment (general hospital)*

*Or What was the nature of the treatment you received for your eating disorder? Check all that apply. = Inpatient treatment (general psychiatric unit)*

*Or What was the nature of the treatment you received for your eating disorder? Check all that apply. = Inpatient treatment (specialist eating disorders unit)*

Treat5 How many inpatient hospitalisations for eating disorders have you had (all types of inpatient treatment)?

________________________________________________________________

---

*Display This Question:*

*If What was the nature of the treatment you received for your eating disorder? Check all that apply. = Inpatient treatment (general hospital)*

*Or What was the nature of the treatment you received for your eating disorder? Check all that apply. = Inpatient treatment (general psychiatric unit)*

*Or What was the nature of the treatment you received for your eating disorder? Check all that apply. = Inpatient treatment (specialist eating disorders unit)*

Treat6 What was the longest duration *(in days)* of inpatient treatment that you have had?

________________________________________________________________

*Display This Question:*

*If What was the nature of the treatment you received for your eating disorder? Check all that apply. = Emergency room visits*

Treat7 How many emergency room visits have you made related to an eating disorder?

________________________________________________________________

*Display This Question:*

*If What was the nature of the treatment you received for your eating disorder? Check all that apply. = Partial hospitalisation (residing at home, but attending a treatment centre up to 7 days per week)*

Treat8 How many partial hospitalisations have you had? *(where you continue to reside at home, but attend a treatment centre up to seven days a week.)*

________________________________________________________________

*Display This Question:*

*If What, if any of the following patterns, would best describe your treatment pathway? != No treatment*

Treat9 How difficult was it for you to access appropriate treatment—including both finding the right professional(s) for you and then obtaining appointments?

- ◯ Much more difficult than for treatment for other conditions (4)
- ◯ More difficult than for treatment for other conditions (3)
- ◯ About the same as for treatment for other conditions (2)
- ◯ Easier than for other conditions (1)
- ◯ Not difficult at all (0)

*Display This Question:*

*If What, if any of the following patterns, would best describe your treatment pathway? != No treatment*

*And Have you ever experienced binge eating? Please note that the term "binge eating" means eating wha... = Yes*

Treat10 Have you ever taken any of the following medications for **binge eating**? (check all that apply):

- ☐ Fluoxetine (Prozac) (1)
- ☐ Fluvoxamine (Faverin, Luvox) (2)
- ☐ Sertraline (Lustral, Zoloft) (3)
- ☐ Citalopram (Cipramil, Celexa) (4)
- ☐ Escitalopram (Cipralex, Lexapro) (5)
- ☐ Paroxetine (Seroxat, Paxil, Pexeva) (6)
- ☐ Vilazodone (Viibryd) (7)
- ☐ Phentermine (Ionamin, Adipex-p, Duromine, Metermine, Suprenza) (8)
- ☐ Orlistat (Alli, Xenical) (9)
- ☐ Phentermine/topiramate (Qsymia) (10)
- ☐ Naltrexone/bupropion (Mysimba, Contrave) (11)
- ☐ Lorcaserin (Lorqess, Belviq) (12)
- ☐ Lisdexamfetamine (Elvanse, Vyvanse) (13)
- ☐ Topiramate (Topamax, Trokendi XR, Qudexy XR) (14)
- ☐ Bupropion (Zyban, Wellbutrin) (15)
- ☐ Duloxetine (Cymbalta, Yentreve, Irenka) (16)
- ☐ Other medication: (17) ________________________________________________
- ☐ ⨂I have never taken any medication for binge eating (18)

☐ ⊗ Prefer not to answer (19)

Impact1 What degree of impact do you think that having an eating disorder has had on each of the following areas of your life?

| | Not applicable (0) | Very little or no impact (1) | Little impact (2) | Some impact (3) | Significant impact (4) | Very significant impact (5) |
|---|---|---|---|---|---|---|
| Your social life (Impact1_1) | | | | | | |
| Your overall wellbeing and quality of life (Impact1_2) | | | | | | |
| Your participation and productivity at work (Impact1_3) | | | | | | |
| Your engagement and attainment in your education (Impact1_4) | | | | | | |
| Your family in general (Impact1_5) | | | | | | |
| Your relationship with your parents (Impact1_6) | | | | | | |
| Your relationship with your siblings (Impact1_7) | | | | | | |
| Your relationship with your partner/spouse (Impact1_8) | | | | | | |

| | | | | | | |
|---|---|---|---|---|---|---|
| Your relationship with your children (Impact1_9) | ○ | ○ | ○ | ○ | ○ | ○ |
| Other family relationships (Impact1_10) | ○ | ○ | ○ | ○ | ○ | ○ |

Impact2 Do you think that your eating disorder had any impact on other family members *(e.g. your siblings)*?

○ Yes (4)

○ No (5)

○ Don't know (6)

---

*Display This Question:*

*If Do you think that your eating disorder had any impact on other family members (e.g. your siblings... = Yes*

*Or Do you think that your eating disorder had any impact on other family members (e.g. your siblings... = Don't know*

Impact2a **How** might your eating disorder have **impacted** your family members?

- ☐ Exhaustion/stress (3)
- ☐ Marital problems (4)
- ☐ Family ill-health (5)
- ☐ Less quality time together (17)
- ☐ Strained relationship with/between family members (6)
- ☐ Family members becoming withdrawn (14)
- ☐ Family members being overly attentive/overbearing (16)
- ☐ Lack of understanding of the eating disorder (15)
- ☐ Became closer with family (9)
- ☐ Became **more** trusting (12)
- ☐ Became **less** trusting (7)
- ☐ Resentment (8)
- ☐ Other (please specify): (10) ________________________________________________
- ☐ ⊗Don't know (13)
- ☐ ⊗Prefer not to answer (11)

Impact3 What is your current occupational status? Please check all that apply.

- ☐ Student (1)
- ☐ Full time paid worker (2)
- ☐ Part time paid worker (3)
- ☐ Stay at home parent (4)
- ☐ Unpaid carer (5)
- ☐ Retired (6)
- ☐ Unemployed (7)
- ☐ Other (8) ________________________________________________

Impact4 What is your approximate annual income?

- ○ I have no income (1)
- ○ Under £10,000 (2)
- ○ £10,000 - £19,999 (3)
- ○ £20,000 - £29,999 (4)
- ○ £30,000 - £39,999 (5)
- ○ £40,000 - £49,999 (6)
- ○ £50,000 - £59,999 (7)
- ○ £60,000 - £69,999 (8)
- ○ £70,000 - £79,999 (9)
- ○ £80,000 - £89,999 (10)
- ○ £90,000 - £99,999 (11)
- ○ £100,000 - £109,999 (12)
- ○ £110,000 - £119,999 (13)
- ○ £120,000 and above (14)
- ○ Prefer not to answer (-99)

Impact5 What was your approximate annual income before your eating disorder?

*Please add your income in thousands (e.g. £20000).*

________________________________________________________________

Impact6 What calendar year was that?

*Please enter in YYYY format.*

________________________________________________________________

Impact7 Consider the last year you experienced the eating disorder (if you currently have the eating disorder, consider the past year). During this time, did your eating disorder cause you to **work (or study) fewer hours** than you would have wanted to? If so, by how many **hours per week on average**?

- ○ None, i.e., no impact (1)
- ○ Up to 10 hours per week, on average, less participation (2)
- ○ 11-20 hours less participation (3)
- ○ 21-30 hours less participation (4)
- ○ 31-40 hours less participation (5)
- ○ Prevented any engagement in paid work or regular study (6)

Impact8 What calendar year was that?

*Please enter in YYYY format.*

________________________________________________________________

Impact9 Consider the last year you experienced the eating disorder (if you currently have the eating disorder, consider the past year). During this time, approximately how many **days** were you **unable to work or study** due to your eating disorder (e.g., sick days off work)?

________________________________________________________________

Impact10 What calendar year was that?

*Please enter in YYYY format.*

________________________________________________________________

Impact11 When you were/are at work/studying, did/does your eating disorder cause you to be less productive?

○ Yes (1)

○ No (0)

Display This Question:

*If When you were/are at work/studying, did/does your eating disorder cause you to be less productive? = Yes*

Impact12 Please estimate the percentage reduction in your productivity.

| | 0% reduction in productivity | 100% reduction in productivity |
|---|---|---|
| | 0 10 20 30 40 50 60 70 80 90 100 | |
| Percentage reduction in productivity () | | |

Impact14 Have you had to take extended leave of absences **(at least 4 weeks of sick leave)** from your work due to your eating disorder?

○ Yes (1)

○ No (0)

Display This Question:

*If Have you had to take extended leave of absences (at least 4 weeks of sick leave) from your work d... = Yes*

Impact15 How long *(in days)* were you off work or out of school?

________________________________________________________________

Impact16 Have you ever had to take a break or permanently leave your education due to your eating disorder?

- ○ Yes - I have taken a temporary break from a course of education (1)
- ○ Yes - I have permanently left a course of education (2)
- ○ No (0)

---

*Display This Question:*

*If Have you ever had to take a break or permanently leave your education due to your eating disorder? = Yes - I have taken a temporary break from a course of education*

*Or Have you ever had to take a break or permanently leave your education due to your eating disorder? = Yes - I have permanently left a course of education*

Impact17 What stage were you at when you left or took a break from your education? *Select all that apply.*

- ▢ I was in secondary school (1)
- ▢ I was in college/sixth form (2)
- ▢ I was an undergraduate at university (4)
- ▢ I was a postgraduate at university (9)

Impact19 Please provide an estimate of the average annual financial costs to you of the factors below related to your eating disorder:

- Expenditure on private treatment (1) ________________________________________________
- Expenditure due to publicly funded treatment (NHS) (2) ________________________________________________
- Travel costs for treatment (3) ________________________________________________
- Loss of income due to taking time off work (4) ________________________________________________
- Loss of income due to impacted educational or professional development (5) ________________________________________________
- Other expenditures (6) ________________________________________________

Impact20 Consider the last year you had an eating disorder (if you currently have an eating disorder, consider the past year). During this time, did you ever receive **professional treatment** for the eating disorder?

- Yes (1)
- No (0)

Impact21 What kind of professional help did you receive? Please indicate all that apply.

- ▢ General practitioner/primary care physician/family physician (1)
- ▢ Paediatrician/adolescent medicine physician (9)
- ▢ Counselor/social worker (2)
- ▢ Psychologist (3)
- ▢ Psychiatrist (4)
- ▢ Dietitian/Nutritionist (5)
- ▢ Eating disorder Specialist (6)
- ▢ Other health professional (7)
- ▢ Non-health professional (8)

---

*Display This Question:*

*If What kind of professional help did you receive? Please indicate all that apply. = Other health professional*

*Or What kind of professional help did you receive? Please indicate all that apply. = Non-health professional*

Impact22 Please specify what type of "other health professional" or a "non-health professional" you saw for your eating disorder:

________________________________________________________________

Impact23 Consider the last year you had an eating disorder (if you currently have an eating disorder, consider the past year). During this time, **how many times** did you **see** a health professional about your condition?

- ◯ 1 to 2 times (1)
- ◯ 3 to 5 times (2)
- ◯ 6 to 10 times (3)
- ◯ 11 to 20 times (4)
- ◯ 21 to 35 times (5)
- ◯ 36 to 50 times (6)
- ◯ More than 50 times (7)

Impact24 Were all costs of your treatment for the eating disorder covered by the **National Health Service (NHS)**?

- ◯ Yes (1)
- ◯ No (0)

---

*Display This Question:*

*If Were all costs of your treatment for the eating disorder covered by the National Health Service (... = No*

Impact25 Please indicate the **percentage of costs** that the NHS **covered** in the following categories:

| | Not covered (-9) | < 20% (1) | 21-40% (2) | 41-60% (3) | 61-80% (4) | 81-100% (5) | N/A (did not see this professional) (6) |
|---|---|---|---|---|---|---|---|
| General practitioner/primary care physician/family physician (Impact25_1) | ○ | ○ | ○ | ○ | ○ | ○ | ○ |
| Nurse/nurse practitioner (Impact25_11) | ○ | ○ | ○ | ○ | ○ | ○ | ○ |
| Paediatrician/adolescent medicine physician (Impact25_2) | ○ | ○ | ○ | ○ | ○ | ○ | ○ |
| Counsellor/social worker (Impact25_3) | ○ | ○ | ○ | ○ | ○ | ○ | ○ |
| Psychologist (Impact25_4) | ○ | ○ | ○ | ○ | ○ | ○ | ○ |
| Psychiatrist (Impact25_5) | ○ | ○ | ○ | ○ | ○ | ○ | ○ |
| Dietitian/nutritionist (Impact25_6) | ○ | ○ | ○ | ○ | ○ | ○ | ○ |
| Eating disorder specialist (Impact25_7) | ○ | ○ | ○ | ○ | ○ | ○ | ○ |
| Hospitalisation (Impact25_8) | ○ | ○ | ○ | ○ | ○ | ○ | ○ |
| Other health professional (Impact25_9) | ○ | ○ | ○ | ○ | ○ | ○ | ○ |
| Non-health professional (Impact25_10) | ○ | ○ | ○ | ○ | ○ | ○ | ○ |

Impact27 How much **money**, in total, have you and/or your family **spent** on the eating disorder **treatment** by health professionals out of your own pocket *(i.e. not NHS funded or reimbursed by private insurance)*?

○ None (0)

○ Up to £250 (1)

○ £251 to £500 (2)

○ £501 to £1000 (3)

○ £1001 to £1500 (4)

○ £1501 to £2000 (5)

○ £2001 to £5000 (7)

○ Over £5001 (6)

Impact28 How much money, in total, have you and/or your family spent on medications out of your own pocket *(i.e. not NHS funded or reimbursed by private health insurance)*?

○ None (0)

○ Up to £250 (1)

○ £251 to £500 (2)

○ £501 to £1000 (3)

○ £1001 to £1500 (4)

○ £1501 to £2000 (5)

○ Over £2000 (6)

Impact30 Consider the last year you had an eating disorder (if you **currently** have an eating disorder, consider the past year). During this time, how many times were you **hospitalised for treatment** related to your condition?

▼ 0 (0) ... 10+ (10)

*Display This Question:*

*If Consider the last year you had an eating disorder (if you currently have an eating disorder, cons... = 1*

*Or Consider the last year you had an eating disorder (if you currently have an eating disorder, cons... = 2*

*Or Consider the last year you had an eating disorder (if you currently have an eating disorder, cons... = 3*

*Or Consider the last year you had an eating disorder (if you currently have an eating disorder, cons... = 4*

*Or Consider the last year you had an eating disorder (if you currently have an eating disorder, cons... = 5*

*Or Consider the last year you had an eating disorder (if you currently have an eating disorder, cons... = 6*

*Or Consider the last year you had an eating disorder (if you currently have an eating disorder, cons... = 7*

*Or Consider the last year you had an eating disorder (if you currently have an eating disorder, cons... = 8*

*Or Consider the last year you had an eating disorder (if you currently have an eating disorder, cons... = 9*

*Or Consider the last year you had an eating disorder (if you currently have an eating disorder, cons... = 10+*

Impact31 What was the **total number of days** for all hospital admissions, if any, during this **year-long** period?

▼ 0 (4) ... 20+ (24)

*Display This Question:*

*If What was the total number of days for all hospital admissions, if any, during this year-long period? = 20+*

Impact31b What was the **total number of weeks** for all hospital admissions during this **year-long** period?

▼ 1 (1) ... 12+ (12)

*Display This Question:*

*If What was the total number of weeks for all hospital admissions during this year-long period? = 12+*

Q1699 What was the total number of months for all hospital admissions during this year-long period?

▼ 1 (1) ... 12 (12)

*Display This Question:*

*If Consider the last year you had an eating disorder (if you currently have an eating disorder, cons... = 1*

*Or Consider the last year you had an eating disorder (if you currently have an eating disorder, cons... = 2*

*Or Consider the last year you had an eating disorder (if you currently have an eating disorder, cons... = 3*

*Or Consider the last year you had an eating disorder (if you currently have an eating disorder, cons... = 4*

*Or Consider the last year you had an eating disorder (if you currently have an eating disorder, cons... = 5*

*Or Consider the last year you had an eating disorder (if you currently have an eating disorder, cons... = 6*

*Or Consider the last year you had an eating disorder (if you currently have an eating disorder, cons... = 7*

*Or Consider the last year you had an eating disorder (if you currently have an eating disorder, cons... = 8*

*Or Consider the last year you had an eating disorder (if you currently have an eating disorder, cons... = 9*

*Or Consider the last year you had an eating disorder (if you currently have an eating disorder, cons... = 10+*

Impact32 Were **all** of your hospital admissions during this year-long period classified as being **due to your eating disorder**?

○ Yes (1)

○ No (0)

○ Don't know (-9)

---

*Display This Question:*

*If Were all of your hospital admissions during this year-long period classified as being due to your... = No*

---

*Display This Question:*

*If Were all of your hospital admissions during this year-long period classified as being due to your... = No*

Impact33 If no or if not all, please indicate what the records said the admission(s) was/were for:

________________________________________________________________

*Display This Question:*

*If Consider the last year you had an eating disorder (if you currently have an eating disorder, cons... = 1*

*Or Consider the last year you had an eating disorder (if you currently have an eating disorder, cons... = 2*

*Or Consider the last year you had an eating disorder (if you currently have an eating disorder, cons... = 3*

*Or Consider the last year you had an eating disorder (if you currently have an eating disorder, cons... = 4*

*Or Consider the last year you had an eating disorder (if you currently have an eating disorder, cons... = 5*

*Or Consider the last year you had an eating disorder (if you currently have an eating disorder, cons... = 6*

*Or Consider the last year you had an eating disorder (if you currently have an eating disorder, cons... = 7*

*Or Consider the last year you had an eating disorder (if you currently have an eating disorder, cons... = 8*

*Or Consider the last year you had an eating disorder (if you currently have an eating disorder, cons... = 9*

*Or Consider the last year you had an eating disorder (if you currently have an eating disorder, cons... = 10+*

Impact34 For any of the admissions during this year-long period, did you and/or your family have to pay for any **hospital costs** out of your own pocket *(i.e. not paid by the NHS or reimbursed by an insurance company)*? If so, how much?

- No (0)
- Up to £250 (1)
- £251 to £500 (2)
- £501 to £1000 (3)
- £1001 to £1500 (4)
- £1501 to £2000 (5)
- £2001 to £2500 (6)
- £2501 to £5000 (8)
- Over £5001 (7)

*Display This Question:*

*If Consider the last year you had an eating disorder (if you currently have an eating disorder, cons... = 1*

*Or Consider the last year you had an eating disorder (if you currently have an eating disorder, cons... = 2*

*Or Consider the last year you had an eating disorder (if you currently have an eating disorder, cons... = 3*

*Or Consider the last year you had an eating disorder (if you currently have an eating disorder, cons... = 4*

*Or Consider the last year you had an eating disorder (if you currently have an eating disorder, cons... = 5*

*Or Consider the last year you had an eating disorder (if you currently have an eating disorder, cons... = 6*

*Or Consider the last year you had an eating disorder (if you currently have an eating disorder, cons... = 7*

*Or Consider the last year you had an eating disorder (if you currently have an eating disorder, cons... = 8*

*Or Consider the last year you had an eating disorder (if you currently have an eating disorder, cons... = 9*

*Or Consider the last year you had an eating disorder (if you currently have an eating disorder, cons... = 10+*

Impact35 As a result of any of the **admissions** during this year-long period, did you and/or your family incur any **travel, accommodation, or relocation expenses** when accessing treatment?

- No (0)
- Up to £250 (1)
- £251 to £500 (2)
- £501 to £1000 (3)
- £1001 to £1500 (4)
- £1501 to £2000 (5)
- £2001 to £5000 (7)
- Over £5000 (6)

*Display This Question:*

*If Have you ever experienced binge eating? Please note that the term "binge eating" means eating wha... = Yes*

Impact37 Consider the last year you had an eating disorder (if you currently have an eating disorder, consider the past year). If regularly **binge eating** during this time, how much do you think your food bill increased over this year-long period?

- Not applicable (-99)
- Up to £250 (1)
- £251 to £500 (2)
- £501 to £1000 (3)
- £1001 to £1500 (4)
- £1501 to £2000 (5)
- £2001 to £5000 (7)
- Over £5000 (6)

Impact39 Have you and/or your family ever had to **access financing, mortgage property, or sell assets** to pay for the eating disorder?

- No (0)
- Yes, less than £1000 (1)
- Yes, £1001 - £5000 (2)
- Yes, £5001 - £10,000 (3)
- Yes, over £10,000 (4)

Impact41 Do you **currently** experience any of the following health consequences as a **result of the eating disorder**?

*Please select all that apply.*

- ▢ Osteoporosis (1)
- ▢ Bone fractures (12)
- ▢ Infertility (2)
- ▢ Digestive disorders - stomach, esophagus, intestinal damage (3)
- ▢ Dental erosion (4)
- ▢ Obesity and obesity related disorders including diabetes (5)
- ▢ Heart disease and cardiac abnormalities (6)
- ▢ Kidney problems (7)
- ▢ Anxiety (8)
- ▢ Depression (9)
- ▢ Other mental health condition (10)
- ▢ Other physical health condition (13)
- ▢ ⊗None of the above (11)

---

*Display This Question:*

*If Do you currently experience any of the following health consequences as a result of the eating di... = Other mental health condition*

Impact42 **Please specify** which other **mental** health condition you **currently** experience as a result of the eating disorder:

________________________________________________________________

*Display This Question:*

*If Do you currently experience any of the following health consequences as a result of the eating di... = Other physical health condition*

Impact42b **Please specify** which other **physical** health condition you **currently** experience as a result of the eating disorder:

________________________________________________________________

Reason1 Below are possible reasons **why** people might develop an **eating disorder**. Please rate the extent to which each reason **applies to you**.

| | Not a reason (0) | Probably a reason (1) | To some extent a reason (2) | Definitely a major reason (3) |
|---|---|---|---|---|
| I was bullied or teased about my weight or appearance (Reason1_1) | ○ | ○ | ○ | ○ |
| I was bullied or teased about other things (Reason1_2) | ○ | ○ | ○ | ○ |
| I had low self-esteem (Reason1_3) | ○ | ○ | ○ | ○ |
| It is a biological or genetic illness (Reason1_4) | ○ | ○ | ○ | ○ |
| I felt pressure to be thin (Reason1_5) | ○ | ○ | ○ | ○ |
| Certain issues that happened to me as a child (Reason1_6) | ○ | ○ | ○ | ○ |
| Life was stressful (Reason1_7) | ○ | ○ | ○ | ○ |
| I was having difficulty with major life changes (Reason1_8) | ○ | ○ | ○ | ○ |
| There was conflict with key people in my life (Reason1_9) | ○ | ○ | ○ | ○ |
| There was no one to share my innermost thoughts and feelings with (Reason1_10) | ○ | ○ | ○ | ○ |
| I couldn't achieve what I wanted to (Reason1_11) | ○ | ○ | ○ | ○ |
| I felt pressure to succeed (Reason1_12) | ○ | ○ | ○ | ○ |

| | | | | |
|---|---|---|---|---|
| I wanted to get control of my life (Reason1_13) | ○ | ○ | ○ | ○ |
| Other reasons (Reason1_14) | ○ | ○ | ○ | ○ |

---

*Display This Question:*

*If Below are possible reasons why people might develop an eating disorder. Please rate the extent to... = Other reasons [ Probably a reason ]*

*Or Below are possible reasons why people might develop an eating disorder. Please rate the extent to... = Other reasons [ To some extent a reason ]*

*Or Below are possible reasons why people might develop an eating disorder. Please rate the extent to... = Other reasons [ Definitely a major reason ]*

Reason1a Please specify other reasons why people might develop an eating disorder:

________________________________________________________________

Reason2 Have you ever received any of the following therapies/treatments for the eating disorder?
*Check all that apply.*

- ☐ Family-based treatment (1)
- ☐ Cognitive behavior therapy (CBT) (2)
- ☐ Dialectical behavior therapy (DBT) (3)
- ☐ Specialist supportive clinical management (SSCM) (4)
- ☐ Psychodynamic (5)
- ☐ Interpersonal (6)
- ☐ Psychotherapy (7)
- ☐ Cognitive remediation (8)
- ☐ Metacognitive therapy (9)
- ☐ Group therapy (10)
- ☐ Other therapy type *(please specify)* (11)

________________________________________________

- ☐ I don't know the name of the therapy (12)
- ☐ ⊗ I have not received therapy/treatment (13)

Reason2a Did you find your treatment helpful or unhelpful?

*Display This Choice:*

*If Have you ever received any of the following therapies/treatments for the eating disorder? Check a... = Family-based treatment*

*Display This Choice:*

*If Have you ever received any of the following therapies/treatments for the eating disorder? Check a... = Cognitive behavior therapy (CBT)*

*Display This Choice:*

*If Have you ever received any of the following therapies/treatments for the eating disorder? Check a... = Dialectical behavior therapy (DBT)*

*Display This Choice:*

*If Have you ever received any of the following therapies/treatments for the eating disorder? Check a... = Specialist supportive clinical management (SSCM)*

*Display This Choice:*

*If Have you ever received any of the following therapies/treatments for the eating disorder? Check a... = Psychodynamic*

*Display This Choice:*

*If Have you ever received any of the following therapies/treatments for the eating disorder? Check a... = Interpersonal*

*Display This Choice:*

*If Have you ever received any of the following therapies/treatments for the eating disorder? Check a... = Psychotherapy*

*Display This Choice:*

*If Have you ever received any of the following therapies/treatments for the eating disorder? Check a... = Cognitive remediation*

*Display This Choice:*

*If Have you ever received any of the following therapies/treatments for the eating disorder? Check a... = Metacognitive therapy*

*Display This Choice:*

*If Have you ever received any of the following therapies/treatments for the eating disorder? Check a... = Group therapy*

*Display This Choice:*

*If Have you ever received any of the following therapies/treatments for the eating disorder? Check a... = Other therapy type <i>(please specify)</i>*

*Display This Choice:*

*If Have you ever received any of the following therapies/treatments for the eating disorder? Check a... = I don't know the name of the therapy*

| | Helpful (1) | Unhelpful (2) | Neither helpful nor unhelpful (3) | Don't know (-88) |
|---|---|---|---|---|
| *Display This Choice: If Have you ever received any of the following therapies/treatments for the eating disorder? Check a... = Family-based treatment* Family-based treatment (1) | ○ | ○ | ○ | ○ |
| *Display This Choice: If Have you ever received any of the following therapies/treatments for the eating disorder? Check a... = Cognitive behavior therapy (CBT)* Cognitive behaviour therapy (CBT) (2) | ○ | ○ | ○ | ○ |
| *Display This Choice: If Have you ever received any of the following therapies/treatments for the eating disorder? Check a... = Dialectical behavior therapy (DBT)* Dialectical behaviour therapy (DBT) (3) | ○ | ○ | ○ | ○ |
| *Display This Choice: If Have you ever received any of the following therapies/treatments for the eating disorder? Check a... = Specialist supportive clinical management (SSCM)* Specialist supportive clinical management (SSCM) (4) | ○ | ○ | ○ | ○ |
| *Display This Choice: If Have you ever received any of the following therapies/treatments for the eating disorder? Check a... = Psychodynamic* Psychodynamic (5) | ○ | ○ | ○ | ○ |
| *Display This Choice: If Have you ever received any of the following therapies/treatments for the eating disorder? Check a... = Interpersonal* Interpersonal (6) | ○ | ○ | ○ | ○ |

| | | | | |
|---|---|---|---|---|
| *Display This Choice:*<br>*If Have you ever received any of the following therapies/treatments for the eating disorder? Check a... = Psychotherapy*<br>Psychotherapy (7) | ○ | ○ | ○ | ○ |
| *Display This Choice:*<br>*If Have you ever received any of the following therapies/treatments for the eating disorder? Check a... = Cognitive remediation*<br>Cognitive remediation (8) | ○ | ○ | ○ | ○ |
| *Display This Choice:*<br>*If Have you ever received any of the following therapies/treatments for the eating disorder? Check a... = Metacognitive therapy*<br>Metacognitive therapy (9) | ○ | ○ | ○ | ○ |
| *Display This Choice:*<br>*If Have you ever received any of the following therapies/treatments for the eating disorder? Check a... = Group therapy*<br>Group therapy (10) | ○ | ○ | ○ | ○ |
| *Display This Choice:*<br>*If Have you ever received any of the following therapies/treatments for the eating disorder? Check a... = Other therapy type <i>(please specify)</i>*<br>Other: ${Reason2/ChoiceTextEntryValue/11} (11) | ○ | ○ | ○ | ○ |
| *Display This Choice:*<br>*If Have you ever received any of the following therapies/treatments for the eating disorder? Check a... = I don't know the name of the therapy*<br>Other therapy (name unknown) (12) | ○ | ○ | ○ | ○ |

*Display This Question:*

*If If Have you ever received any of the following therapies/treatments for the eating disorder? Check a... q://QID1920/SelectedChoicesCount Is Greater Than or Equal to 1*

*And Have you ever received any of the following therapies/treatments for the eating disorder? Check a...*

Reason4 Have you experienced **negative** or **unhelpful** things from the treatment you received?

*Please select all that apply.*

- ▢ More tense, sad, or unhappy (8)
- ▢ Feeling less hopeful (9)
- ▢ Symptoms worsened (10)
- ▢ More difficult to trust others (11)
- ▢ More difficult to make decisions on my own (18)
- ▢ Suicidal thoughts/intentions for the first time (12)
- ▢ Harder time dealing with past events (19)
- ▢ More conflict in relationships *(e.g. family or friends)* (20)
- ▢ Felt hurt or ridiculed by the therapist (13)
- ▢ Therapist violated confidentiality (14)
- ▢ Anxious that others could find out about therapy (17)
- ▢ Other (please specify): (15) ________________________________________________

- ▢ ⊗None of the above (16)

Recovery1 If you have recovered from an eating disorder or improved, please indicate the extent to which the following factors assisted the recovery process:

| | Not a factor (0) | Probably not a factor (1) | To some extent a factor (2) | Definitely an important factor (3) |
|---|---|---|---|---|
| Own motivation (Recovery1_1) | ○ | ○ | ○ | ○ |
| Family / partner involvement in treatment (Recovery1_2) | ○ | ○ | ○ | ○ |
| Family / partner support (Recovery1_3) | ○ | ○ | ○ | ○ |
| Support of friends (Recovery1_4) | ○ | ○ | ○ | ○ |
| Support groups / organisations (Recovery1_5) | ○ | ○ | ○ | ○ |
| Specific type of therapy received (Recovery1_6) | ○ | ○ | ○ | ○ |
| Relationship with treatment team (Recovery1_7) | ○ | ○ | ○ | ○ |
| New relationship (Recovery1_8) | ○ | ○ | ○ | ○ |
| New direction in education or new job (Recovery1_9) | ○ | ○ | ○ | ○ |
| Having a child (Recovery1_10) | ○ | ○ | ○ | ○ |
| Changing another important aspect of life (Recovery1_11) | ○ | ○ | ○ | ○ |
| Other (Recovery1_12) | ○ | ○ | ○ | ○ |

*Display This Question:*

*If If you have recovered from an eating disorder or improved, please indicate the extent to which th... = Other [ Probably not a factor ]*

*Or If you have recovered from an eating disorder or improved, please indicate the extent to which th... = Other [ To some extent a factor ]*

*Or If you have recovered from an eating disorder or improved, please indicate the extent to which th... =*

Recovery1a Please describe **other factors** that assisted with your recover process:

________________________________________________________________

### Medication Module

4) Have you ever taken antidepressants (even if it wasn't for depression or anxiety)?

☐Yes

☐No

☐Don't know

☐Prefer not to say

**4.1 only displayed if ‘yes’ is selected in 4**

4.1) Would you be willing to answer additional questions about antidepressants?

*Reminder: if you do not have time right now, but would like to answer questions about antidepressants later, you can always leave the survey and come back when you are ready.*

☐Yes
☐No

**4.2 only displayed if ‘yes’ is selected in 4.1**

4.2 Have you ever taken any of the following antidepressants (even if it wasn't for depression or anxiety)? *Please select all that apply.*

☐Citalopram (e.g. Cipramil)

☐Sertraline (e.g. Lustral, Zoloft)

☐Fluoxetine (e.g. Prozac, Olena, Oxactin, Prozep)

☐Paroxetine (e.g. Seroxat)

☐Mirtazapine (e.g. Zispin)

☐Amitriptyline (e.g. Tryptizol)

☐Dosulepin (e.g. Prothiaden)

☐Duloxetine (e.g. Cymbalta, Duciltia, Yentreve)

☐Venlafaxine (e.g. Efexor XL, Depefex XL, Foraven XL, Politid XL, Sunveniz XL, Tonpular XL, Venaxx XL, Venladex XL, Venlalic XL, Venzip XL, ViePax XL)

☐Trazodone (e.g. Molipaxin)

☐Escitalopram (e.g. Cipralex, Lexapro)

☐Nortriptyline (e.g. Allegron)

☐Clomipramine (e.g. Anafranil)

☐A different antidepressant that isn't listed above

**4.2a only displayed if ‘yes’ is selected in 4.2**

4.2a Have you ever taken any of the following antidepressants (even if it wasn't for depression or anxiety)? *Please select all that apply.*

☐Lofepramine (e.g. Lomont)

☐Imipramine (e.g. Tofranil)

☐Trimipramine (e.g. Surmontil)

☐Doxepin (e.g. Sineprin)

☐Reboxetine (e.g. Edronax)

☐Fluvoxamine (e.g. Faverin)

☐Moclobemide (e.g. Manerix)

☐Phenelzine (e.g. Nardil)

☐Tranylcypromine (e.g. Parnate)

☐Mianserin

☐Dapoxetine (e.g. Priligy, Westoxetin)

☐Tryptophan (e.g. Optimax)

☐Isocarboxazid

☐Hypericum perforatum (e.g. St John's wort)

☐Vortioxetine (e.g. Brintellix)

☐A different antidepressant that isn't listed above

4.2b What other antidepressants have you ever taken that weren't listed previously (even if it wasn't for depression or anxiety)?

______________________________________________________

4.3 When you were taking these antidepressants, were you also taking any other prescribed medication?

☐No
☐Yes
☐Don't know

4.3-1 If you answered yes to any of the above, please answer the following questions for EACH medication you have taken previously:

4.3a Why were you prescribed each medication? (e.g. Sertaline for anxiety)

4.3b How old were you when you started taking each medication? (e.g. Sertaline, 18 years)

4.3c How long did you take/ have you been taking each medication? (e.g. Sertaline for 5 months)

**4.4 displayed if 'yes' is selected in 4.3**

4.4 Have you ever taken any of the following medications while you were also taking antidepressants?

☐Quetiapine (e.g. Brancico, Psyquet, Seroquel (XL), Atrolak XL, Biquelle (XL), Mintreleq, Sondate XL, Zaluron XL, Atrolak XL, Ebesque, Tenprolide)
☐Olanzapine (e.g. ZypAdhera, Zalasta, Zyprexa)
☐Prochlorperazine (e.g. Buccastem, Stemetil)
☐Risperidone (e.g. Risperdal, Risperdal Consta)
☐Aripiprazole (e.g. Abilify)
☐Amisulpride (e.g. Solian)
☐Haloperidol (e.g. Haldol decanoate, Dozic, Serenace)
☐Chlorpromazine (e.g. Largactil)
☐Flupentixol (e.g. Fluanxol, Depixol, Psytixol)
☐Promazine
☐Zuclopenthixol (e.g Ciatyl-Z, Clopixol Acuphase/Concentrate)
☐ Levomepromazine (e.g. Nozinan)
☐ Trifluoperazine (e.g. Stelazine)
☐ Pericyazine (e.g. Neulactil)
☐ Fluphenazine (e.g. Modecate)
☐ Benperidol (e.g. Anquil)
☐ Clozapine (e.g. Denzapine, Zaponex, Clozaril)
☐ Pimozide (e.g. Orap)
☐ Lurasidone (e.g Latuda)
☐ Perphenazine (e.g. Fentazin)
☐ Paliperidone (e.g. Invega, Trevicta, Xeplion)

4.5 Have you ever taken any of the following medications while you were also taking antidepressants?

☐ Zopiclone (e.g. Zimovane)
☐ Pregabalin (e.g. Axalid, Lyrica, Rewisca, Lecaent, Alzain)
☐Propranolol (e.g. Bedranol, Beta-Prograne)
☐ Zolpidem (e.g. Stilnoct)
☐Melatonin (e.g. Circadin)
☐ Buspirone
☐ Oxprenolol (e.g. Slow-Trasicor)
☐Zaleplon

4.6 Have you ever taken any of the following medications while you were also taking antidepressants?

☐ Diazepam (e.g. Stesolid)
☐Temazepam
☐Lorazepam
☐Clonazepam
☐Nitrazepam
☐ Clobazam (e.g. Perizam, Tapclob, Zacco, Frisium)
☐Oxazepam
☐Alprazolam (e.g. Xanax)
☐Chlordiazepoxide (e.g. Librium)
☐Loprazolam
☐Lormetazepam

4.7 Have you ever taken any of the following medications while you were also taking antidepressants?

☐Valproate/valproic acid (e.g. Depakote, Convulex, Epilim (Chronosphere), Epival, Episenta)
☐Carbamazepine (e.g Carbagen, Tegretol)
☐Lithium carbonate/citrate (e.g. Priadel, Liskonum, Camcolit, Li-Liquid)

5 After taking each of the antidepressants for a period of time, did you ever experience any further symptoms associated with the condition for which you were prescribed antidepressants?

☐ Yes ☐ No ☐ Don’t know ☐ Prefer not to say

6 How long did the improvements in symptoms you experienced after taking the antidepressants last for?

☐ Less than a month
☐ 1 to 2 months
☐ 3 to 6 months
☐ 7 to 12 months
☐ More than 12 months
☐ Not sure
☐ I didn't have any improvement in symptoms

7 Overall, how would you rate the benefits of taking antidepressants?

☐ 1 (lowest/worst) ☐ 2 ☐ 3 ☐ 4 ☐ 5 (highest/best)

8 What were the worst aspects of taking the antidepressant(s)?

☐ New side effects like nausea, headache, drowsiness, fatigue, sexual dysfunction
☐ Increased depressive symptoms like anxiety, agitation, sleep disturbance
☐ Knowing that I needed to take medications to get well
☐ Telling others that I needed to take medications
☐ Increased suicidal thoughts or actions
☐ Interfered with my capacity to do normal daily activities
☐ Increased direct costs (e.g. seeing private doctors, buying medications)
☐ Other (please specify): ____________________________

9 Did you experience side effects from any antidepressant?

☐ Yes (continue to #11) ☐ No (skip to Section 5)

10 Please list the side effects you experienced from taking antidepressants:

______________________________________________________________________
______________________________________________________________________
______________________________________________________________________

11 Did you have to stop taking any antidepressant because of side effects?

☐ Yes ☐ No

12a. If yes, which antidepressant did you stop taking? ______________________

12 Overall, how would you rate the side effects of taking antidepressants?

☐ 1 (lowest/worst) ☐ 2 ☐ 3 ☐ 4 ☐ 5 (highest/best)

13 What were the worst aspects of taking the antidepressant(s)?

☐ New side effects like nausea, headache, drowsiness, fatigue, sexual dysfunction
☐ Increased depressive symptoms like anxiety, agitation, sleep disturbance
☐ Knowing that I needed to take medications to get well
☐ Telling others that I needed to take medications
☐ Increased suicidal thoughts or actions
☐ Interfered with my capacity to do normal daily activities
☐ Increased direct costs (e.g. seeing private doctors, buying medications)
☐ Other (please specify): __________________________
